# US Exceptionalism? International Trends in Midlife Mortality

**DOI:** 10.1101/2023.07.25.23293099

**Authors:** Jennifer Beam Dowd, Katarzyna Doniec, Luyin Zhang, Andrea Tilstra

## Abstract

**Background:** Rising midlife mortality in the United States (US) has raised concerns, particularly the increase in “deaths of despair” (due to drugs, alcohol, and suicide). While life expectancy is also stalling in other countries such as the UK, whether midlife mortality is rising outside the US is not known.

**Methods:** We document trends in midlife mortality in the US, UK and a group of 16 high-income countries in Western Europe, Australia, Canada, New Zealand, and Japan, as well as 7 Central and Eastern European (CEE) countries from 1990-2019. We use annual mortality data from the World Health Organization Mortality Database to analyze sex and age-specific (25-44, 45-54, and 55-64) age-standardized death rates across 13 major cause-of-death categories.

**Findings:** US midlife mortality rates worsened since 1990 for several causes of death including drug-related, alcohol-related, suicide, metabolic disease, nervous system disease, respiratory disease, and infectious/parasitic diseases. Deaths due to homicide, transport accidents, and cardiovascular disease declined overall since 1990 but saw recent increases or stalling of improvements. Midlife mortality has also recently increased in the UK for 45-54-year-olds, and in Canada, Poland, and Sweden among 25-44-year-olds.

**Conclusion:** The US is increasingly falling behind not only high-income but also CEE countries heavily impacted by the post-Soviet mortality crisis of the 1990s. While levels of midlife mortality in the UK are substantially lower than in the US overall, there are signs that UK midlife mortality is worsening relative to the rest of Europe.

## Introduction

High-income countries experienced unprecedented improvements in life expectancy for over a hundred years, at a pace of almost 2.5 years per decade (Vaupel et al., 2021). The seemingly unstoppable gains faltered in the 2010s with life expectancy reversals in the US and stagnation in the UK (Marmot Indicators 2017, Raleigh, 2018, Harris et al., 2021), despite continued improvements for most other countries. Prior to the COVID-19 pandemic, life expectancy losses in the US were attributed to rising mortality in midlife due to slowing improvements in cardiovascular conditions and increases in deaths from “despair” related causes, including drugs, alcohol, and suicide (Case & Deaton, 2015, 2017, Ho, 2019, Woolf & Schoomaker, 2019, Harris et al., 2021, Masters et al., 2018). In the UK, stalling life expectancy in the 2010s has been attributed to austerity policies and declining investments in health and social care, though these hypotheses are difficult to test empirically (McCartney et al., 2022, BMJ, 2019). Cross-country comparisons of overall life expectancy trends are well-documented (for examples, Ho & Hendi, 2018, Leon et al., 2019), but less is known about how trends in midlife mortality compare across countries. How the US and other high-income countries compare to CEE countries that experienced a relatively recent mortality crisis is not known. While life expectancy changes in higher income countries are often heavily influenced by mortality at older ages where most deaths occur, trends in midlife mortality can identify changing mortality dynamics in younger cohorts that foreshadow future trends in life expectancy.

We aim to provide an international comparative perspective on midlife mortality to better understand recent life expectancy declines in the US and stagnation in the UK. We analyzed trends in cause-specific midlife mortality in the US, the UK and a group of 16 high-income (HI) and 7 Central and Eastern European countries (CEE) from 1990-2019. The selected CEE countries traditionally had among the highest rates of despair-related deaths globally, particularly among working-age men (Scheiring et al., 2021).

## Data and Methods

We used cause-specific mortality and population counts from the World Health Organization (WHO) Mortality Database for 25 countries from 1990 up to the most recent year available (from 2016 to 2019, see Tables 2-3 in Appendix 1 for detailed information on death counts and population estimates data availability). The 18 HI countries included: Austria, Australia, Belgium, Canada, Denmark, France, Finland, Germany, Italy, Japan, the Netherlands, Norway, Portugal, Spain, Sweden, Switzerland, the United Kingdom, and the United States. The 7 Central and Eastern European (CEE) countries analyzed included Bulgaria, Czech Republic, Hungary, Poland, Romania, Slovakia, and Slovenia. For Canada 2006–2019, the US 2008–2019, and France 2015–2019, population counts were not available in the WHO database and were drawn from the respective official vital statistics agencies of each country (i.e, Statistics Canada, the US Census Bureau, and the France National Institute of Statistics and Economic Studies).

We examined all-cause mortality and 13 mutually exclusive major causes of death, (see Table 1 in Appendix 1 for the corresponding ICD-9 and ICD-10 codes): (i) infectious and parasitic diseases, excluding HIV/AIDS, (ii) HIV/AIDS, (iii) respiratory diseases, (iv) trachea/bronchus and lung cancers (v) all other cancers, (vi) nervous system diseases, (vii) metabolic diseases, (viii) cardiovascular disease, (ix) suicide, (x) homicide, (xi) transport accidents, (xii) all other external causes, and (xiii) a residual category, capturing all other causes of death. Due to the limitations of the simplified ICD-9 classification (ICD-9 BTL) used by WHO, we were not able to match ICD-9 and ICD-10 codes for (xii) drug- and (xiii) alcohol-related causes. For this reason, we analyzed trends in these two categories for the period 2000-2019, when ICD-10 classification was introduced in most countries in our sample.

We calculated all-cause and cause-specific mortality rates as deaths per 100,000 individuals for three age groups (25-44, 45-54, 55-64) by country, sex, and year. Rates were age-standardized within each age category using the 2013 European Standard Population. To smooth short-term fluctuations with small counts, we report 3-year moving averages of standardized mortality rates. We also calculated and visualized the percent change in mortality from the baseline for each year to show relative changes over time.

All analyses are fully reproducible, the data and code can be accessed here.

## Results

Because of the large number of country by age-sex-year-cause of death combinations, our figures highlight trends in the US and UK compared to the mean of HI countries (excluding the US and UK) as well as the mean of CEE countries. Full results for absolute death rates for each cause of death are available in Appendix 2. In addition, calculations of relative mortality change for each age group, sex and cause are available in Appendix 3. Individual country figures, showing absolute levels of age-standardized mortality for each cause of death, sex and age are available in Appendix 4.

### All-cause mortality

Figure 1 shows trends in all-cause mortality rates for males and females in three age groups (25-44, 45-54, 55-64) in the 25 countries from 1990-2019. Most countries experienced substantial declines in midlife mortality since 1990 but at different speeds. CEE countries showed dramatic improvements over this period, recovering from the high mortality of the post-Soviet mortality crisis. During the 1990s, the highest overall mortality rates among CEE countries were observed in Hungary, Poland, Romania, and Bulgaria, with Slovenia having the lowest rates. Despite these improvements, the average mortality rate in the CEE group in 2019 remained substantially higher than the average for high-income countries (about twice as high for CEE males and 1.5 times higher for CEE females, depending on the age group), excluding the US and the UK.

**Figure 1.**
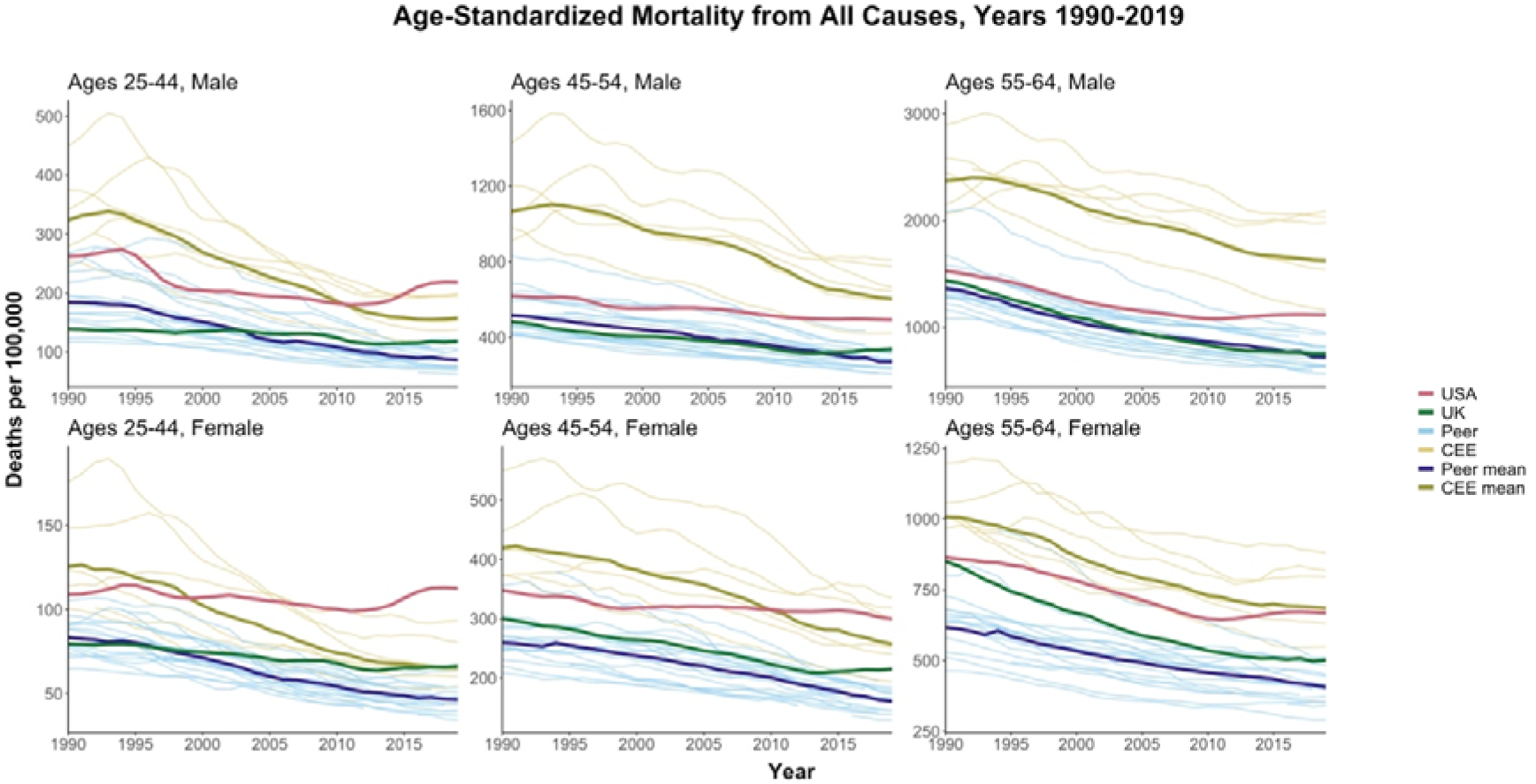
All-cause mortality by age and sex.

US trends diverged substantially from other countries. In contrast to the consistent declines in HI peer and CEE countries, all-cause mortality trends in the US were flat or increasing in most age-sex groups. The divergence was most noticeable for 25–44-year-olds, where US all-cause mortality rates rose above the CEE mean in the late 1990s for females and in 2011 for males. By 2019, US mortality was not only above the CEE mean in this age group, but also higher than each individual CEE country. Compared to peer countries, US all-cause rates in 2019 were 2.5 times higher for both males and females. At ages 45-54, all-cause mortality rates in 2019 were 88% higher for US females than the high-income peer mean, and 85% higher for US males compared to the peer mean. CEE countries consistently had the highest mortality for ages 55-64, but US females in this age group were roughly equal to the CEE mean by 2019. Overall, US midlife mortality worsened on a relative basis compared to both peer and CEE countries, most noticeably for females and younger middle-aged groups.

Figure 2 shows the percent change in all-cause mortality rates across age, sex and country relative to 1990 (or the first year after 1990 for which data is available). These relative changes highlight the lack of improvements among US women aged 25-44 since 1990, and the recent stagnation of improvements compared to other countries overall in the 25-44 and 45-54 age groups. For 55–64-year-olds in the US, relative mortality improvements tracked other countries until around 2010, when mortality improvements reversed.

**Figure 2.**
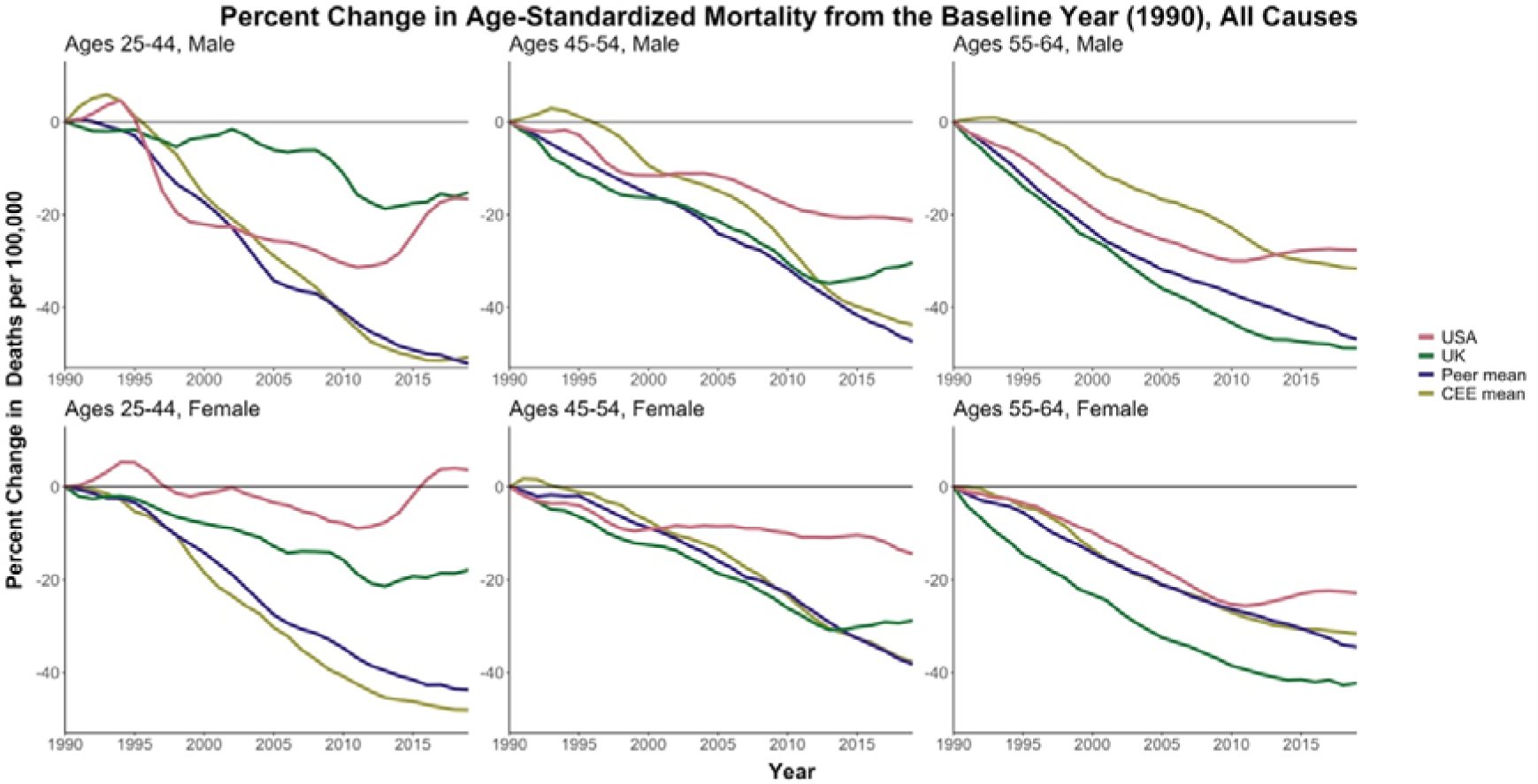
Percent change in all-cause mortality rates since 1990.

The UK fared better than the US, but its performance worsened over time compared to its high-income peers, particularly at the youngest ages. Mortality in UK males aged 25-44 was mostly flat between 1990 and 2003 and then rose above the peer mean. UK male mortality at ages 45–54 increased starting in 2015, in contrast to continued declines in peer countries.

Despite the relative and absolute deterioration, all-cause mortality rate for UK males ages 45- 54 in 2019 (329 deaths per 100,000) was well below both the US (490 per 100,000) and CEE mean (594 per 100,000). Across the analysis years, UK females in younger midlife fared worse than peer and CEE countries. Despite the good relative standing, death rates among UK 25-44- and 45–54-year-olds of both sexes were mostly stable rather than improving since the early 2010s, a possible contributor to stalling life expectancy.

### Cause-specific Mortality

There was more variation in patterns for cause-specific mortality, so we summarize the main points of interest, with full results available in Appendix 2. The US stood apart with higher mortality for many causes compared to both HI peers *and* the CEE countries. For example, while deaths from transport accidents fell since 1990 across most countries, the US rates for males aged 25-44 stagnated in the late 1990s and early 2000s, and increased since 2010, with levels around 3.5 times as high as peer high-income countries and 70% higher than CEE countries by 2019 (Figure 3). These patterns were similar at older ages and for US females, where mortality from transport accidents was well above the CEE mean for most of the period. In contrast, the UK has among the lowest death rates from transport accidents among all countries.

**Figure 3.**
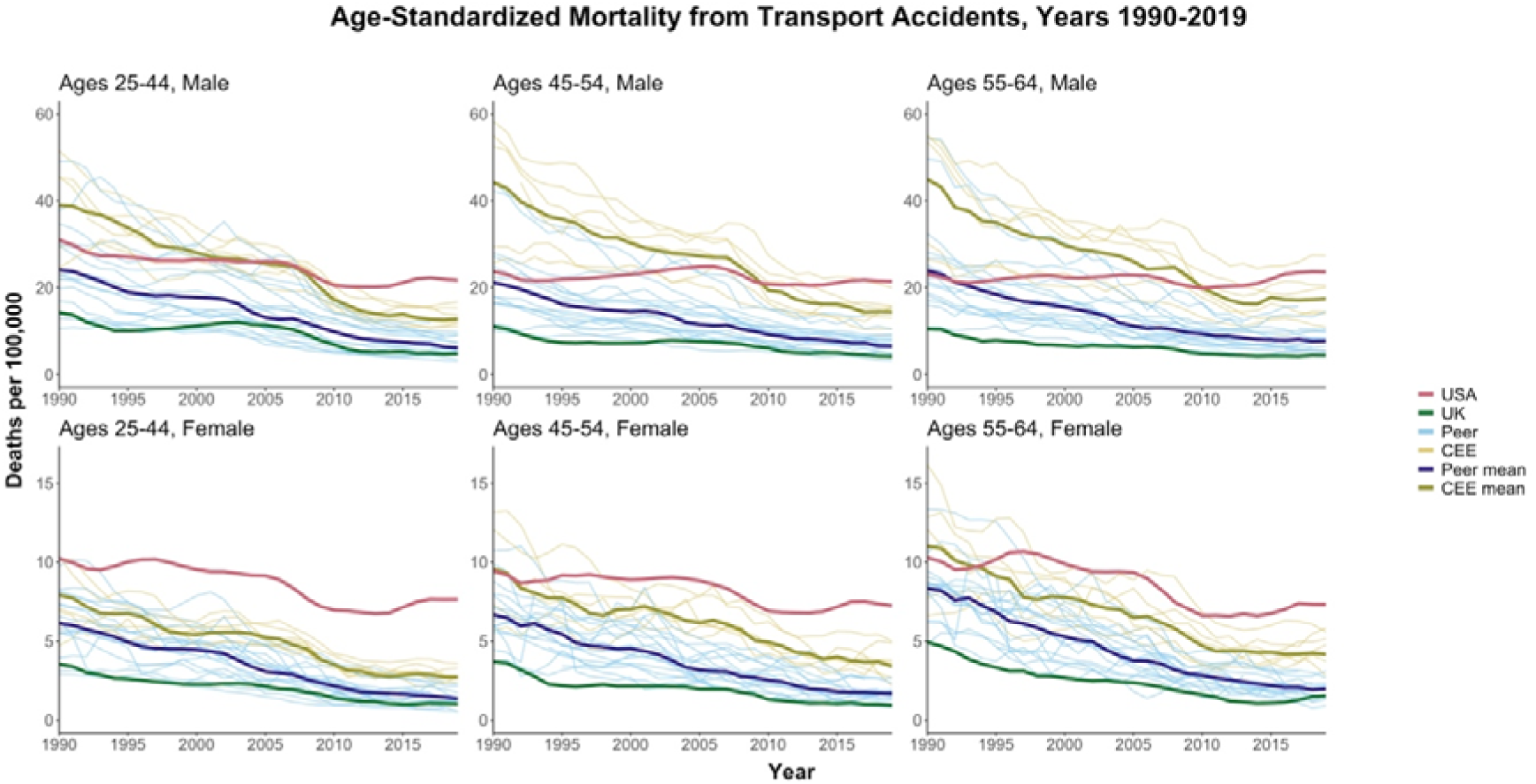
Age-standardized mortality rates from transport accidents.

This negative US exceptionalism also carried over to death by homicide (Figure 4). US homicide rates were far above all other countries for both sexes in all age groups. While US homicide mortality declined substantially from 1990 to 2000, rates have then stabilized and crept upward since around the mid-2010s. For 25–44-year-old males, the homicide mortality rate for US males was nearly 15 times higher than both the peer and CEE mean in 2019. The UK homicide deaths were below peer means across all age groups, though the rate has risen slightly in recent years.

**Figure 4.**
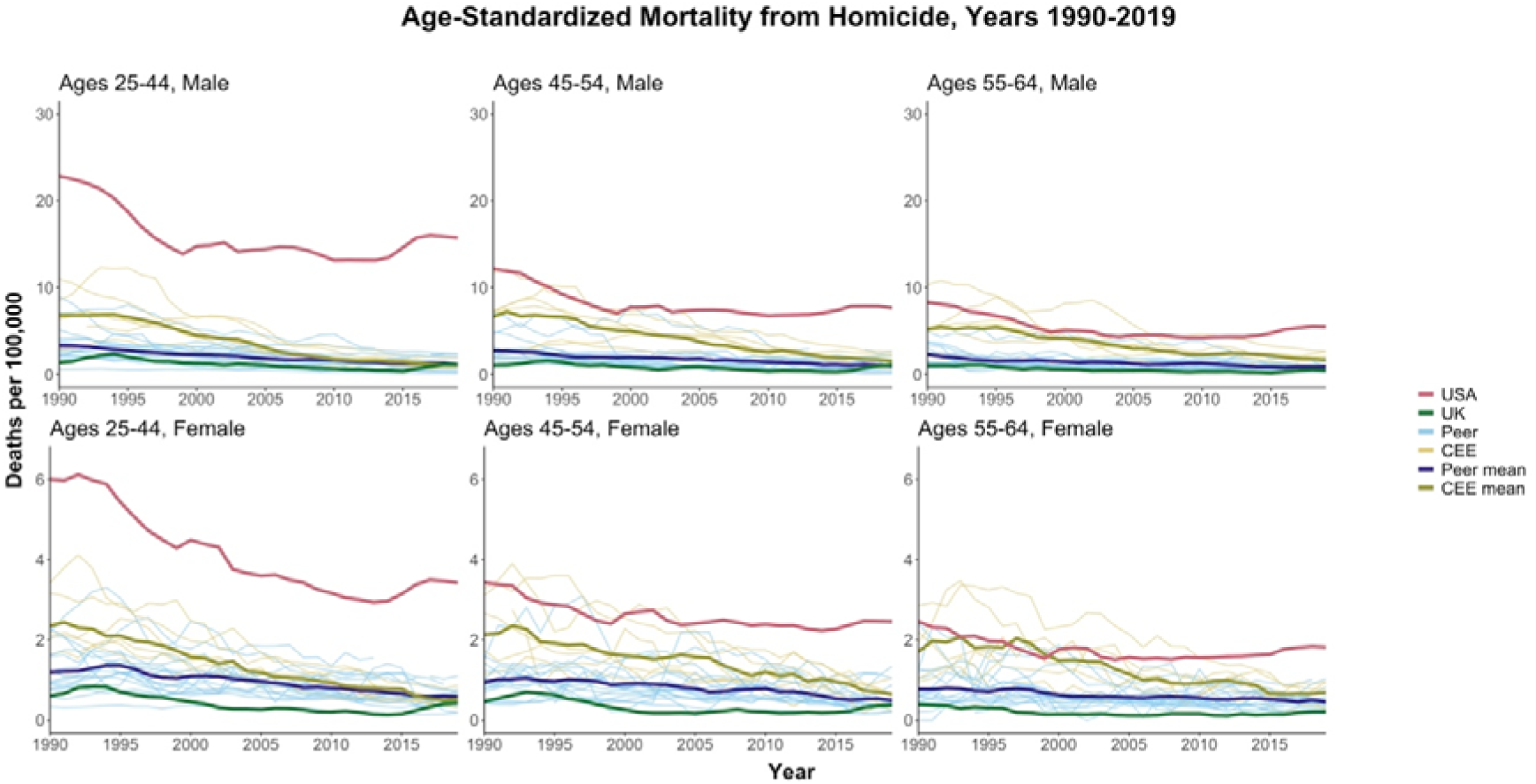
Age-standardized mortality rates from homicide.

“Despair”-related causes of death Suicide

In 1990, the US had suicide mortality rates mostly below peer means (Figure 5). A gradual uptick since the late 1990s/early 2000s, coupled with continued declines elsewhere, left the US with rates above both peer and CEE means by 2019. The US divergence from peers was largest for 25–44-year-old males, where 2019 rates were 1.65 times higher than the peer mean and 1.72 times higher than the CEE mean. The UK suicide mortality was generally low compared to other countries, but there were some increases at younger ages since 2010, bringing it closer to high-income peers and CEE means, but still far below the US.

**Figure 5.**
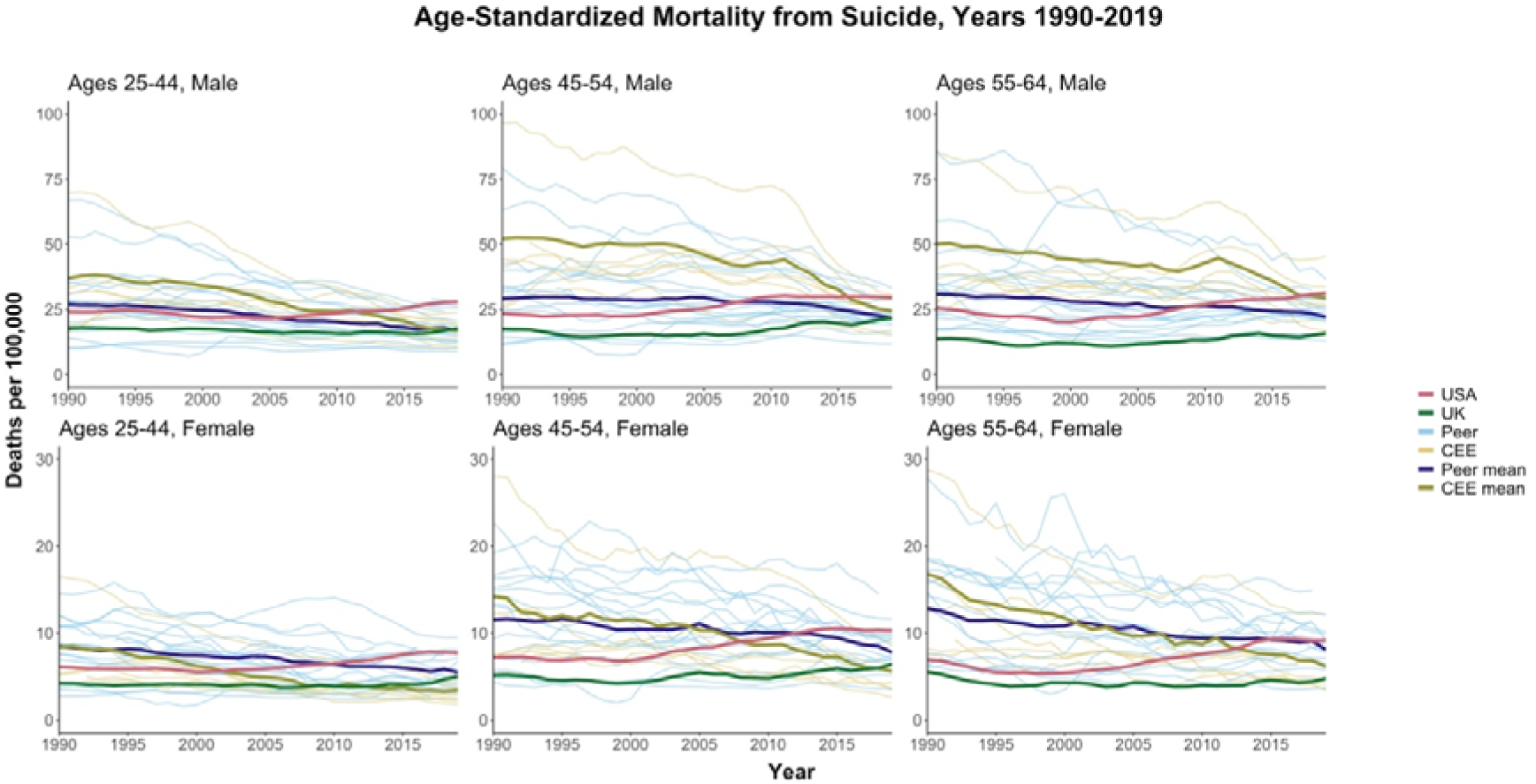
Age-standardized mortality rates from suicide.

**Figure 6.**
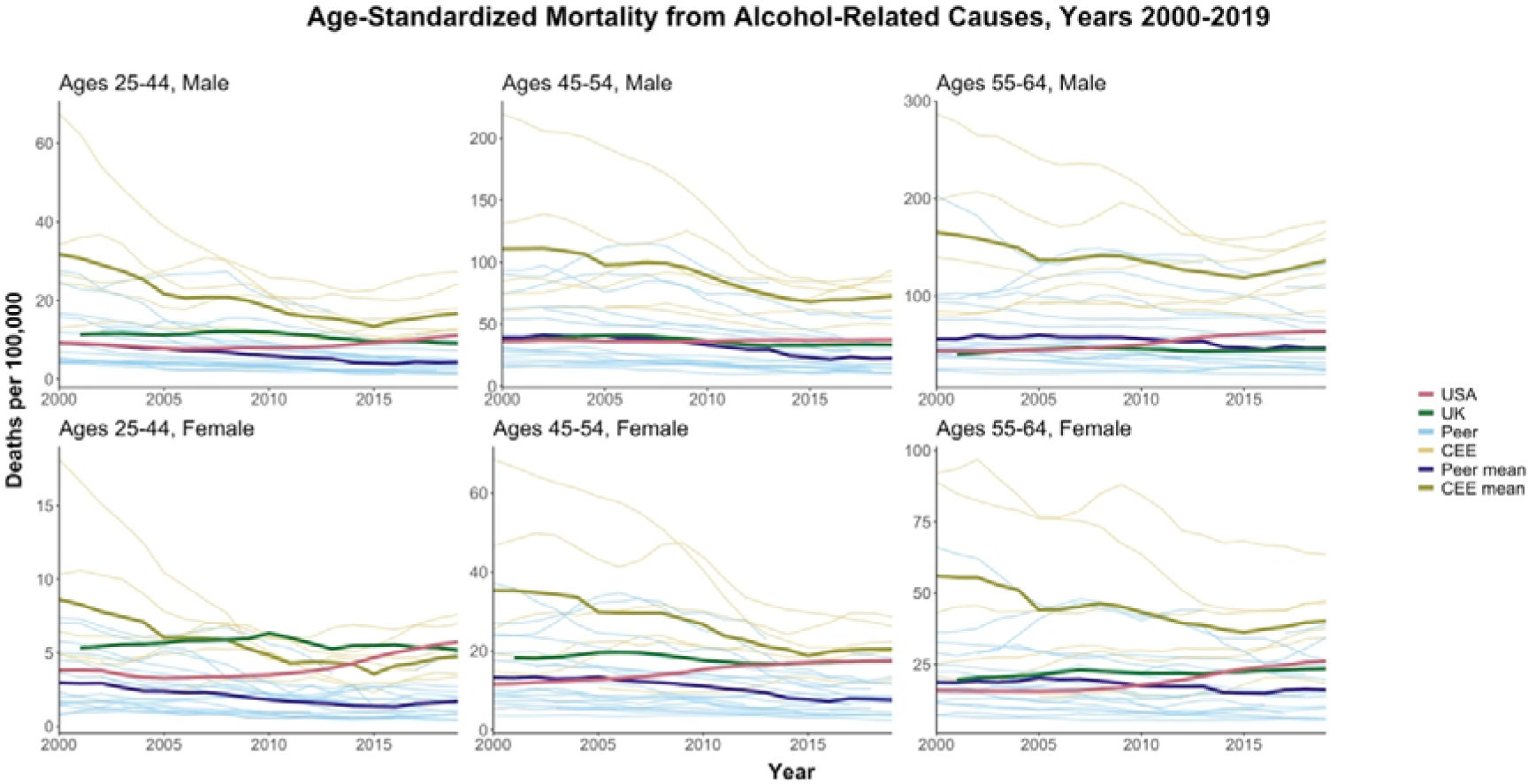
Age-standardized alcohol-related mortality.

### Alcohol-related deaths

The CEE countries continued to have the highest levels of alcohol-related mortality, especially at older ages, but these have generally decreased over time. The US began the period below the peer mean for alcohol-related deaths in most age-sex groups but trended upward since late 2000s, ending the period above both the UK and peer mean in most age-sex categories. Alcohol-related mortality was higher in the UK compared to the peer means in most age-sex categories but has remained mostly flat since 2000s.

### Drug-related deaths

US trends in drug-related mortality diverged noticeably from other countries, with levels increasing dramatically from 2000 to 2019. The highest US levels were seen in 45–54-year-old males, but the dramatic increases were universal across all age-sex groups. Drug deaths also increased in the UK since the late 2000/early 2010s and worsened compared to peer means, but absolute levels were still much lower than in the US. Trends in drug-related deaths in the HI peer and CEE mean countries were relatively flat, confirming the unique nature of high drug-related mortality in the U.S.

### Cardiovascular disease

Cardiovascular disease (CVD) mortality was highest in CEE countries in the 55-64 age group, but dramatic declines brought the CEE closer to overall levels in the US by 2019 (Figure 8). CVD mortality in the oldest group (55-64) in the US was similar or better than the UK in 1990 but both countries witnessed gradual declines followed by stagnation for both sexes by 2019. By 2019, CVD mortality in the US in this age group was worse than in all peer countries and some individual CEE countries. At younger ages, the US stagnation in CVD mortality improvements was more evident, particularly for females. By 2019, CVD mortality for US women at ages 25-44 and 45-54 was higher than the CEE mean despite much higher CEE levels in 1990. UK CVD mortality at the same ages was above the peer mean by 2019, with initial declines flattening out in recent years. The slower declines in the US compared to its peers were particularly clear looking at relative changes (Appendix 3, Figure 8), as is the relative stagnation for 25–44-year-olds in the UK.

### Lung Cancer

For males in all three age groups, lung cancer mortality consistently declined in most countries, with levels in the CEE still higher in 2019 than all high-income peers (Figure 9). For example, in the 55-64 group in 2019, the mortality rate in the US (74 per 100,000) was close to the average of its high-income peers (73 per 100,000), higher than the UK (61 per 100,000), yet significantly lower than the CEE mean (154 per 100,000).

Patterns were more varied for females. For those aged 55-64, lung cancer mortality rose in the CEE and HI peer mean over this period, reflecting different patterns of smoking initiation and cessation compared to men (Peters et al., 2014). US females aged 55-64 saw substantial declines from 1990 levels that were much higher than peer countries. In contrast, rates for UK females aged 55-64 were above peers for most of this period. By 2019, rates for females of all ages converged to similar levels among countries.

### Other causes of death

Figures for all other cause of death categories are available in Appendix 2. Here we highlight a few notable results. Mortality due to metabolic disease, which includes diabetes, was noticeably higher in the US compared to the UK, other high-income peers, and the CEE mean (Appendix 2, Figure 7). Similarly, deaths from respiratory disease for females were higher in the US and UK than high-income peers or the CEE mean, and these deaths trended up in the last decade in the US. For males, the CEE countries did worst, but the US and UK were far above the other HI peer means (Appendix 2, Figure 3). The US also stood out with much higher levels of mortality from (non-HIV) infectious and parasitic diseases (Appendix 2, Figure 2). These levels rose considerably over time in the US in the older age groups, while remaining relatively flat elsewhere. The UK rates, in comparison, were generally lower than peers for infectious and parasitic diseases. Cancer was one cause of death for which the US performed relatively well, with rates below the UK and peer mean for most of the period, except for 25-44 year-olds (Appendix 2, Figure 5).

**Figure 7.**
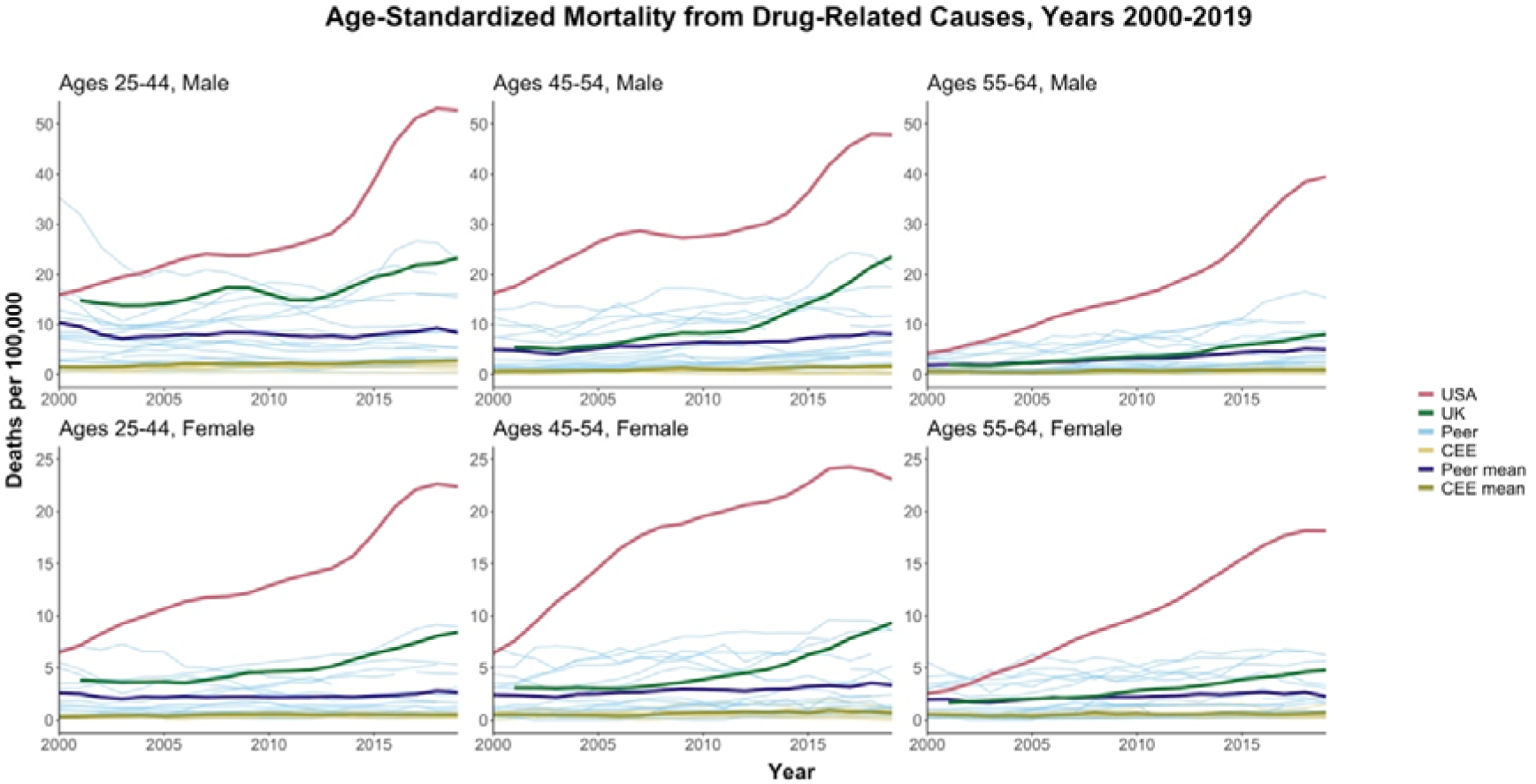
Age-standardized drug-related mortality.

**Figure 8.**
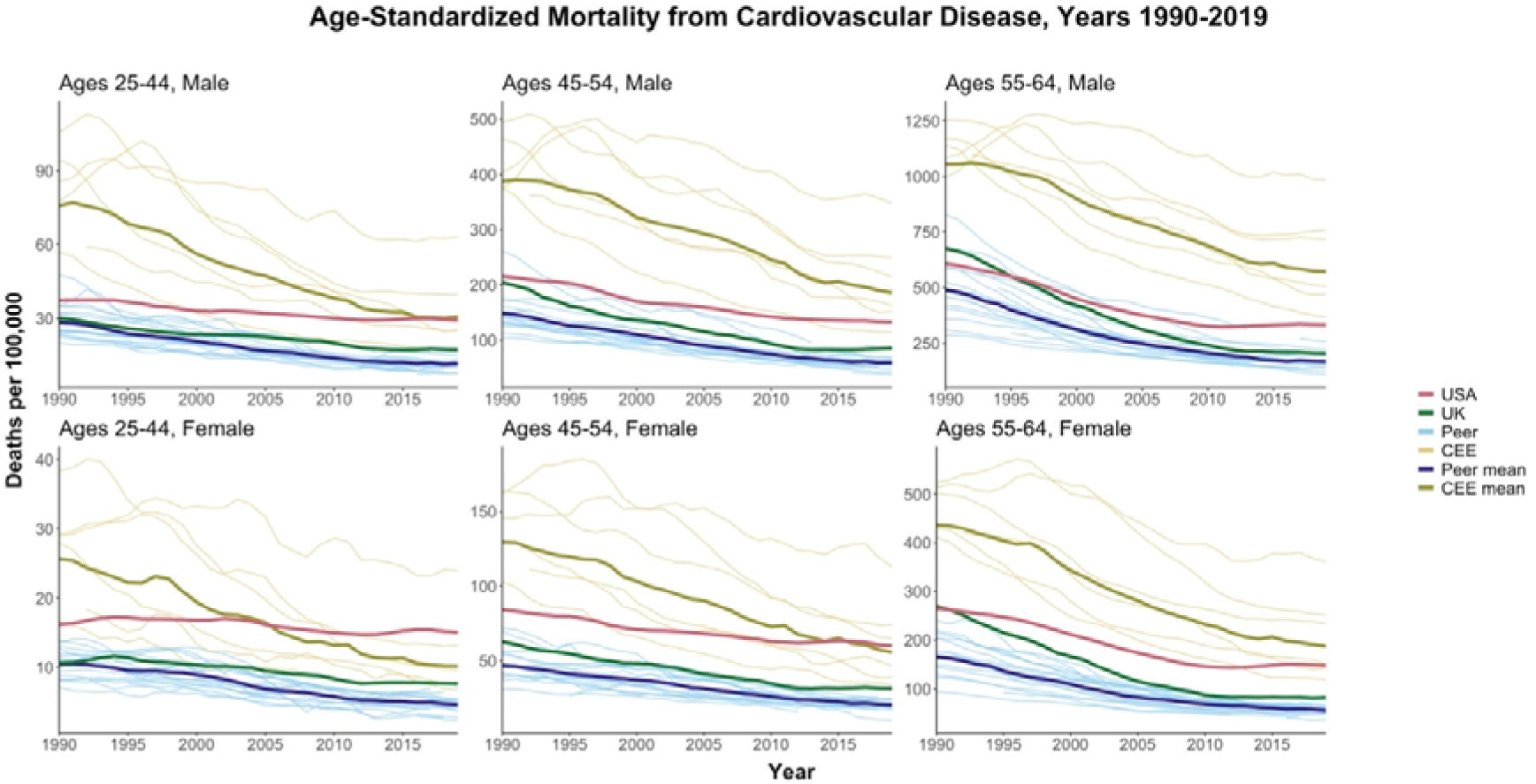
Age-standardized CVD mortality.

**Figure 9.**
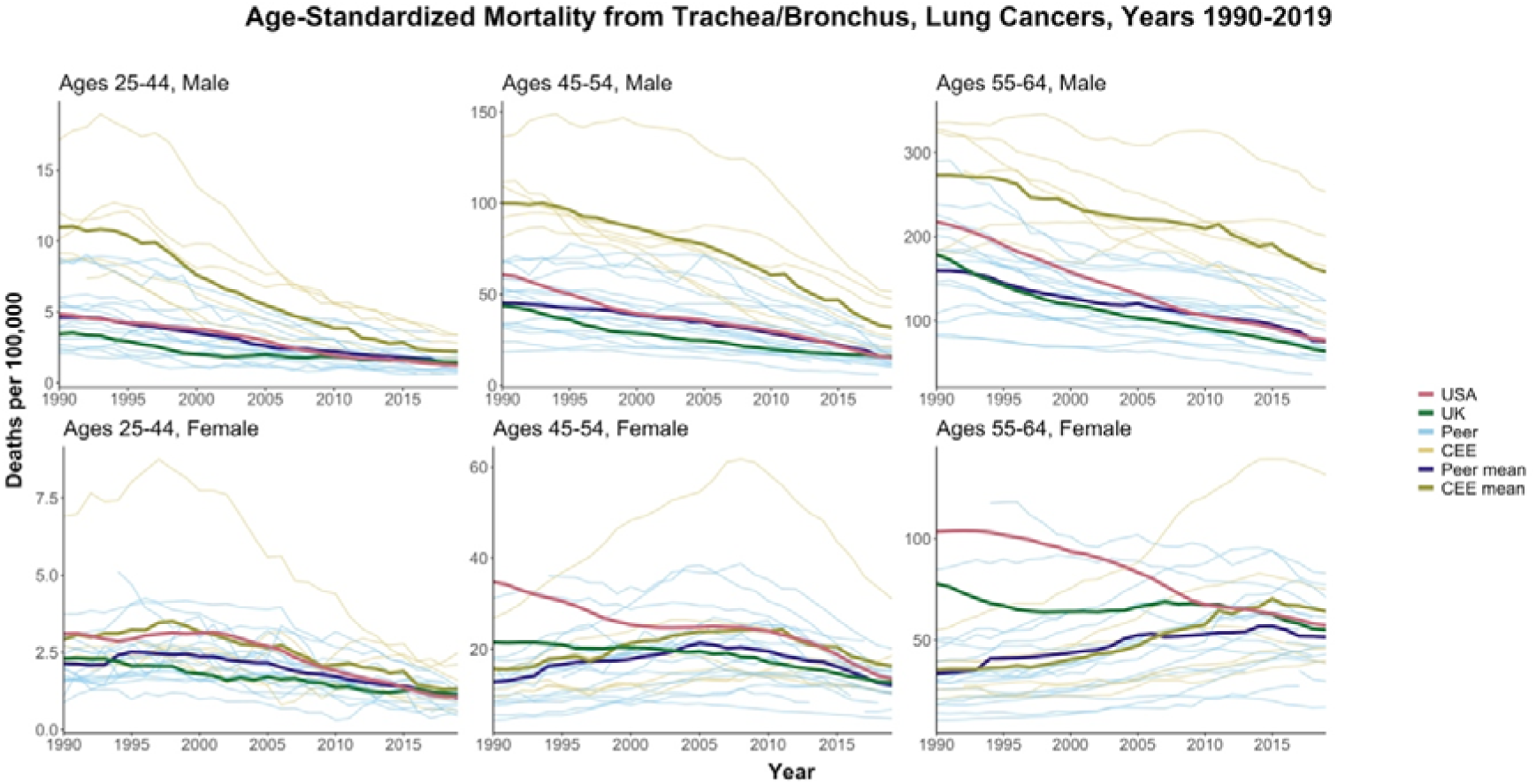
Age-standardized mortality from lung cancer.

The US and UK were not the only high-income countries experiencing rising or stalling midlife mortality. In Canada, all-cause mortality among 25-44 -years-old males and females has been rising since 2013. For males, it increased from 101.3 deaths per 100,000 in 2013 to 118.7 in 2019. For females, mortality increased from 57.4 deaths per 100,000 in 2013 to 62.6 in 2019. Other countries experiencing increases in midlife mortality were Poland (in 25-44- years-old males, from 190.3 per 100,000 in 2016 to 197.7 per 100,000 in 2019) and Sweden (in 25-44-years-old males from 81.3 per 100,000 in 2012 to 84.3 per 100,000 in 2018).

## Discussion

We examined trends in midlife mortality in the US, the UK, and peer high-income countries from 1990-2019. We also compared these trends to countries from Central and Eastern European countries that had suffered from the post-Soviet mortality crisis. Over three decades, most countries experienced declines in all-cause midlife mortality. The US was the notable exception to this pattern, with increases in all-cause mortality seen in some age-sex groups over at least part of this period. Most striking was all-cause mortality for 25–44-year-old females in the US, which was higher in 2019 than it was in 1990. UK midlife mortality remained lower than the US but fell behind compared to high-income peers.

US midlife mortality rates worsened over the period for several causes of death including drug-related, alcohol-related, suicide, metabolic disease, nervous system disease, respiratory disease, and infectious/parasitic diseases. Despite overall declines in the US for homicide, transport accidents, and cardiovascular disease, we documented recent increases or stalling improvements. This led to a deterioration of the relative standing of the US, with mortality rates now surpassing the CEE countries for several causes, a pattern that is more pronounced for US females compared to males, and for younger age groups. The bright spot for the US were cancer deaths, which were more comparable to peer countries.

Our findings are consistent with previous work showing the relative mortality disadvantage of the United States (Masters et al., 2018). Life expectancy is lower in the US than in other high-income countries and its standing has deteriorated substantially since 2010 (Ho, 2013). The recent comprehensive US 2020 National Academy of Sciences report documented diverging life expectancy compared to peers from 1950-2016, and that mortality in ages 18- 60 contributed most to this gap (Harris et al., 2021). In particular, the diverging trends in age-standardized mortality among 25–64-year-olds in the US from 16 peer countries accelerated over time. We confirmed these findings and highlighted that the US disadvantage is most severe for 25-44- and 45–54-year-olds. Besides high-income peers, we found that the US increasingly lags behind Central and Eastern European countries. Higher US mortality was especially noticeable for highly preventable external causes of death including drug overdose, homicide, suicide, and transport accidents. While narratives around “deaths of despair” due to economic and social distress have garnered much attention (Gaydosh et al., 2019, Case & Deaton, 2015, Ruhm, 2019), specific aspects of the US risk environment, especially concerning guns, vehicles, and drugs, merit research and policy attention. In addition to external causes, the US is falling behind due to stalling improvements in deaths due to cardiovascular disease and increases in deaths due to metabolic, respiratory, and nervous system causes. The US disadvantage in these causes suggests longer-term processes that contribute to chronic disease such as high levels of obesity, which are more extreme in the US compared to peer countries (Preston et al., 2018).

There has been less work comparing trends in UK mortality to its peers. The rate of improvement in life expectancy slowed sharply in England and Wales from 2011-2016, relative to OECD peer countries (Leon et al., 2019). Additionally, overall mortality rates in England and Wales in people aged 25–50 years were appreciably higher than peer countries since the 2010s, consistent with our findings. While some have implied that austerity measures after the 2008 financial crisis contributed to mortality deterioration, the empirical grounding for such claims is not strong (Darlington-Pollock & Norman, 2019). Our results suggest that while the UK performs relatively well on external causes such as suicide, homicide, and transport accidents, this is countered by stalling improvements in cardiovascular disease and cancer, and drug deaths are also increasing. Our analyses did not separate the UK into its constituent countries, but previous work has shown much higher midlife mortality in Scotland, with drug mortality in recent years even higher than in the US (Dowd et al., 2022). Overall, our results confirm the divergence of UK mortality from peer countries, which, like the US, is more pronounced in the younger middle-aged groups and in females. While the US mortality crisis has received substantial attention, a greater understanding of the causes behind the stagnation in the UK and prospects for the future is needed.

The mortality disadvantage of the Central and Eastern European countries, although still significant for most common causes of death, diminished significantly in the last twenty years. In the last decade, the mean death rate among CEE countries for some causes such as metabolic, suicides, and transport accidents dropped below the mean rates of the high-income group. The CEE countries, which have historically suffered from high levels of alcohol abuse, are again facing increasing midlife mortality from alcohol-related causes, but without rising drug-related mortality. High-income countries, on the other hand, experienced the inverse: alcohol-related deaths have been relatively stable over the last three decades, but drug-related mortality is gradually rising in several countries (for example Canada, Australia, and Sweden, in addition to the US and UK).

### Limitations

The analyses rely on the comparability of cause-specific mortality data across countries. Although countries included in this analysis have well-developed vital registration systems, some variations in the coding, especially for external causes are likely. Second, harmonization between ICD-9 and ICD-10 coding schemes introduces an additional risk of bias, as the codes are not always directly translatable between classifications. Some ICD-10 codes have no ICD-9 equivalents and vice versa, which meant it was not possible to harmonize codes for alcohol and drug-related mortality. For most other causes of death, the comparability between coding schemes is high (Anderson et al., 2001). Third, our data did not allow examination of variation in mortality by subgroups and regions within countries. Previous studies have suggested large mortality differences between socioeconomic groups (Katikireddi et al., 2017, Masters et al., 2012) and regions in the US and UK (Sheehan et al., 2018, Dowd et al, 2022) so analysis at the national level alone may obscure important trends in certain groups. Where the data are available, within-country analysis of midlife mortality trends by socioeconomic or race/ethnic subgroup could help illuminate potential mechanisms driving the differences in mortality levels and trends.

## Conclusions

Worsening midlife mortality trends in the United States increasingly stand apart not only from high-income peers, but also from former countries of the Soviet Union in Central and Eastern Europe which suffered a mortality crisis in the not-so-distant past. While levels of midlife mortality in the UK are substantially lower than in the US, there are signs of trouble on the horizon relative to the rest of Europe. The more favorable mortality levels in high-income peer countries imply significant room for mortality improvement in both the US and the UK. Comparative work analyzing changes in midlife mortality and policy changes or other trends in risk such as obesity across countries is needed.

## Supporting information

Supplemental Materials-All Figures

Supplemental Materials-Full Results

## Data Availability

All analyses are fully reproducible, the data and code can be accessed here (https://github.com/OxfordDemSci/International-Mortality).

## Funding Acknowledgement

The authors acknowledge funding support from the European Research Council (ERC-2021- CoG-101002587) and The Leverhulme Trust (Large Centre Grant). The funders played no direct role on this research.

## References

Anderson, R. N., Miniño, A. M., Hoyert, D. L., & Rosenberg, H. M. (2001). Comparability of cause of death between ICD-9 and ICD-10: Preliminary estimates. *National Vital Statistics Reports: From the Centers for Disease Control and Prevention, National Center for Health Statistics*, National Vital Statistics System, 49(2), 1–32.

BMJ. (2019, March 19). Danny Dorling: Austerity bites—falling life expectancy in the UK. The BMJ. https://blogs.bmj.com/bmj/2019/03/19/danny-dorling/

Case, A., & Deaton, A. (2015). Rising morbidity and mortality in midlife among white non-Hispanic Americans in the 21st century. Proceedings of the National Academy of Sciences, 112(49), 15078–15083. https://doi.org/10.1073/pnas.1518393112

Case, A., & Deaton, A. (2017). Mortality and morbidity in the 21st century. Brookings Papers on Economic Activity, 2017, 397–476.

Darlington-Pollock, F., & Norman, P. (2019). Stalling life expectancy and increased mortality in working ages deserve urgent attention. The Lancet Public Health, 4(11), e543–e544. https://doi.org/10.1016/S2468-2667(19)30207-5

Dowd, J. B., Angus, C., Zajacova, A., & Tilstra, A. M. (2022). Midlife “Deaths of Despair” Trends in the US, Canada, and UK, 2001-2019: Is the US an Anomaly? MedRxiv, 2022.10.10.22280916. https://doi.org/10.1101/2022.10.10.22280916

Gaydosh, L., Hummer, R. A., Hargrove, T. W., Halpern, C. T., Hussey, J. M., Whitsel, E. A., Dole, N., & Harris, K. M. (2019). The Depths of Despair Among US Adults Entering Midlife. American Journal of Public Health, 109(5), 774–780. https://doi.org/10.2105/AJPH.2019.305002

Harris, K. M., Majmundar, M. K., & Becker, T. (Eds.). (2021). High and Rising Mortality Rates Among Working-Age Adults. National Academies Press. https://doi.org/10.17226/25976

Ho, J. Y. (2013). Mortality Under Age 50 Accounts For Much Of The Fact That US Life Expectancy Lags That Of Other High-Income Countries. Health Affairs, 32(3), 459– 467.

Ho, J. Y. (2019). The Contemporary American Drug Overdose Epidemic in International Perspective. Population and Development Review, 45(1), 7–40. https://doi.org/10.1111/padr.12228

Ho, J. Y., & Hendi, A. S. (2018). Recent trends in life expectancy across high income countries: Retrospective observational study. BMJ, 362, k2562. https://doi.org/10.1136/bmj.k2562

Katikireddi, S. V., Leyland, A. H., McKee, M., Ralston, K., & Stuckler, D. (2017). Patterns of mortality by occupation in the UK, 1991–2011: A comparative analysis of linked census and mortality records. The Lancet Public Health, 2(11), e501–e512. https://doi.org/10.1016/S2468-2667(17)30193-7

Leon, D. A., Jdanov, D. A., & Shkolnikov, V. M. (2019). Trends in life expectancy and age-specific mortality in England and Wales, 1970–2016, in comparison with a set of 22 high-income countries: An analysis of vital statistics data. The Lancet Public Health, 4(11), e575–e582. https://doi.org/10.1016/S2468-2667(19)30177-X

Marmot Indicators 2017—Institute of Health Equity Briefing. (n.d.). Institute of Health Equity. Retrieved 24 March 2023, from https://www.instituteofhealthequity.org/resources-reports/marmot-indicators-2017-institute-of-health-equity-briefing

Masters, R. K., Hummer, R. A., & Powers, D. A. (2012). Educational Differences in U.S. Adult Mortality: A Cohort Perspective. American Sociological Review, 77(4), 548–572. https://doi.org/10.1177/0003122412451019

Masters, R. K., Tilstra, A. M., & Simon, D. H. (2018). Explaining recent mortality trends among younger and middle-aged White Americans. International Journal of Epidemiology, 47(1), 81–88. https://doi.org/10.1093/ije/dyx127

McCartney, G., McMaster, R., Popham, F., Dundas, R., & Walsh, D. (2022). Is austerity a cause of slower improvements in mortality in high-income countries? A panel analysis. Social Science & Medicine (1982), *313*, 115397. https://doi.org/10.1016/j.socscimed.2022.115397

Peters, S. A. E., Huxley, R. R., & Woodward, M. (2014). Do smoking habits differ between women and men in contemporary Western populations? Evidence from half a million people in the UK Biobank study. BMJ Open, 4(12), e005663. https://doi.org/10.1136/bmjopen-2014-005663

Preston, S. H., Vierboom, Y. C., & Stokes, A. (2018). The role of obesity in exceptionally slow US mortality improvement. Proceedings of the National Academy of Sciences, 115(5), 957–961. https://doi.org/10.1073/pnas.1716802115

Raleigh, V. S. (2018). Stalling life expectancy in the UK. BMJ, 362, k4050. https://doi.org/10.1136/bmj.k4050

Ruhm, C. J. (2019). Drivers of the fatal drug epidemic. Journal of Health Economics, 64, 25–42. https://doi.org/10.1016/j.jhealeco.2019.01.001

Scheiring, G., Azarova, A., Irdam, D., Doniec, K., McKee, M., & King, L. (2021). Deindustrialization and the Postsocialist Mortality Crisis. PERI Working Papers. https://doi.org/10.7275/27135476

Sheehan, C., Montez, J. K., & Sasson, I. (2018). Does the Functional Form of the Association Between Education and Mortality Differ by U.S. Region? Biodemography and Social Biology, 64(1), 63–81. https://doi.org/10.1080/19485565.2018.1468239

Vaupel, J. W., Villavicencio, F., & Bergeron-Boucher, M.-P. (2021). Demographic perspectives on the rise of longevity. Proceedings of the National Academy of Sciences, 118(9), e2019536118. https://doi.org/10.1073/pnas.2019536118

Woolf, S., & Schoomaker, H. (2019). LIFE EXPECTANCY AND MORTALITY RATES IN THE UNITED STATES, 1959-2017. JAMA, 322(20), 1996–2016. https://doi.org/10.1001/jama.2019.16932

