## Supplemental Materials-All Figures for "US Exceptionalism? International Trends in Midlife Mortality"

#### Table of Contents

***Appendix 1*** ***1***

Table 1 ICD9-ICD10 harmonization 2

Table 2 Availability of countries-years in mortality data 4

Table 3 Availability of countries-years in population data 5

Table 4 Countries-years with 4-digit ICD10 codes unavailable 7

***Appendix 2*** ***8***

Figure 1 Age−Standardized Mortality from Infectious and Parasitic Diseases, Years 1990−2019 9

Figure 2 Age−Standardized Mortality from HIV/AIDS, Years 1990−2019 10

Figure 3 Age−Standardized Mortality from Respiratory Diseases, Years 1990−2019 11

Figure 4 Age−Standardized Mortality from Trachea/Bronchus, Lung Cancers, Years 1990−2019 12

Figure 5 Age−Standardized Mortality from All Other Cancers, Years 1990−2019 13

Figure 6 Age−Standardized Mortality from Nervous System Diseases, Years 1990−2019 14

Figure 7 Age−Standardized Mortality from Metabolic Diseases, Years 1990−2019 15

Figure 8 Age−Standardized Mortality from Cardiovascular Disease, Years 1990−2019 16

Figure 9 Age−Standardized Mortality from Suicide, Years 1990−2019 17

Figure 10 Age−Standardized Mortality from Homicide, Years 1990−2019 18

Figure 11 Age−Standardized Mortality from Transport Accidents, Years 1990−2019 19

Figure 12 Age−Standardized Mortality from Other External Causes, Years 1990−2019 20

Figure 13 Age−Standardized Mortality from All Other Causes, Years 1990−2019 21

Figure 14 Age−Standardized Mortality from All Causes, Years 1990−2019 22

Figure 15 Age−Standardized Mortality from Alcohol-Related Causes, Years 2000−2019 23

Figure 16 Age−Standardized Mortality from Drug-Related Causes, Years 2000−2019 24

***Appendix 3*** ***25***

Figure 1 Percent Change in Age−Standardized Mortality from the Baseline Year (1990), Infectious and Parasitic Diseases 26

Figure 2 Percent Change in Age−Standardized Mortality from the Baseline Year (1990), HIV/AIDS 27

Figure 3 Percent Change in Age−Standardized Mortality from the Baseline Year (1990), Respiratory Diseases 28

Figure 4 Percent Change in Age−Standardized Mortality from the Baseline Year (1990), Trachea/Bronchus, Lung Cancers 29

Figure 5 Percent Change in Age−Standardized Mortality from the Baseline Year (1990), All Other Cancers 30

Figure 6 Percent Change in Age−Standardized Mortality from the Baseline Year (1990), Nervous System 31

Figures 1-96 Three-year moving average mortality figures by country, cause, sex and age.........49-144

### Appendix 1

Table 1. ICD9 BTL - ICD10 harmonization.

| CATEGORIES | ICD9 BTL | ICD10 CODES | NOTES |
|---|---|---|---|
| **1. Infectious and parasitic (excluding HIV/AIDS)** | B01 - B07 | A00 - A99, B00 - B19, B25 -B99 | |
| **2. HIV/AIDS** | B184, B185 | B20 - B24 | |
| **3. Respiratory System** | B31 - B32 | J00 - J98 | |
| **4. All cancers (excluding trachea/bronchus, lung and liver cancers)** | B08 - B09, B10, B100, B109, B11 - B17 | C00 - D48, excluding C33, C34 | |
| **5. Trachea/ bronchus, lung cancers** | B101 | C33 -- C34 | |
| **6. Nervous system** | B22 | G00 - G98 | |
| **7. Endocrine, Nutritional & Metabolic** | B180-B183, B189, B19 | E00 - E88 | |
| **8. Circulatory system** | B25 – B30 | I00 - I99 | |
| **9. Suicide** | B54 | X60–X84, Y87.0 | |
| **10. Drug Poisoning** | N/A in ICD9 BTL coding scheme | F11-F16, F18-F19, X40–X44, X85, Y10–Y14 | Due to ICD9 BTL scheme limitations this category is only kept for country-years where ICD10 scheme is available. For country-years with ICD9 BTL scheme drug-related deaths are set to 'other external causes' category. |

| | | | |
|---|---|---|---|
| **11. Alcohol-induced** | N/A in ICD9 BTL coding scheme | K70, K73-74, F10, X45, Y15 | Due to ICD9 BTL scheme limitations this category is only kept for country-years where ICD10 scheme is available. For country-years with ICD9 BTL scheme drug-related deaths are set to 'other external causes' category. |
| **12. Homicide** | B55 | X86 - X99, Y00 - Y09, Y87.1 | |
| **13. Transport accidents** | B47 | V01–V99, Y85 | |
| **14. Other external causes** | B480-B482, B49, B50, B51, B52, B53, B56 | W00–W99, X00–X39, X46–X59, Y16–Y36, Y40–Y84, Y86, Y87.2, Y88, Y89 | |

Table 2. Availability of countries-years in mortality data as of 1st June 2023.
Source: https://www.who.int/data/data-collection-tools/who-mortality-database

| Country Code | Name | Year | List | Icd | Note |
|---|---|---|---|---|---|
| 2090 | Canada | 1979-1999<br>2000-2019 | 09B<br>104 | Icd9<br>Icd10 | |
| 2450 | United States of America | 1979-1998<br>1999-2019 | 09B<br>104 | Icd9<br>Icd10 | |
| 3160 | Japan | 1979-1994<br>1995-2017<br>2018-2019 | 09B<br>104<br>103 | Icd9<br>Icd10<br>Icd10 | |
| 4010 | Austria | 1980-2001<br>2002-2019 | 09B<br>104 | Icd9<br>Icd10 | |
| 4020 | Belgium | 1979-1997<br>1998-1999<br>2000-2018 | 09B<br>103<br>104 | Icd9<br>Icd10<br>Icd10 | |
| 4050 | Denmark | 1994-2019 | 104 | Icd10 | |
| 4070 | Finland | 1987-1994<br>1996-2019 | 09B<br>103 | Icd9<br>Icd10 | |
| 4080 | France | 1979-1999<br>2000-2017 | 09B<br>104 | Icd9<br>Icd10 | |
| 4085 | Germany | 1990-1997<br>1998-2019 | 09B<br>104 | Icd9<br>Icd10 | |
| 4180 | Italy | 1979-2002<br>2003-2019 | 09B<br>104 | Icd9<br>Icd10 | |
| 4210 | Netherlands | 1979-1995<br>1996-1999<br>2000-2019 | 09B<br>10M<br>104 | Icd9<br>Icd10<br>Icd10 | |
| 4220 | Norway | 1986-1995<br>1996-2016 | 09B<br>104 | Icd9<br>Icd10 | |
| 4240 | Portugal | 1980-2001<br>2002-2003<br>2004-2005<br>2007-2019 | 09B<br>104<br>UE1<br>104 | Icd9<br>Icd10<br>Icd10<br>Icd10 | Mortality data for 2004-2005 are not disaggregated by age; mortality data are not available for 2006 |
| 4280 | Spain | 1980-1998<br>1999-2019 | 09B<br>104 | Icd9<br>Icd10 | |
| 4290 | Sweden | 1987-1996<br>1997-1998<br>1999-2018 | 09B<br>10M<br>104 | Icd9<br>Icd10<br>Icd10 | |
| 4300 | Switzerland | 1995-2019 | 104 | Icd10 | |
| 4308 | United Kingdom | 1979-1999<br>2001-2019 | 09B<br>104 | Icd9<br>Icd10 | Mortality data are not available for 2000 |

| | | | | | |
|---|---|---|---|---|---|
| 5020 | Australia | 1979-1997<br>1998-2019 | 09B<br>104 | Icd9<br>Icd10 | Mortality data are not available for 2005 |
| 4030 | Bulgaria | 1980-2004<br>2005-2019 | 09B<br>104 | Icd9<br>Icd10 | |
| 4045 | Czech Republic | 1986-1993<br>1994-2019 | 09B<br>104 | Icd9<br>Icd10 | |
| 4150 | Hungary | 1979-1995<br>1996-2019 | 09B<br>104 | Icd9<br>Icd10 | |
| 4230<br>- | Poland<br>- | 1980-1996<br>1999-2019 | 09B<br>104 | Icd9<br>Icd10 | No data for 1997-1998 |
| 4270 | Romania | 1980-1998<br>1999-2019 | 09B<br>104 | Icd9<br>Icd10 | |
| 4274 | Slovakia | 1992-1993<br>1994-2009<br>2010-2019 | 09B<br>103<br>104 | Icd9<br>Icd10<br>Icd10 | No data for 1990, 2011, 2015 |
| 4276 | Slovenia | 1985-1996<br>1997-2019 | 09B<br>103 | Icd9<br>Icd10 | |

Table 3. Availability of countries-years (1990-2019) in population data as of 1st June 2023.
Source: https://www.who.int/data/data-collection-tools/who-mortality-database

| Country Code | Name | Year | Note | Link |
|---|---|---|---|---|
| 2090 | Canada | 1990-2005 | Data for 2006-2019 are unavailable; extract from Statistics Canada instead | https://www150.statcan.gc.ca/t1/tbl1/en/cv.action?pid=1710000501 |
| 2450 | United States of America | 1990-2007 | Data for 2008-2019 are unavailable; extract from US Census Bureau instead | https://www.census.gov/programs-surveys/popest/technical-documentation/research/evaluation-estimates/2020-evaluation-estimates/2010s-national-detail.html |
| 3160 | Japan | 1990-2019 | | |
| 4010 | Austria | 1990-2019 | | |
| 4020 | Belgium | 1990-2019 | | |
| 4050 | Denmark | 1990-2019 | | |
| 4070 | Finland | 1990-2019 | | |

| | | | | |
|---|---|---|---|---|
| 4080 | France | 1990-2014,2017 | Data for 2015-2016 are unavailable; extract from France National Institute of Statistics and Economic Studies instead | https://www.insee.fr/en/statistiques/pyramide/3312960/xls/pyramides-des-ages_bilan-demo_2019.xls |
| 4085 | Germany | 1990-2019 | | |
| 4180 | Italy | 1990-2019 | | |
| 4210 | Netherlands | 1990-2019 | | |
| 4220 | Norway | 1990-2019 | | |
| 4240 | Portugal | 1990-2018 | | |
| 4280 | Spain | 1990-2019 | | |
| 4290 | Sweden | 1990-2018 | | |
| 4300 | Switzerland | 1990-2019 | | |
| 4308 | United Kingdom | 1990-2019 | | |
| 5020 | Australia | 1990-2019 | | |
| 4030 | Bulgaria | 1990-2019 | | |
| 4045 | Czech Republic | 1990-2019 | | |
| 4150 | Hungary | 1990-2019 | | |
| 4230 | Poland | 1990-2019 | | |
| 4270 | Romania | 1990-2019 | | |
| 4274 | Slovakia | 1990-2019 | Year 2015 missing | |
| 4276 | Slovenia | 1990-2019 | | |

Table 4. Countries-years for which ICD10 4-digit codes are unavailable.

| country | year |
|---|---|
| Japan | 2018 |
| Bulgaria | 2008 |
| Bulgaria | 2009 |
| Bulgaria | 2010 |
| Bulgaria | 2011 |
| Bulgaria | 2012 |
| Finland | 2008 |
| Finland | 2009 |
| Finland | 2010 |
| Finland | 2011 |
| Finland | 2012 |
| Finland | 2013 |
| Finland | 2014 |
| Finland | 2015 |
| Finland | 2016 |
| Finland | 2017 |
| Finland | 2018 |
| Slovakia | 2008 |
| Slovakia | 2009 |
| Slovenia | 2008 |
| Slovenia | 2009 |
| Slovenia | 2010 |
| Slovenia | 2011 |
| Slovenia | 2012 |
| Slovenia | 2013 |
| Slovenia | 2014 |
| Slovenia | 2015 |
| Slovenia | 2016 |
| Slovenia | 2017 |
| Slovenia | 2018 |
| Slovenia | 2019 |
| Slovenia | 2020 |

### Appendix 2

### Figure 1 Age−Standardized Mortality from Infectious and Parasitic Diseases, Years 1990−2019

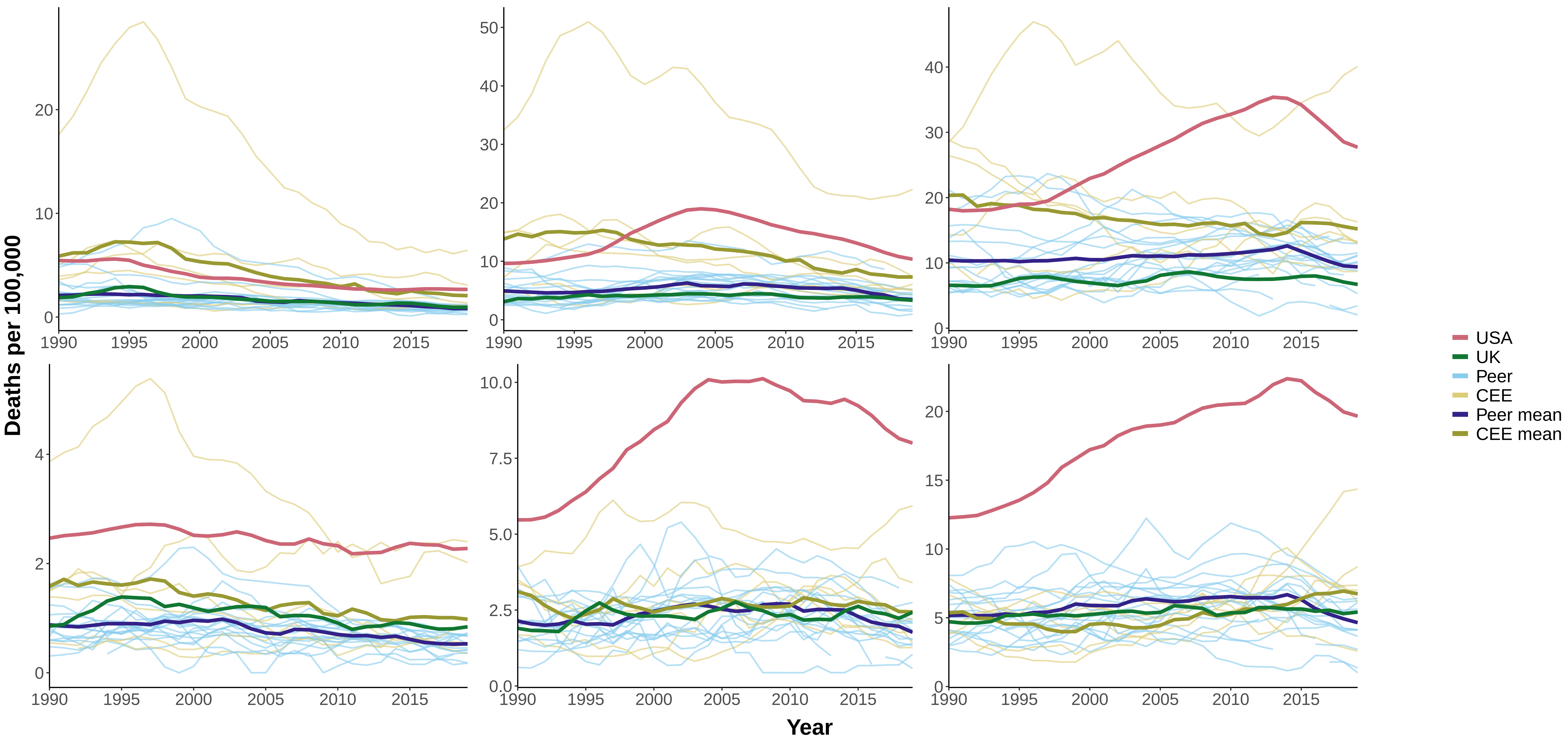

Figure 2 Age−Standardized Mortality from HIV/AIDS, Years 1990−2019

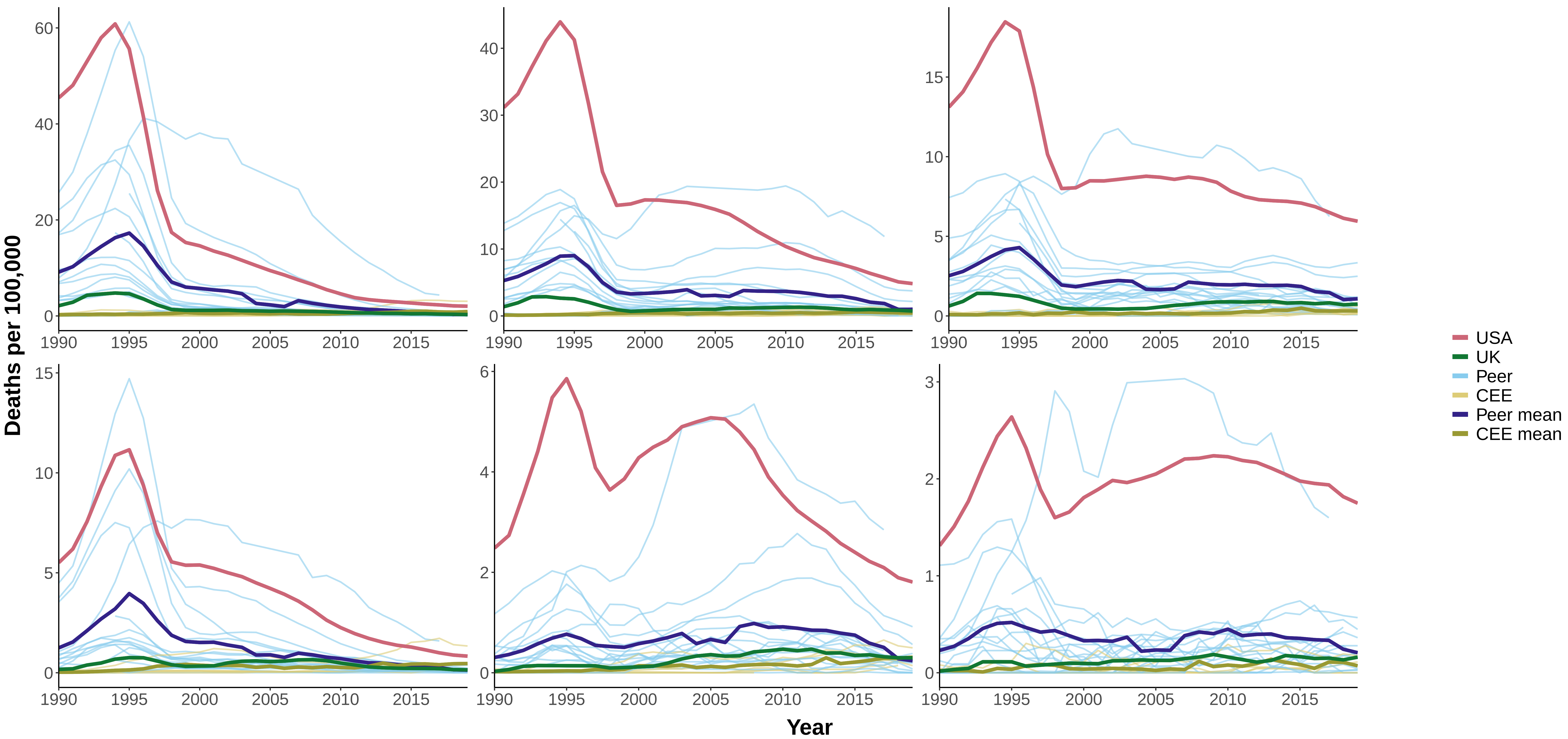

### Figure 3 Age−Standardized Mortality from Respiratory Diseases, Years 1990−2019

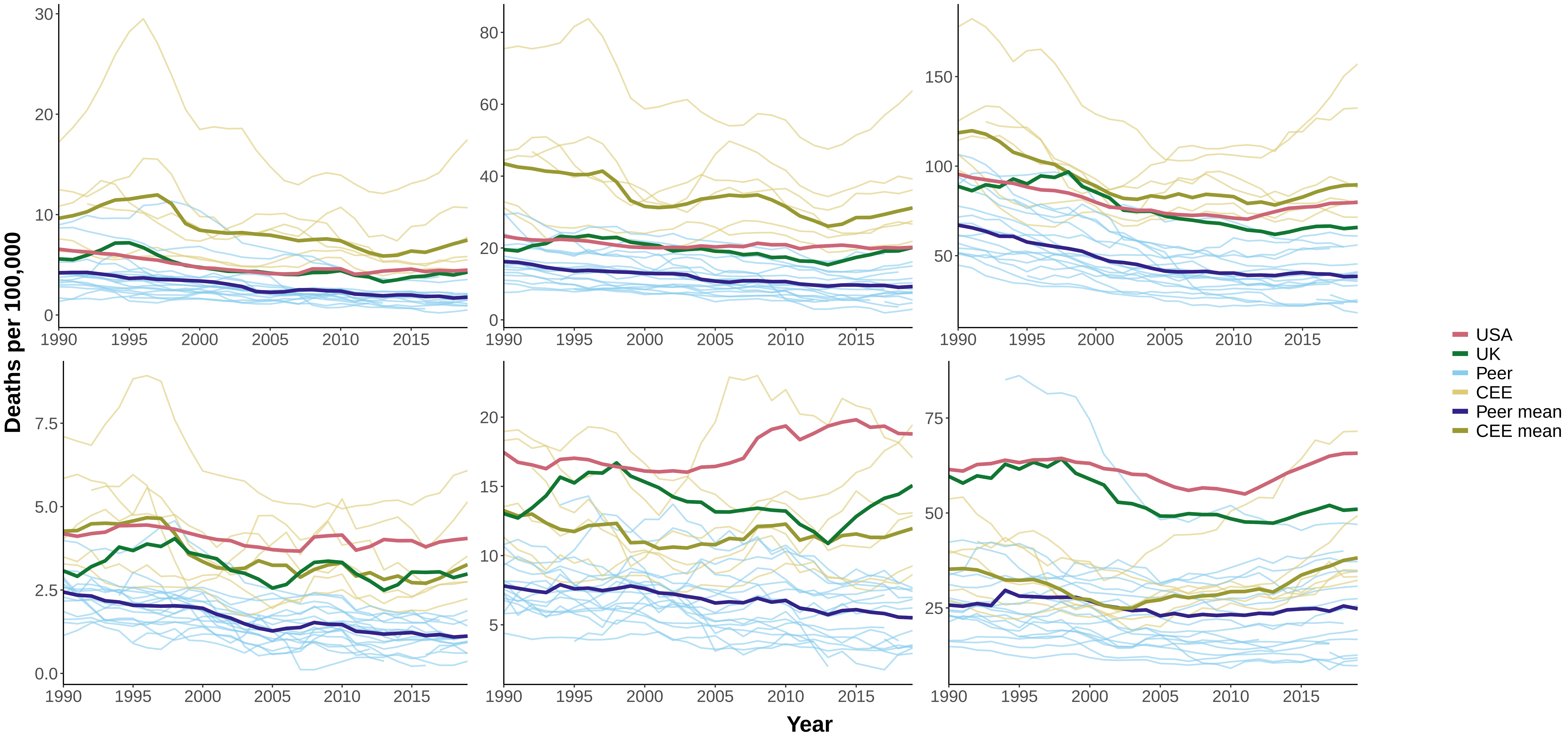

### Figure 4 Age−Standardized Mortality from Trachea/Bronchus, Lung Cancers, Years 1990−2019

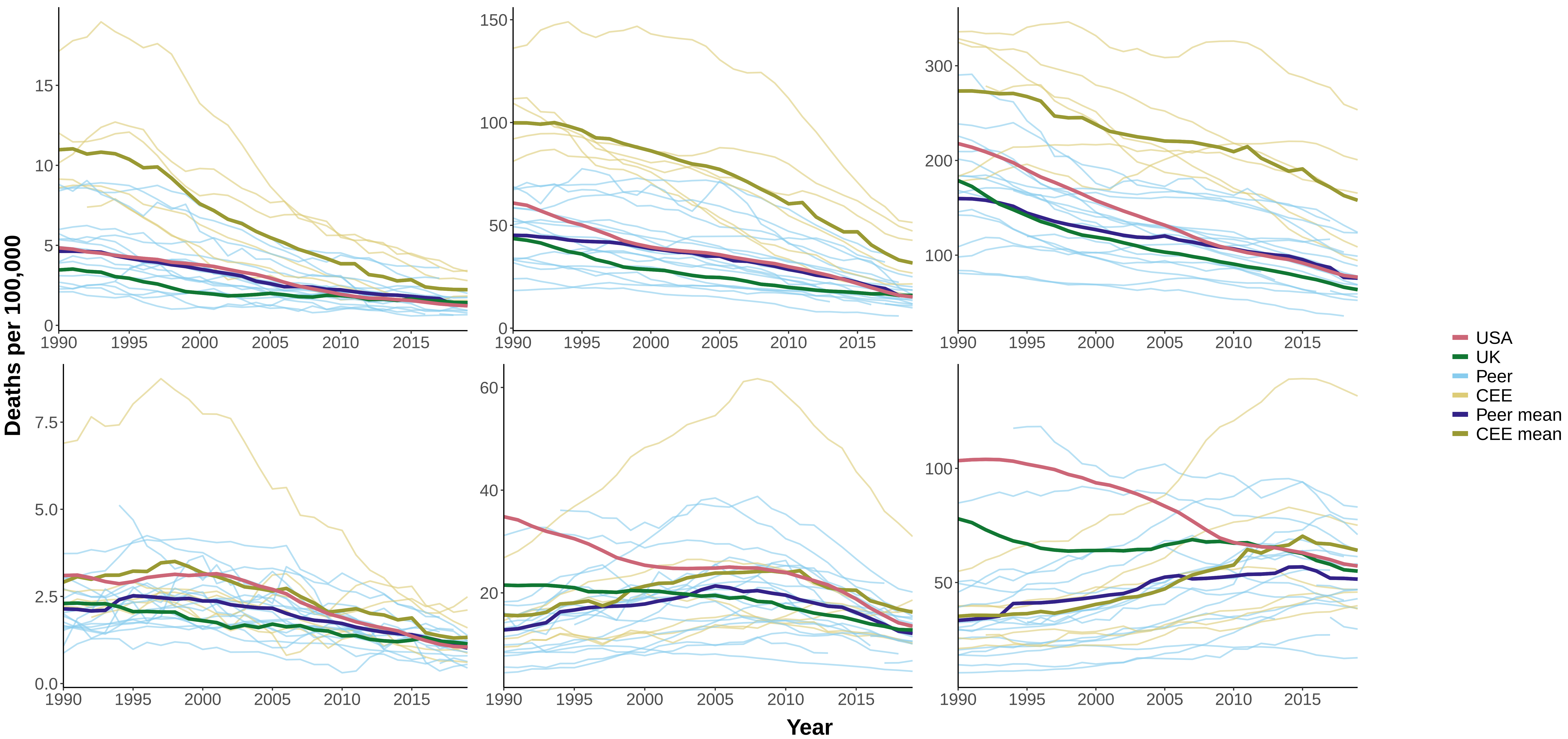

### Figure 5 Age−Standardized Mortality from All Other Cancers, Years 1990−2019

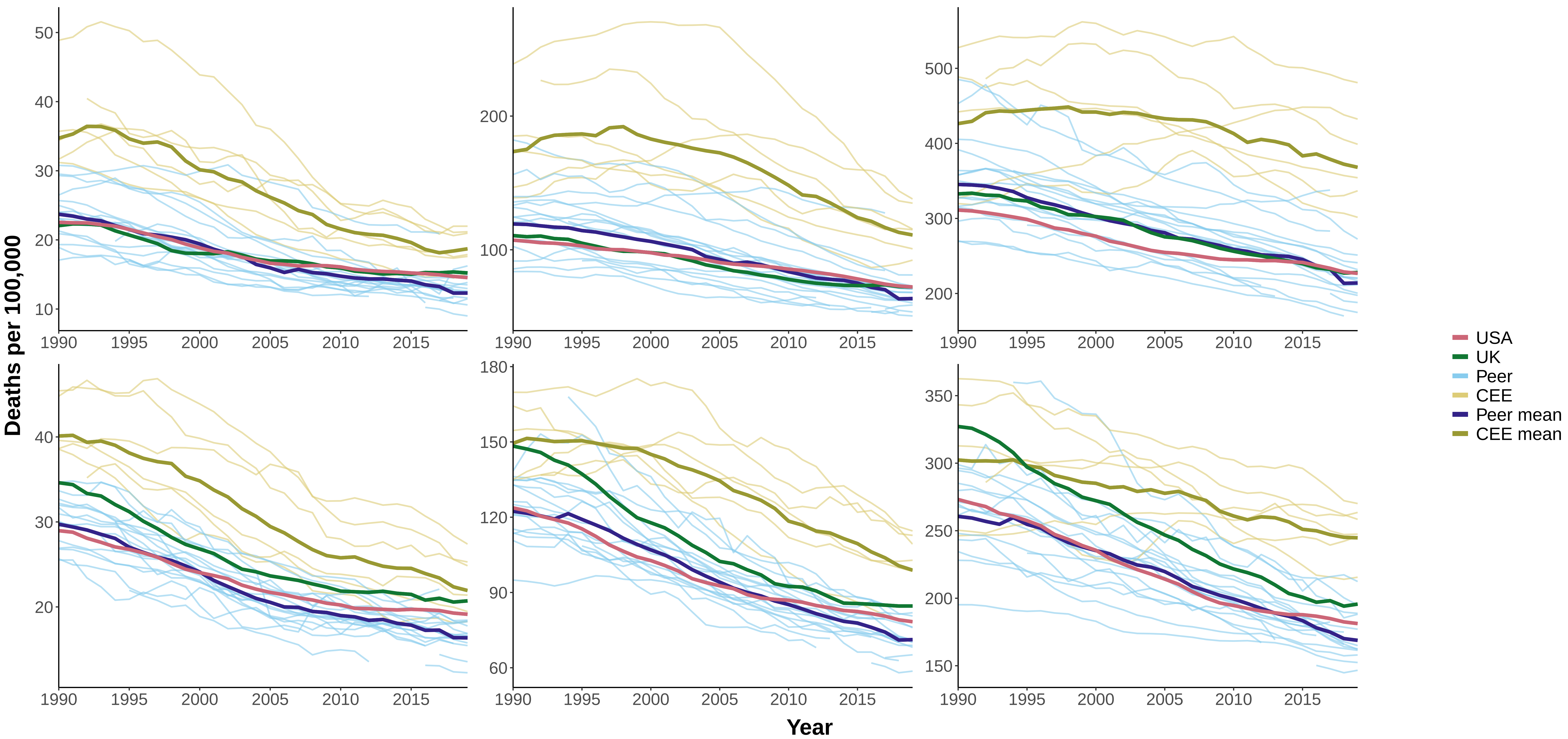

### Figure 6 Age−Standardized Mortality from Nervous System Diseases, Years 1990−2019

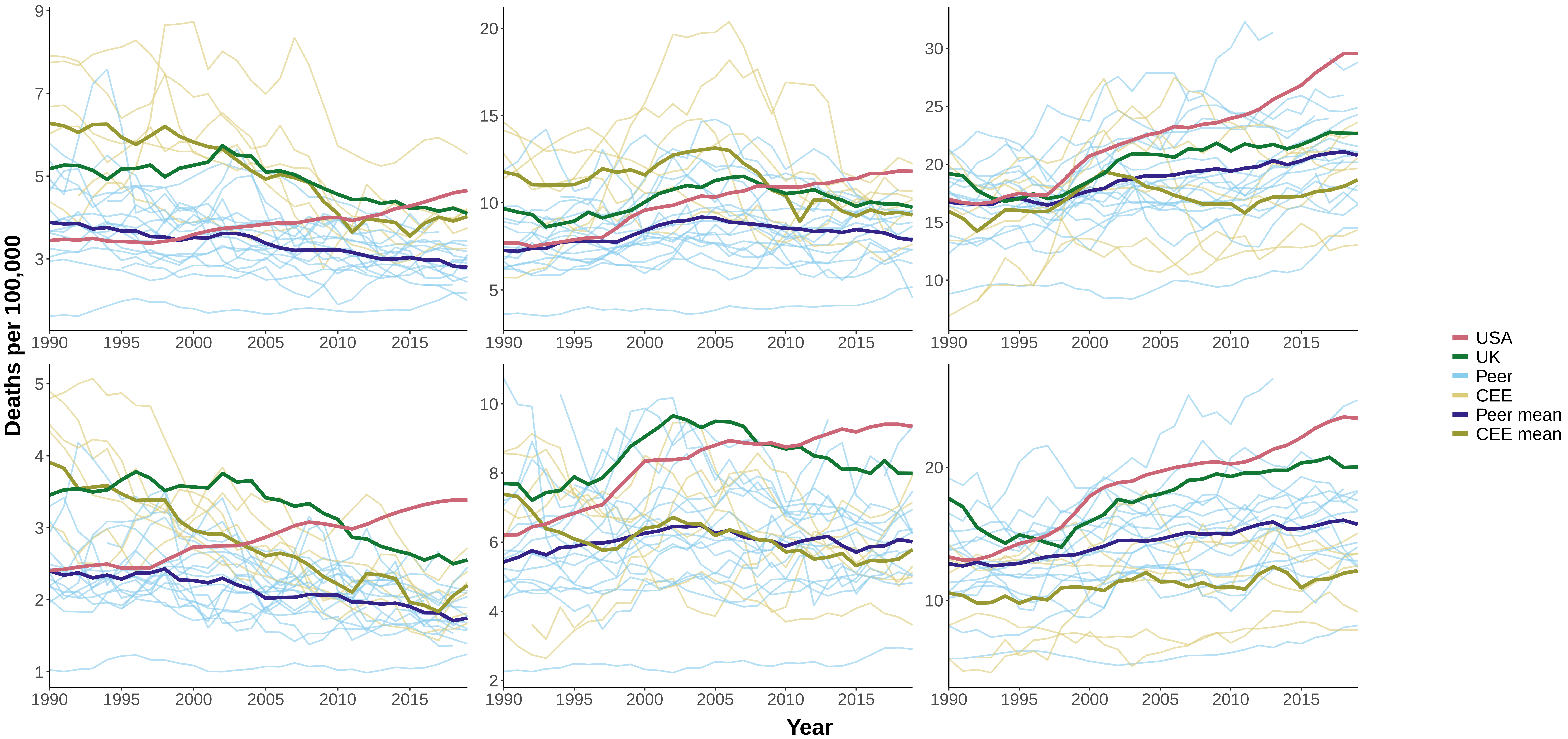

#### Figure 7 Age−Standardized Mortality from Metabolic Diseases, Years 1990−2019

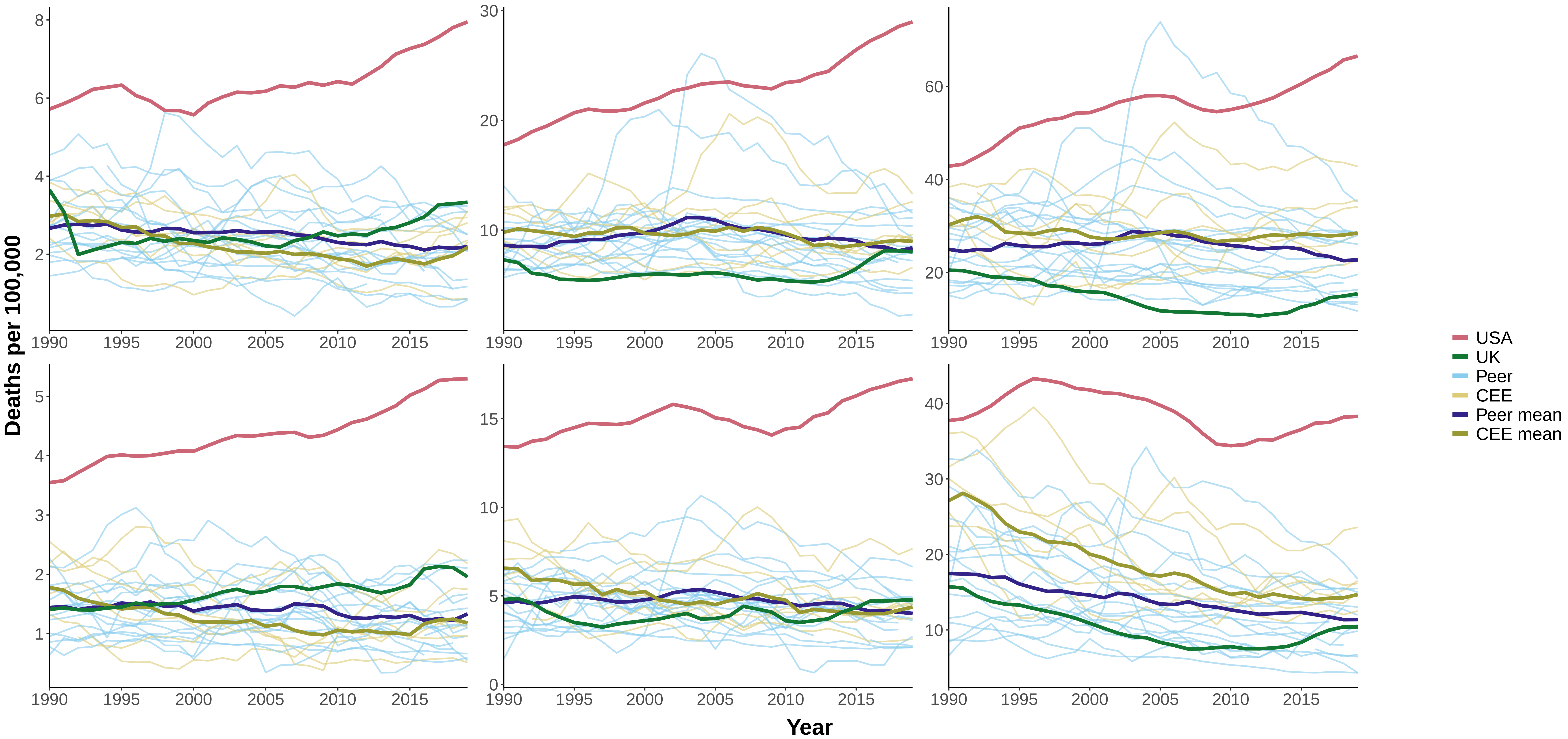

### Figure 8 Age−Standardized Mortality from Cardiovascular Disease, Years 1990−2019

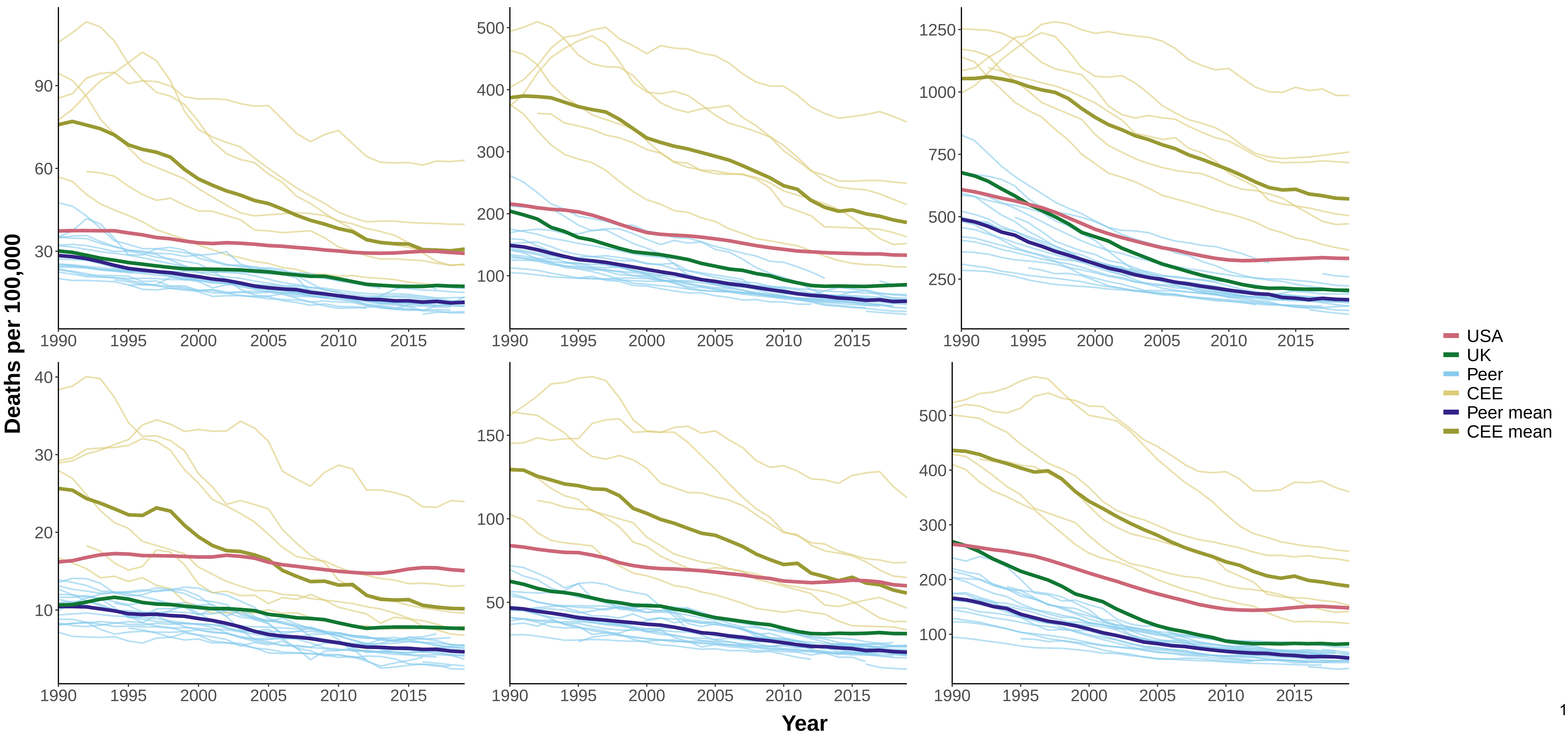

### Figure 9 Age−Standardized Mortality from Suicide, Years 1990−2019

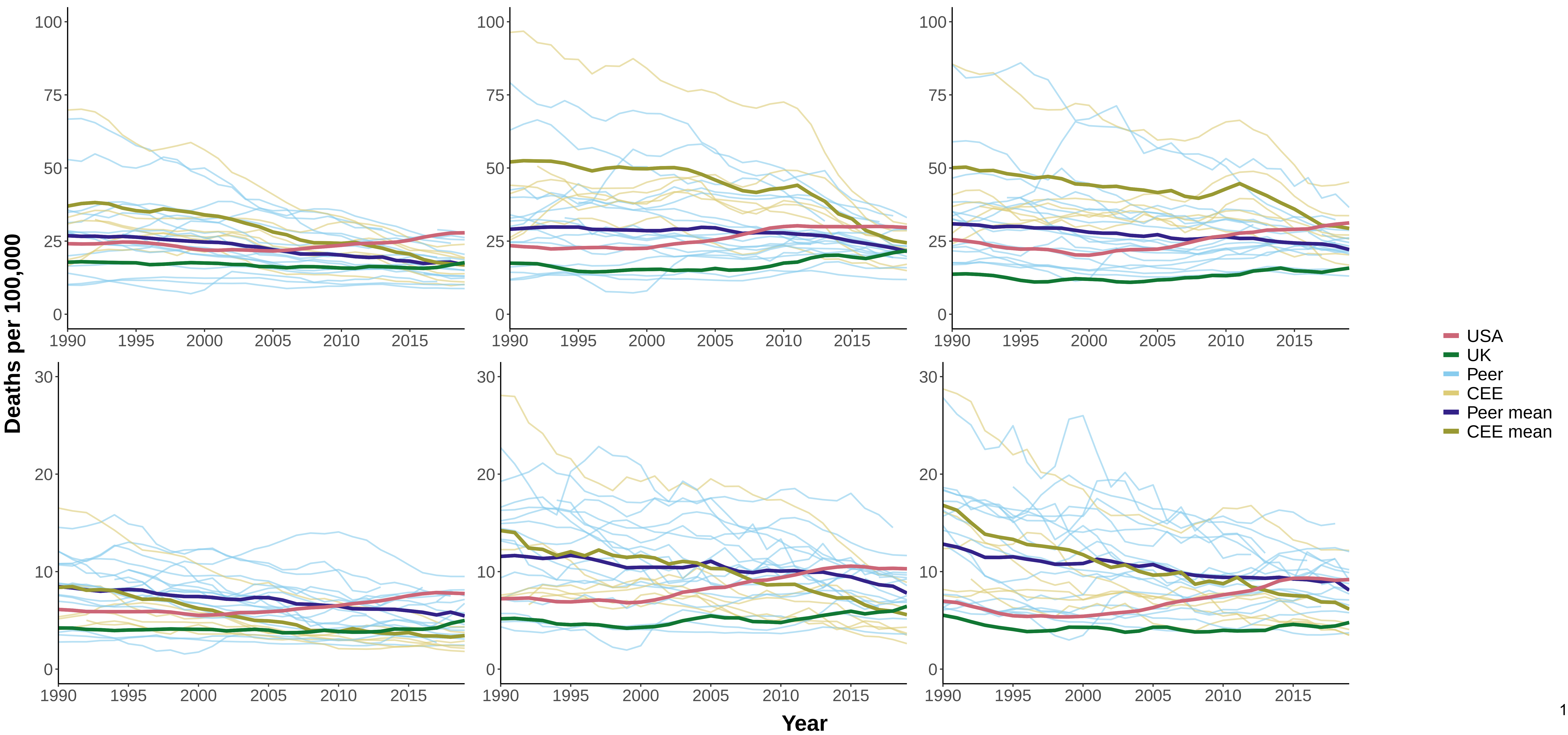

### Figure 10 Age−Standardized Mortality from Homicide, Years 1990−2019

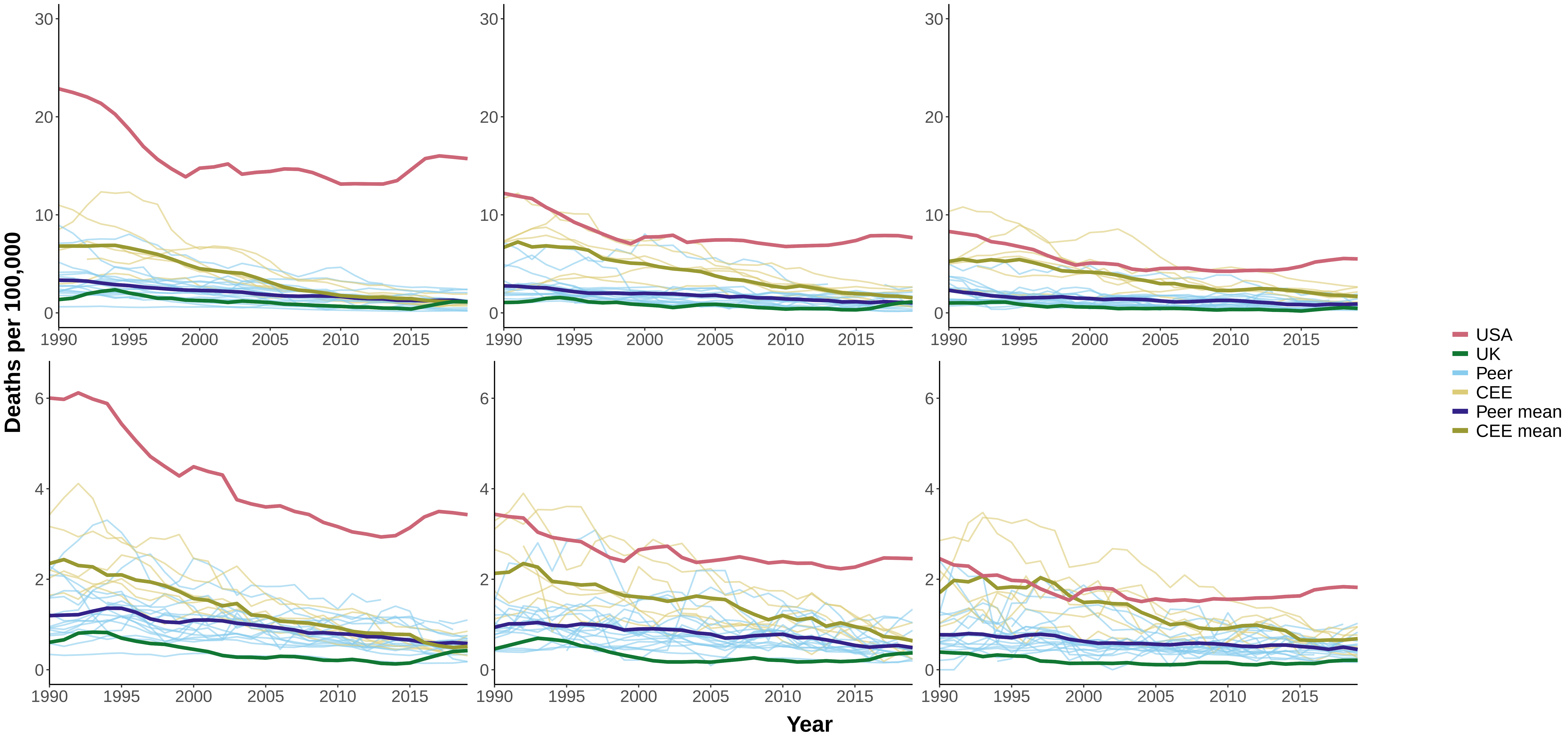

**Figure 11 Age−Standardized Mortality from Transport Accidents, Years 1990−2019**

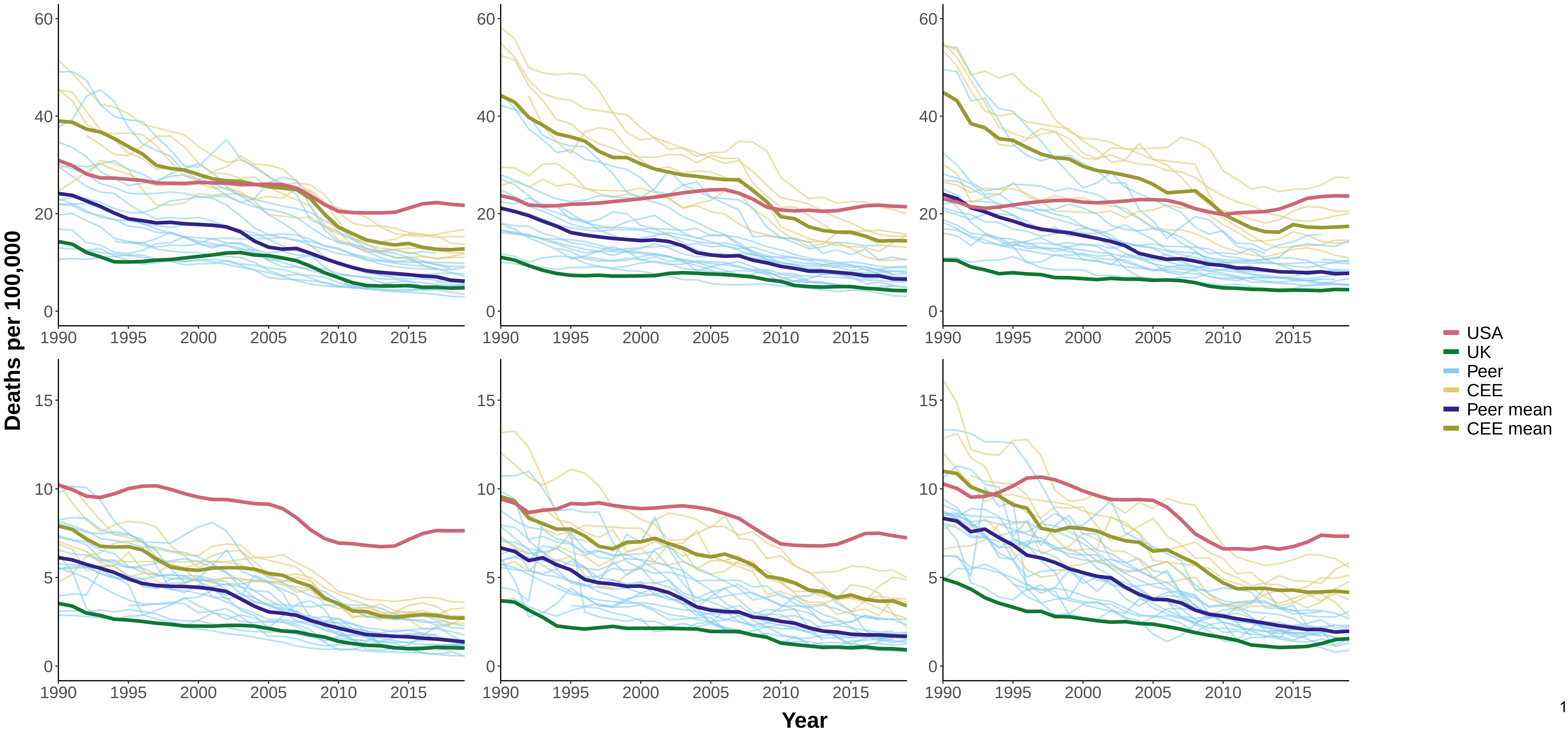

### Figure 12 Age−Standardized Mortality from Other External Causes, Years 1990−2019

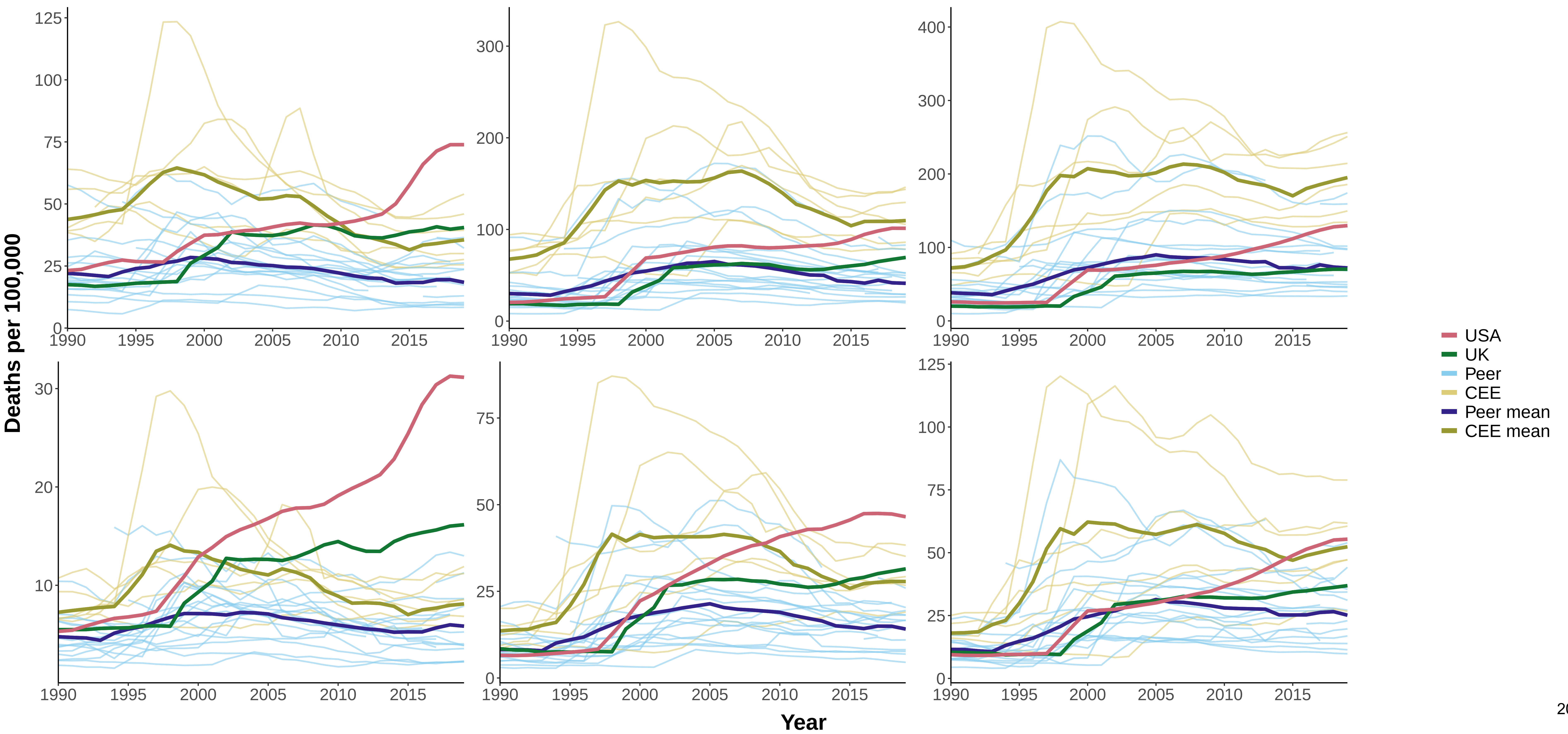

### Figure 13 Age−Standardized Mortality from All Other Causes, Years 1990−2019

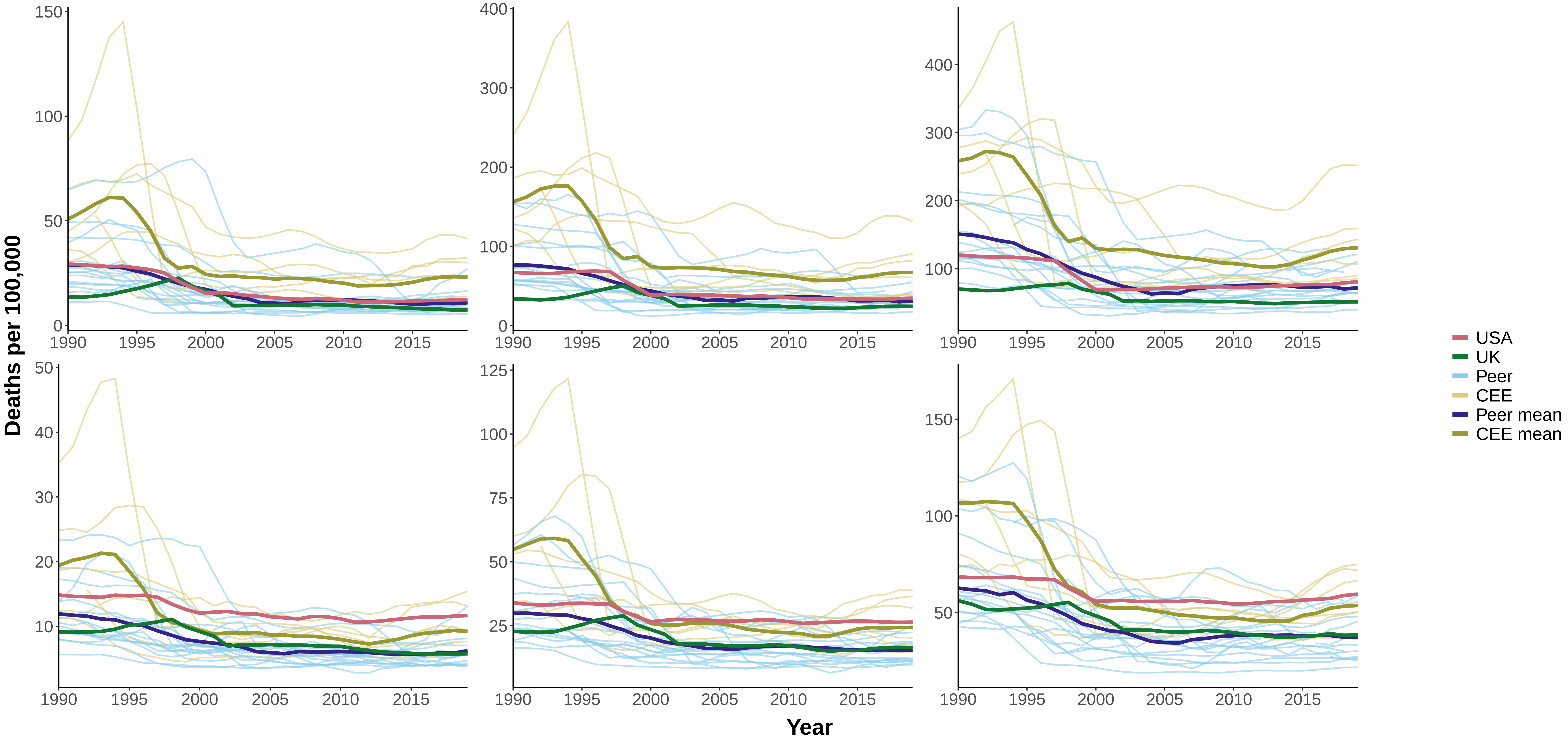

### Figure 14 Age−Standardized Mortality from All Causes, Years 1990−2019

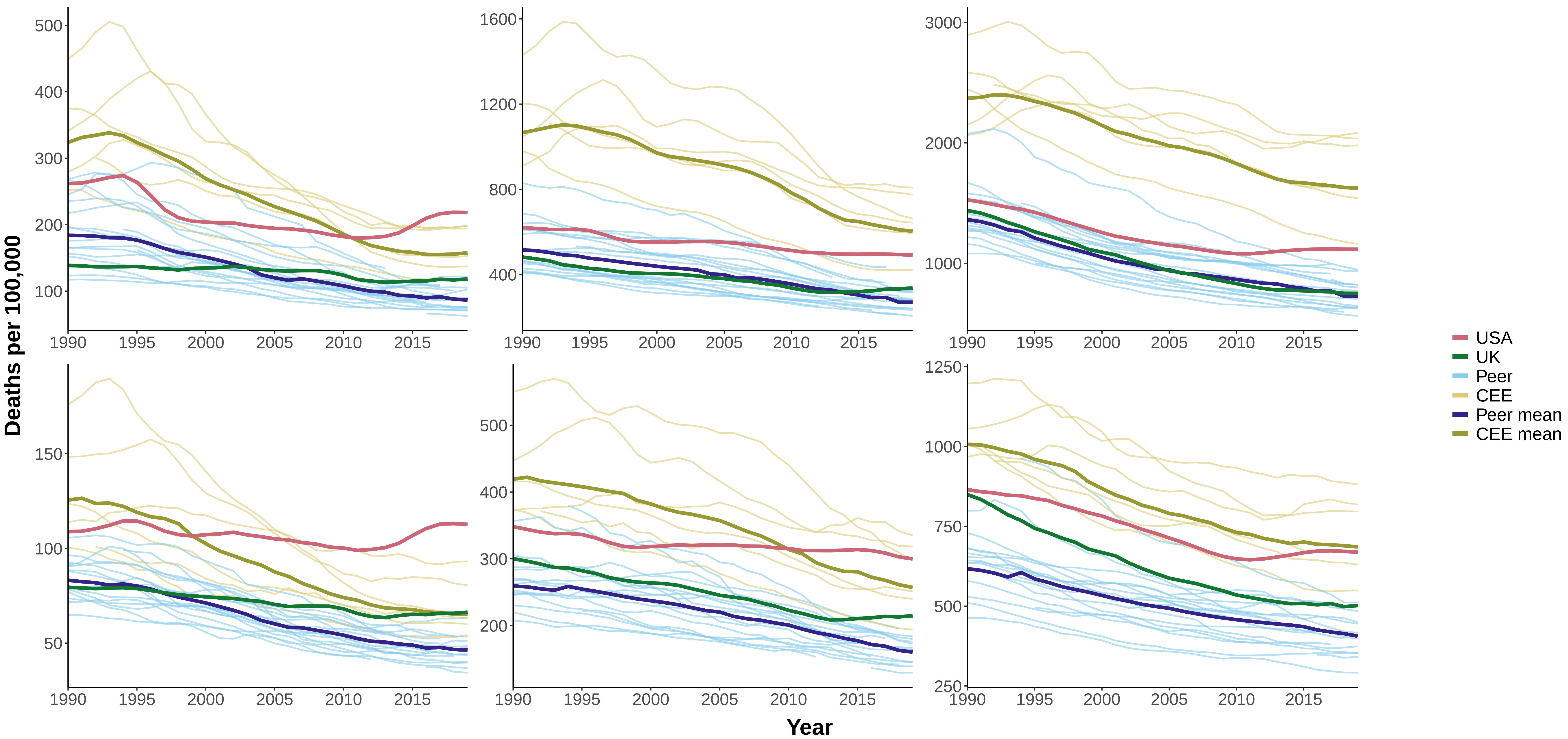

### Figure 15 Age−Standardized Mortality from Drug−Related Causes, Years 2000−2019

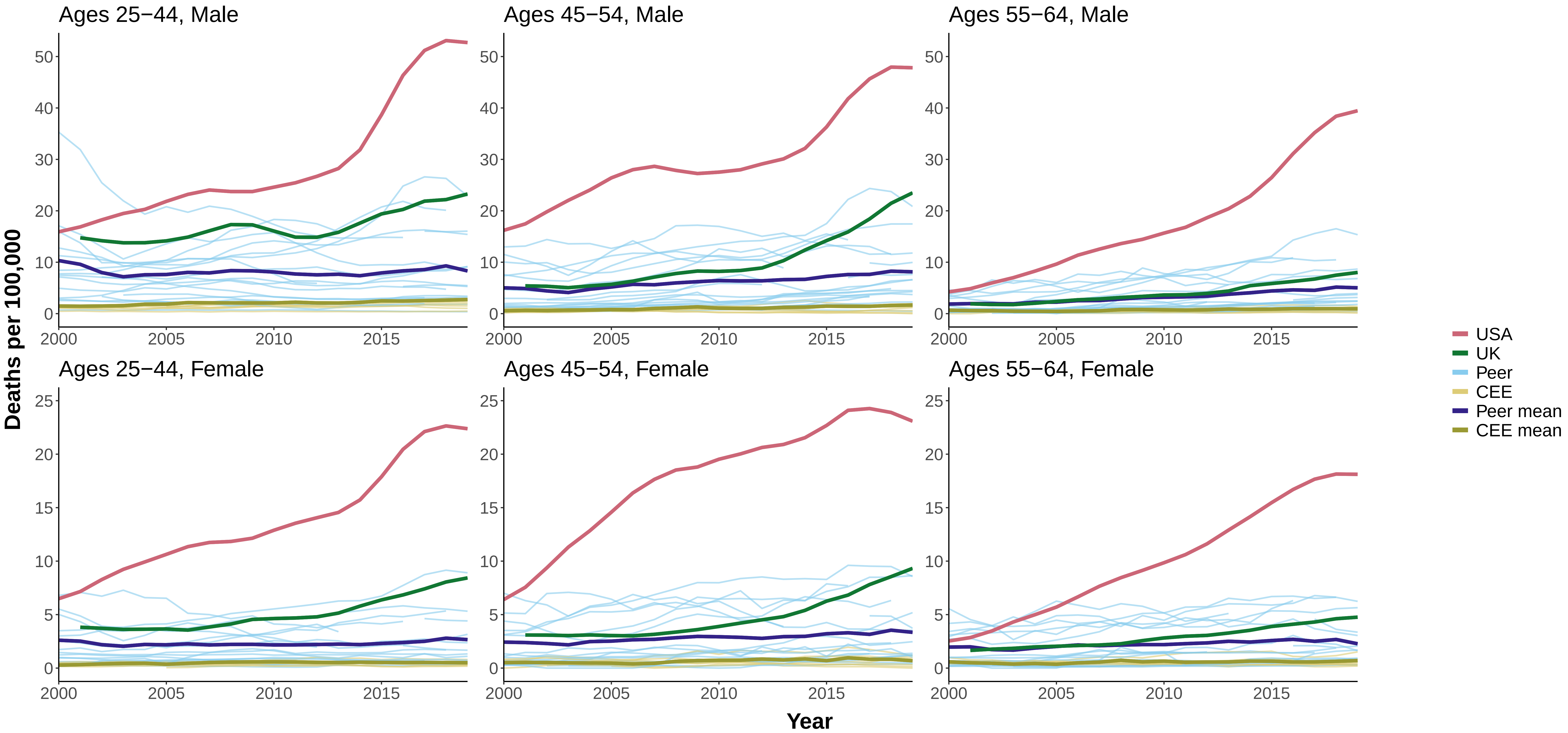

### Figure 16 Age−Standardized Mortality from Alcohol−Related Causes, Years 2000−2019

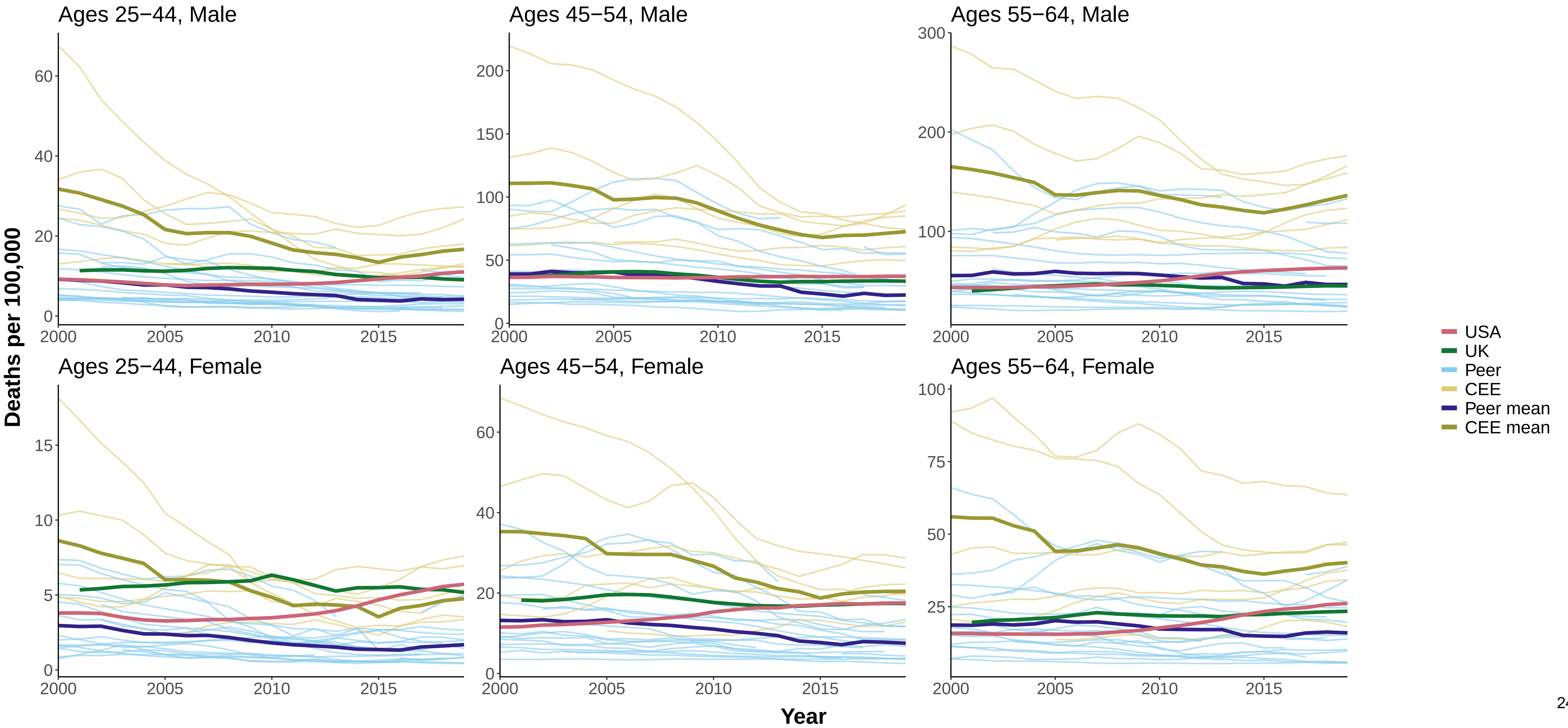

### Appendix 3

**Figure 1 Percent Change in Age−Standardized Mortality from the Baseline Year (1990), Infectious and Parasitic Diseases**

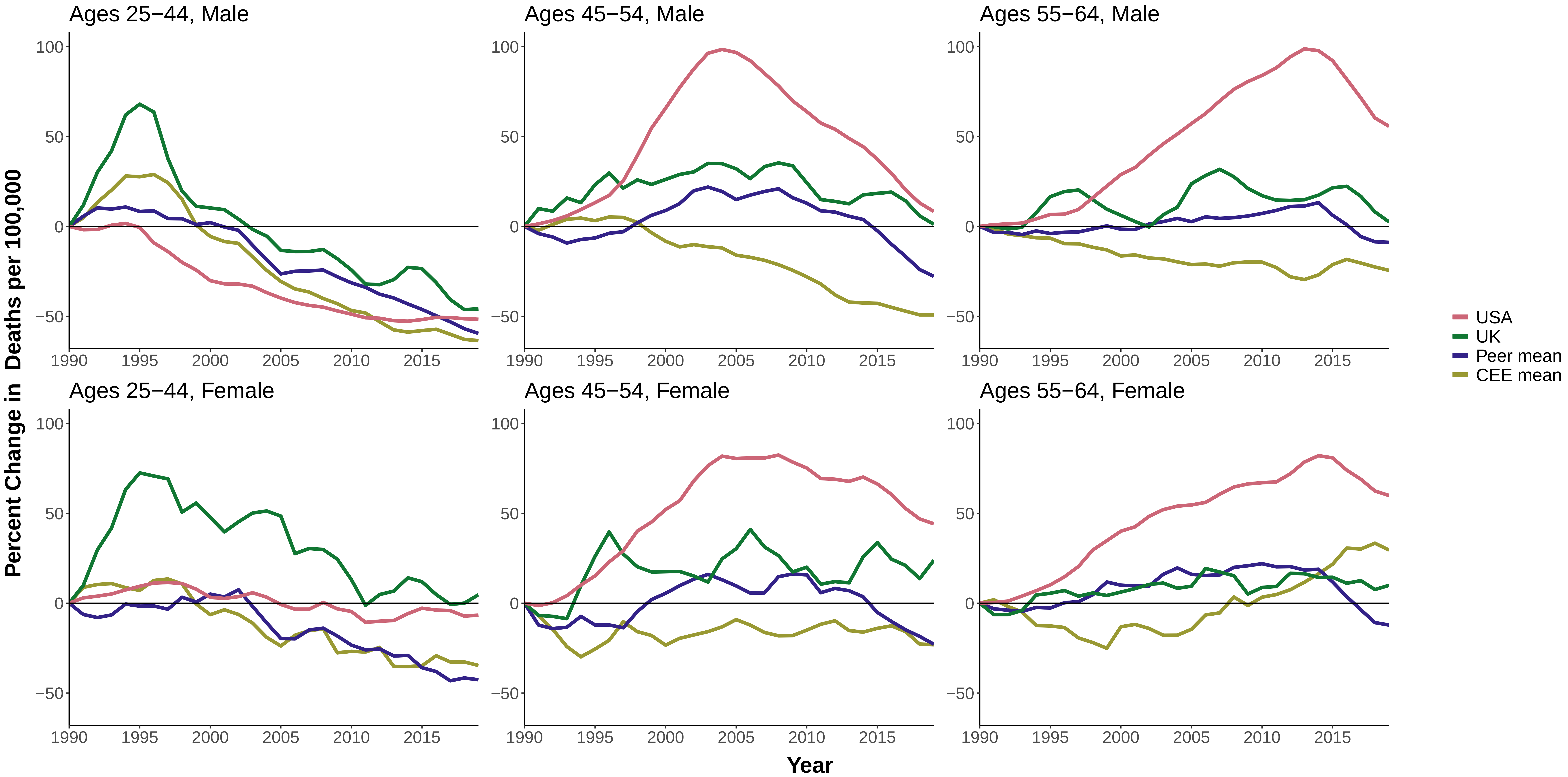

**Figure 2 Percent Change in Age−Standardized Mortality from the Baseline Year (1990), HIV/AIDS**

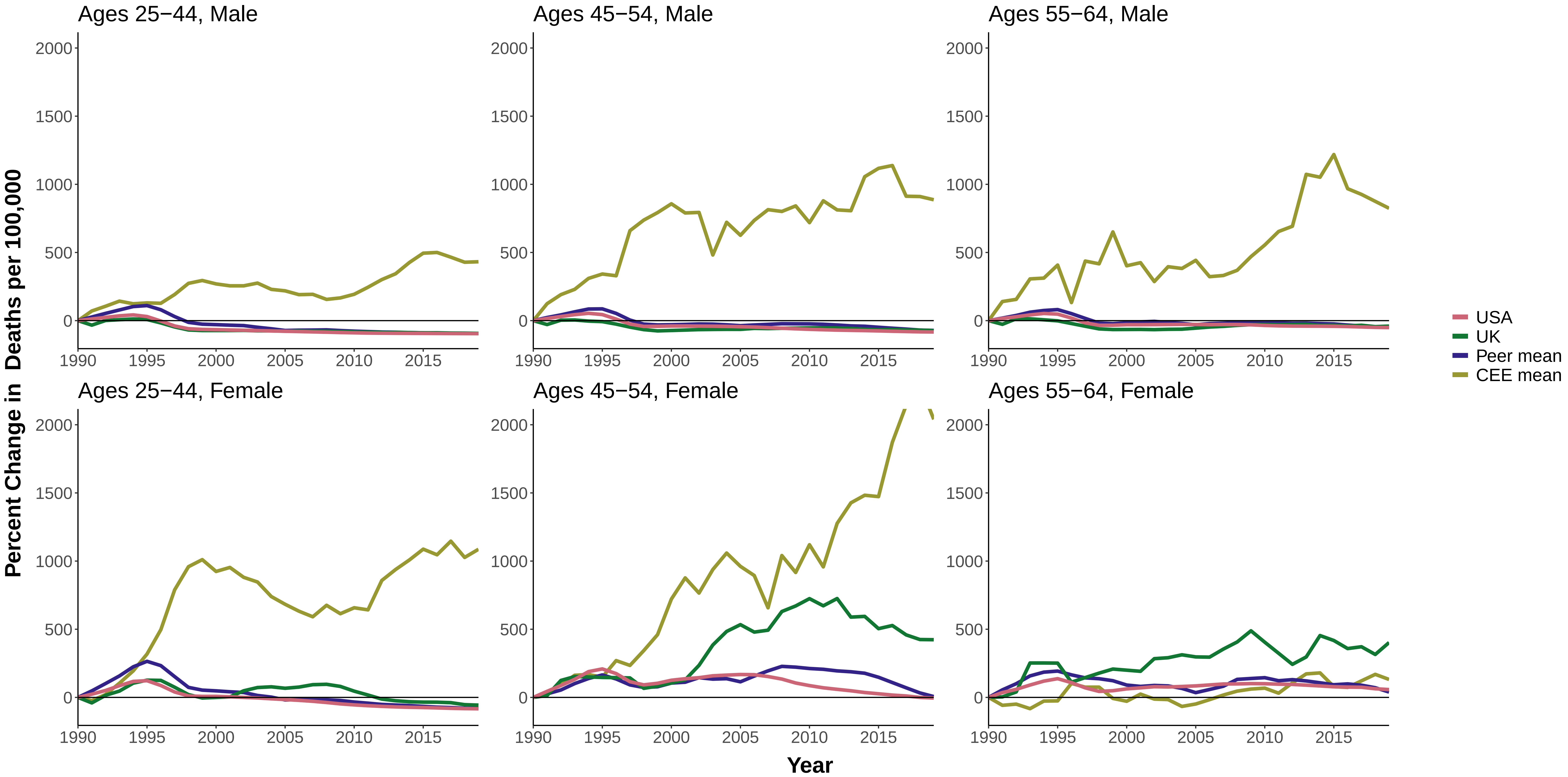

**Figure 3 Percent Change in Age−Standardized Mortality from the Baseline Year (1990), Respiratory Diseases**

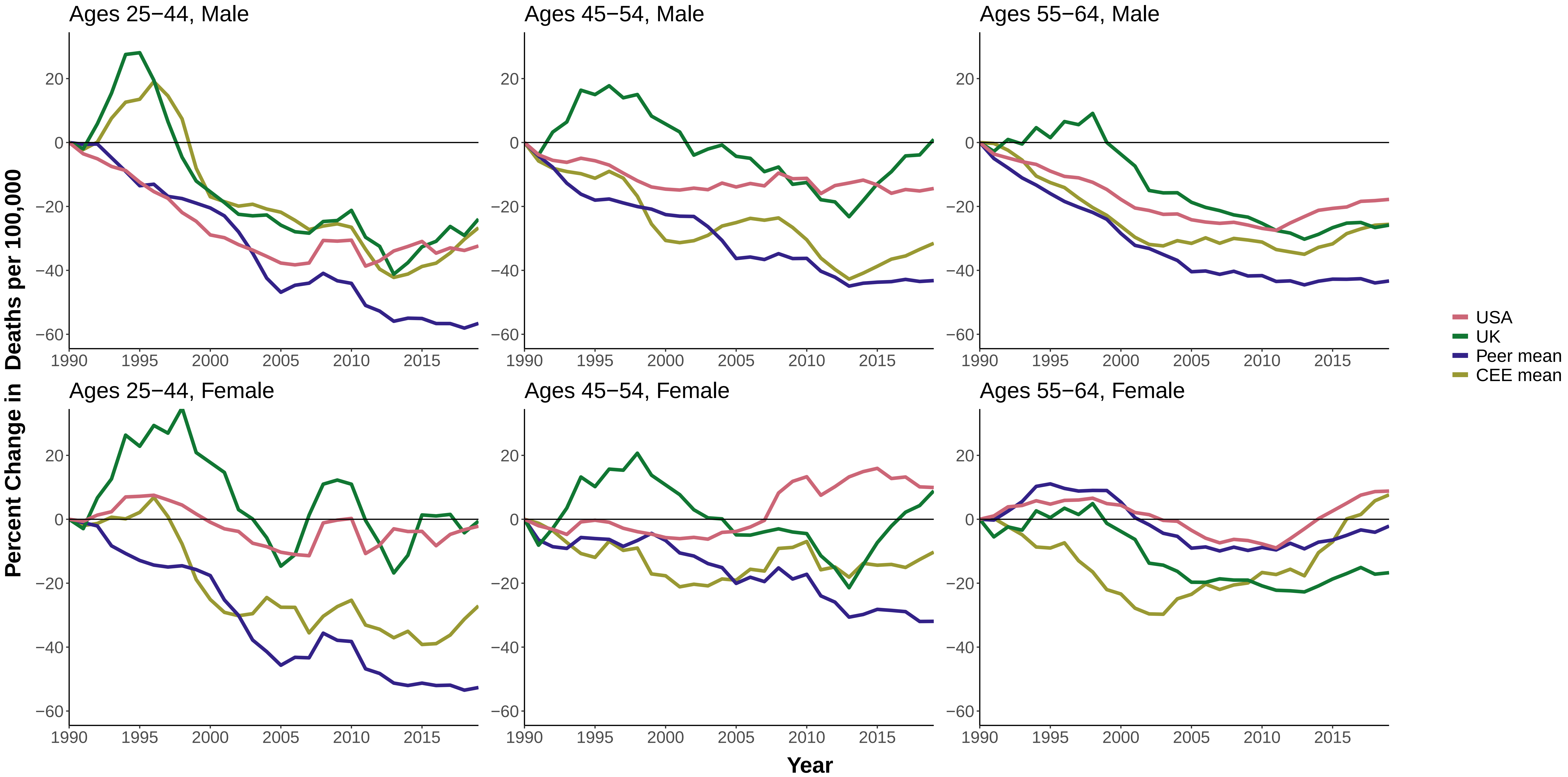

#### Figure 4 Percent Change in Age−Standardized Mortality from the Baseline Year (1990), Trachea/Bronchus, Lung Cancers

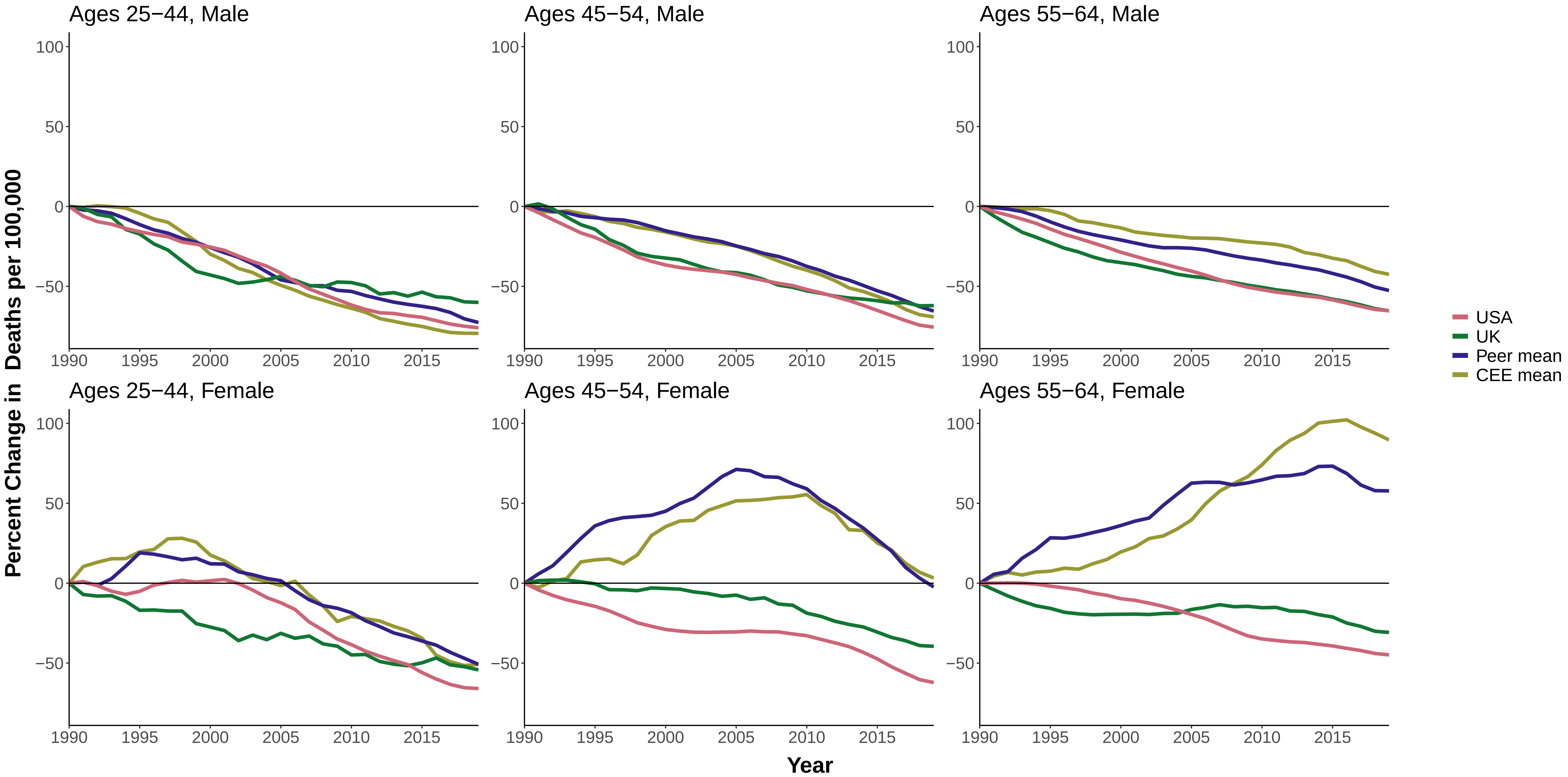

Figure 5 Percent Change in Age−Standardized Mortality from the Baseline Year (1990), All Other Cancers

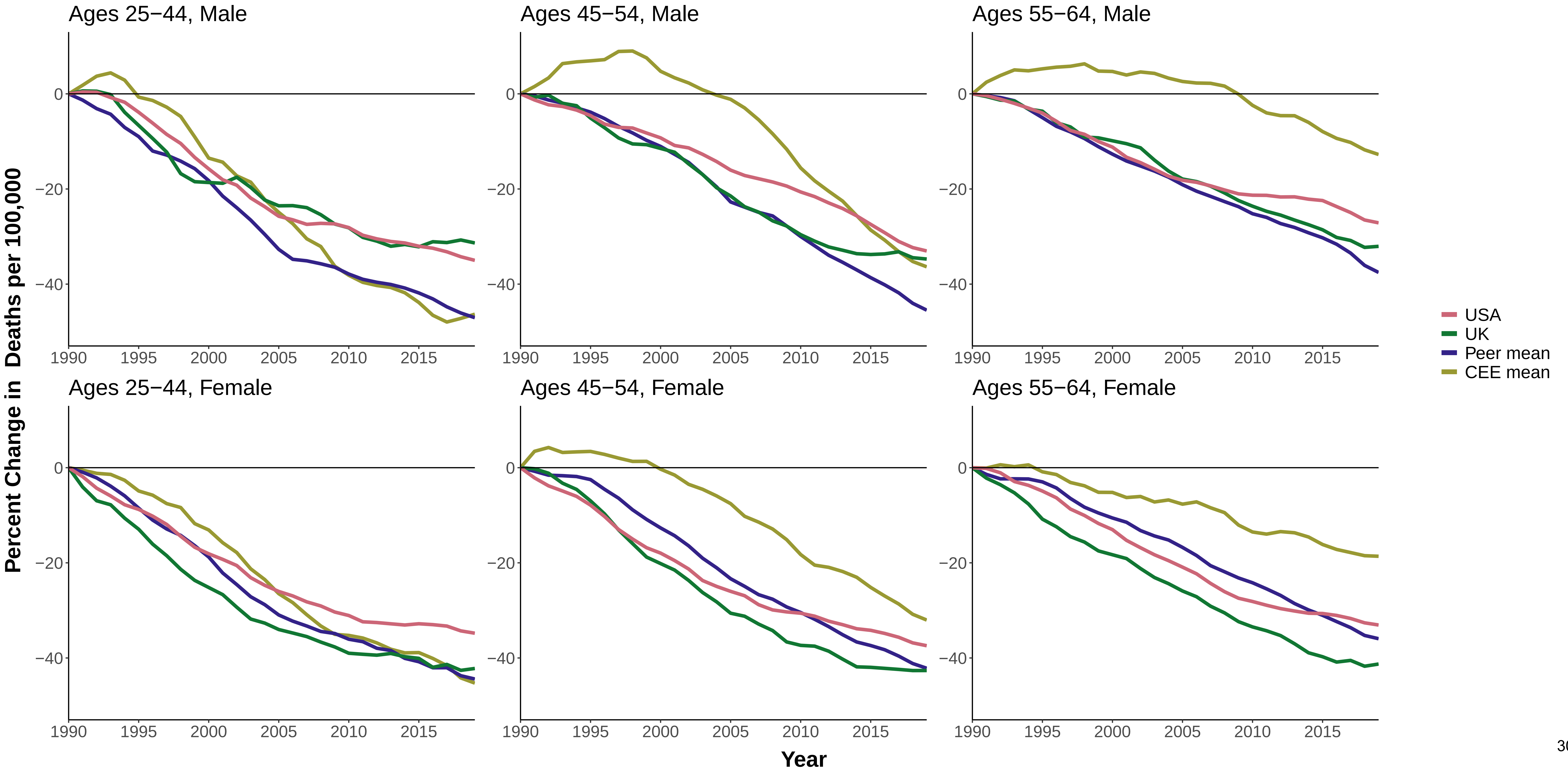

**Figure 6 Percent Change in Age−Standardized Mortality from the Baseline Year (1990), Nervous System Diseases**

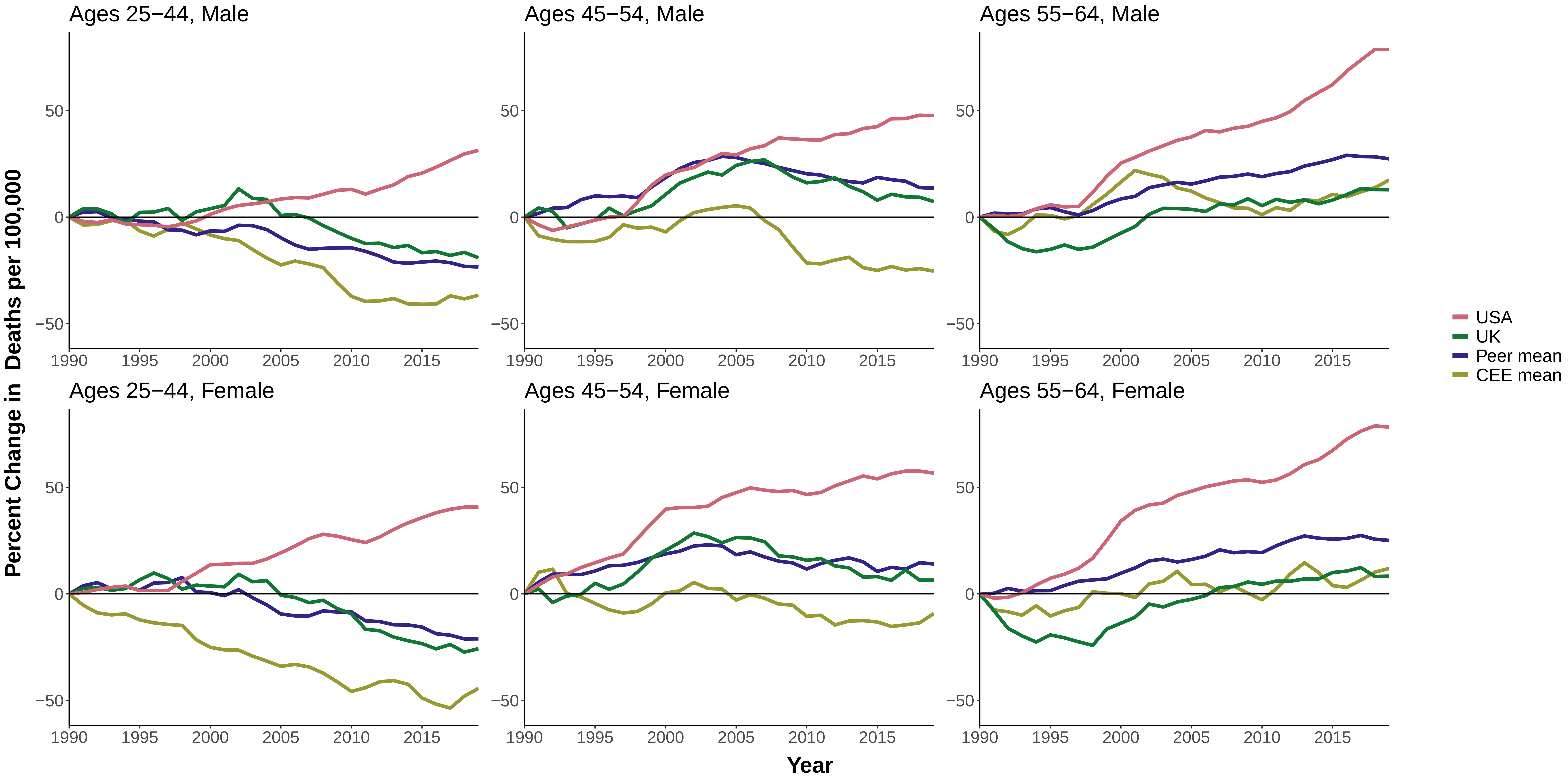

**Figure 7 Percent Change in Age−Standardized Mortality from the Baseline Year (1990), Metabolic Diseases**

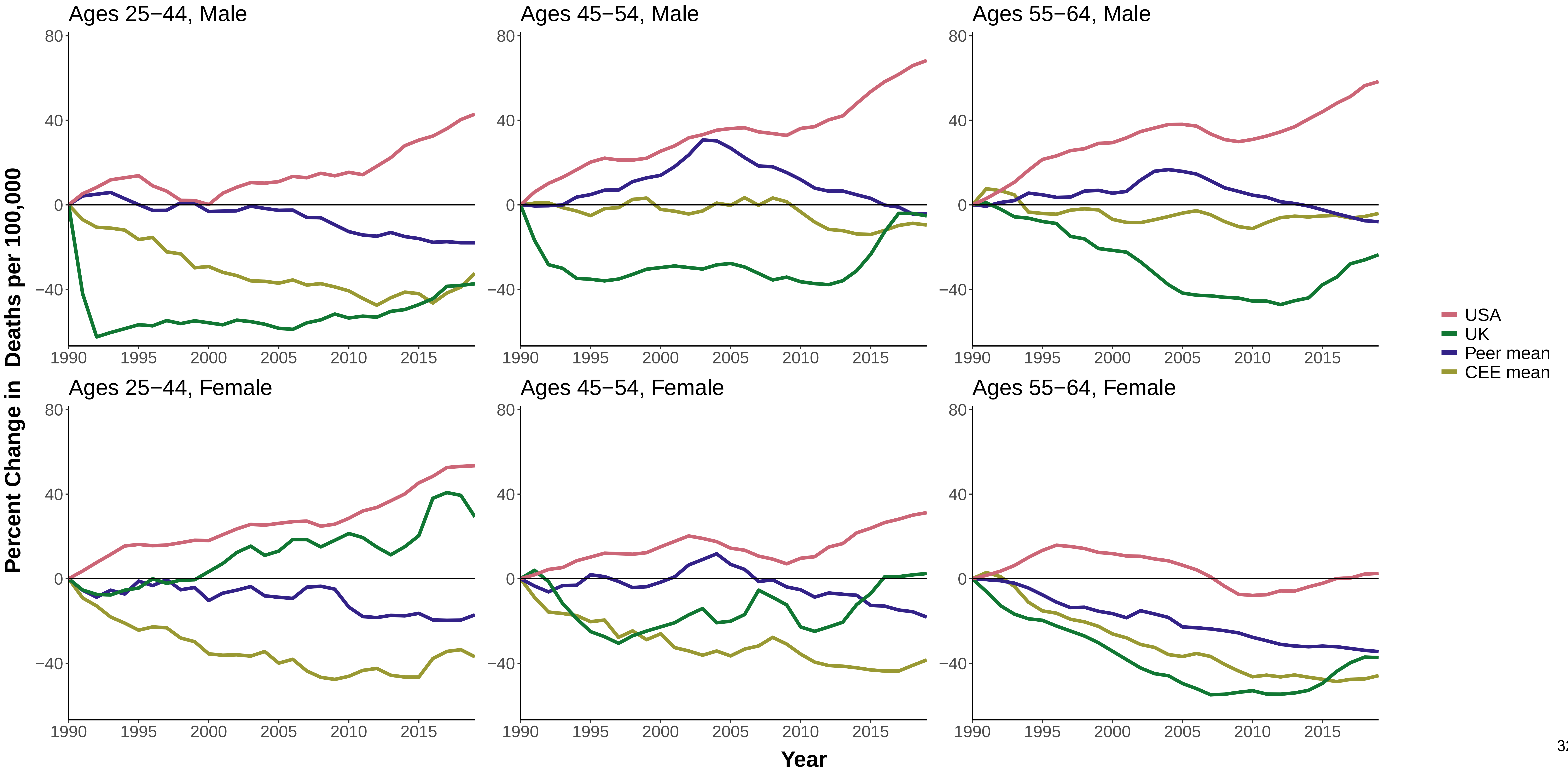

**Figure 8 Percent Change in Age−Standardized Mortality from the Baseline Year (1990), Cardiovascular Disease**

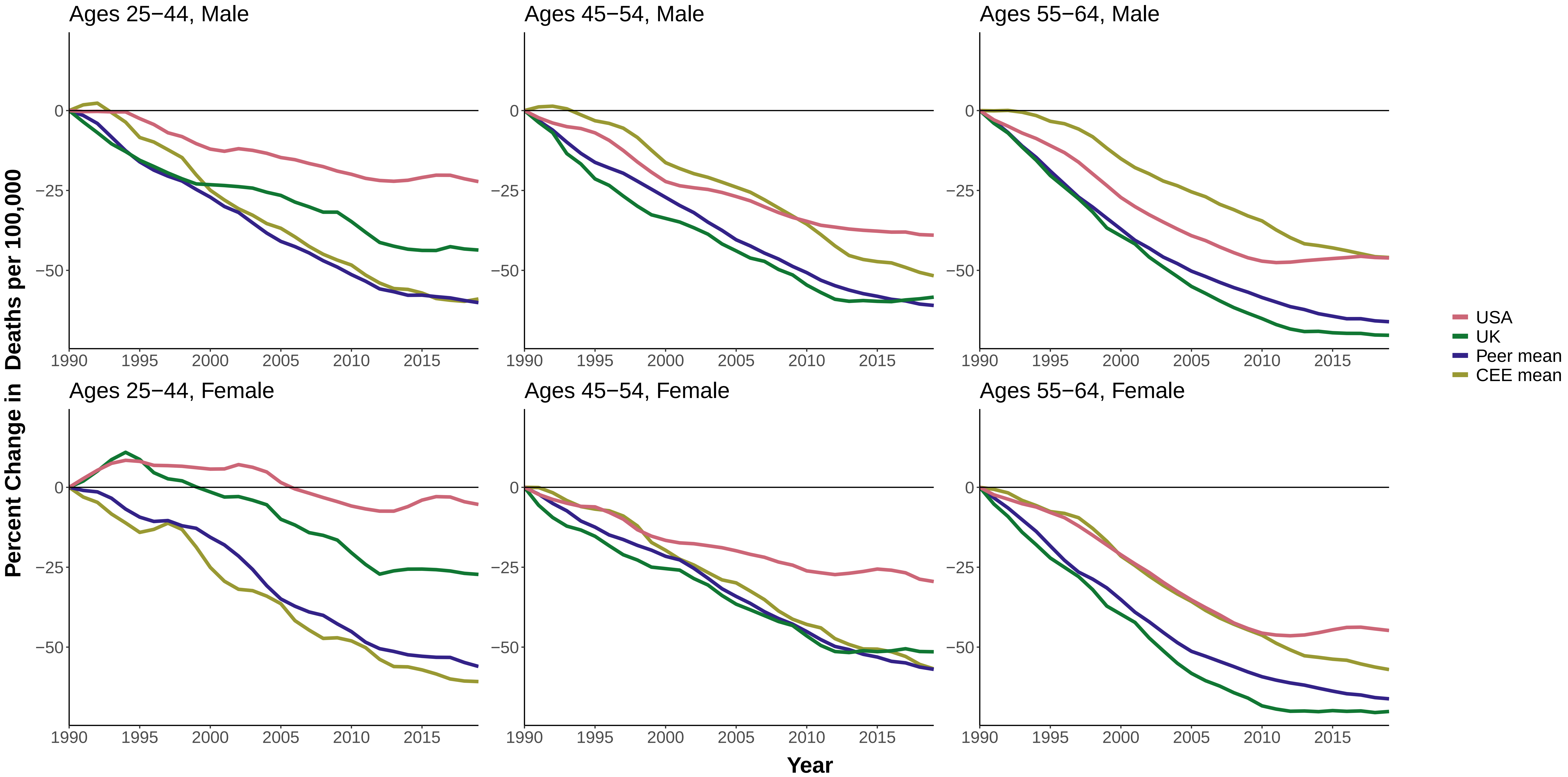

### Figure 9 Percent Change in Age−Standardized Mortality from the Baseline Year (1990), Suicide

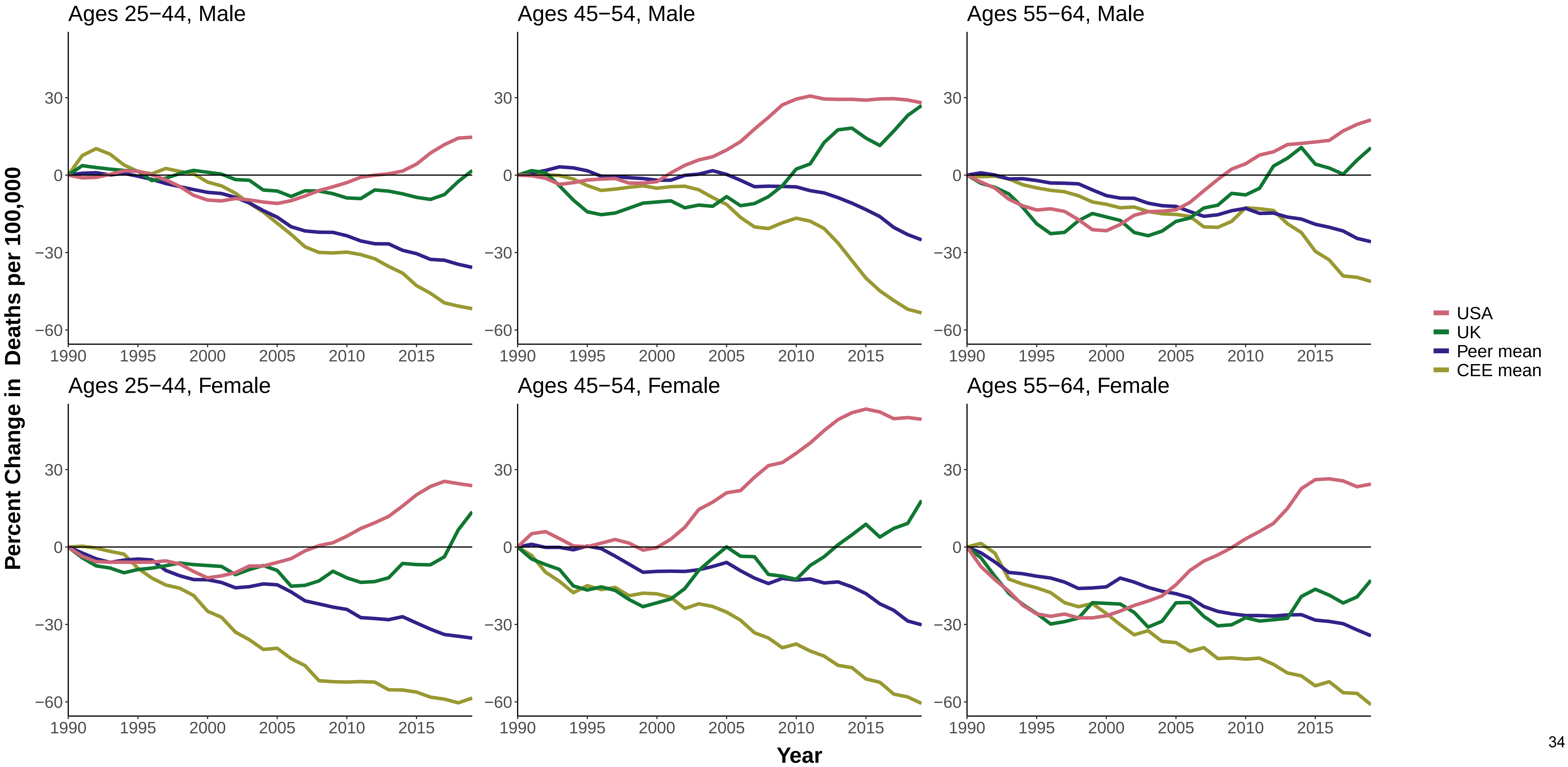

**Figure 10 Percent Change in Age−Standardized Mortality from the Baseline Year (1990), Homicide**

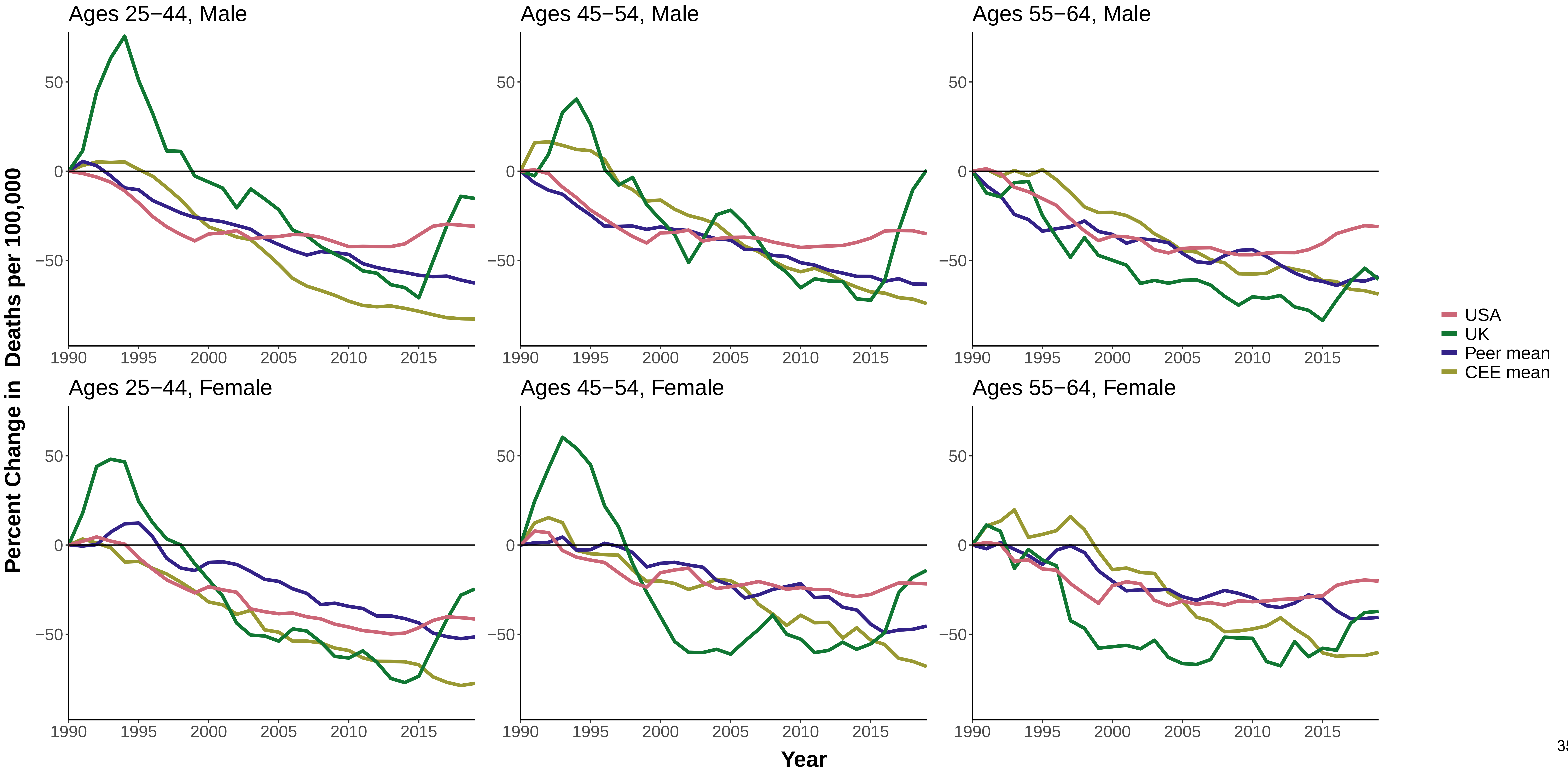

#### Figure 11 Percent Change in Age−Standardized Mortality from the Baseline Year (1990), Transport Accidents

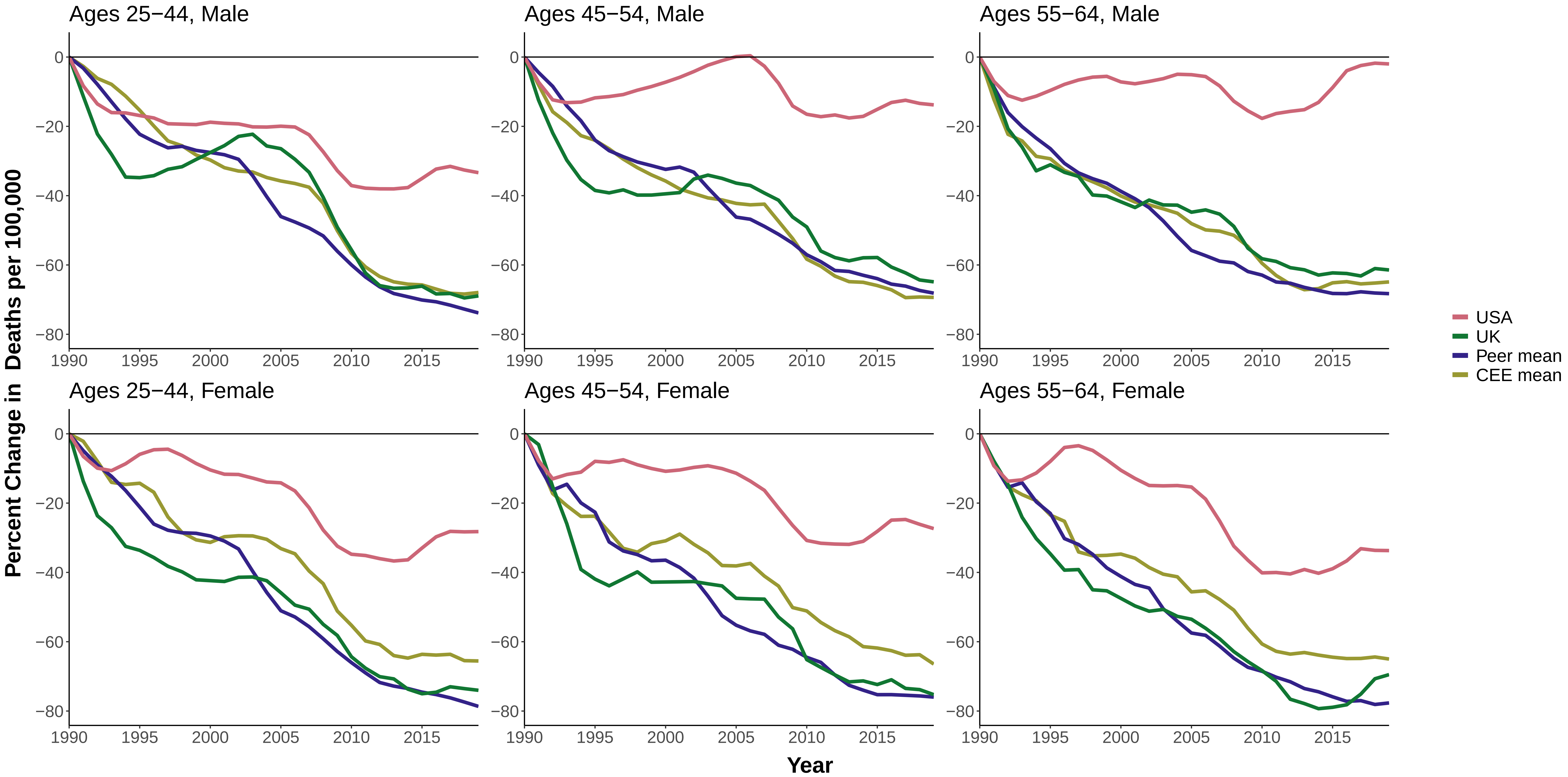

**Figure 12 Percent Change in Age−Standardized Mortality from the Baseline Year (1990), Other External Causes**

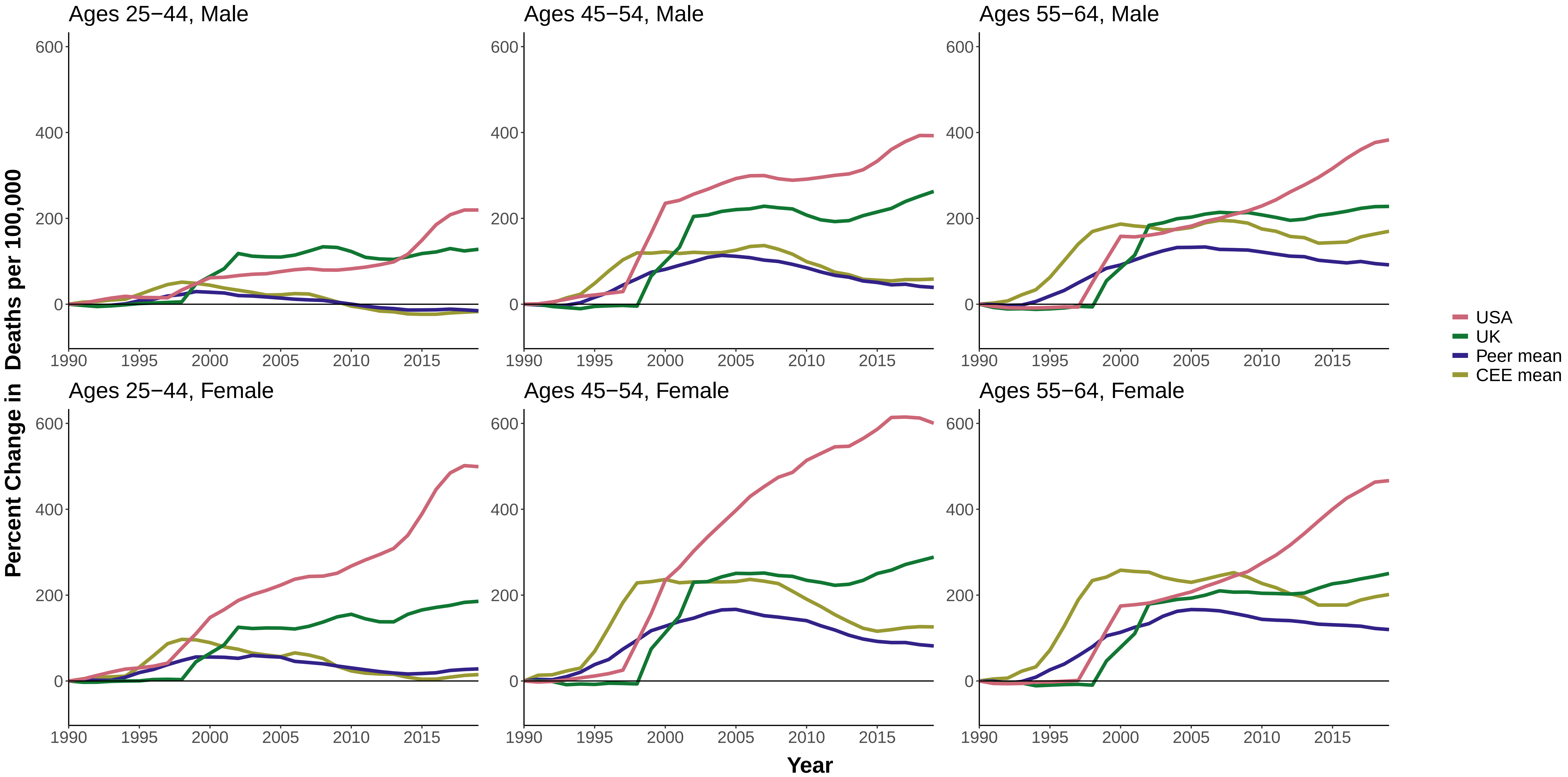

**Figure 13 Percent Change in Age−Standardized Mortality from the Baseline Year (1990), All Other Causes**

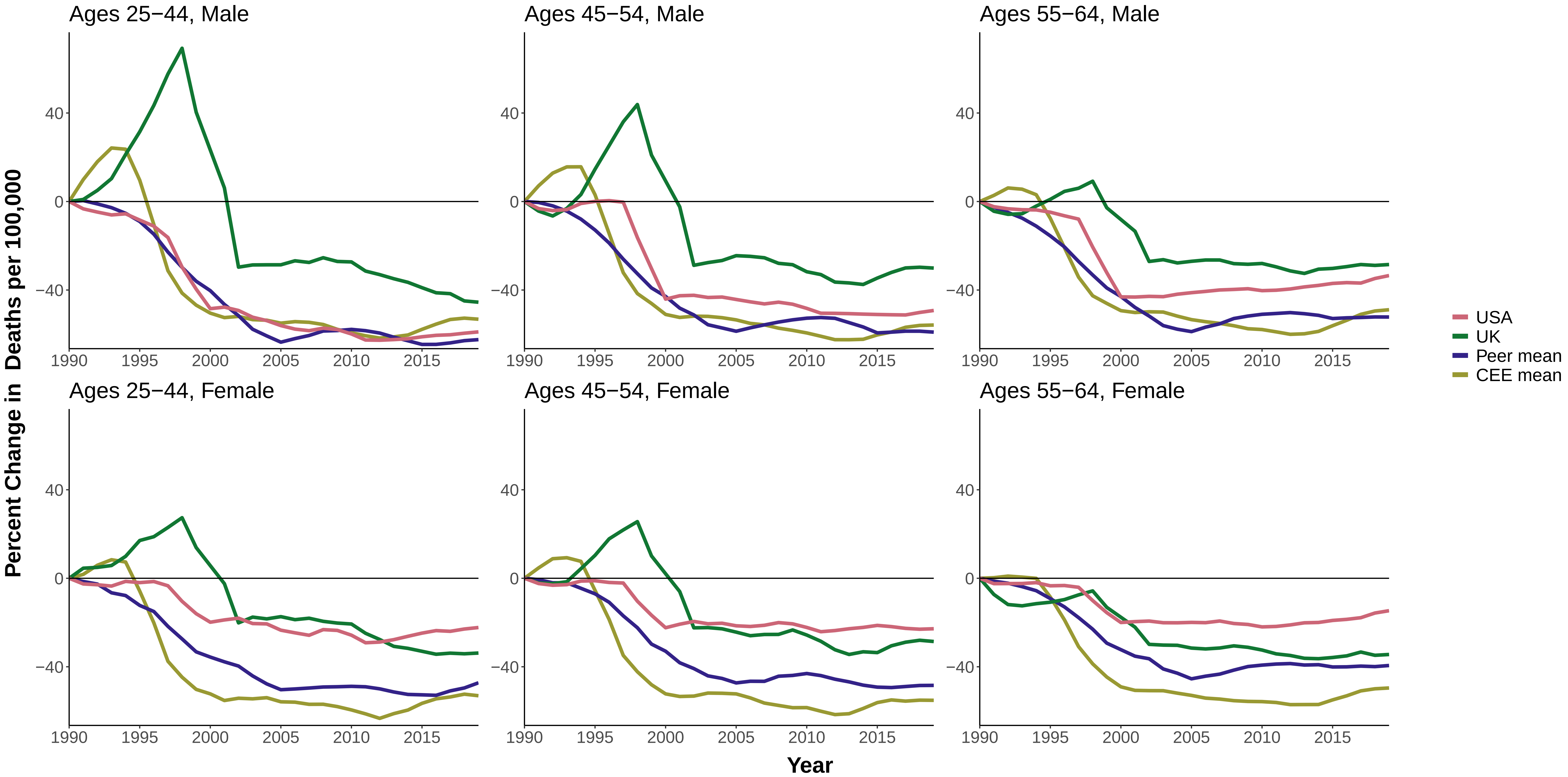

#### Figure 14 Percent Change in Age−Standardized Mortality from the Baseline Year (1990), All Causes

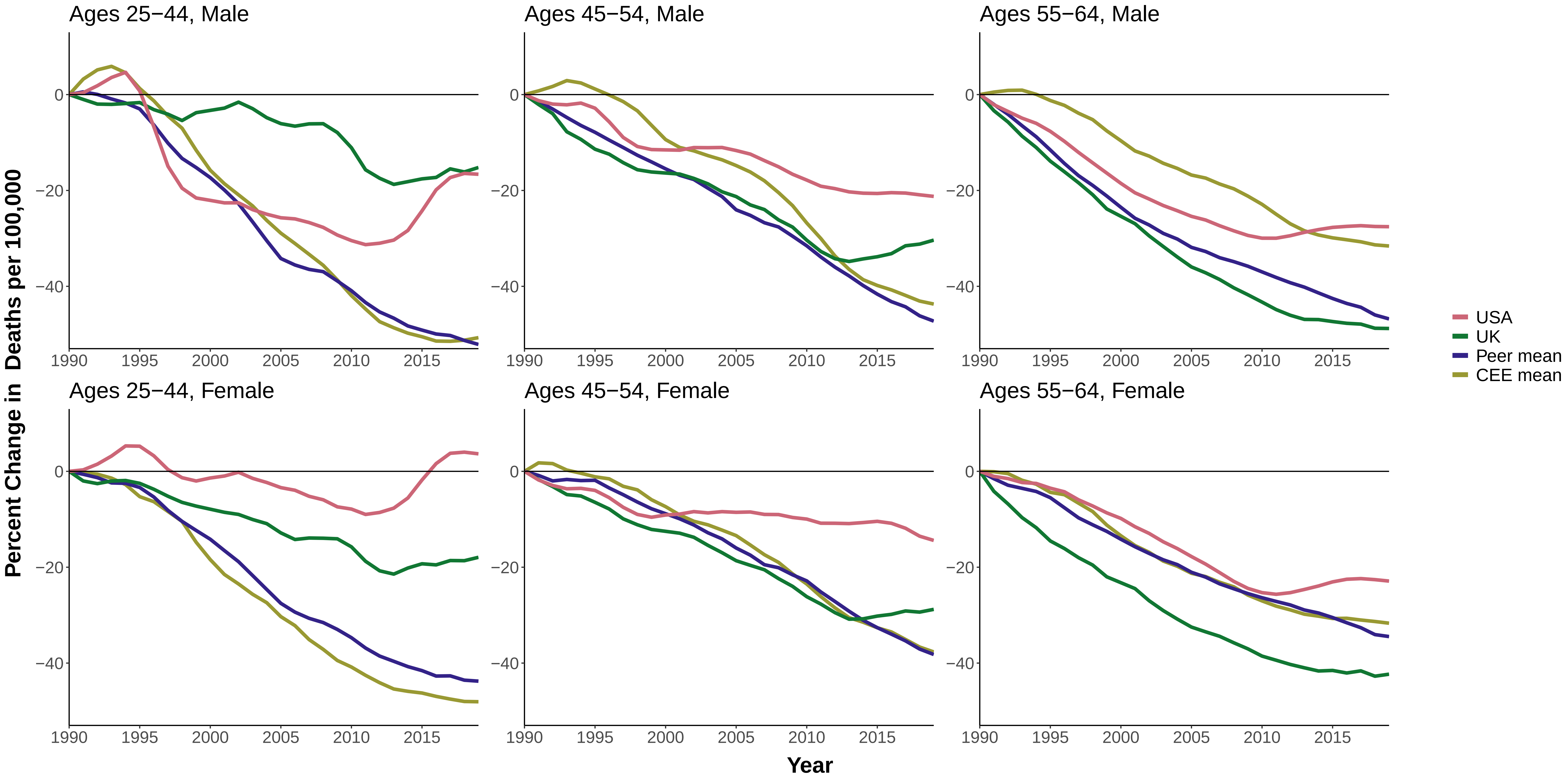

### Figure 15 Percent Change in Age−Standardized Mortality from the Baseline Year (2000), Drug−Related Causes

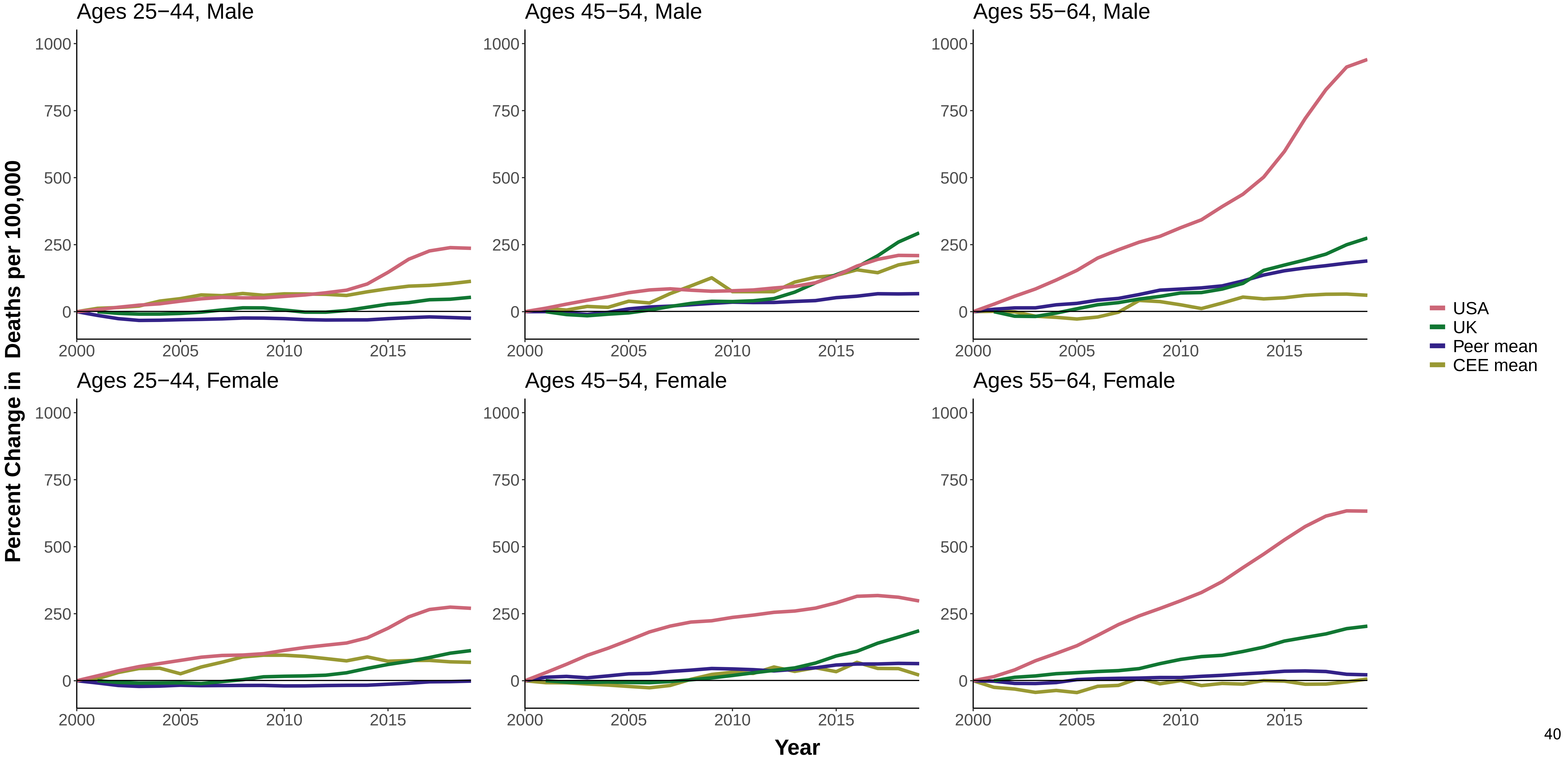

### Figure 16 Percent Change in Age−Standardized Mortality from the Baseline Year (2000), Alcohol−Related Causes

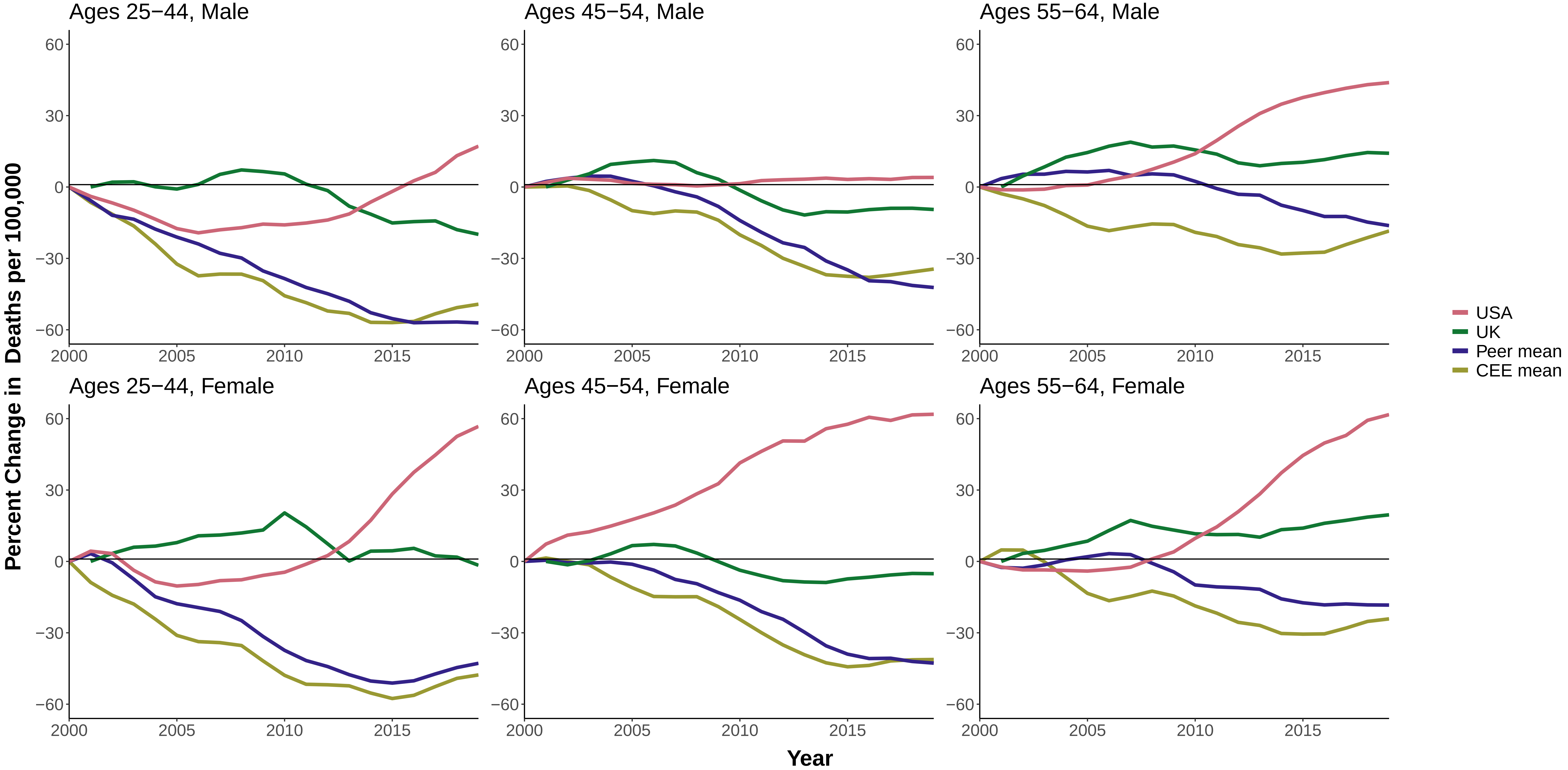

**Figure 17 Percent Change in All−Cause Mortality Between 1990 and 2019/Males, Ages 25−44**

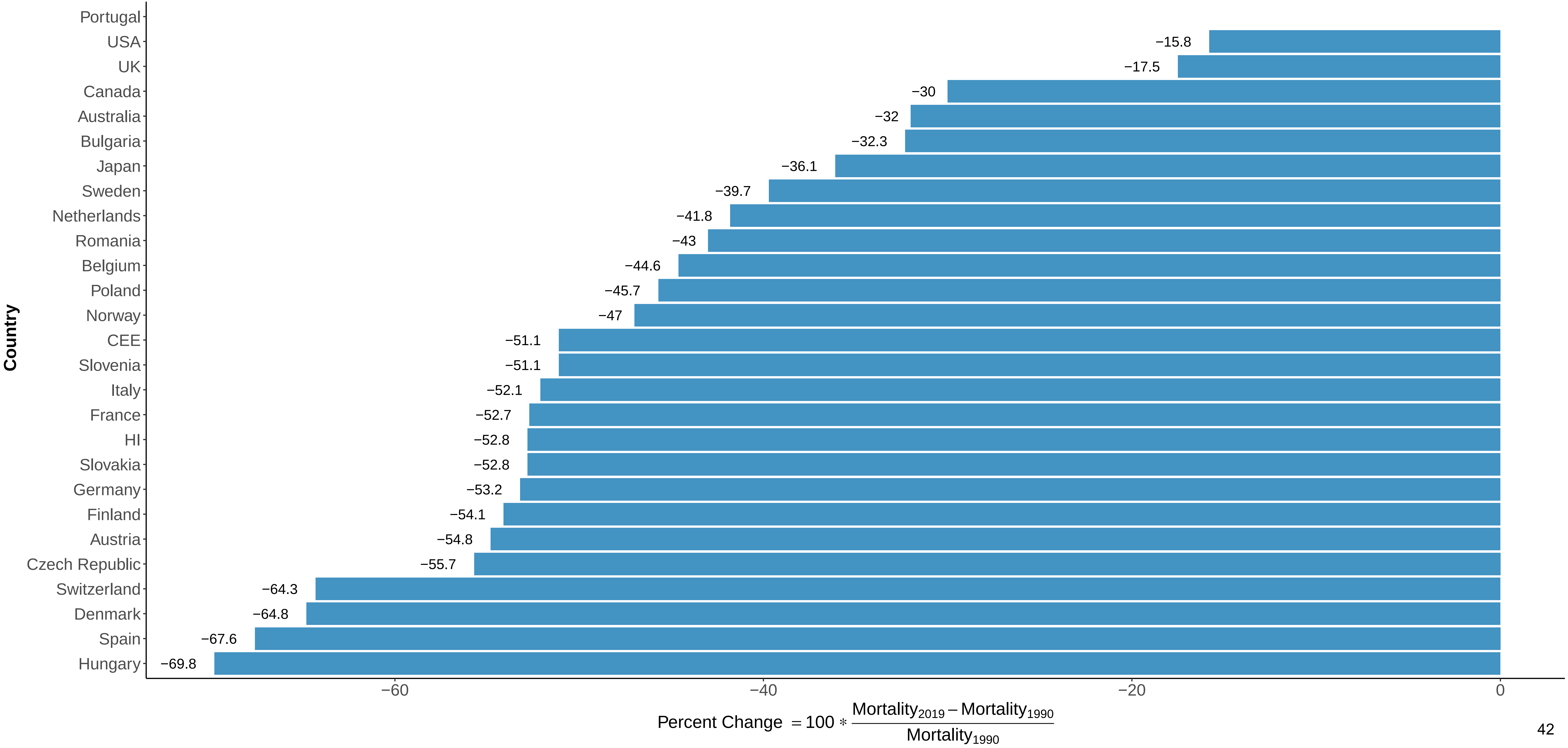

**Figure 18 Percent Change in All−Cause Mortality Between 1990 and 2019/Males, Ages 45−54**

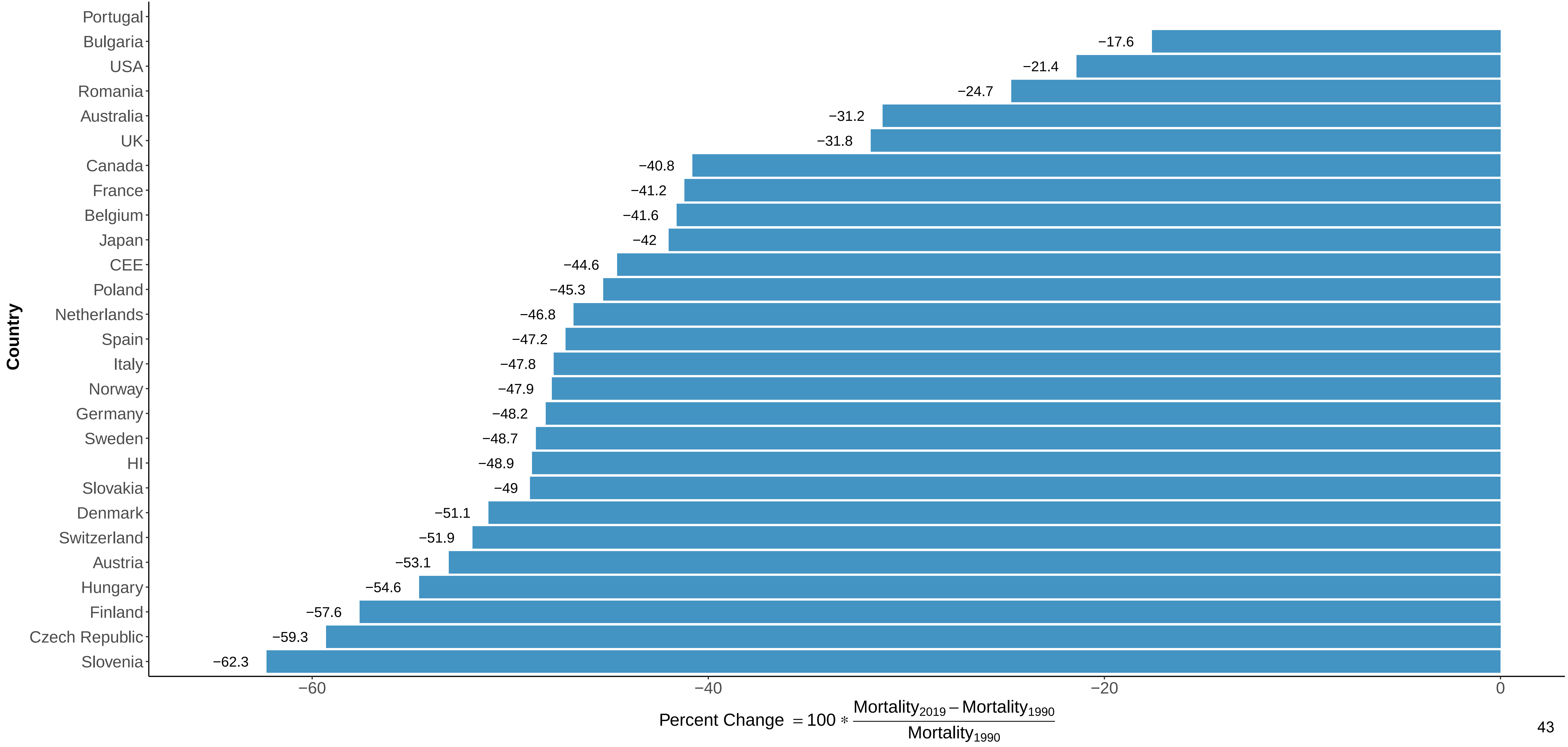

**Figure 19 Percent Change in All−Cause Mortality Between 1990 and 2019/Males, Ages 55−64**

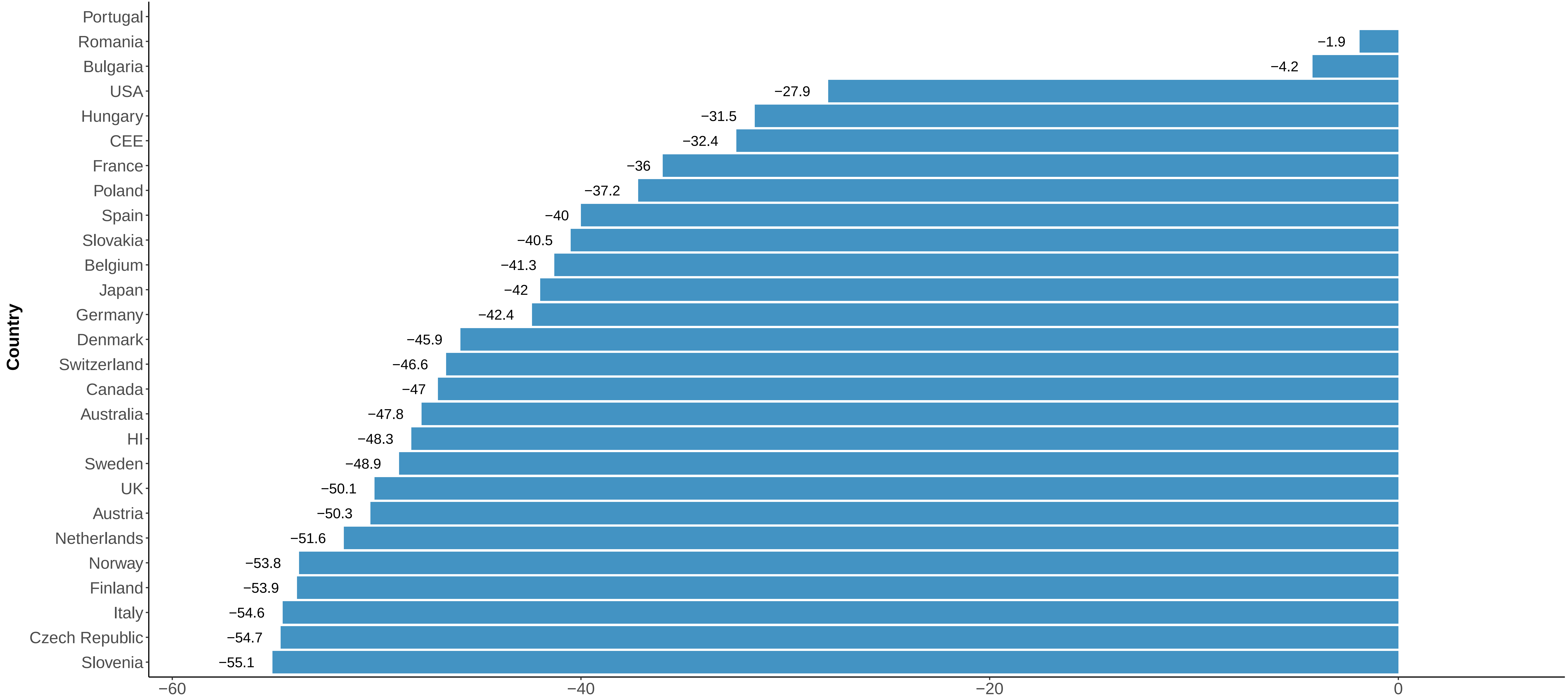

$$\text{Percent Change} = 100 * \frac{\text{Mortality}_{2019} - \text{Mortality}_{1990}}{\text{Mortality}_{1990}}$$

#### Figure 20 Percent Change in All−Cause Mortality Between 1990 and 2019/Females, Ages 25−44

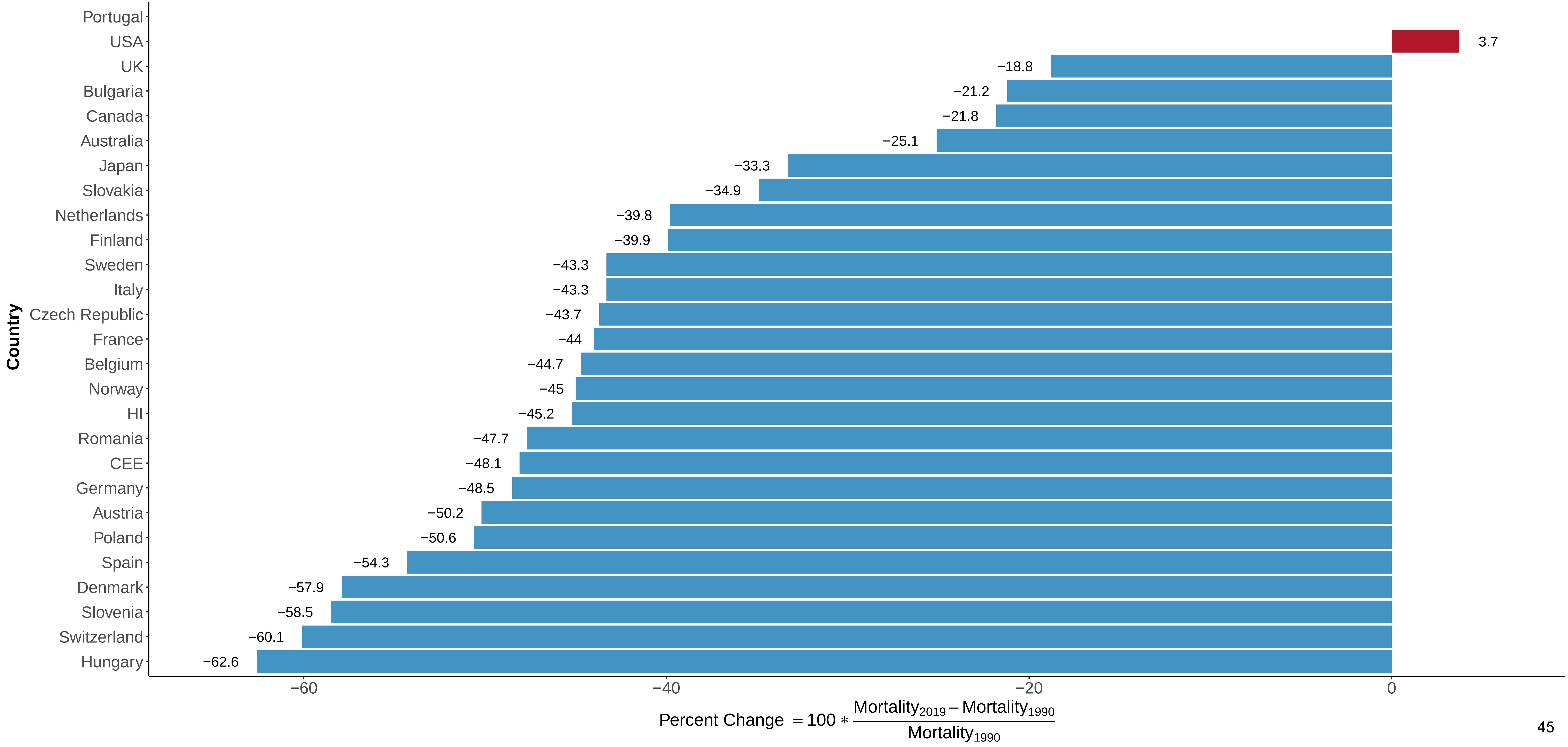

#### Figure 21 Percent Change in All−Cause Mortality Between 1990 and 2019/Females, Ages 45−54

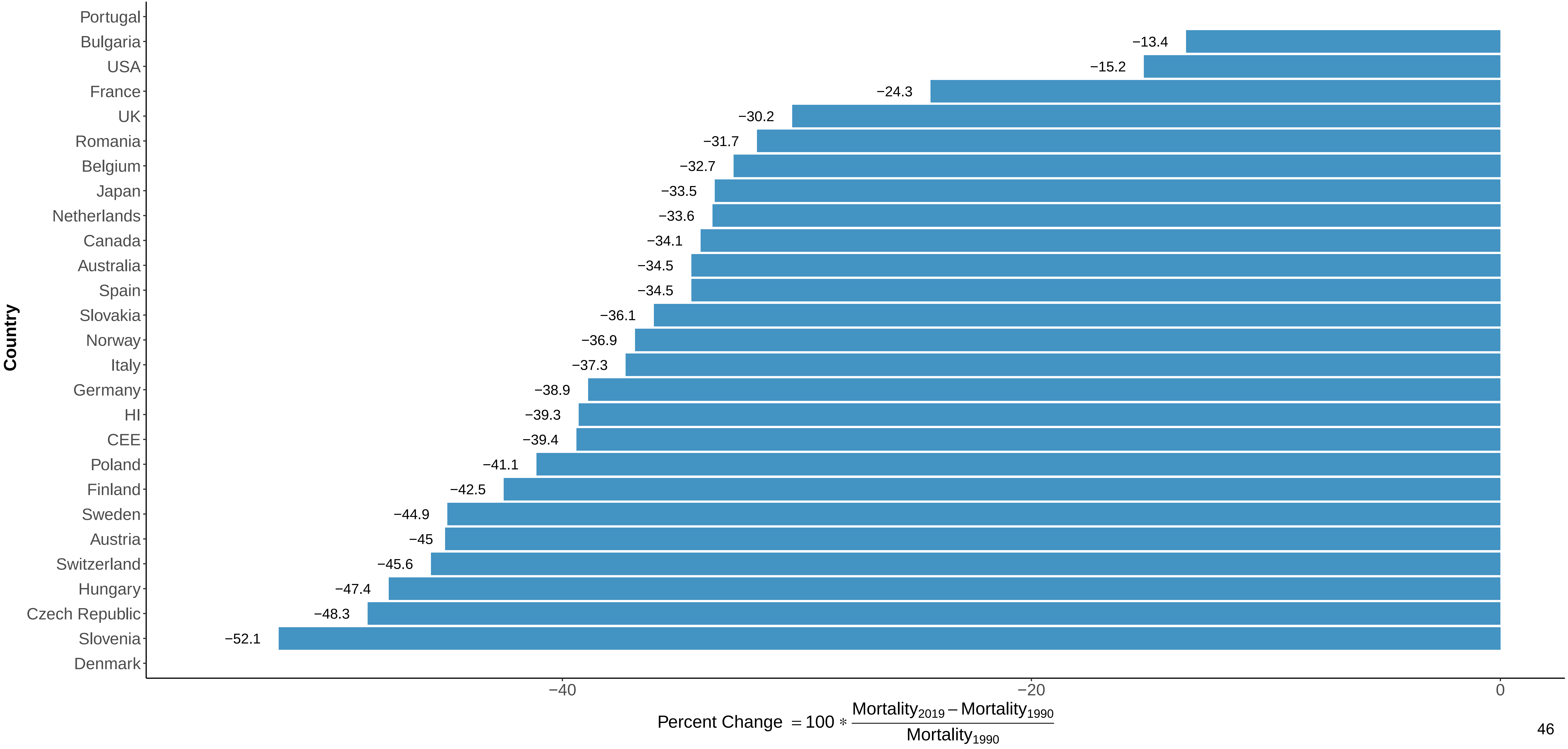

**Figure 22 Percent Change in All−Cause Mortality Between 1990 and 2019/Females, Ages 55−64**

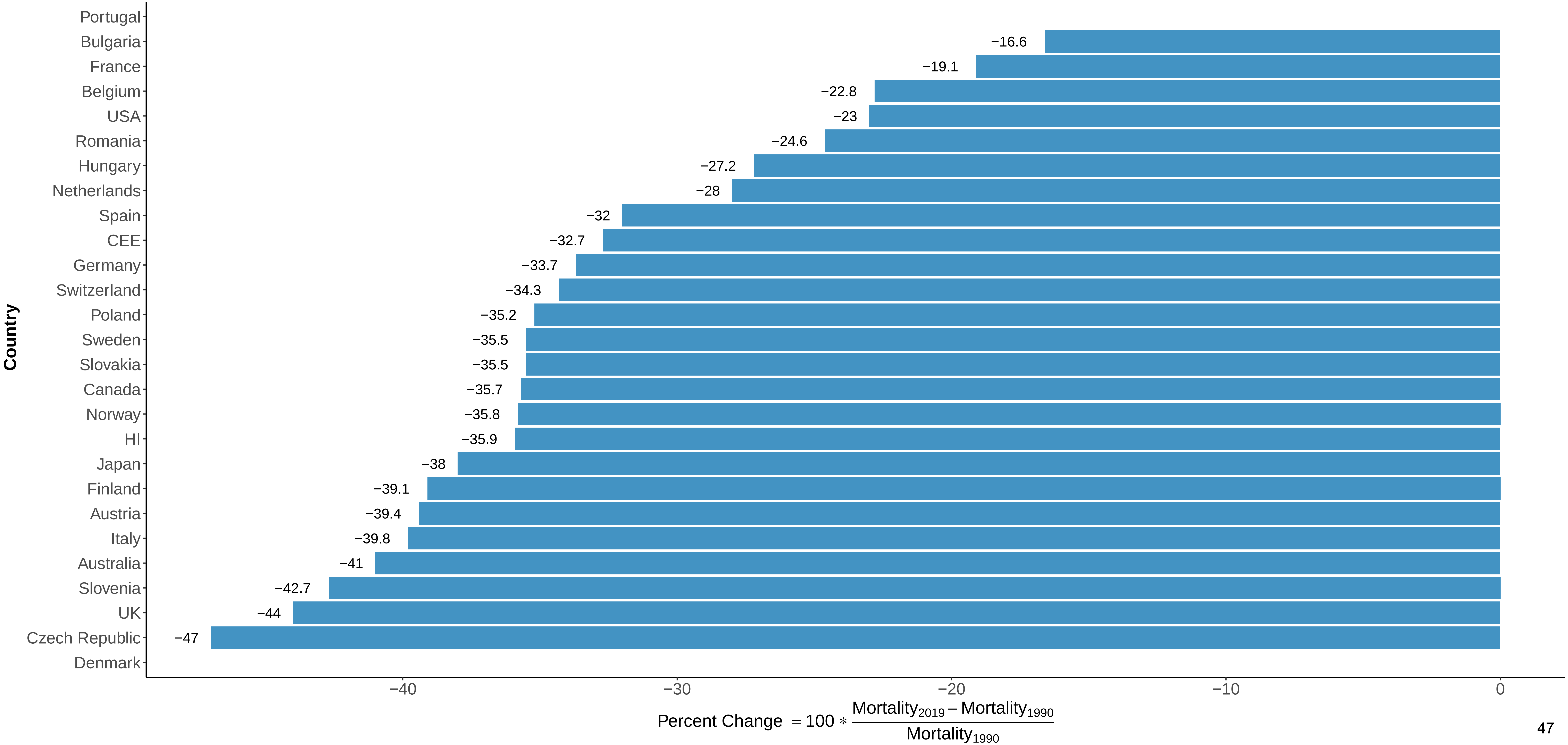

### Appendix 4

Figure 1 Three−Year Moving Average of Male Mortality from Infectious and Parasitic Diseases at Ages 25−44

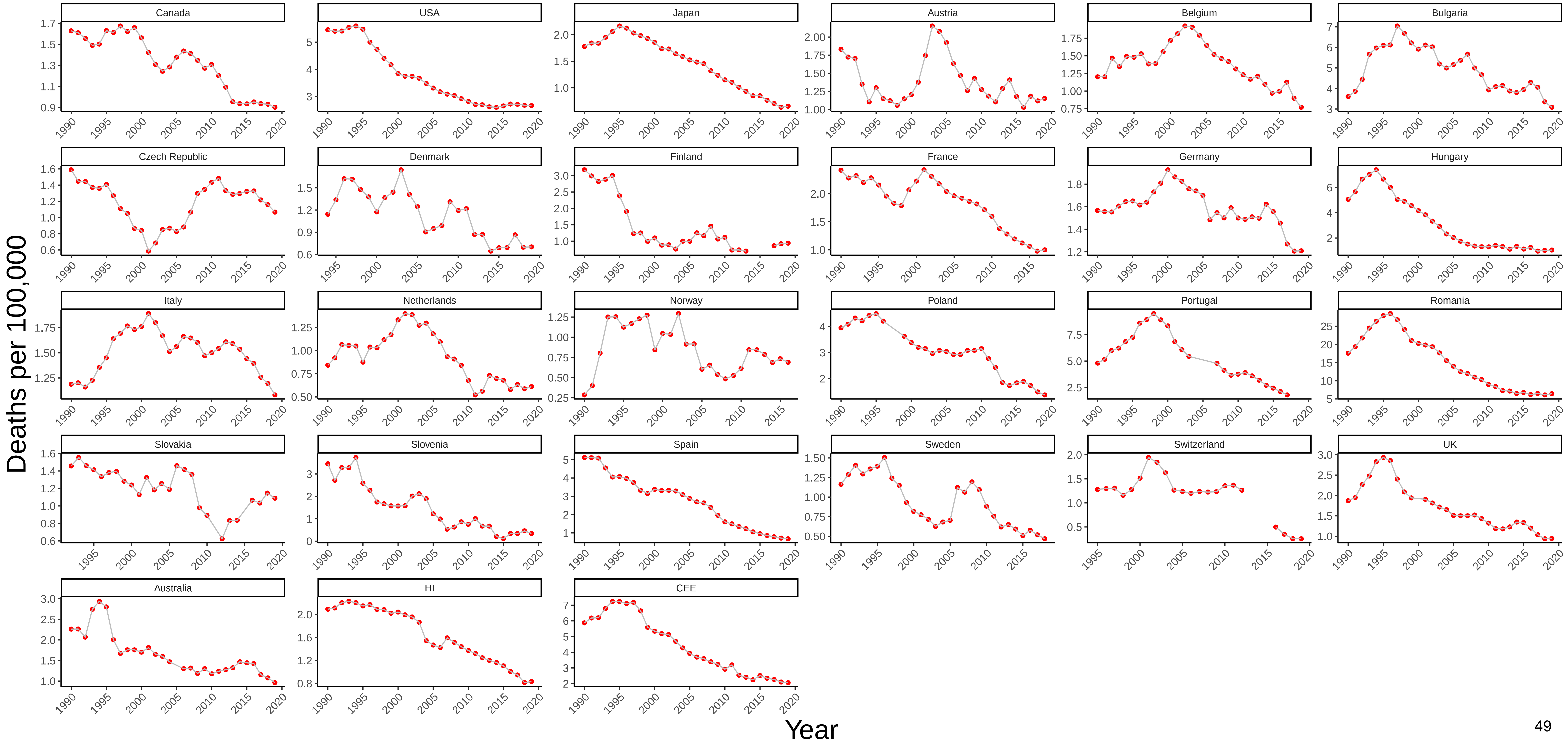

Figure 2 Three−Year Moving Average of Female Mortality from Infectious and Parasitic Diseases at Ages 25−44

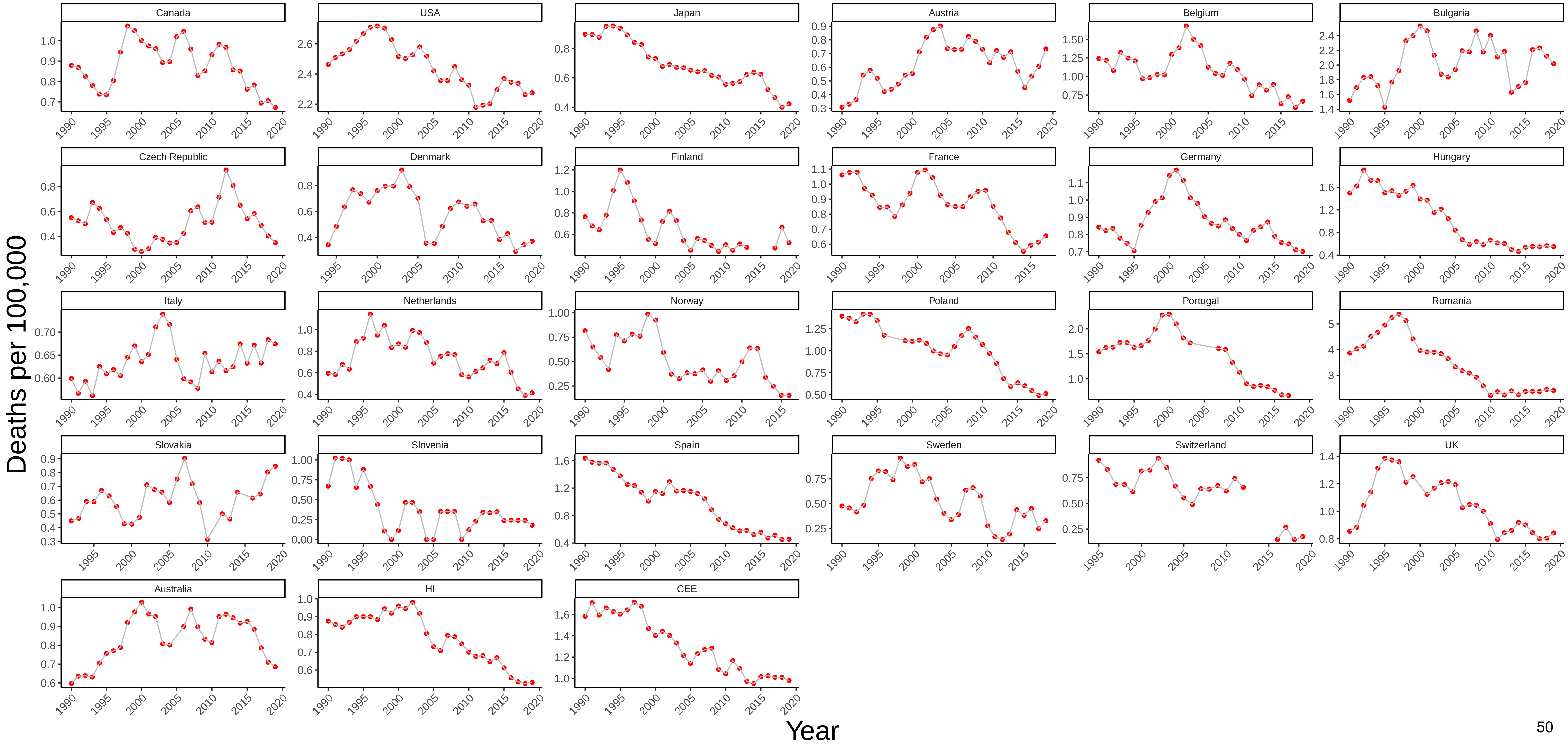

### Figure 3 Three−Year Moving Average of Male Mortality from HIV/AIDS at Ages 25−44

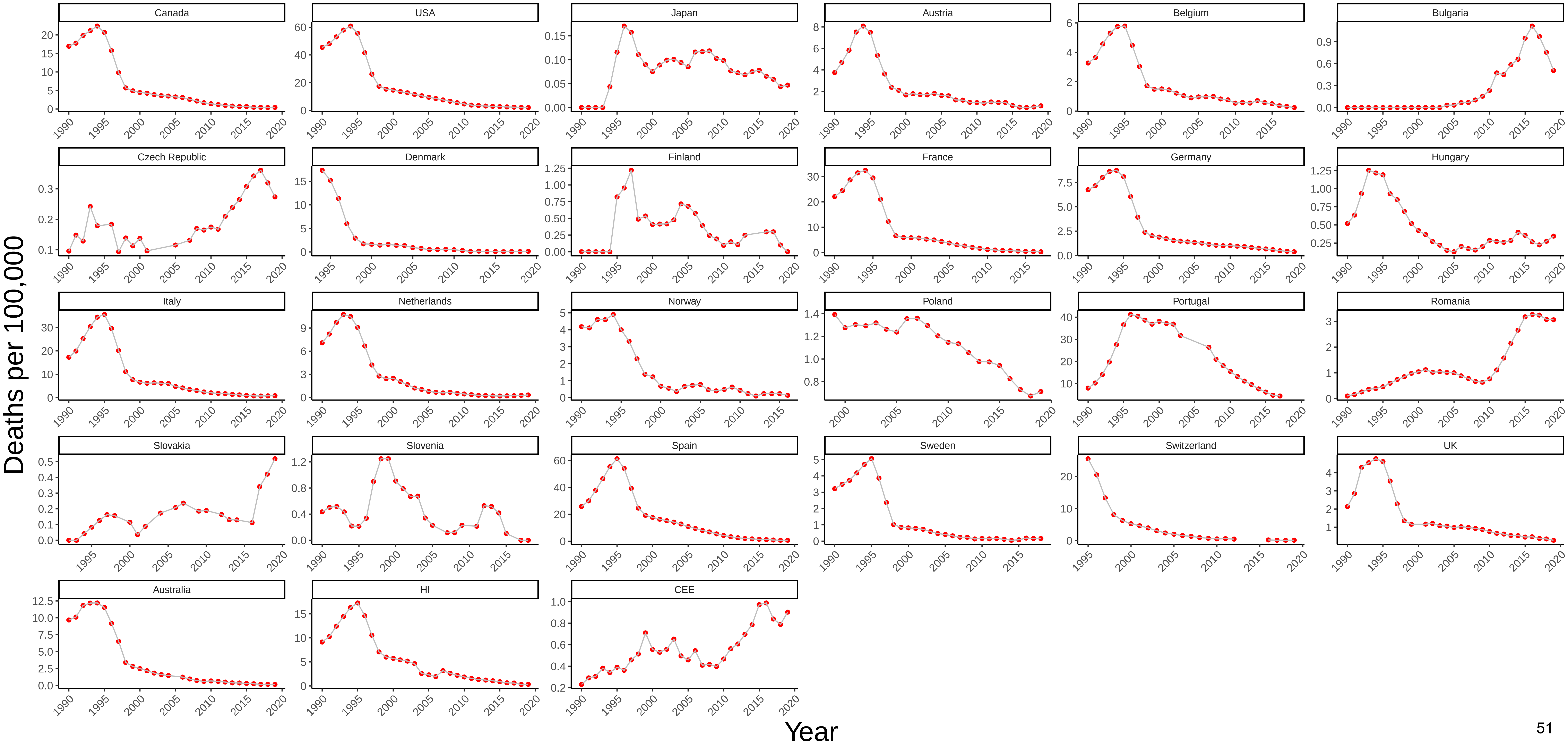

Figure 4 Three−Year Moving Average of Female Mortality from HIV/AIDS at Ages 25−44

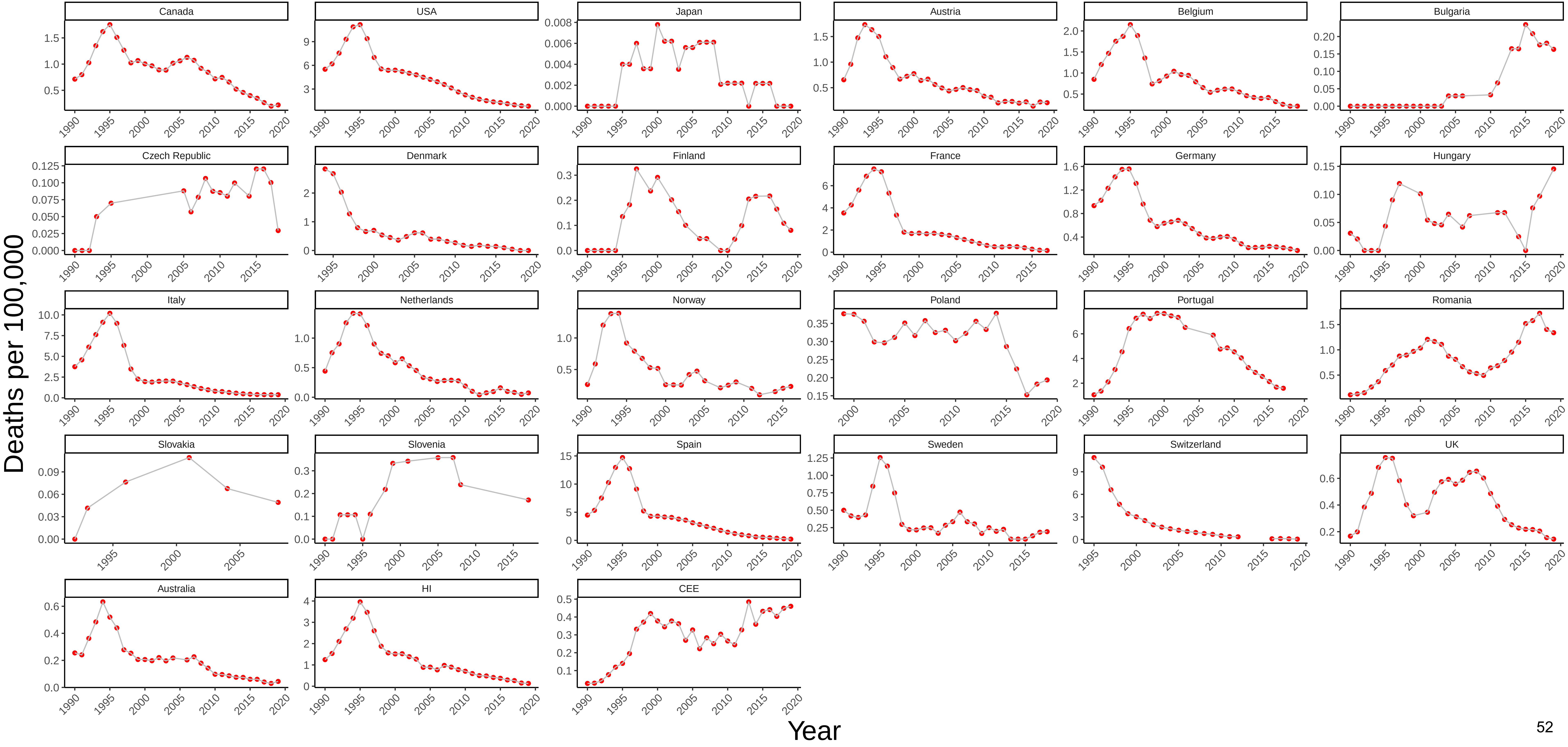

Figure 5 Three−Year Moving Average of Male Mortality from Respiratory Diseases at Ages 25−44

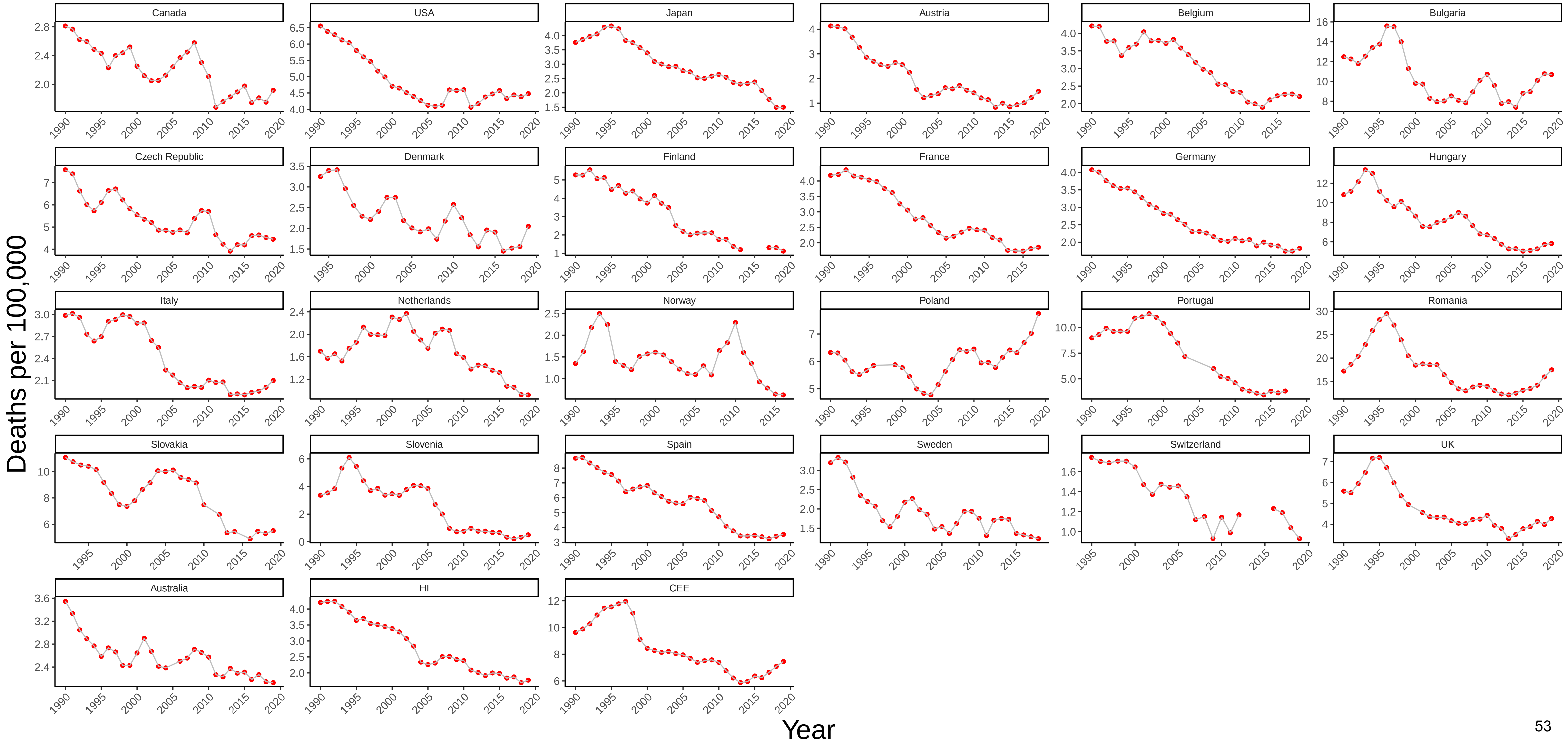

Figure 6 Three−Year Moving Average of Female Mortality from Respiratory Diseases at Ages 25−44

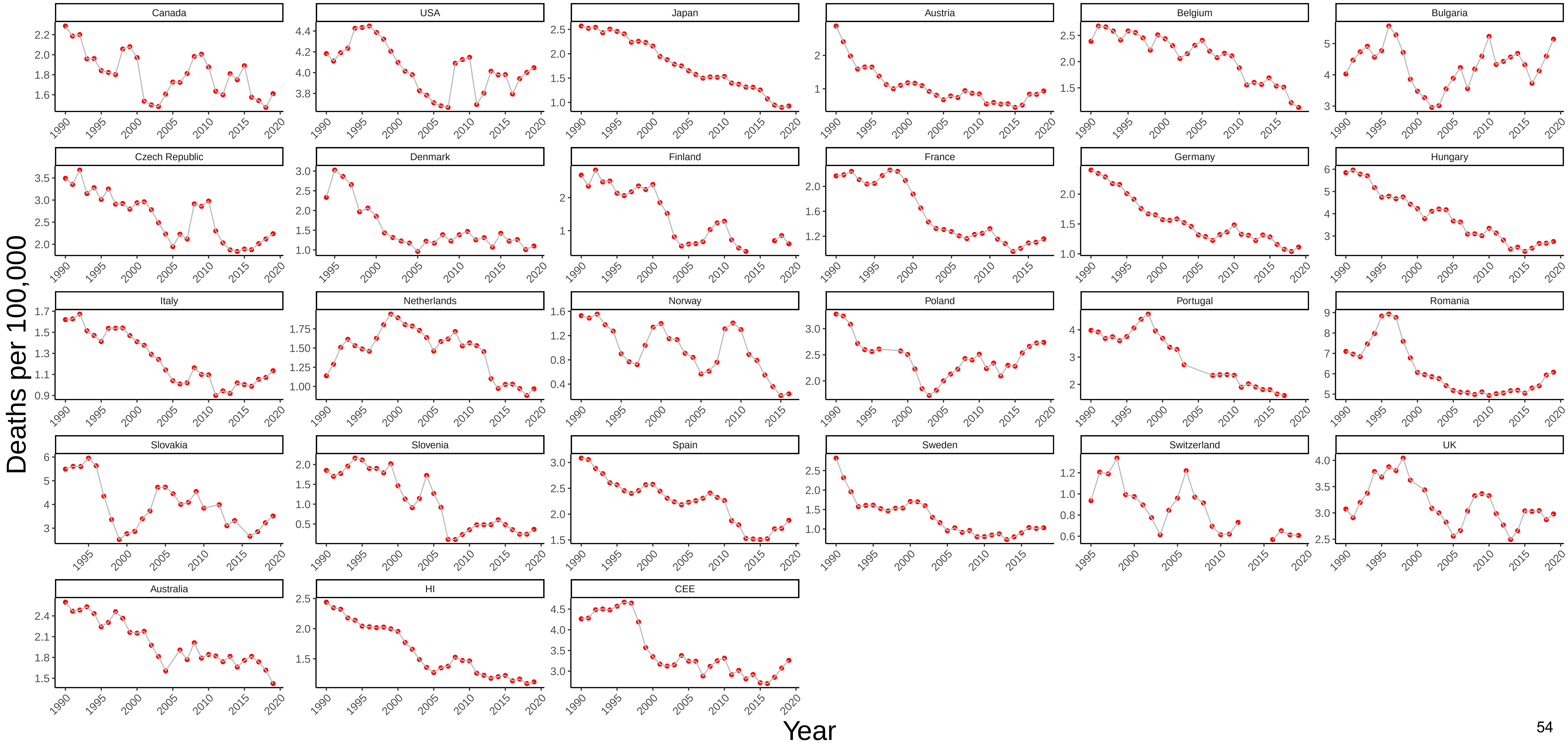

### Figure 7 Three−Year Moving Average of Male Mortality from Trachea/Bronchus, Lung Cancers at Ages 25−44

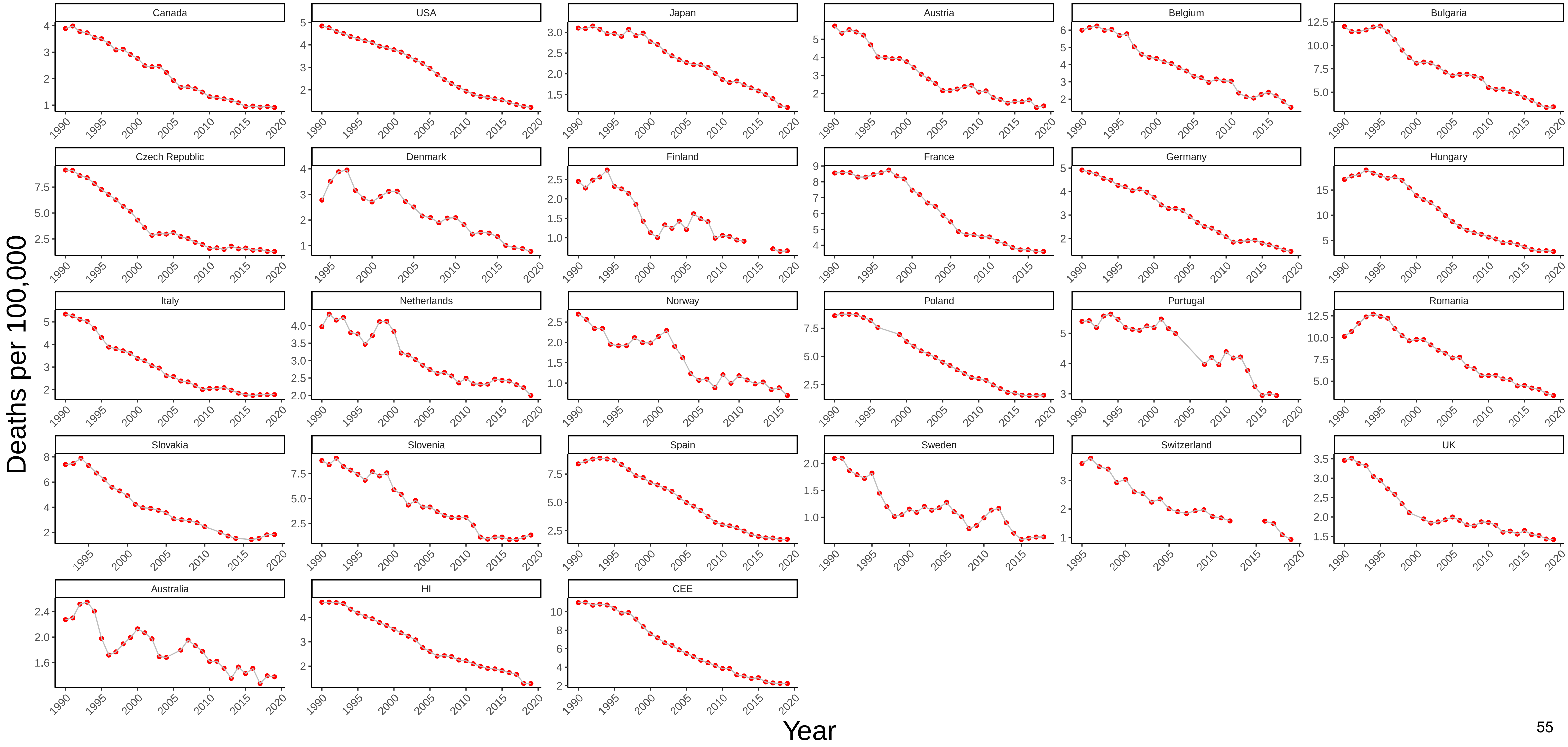

Figure 8 Three−Year Moving Average of Female Mortality from Trachea/Bronchus, Lung Cancers at Ages 25−44

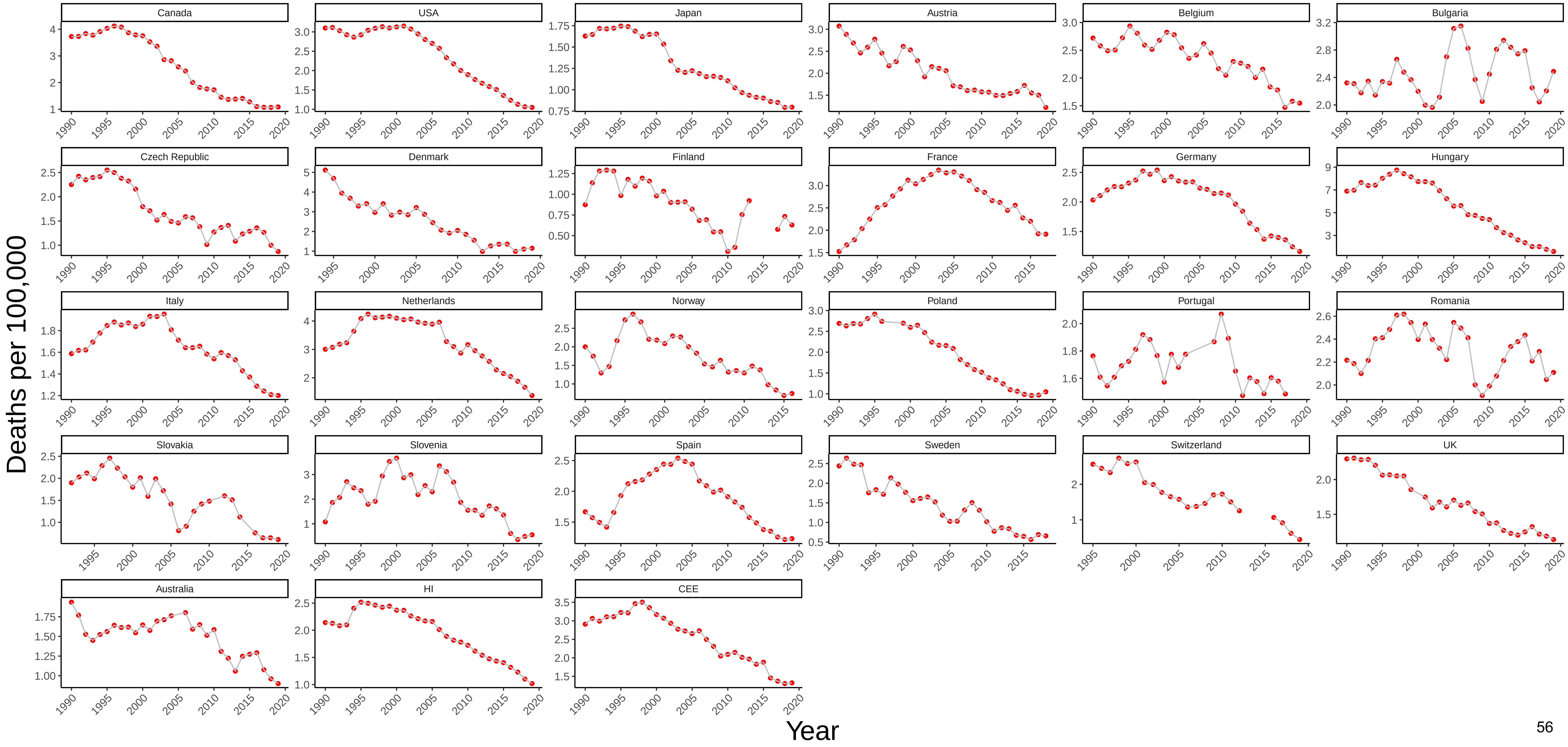

### Figure 9 Three−Year Moving Average of Male Mortality from All Other Cancers at Ages 25−44

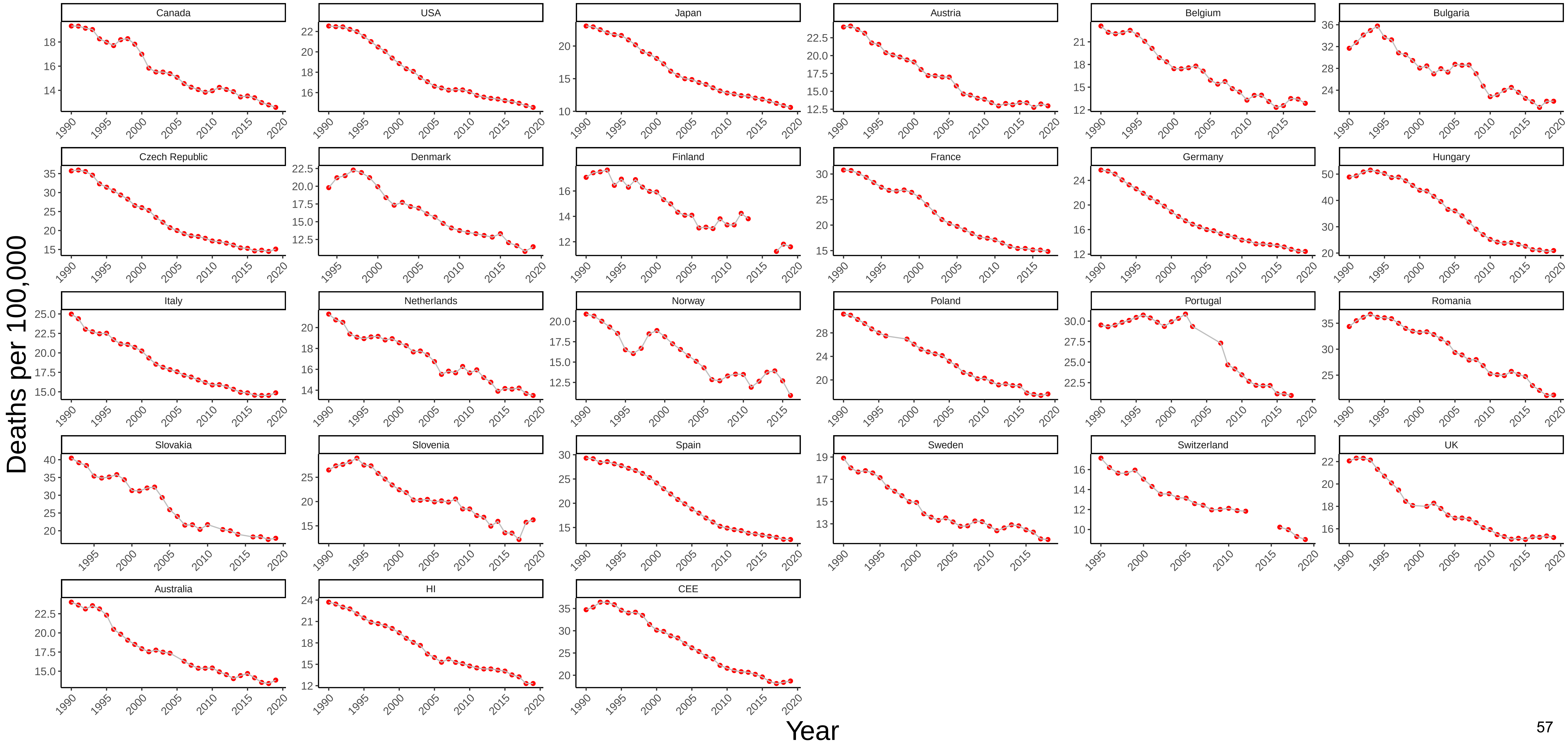

### Figure 10 Three−Year Moving Average of Female Mortality from All Other Cancers at Ages 25−44

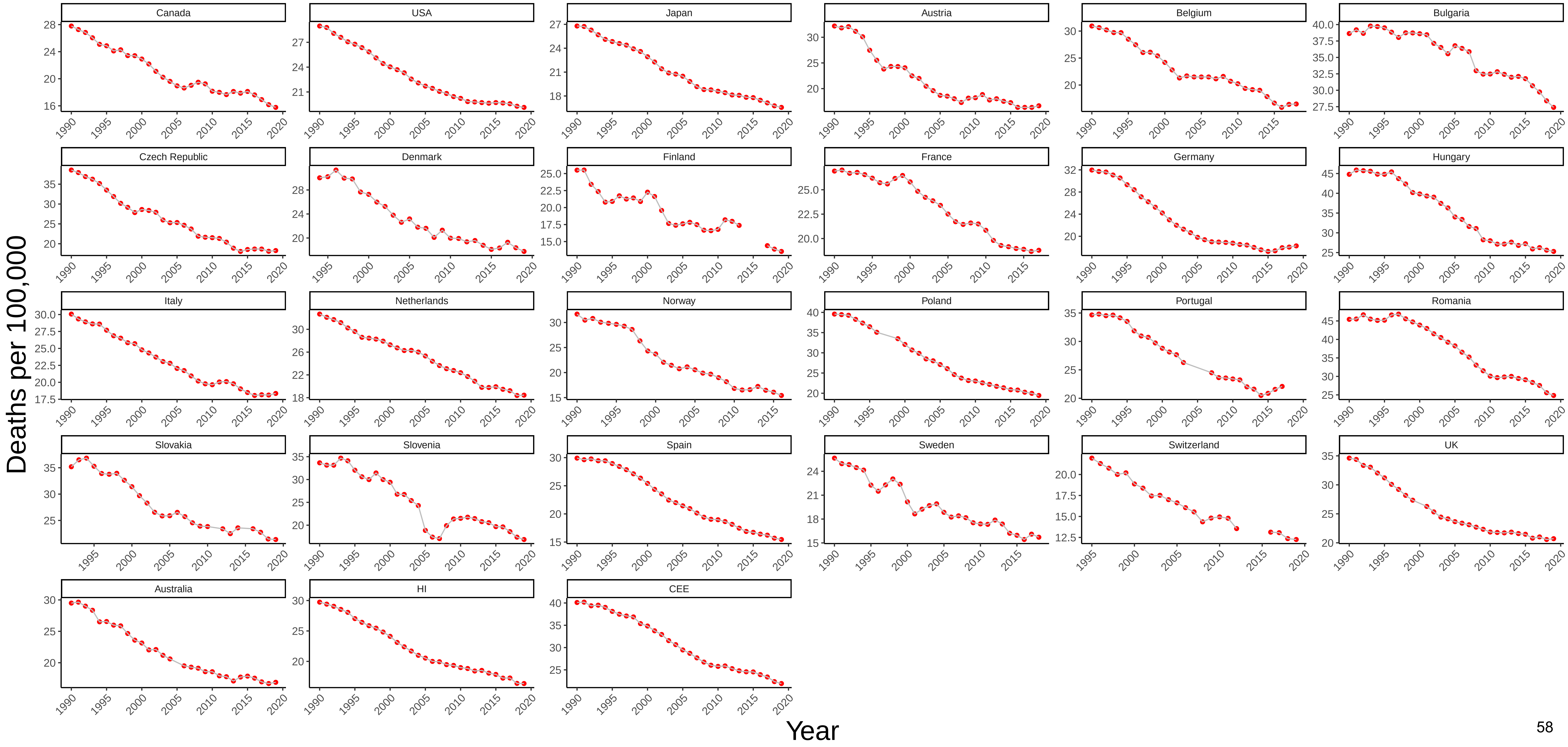

Figure 11 Three−Year Moving Average of Male Mortality from Nervous System Diseases at Ages 25−44

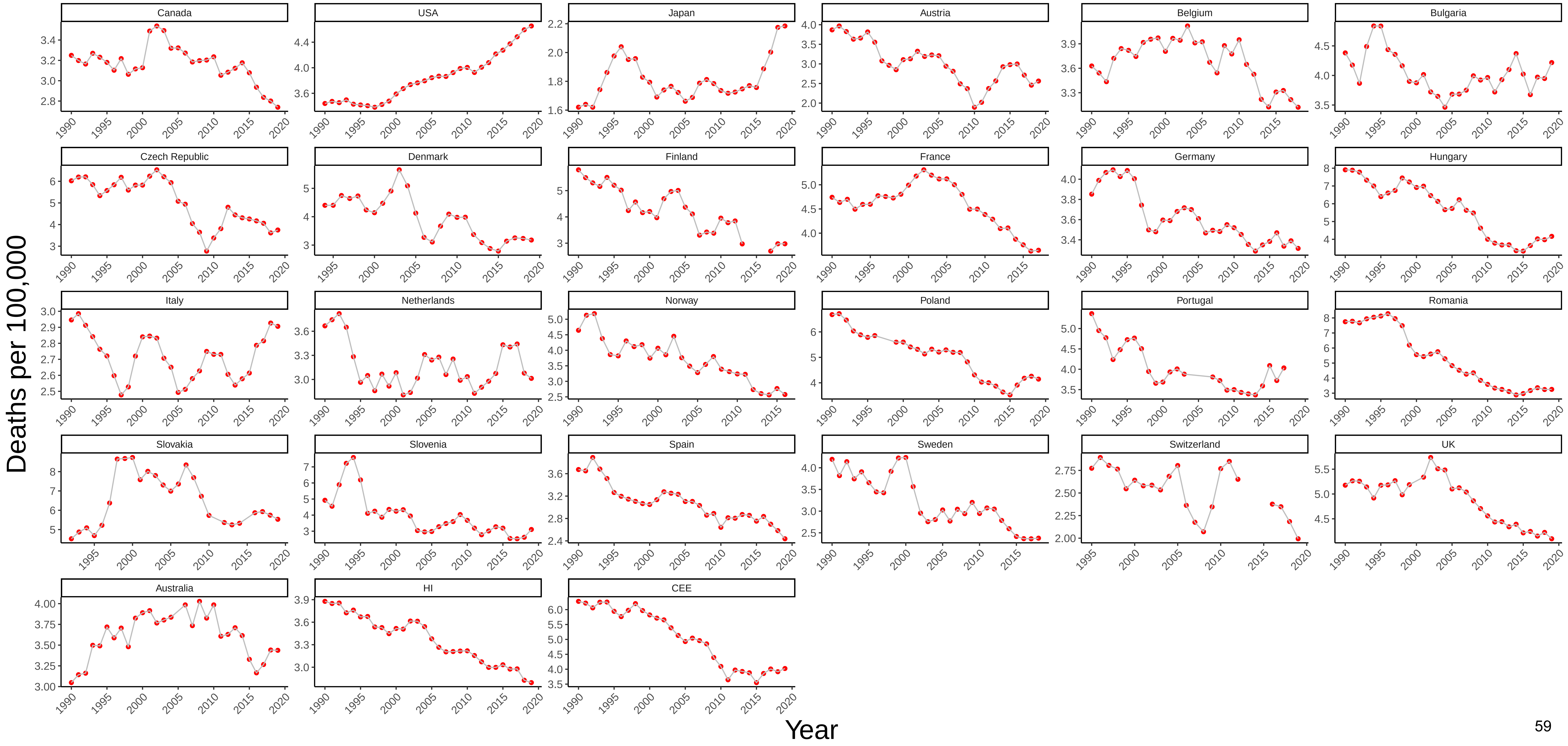

### Figure 12 Three−Year Moving Average of Female Mortality from Nervous System Diseases at Ages 25−44

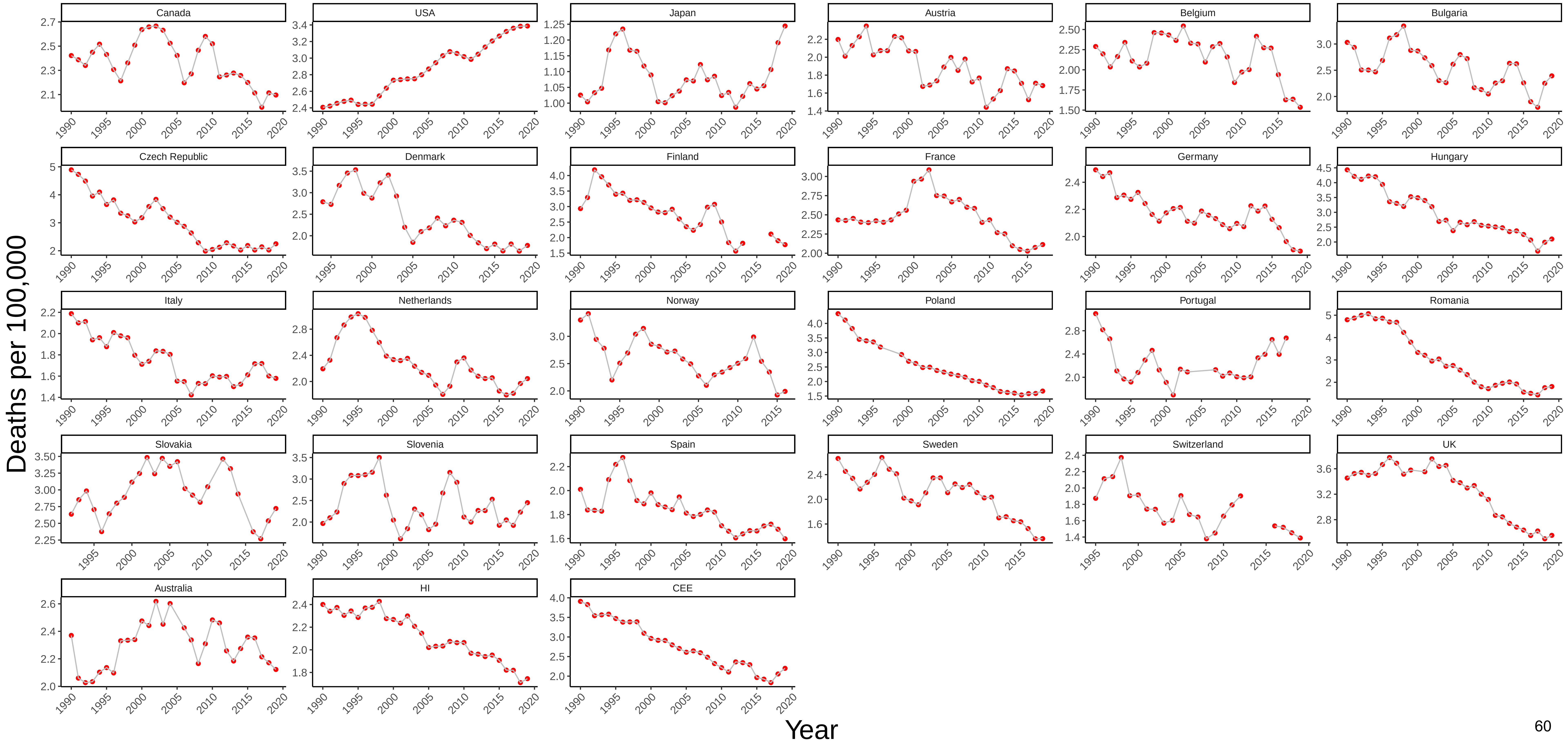

Figure 13 Three−Year Moving Average of Male Mortality from Metabolic Diseases at Ages 25−44

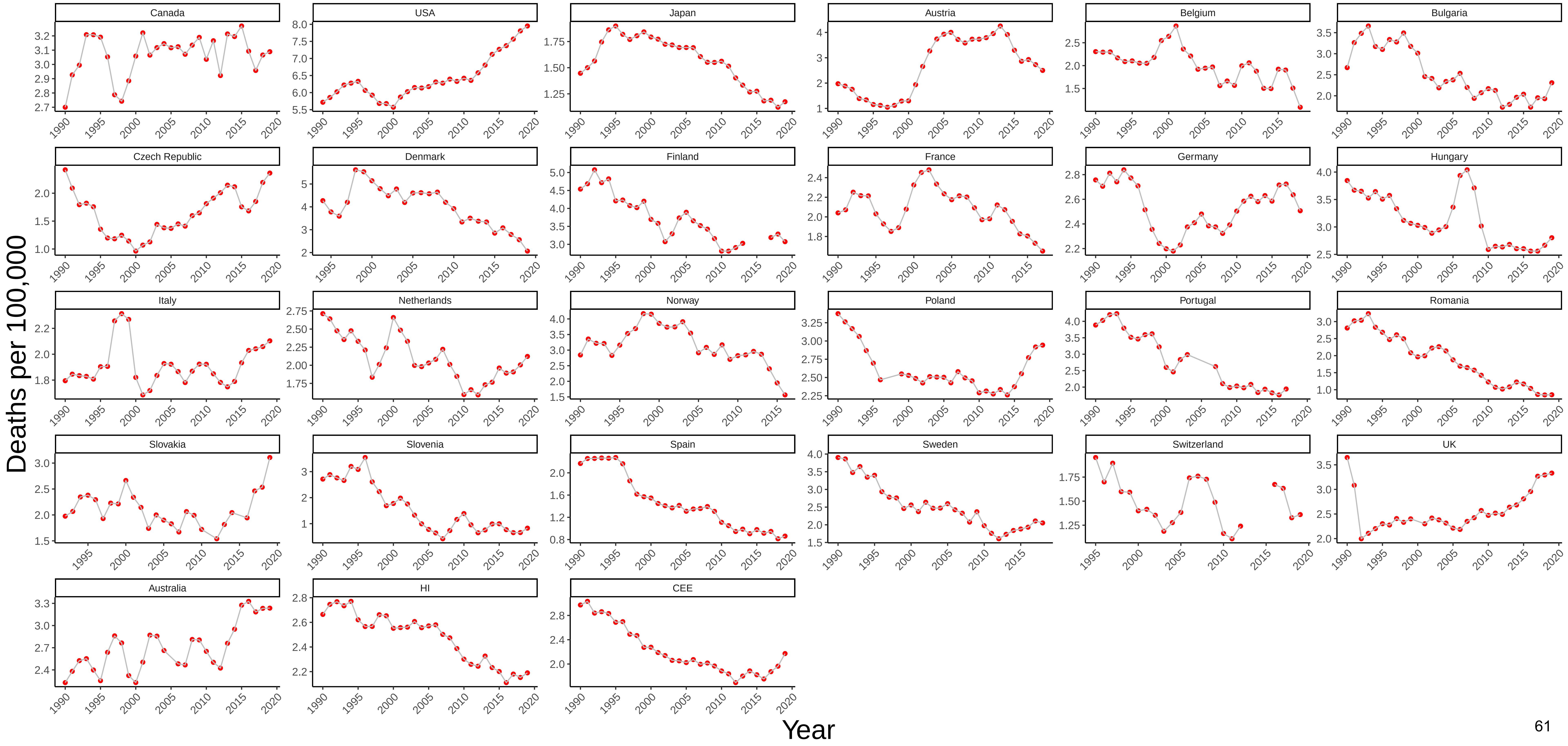

### Figure 14 Three−Year Moving Average of Female Mortality from Metabolic Diseases at Ages 25−44

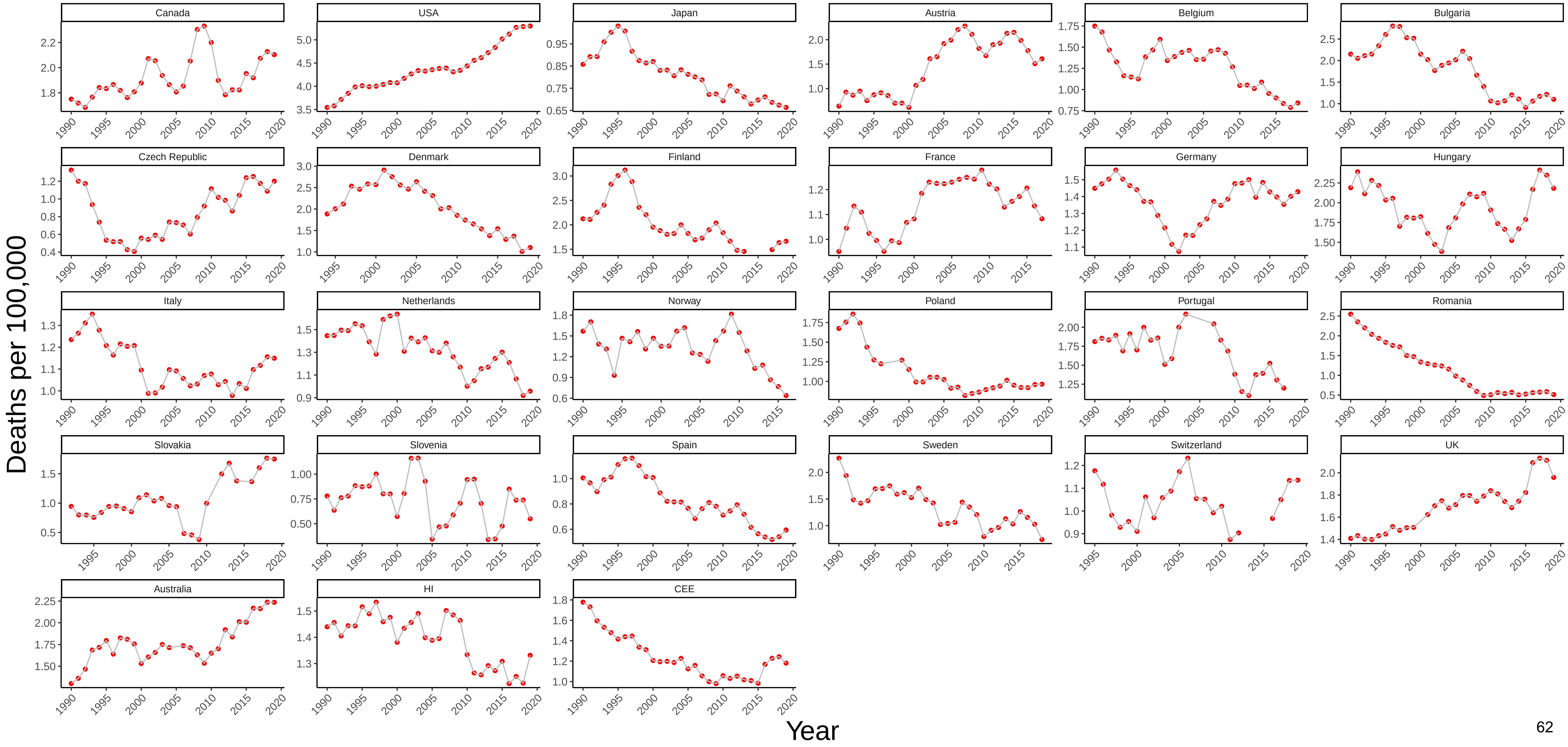

Figure 15 Three−Year Moving Average of Male Mortality from Cardiovascular Disease at Ages 25−44

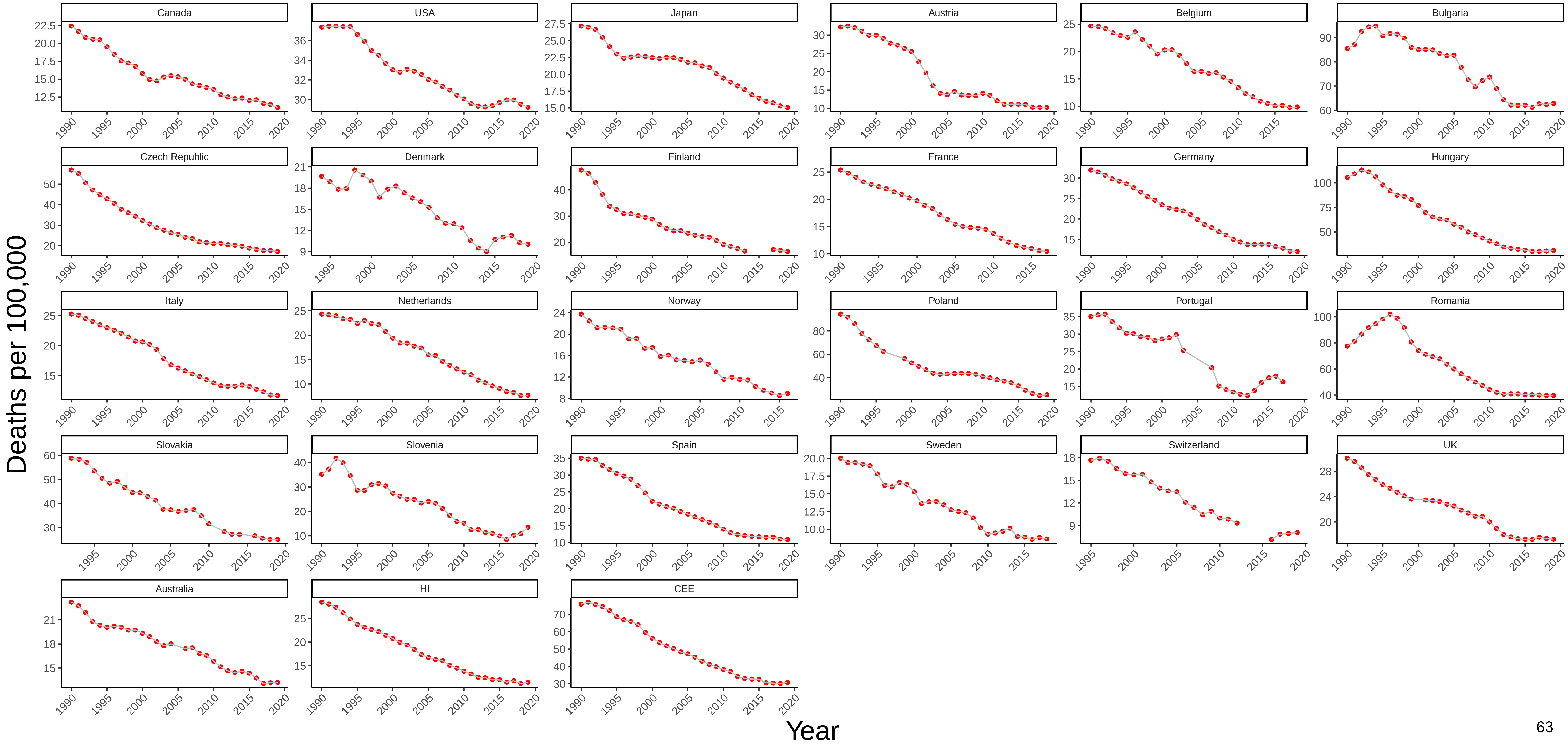

### Figure 16 Three−Year Moving Average of Female Mortality from Cardiovascular Disease at Ages 25−44

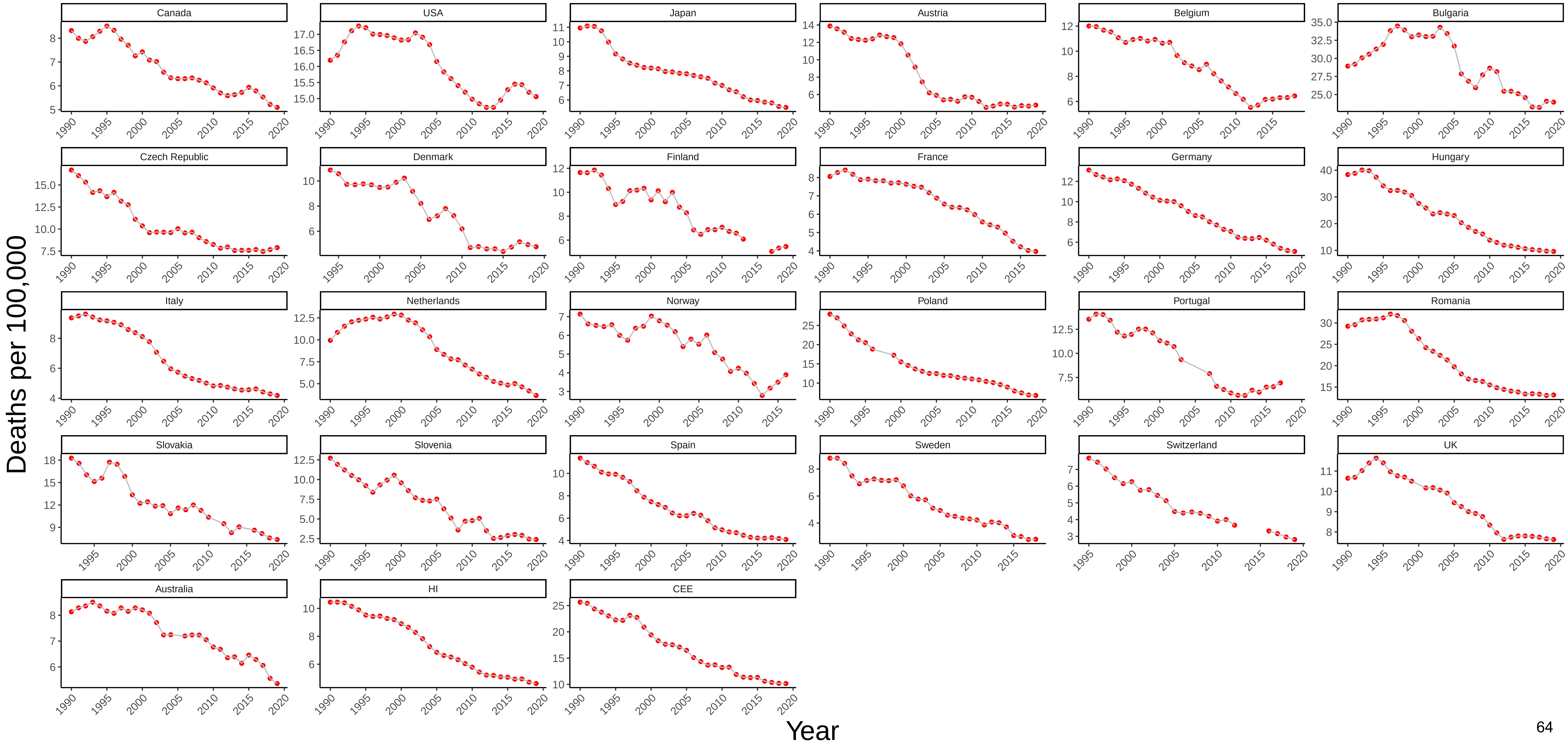

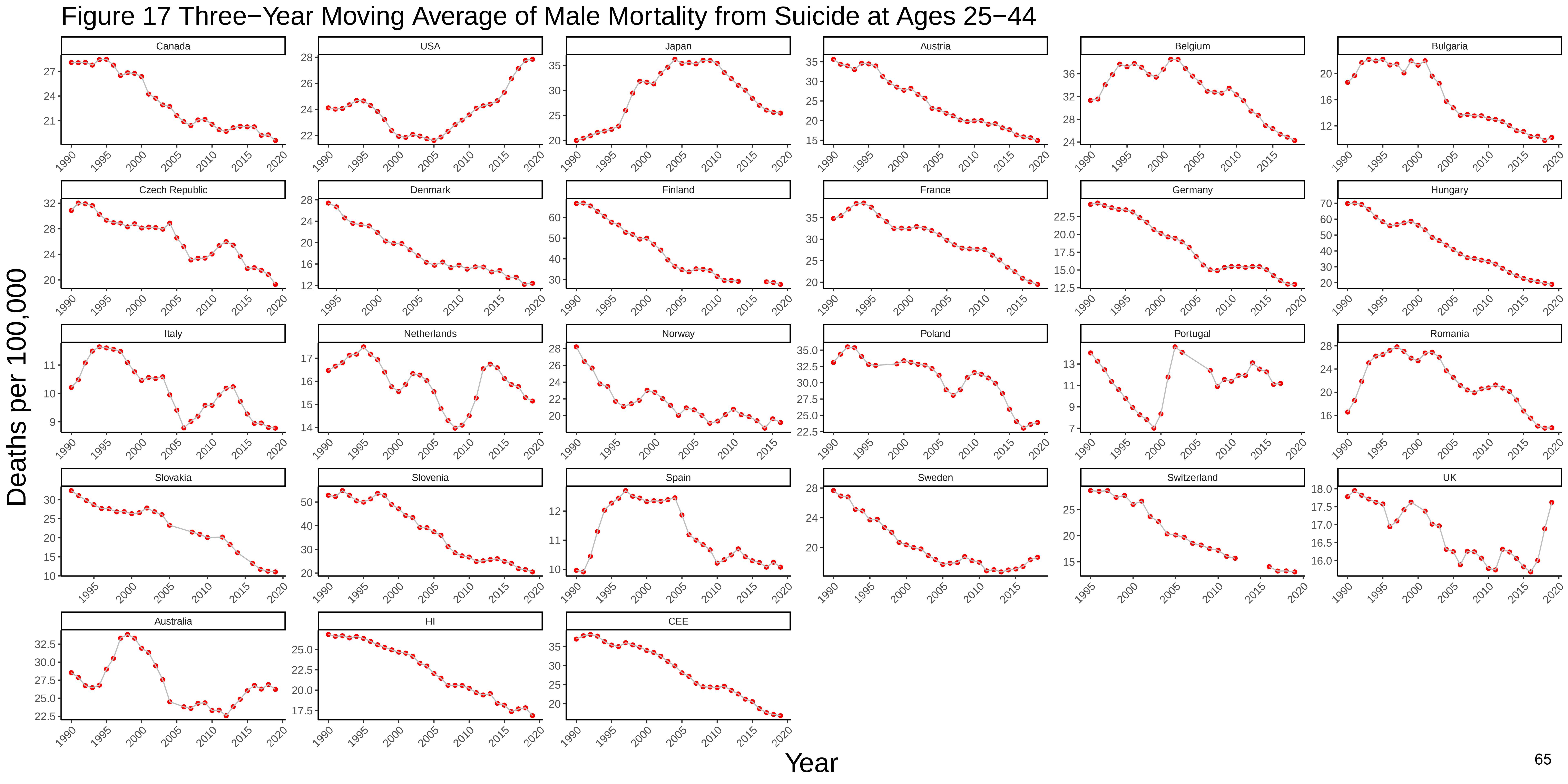
Figure 17 Three−Year Moving Average of Male Mortality from Suicide at Ages 25−44
Canada
USA
Japan
Austria
Belgium
Bulgaria
Czech Republic
Denmark
Finland
France
Germany
Hungary
Italy
Netherlands
Norway
Poland
Portugal
Romania
Slovakia
Slovenia
Spain
Sweden
Switzerland
UK
Australia
HI
CEE
Deaths per 100,000
Year

### Figure 18 Three−Year Moving Average of Female Mortality from Suicide at Ages 25−44

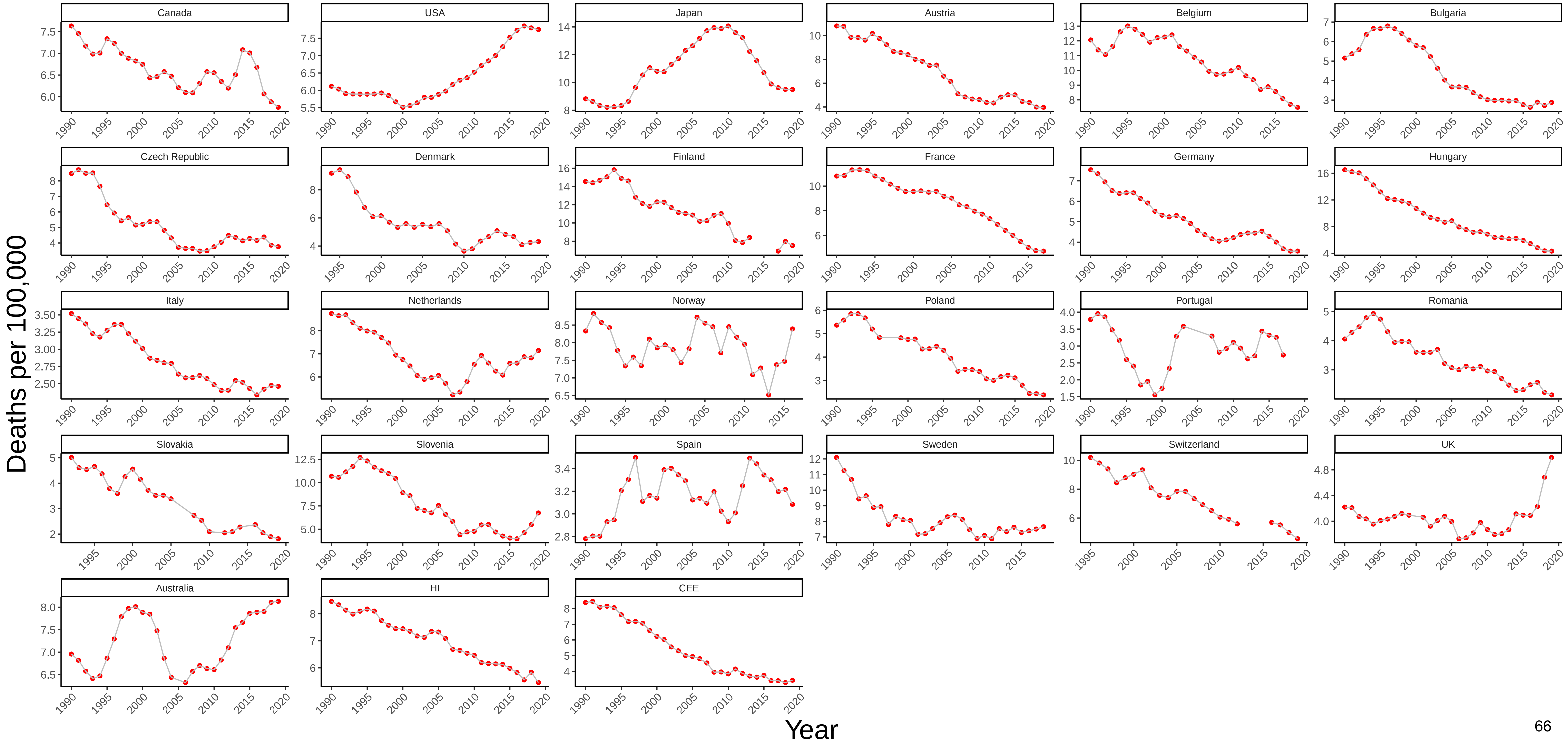

### Figure 19 Three−Year Moving Average of Male Mortality from Homicide at Ages 25−44

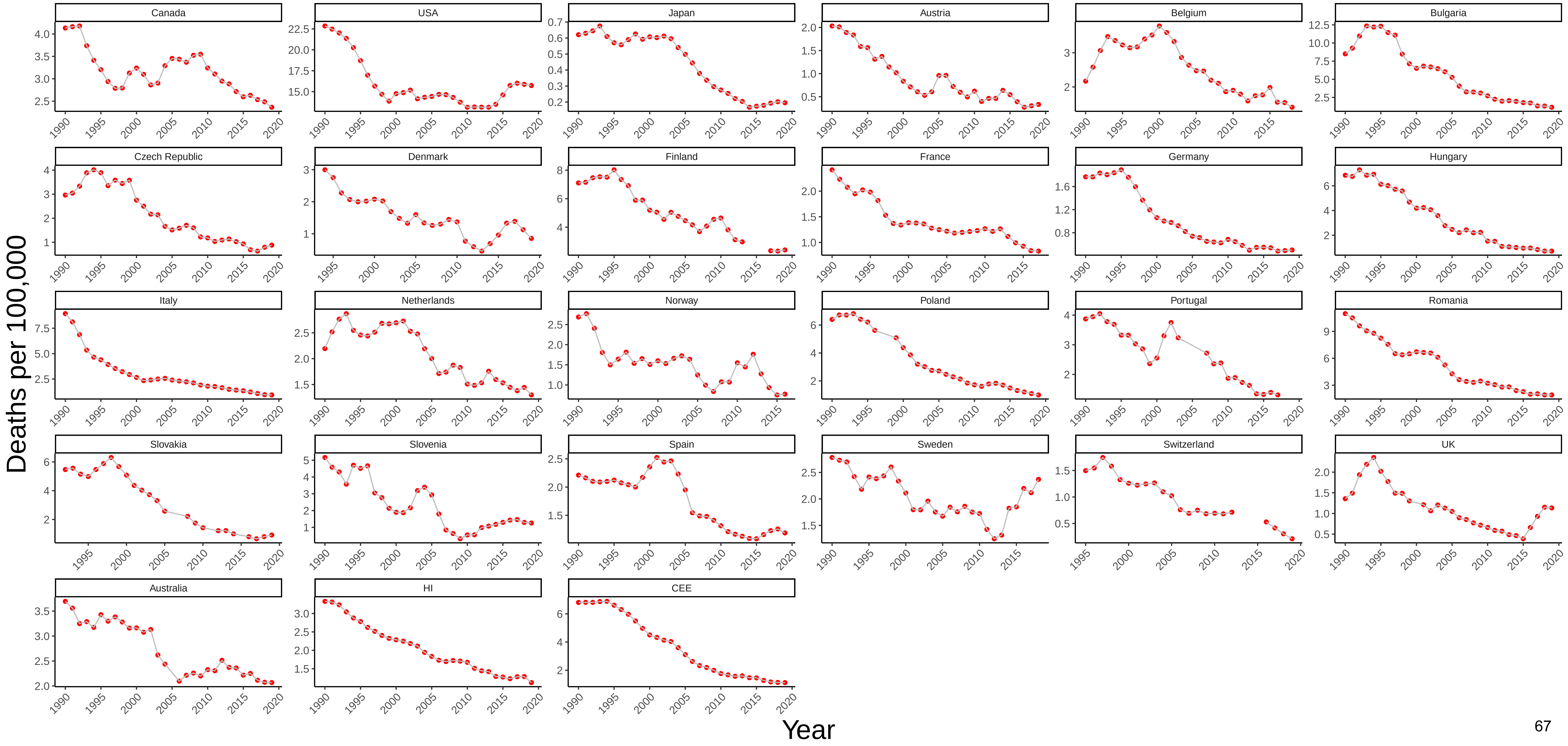

### Figure 20 Three−Year Moving Average of Female Mortality from Homicide at Ages 25−44

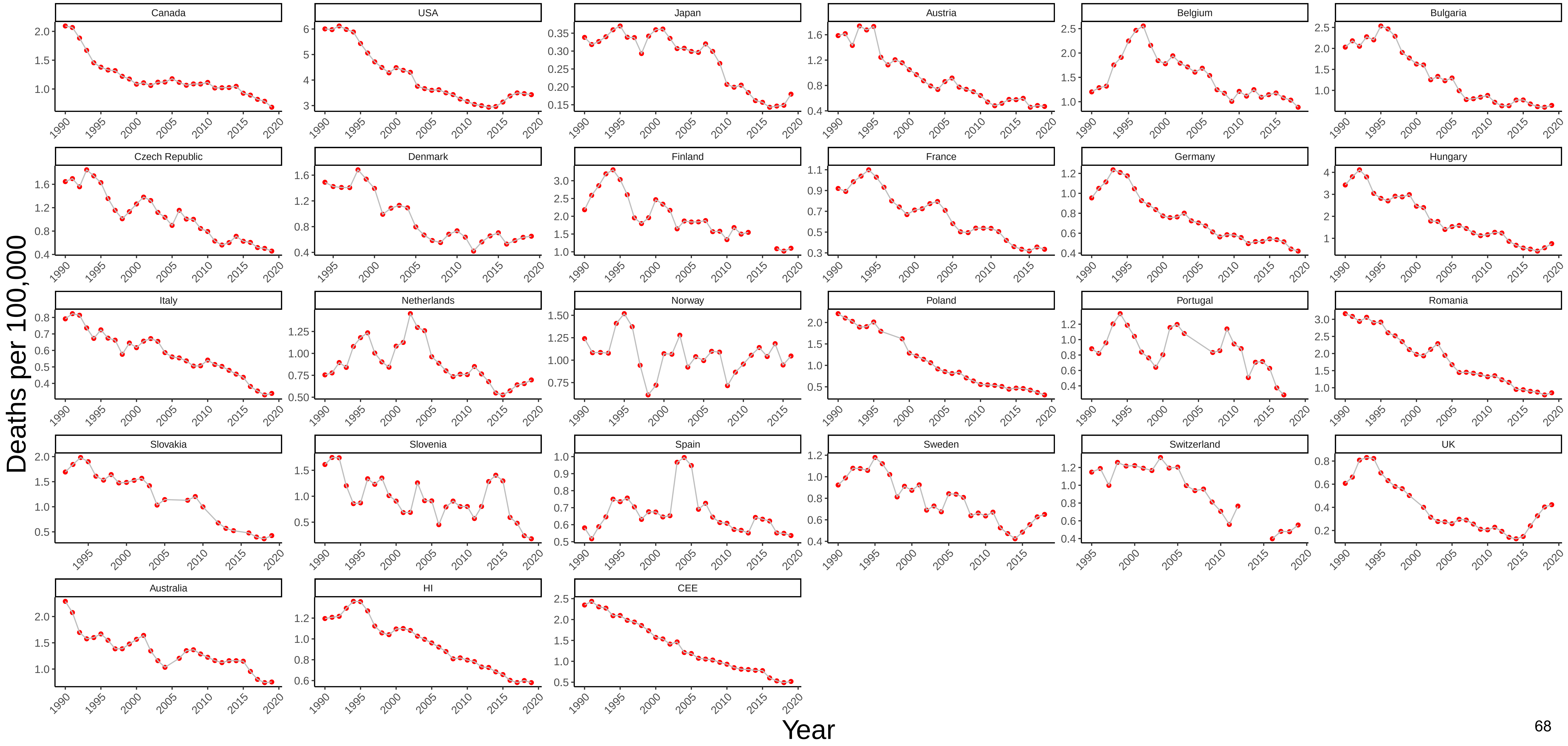

### Figure 21 Three−Year Moving Average of Male Mortality from Transport Accidents at Ages 25−44

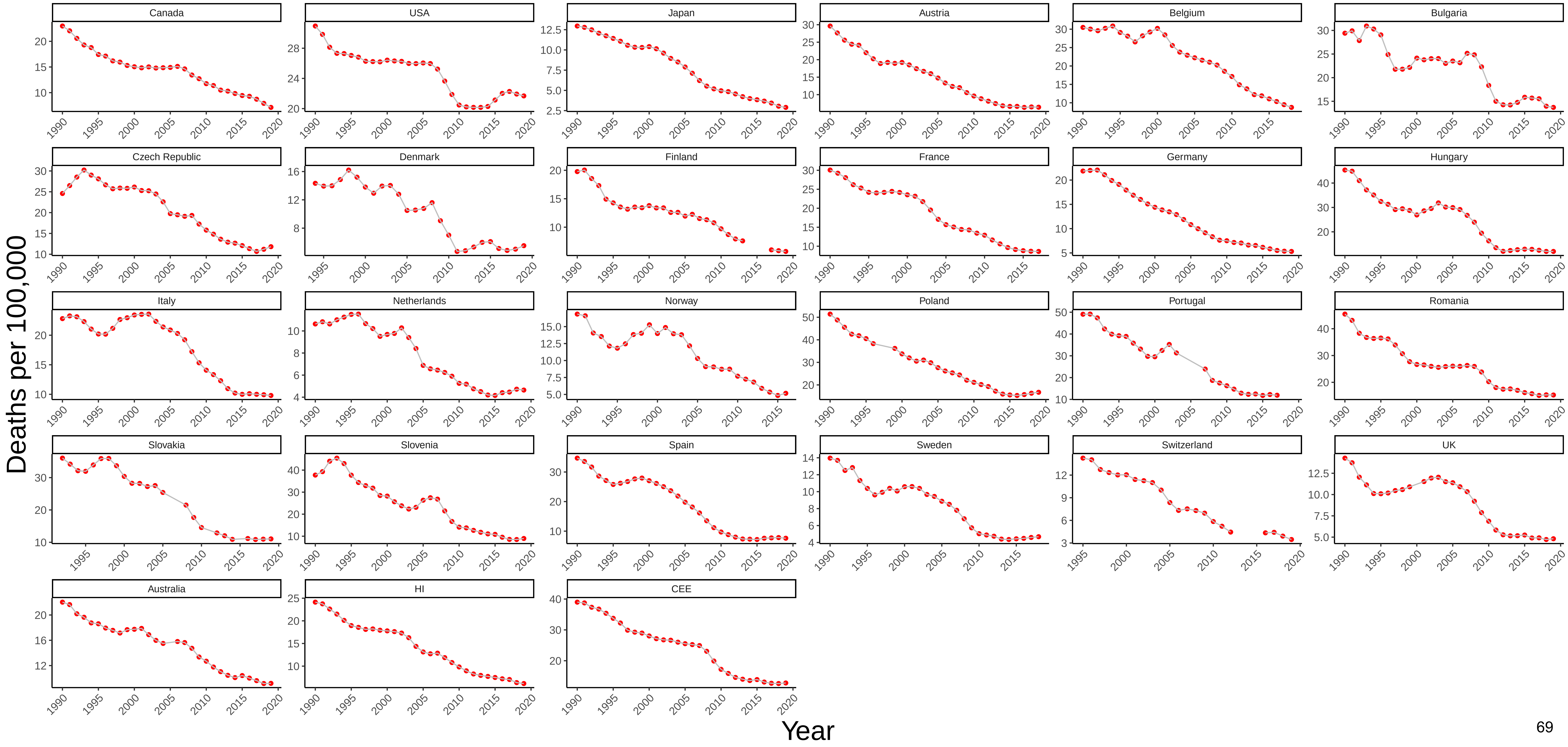

### Figure 22 Three−Year Moving Average of Female Mortality from Transport Accidents at Ages 25−44

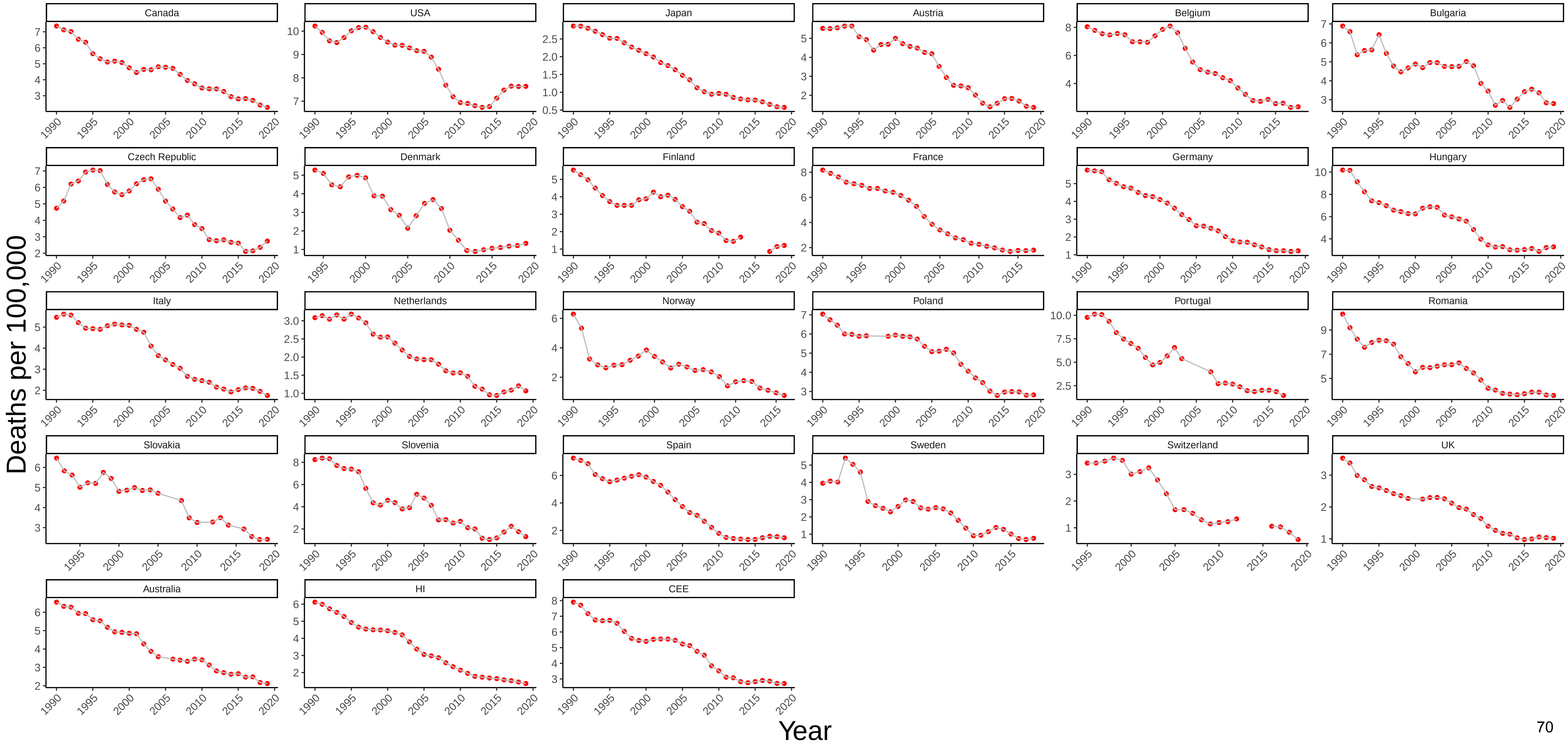

### Figure 23 Three−Year Moving Average of Male Mortality from Other External Causes at Ages 25−44

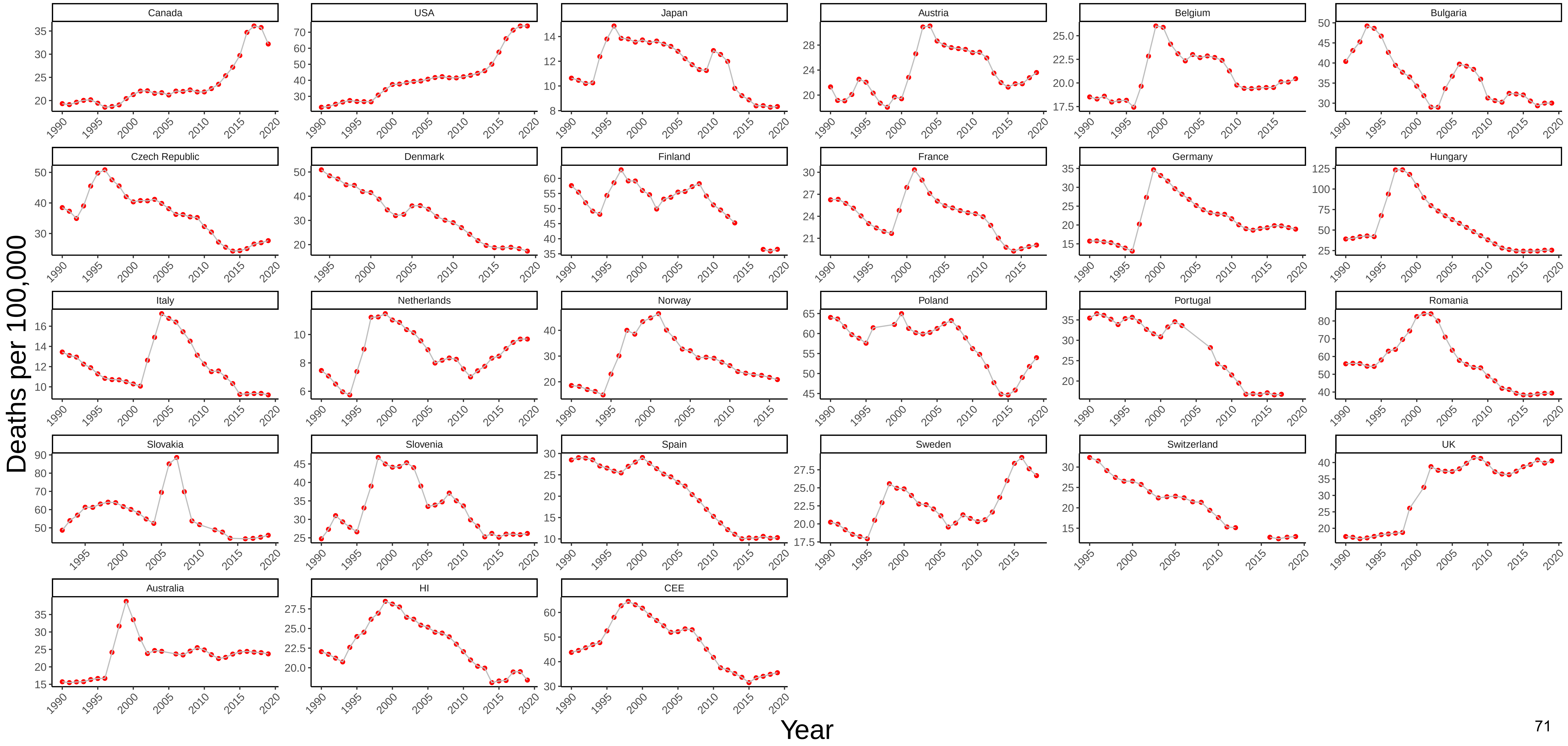

Figure 24 Three−Year Moving Average of Female Mortality from Other External Causes at Ages 25−44

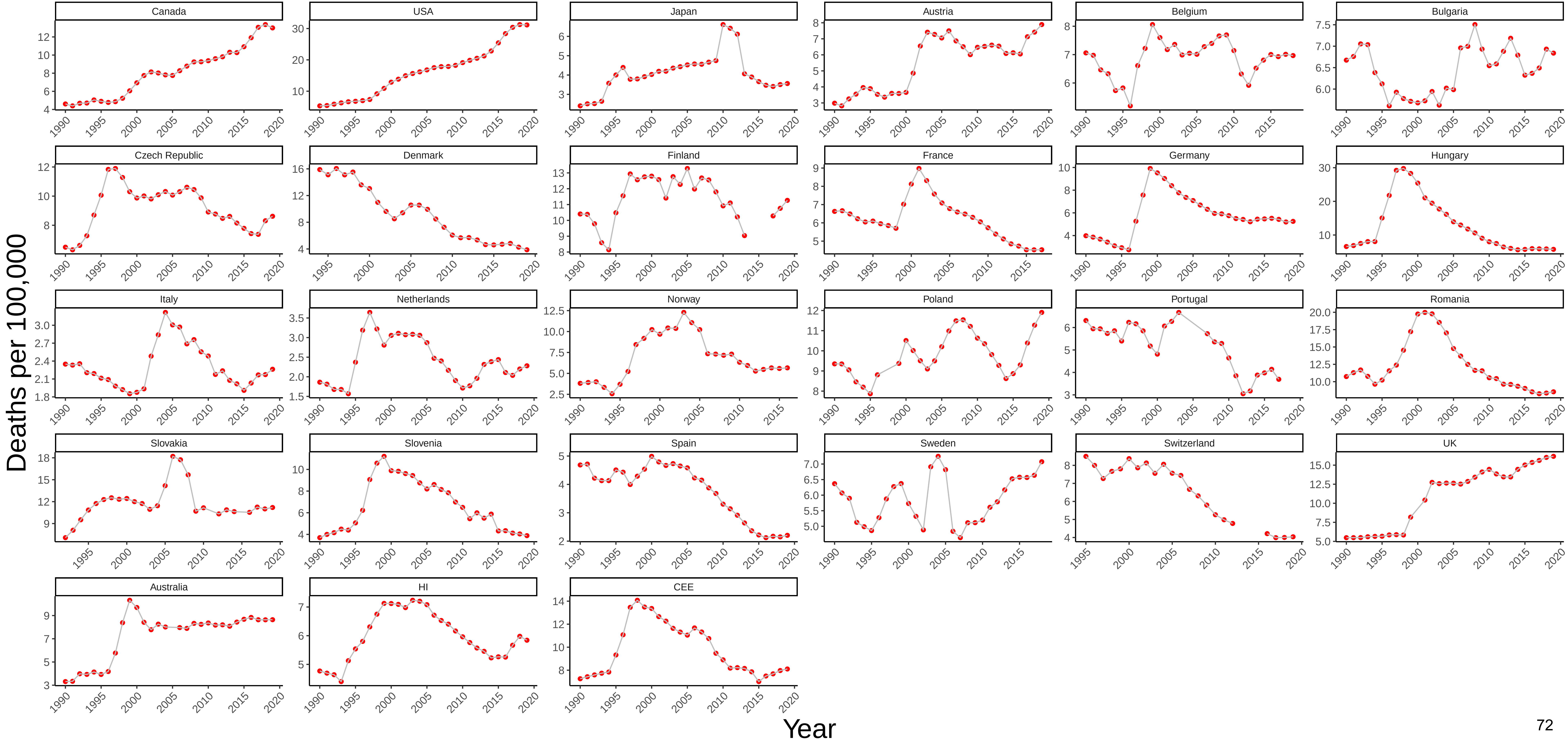

### Figure 25 Three−Year Moving Average of Male Mortality from All Other Causes at Ages 25−44

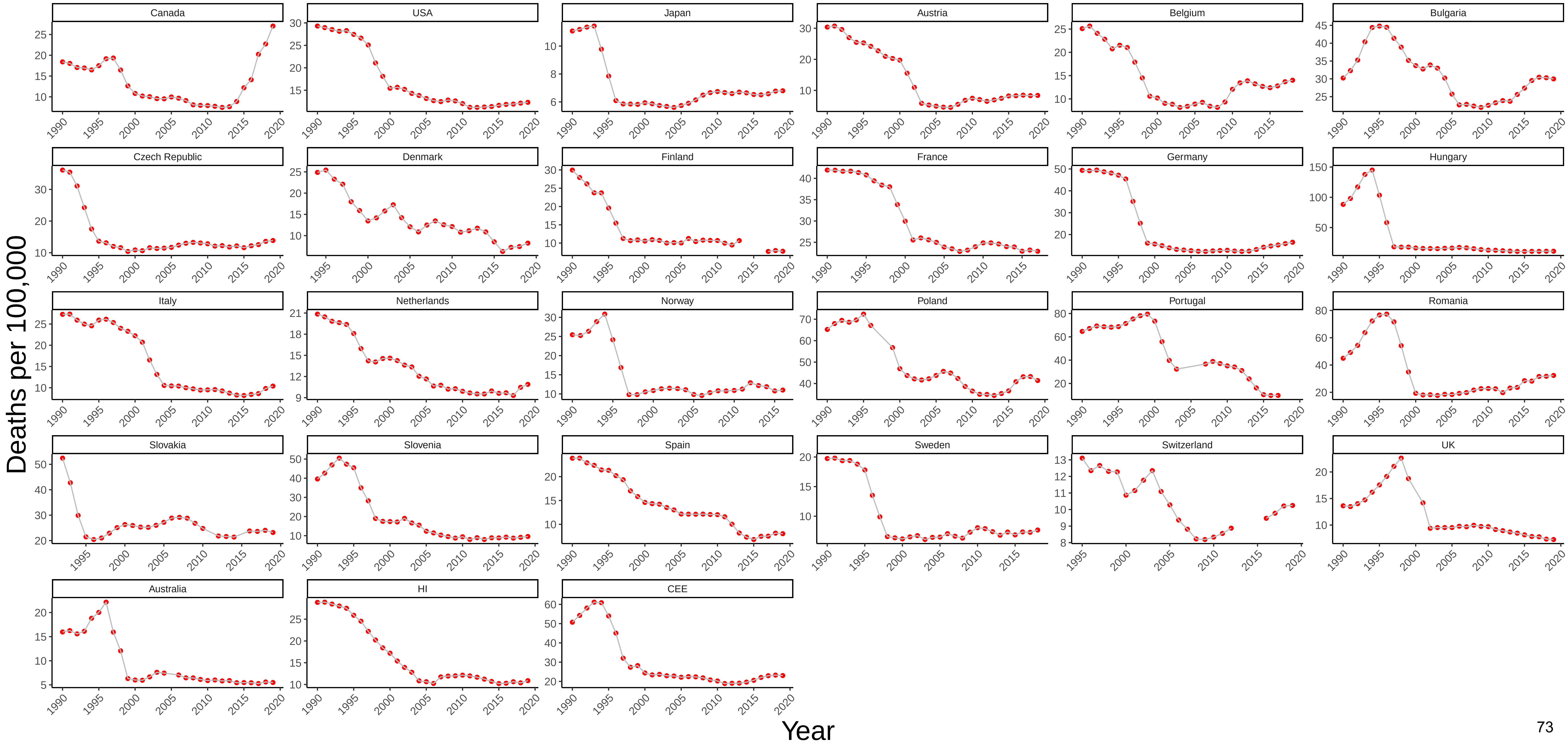

### Figure 26 Three−Year Moving Average of Female Mortality from All Other Causes at Ages 25−44

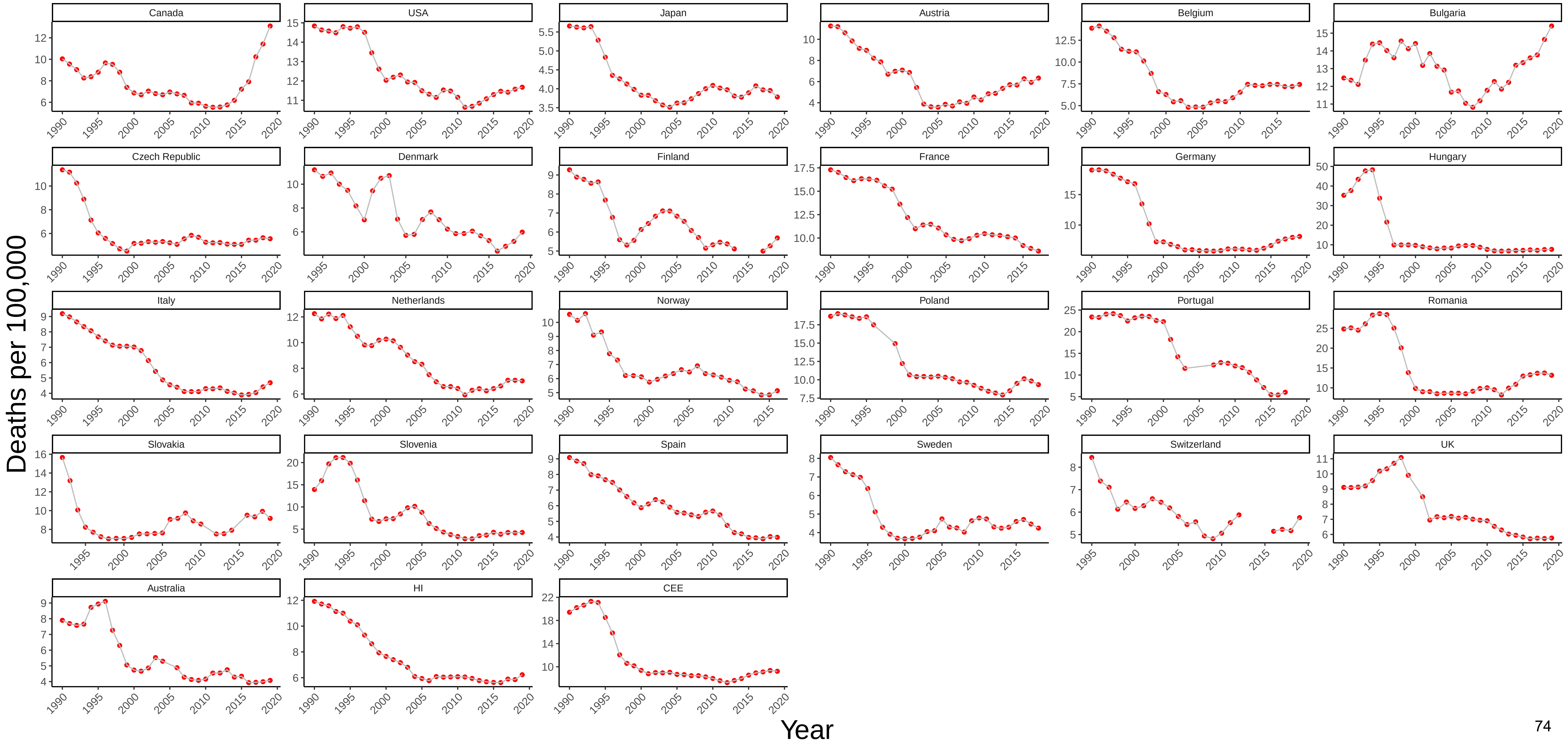

### Figure 27 Three−Year Moving Average of Male Mortality from All Causes at Ages 25−44

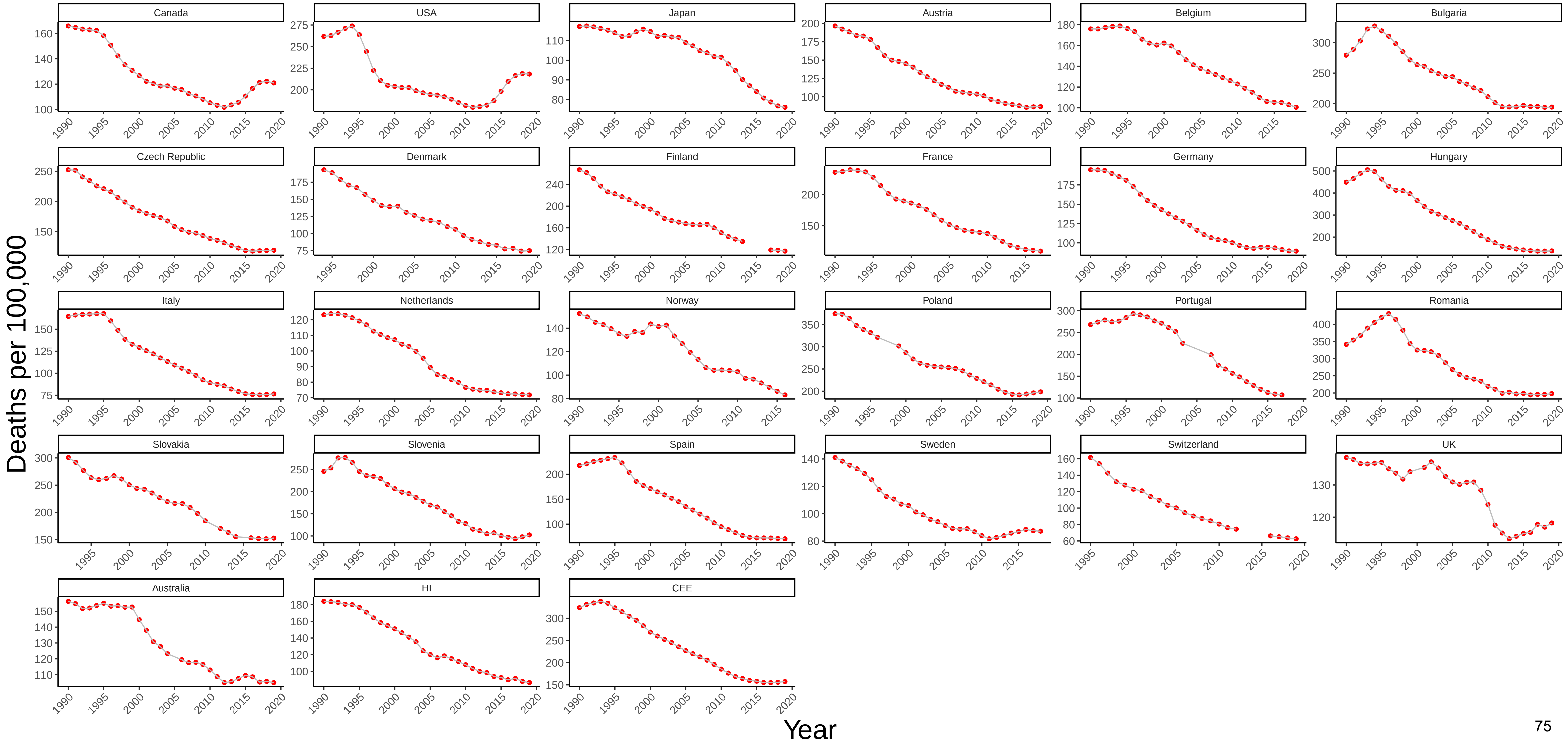

### Figure 28 Three−Year Moving Average of Female Mortality from All Causes at Ages 25−44

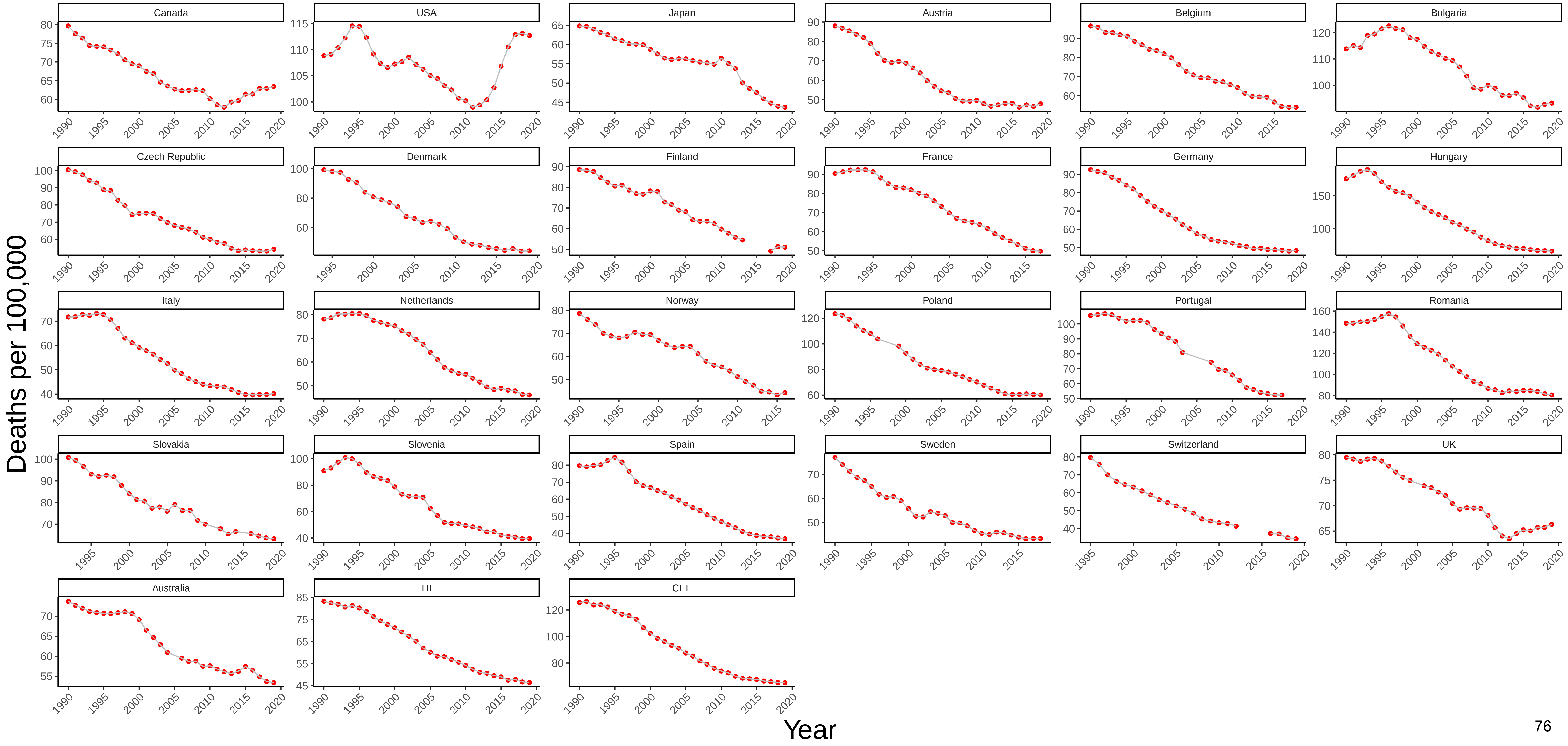

### Figure 29 Three−Year Moving Average of Male Mortality from Infectious and Parasitic Diseases at Ages 45−54

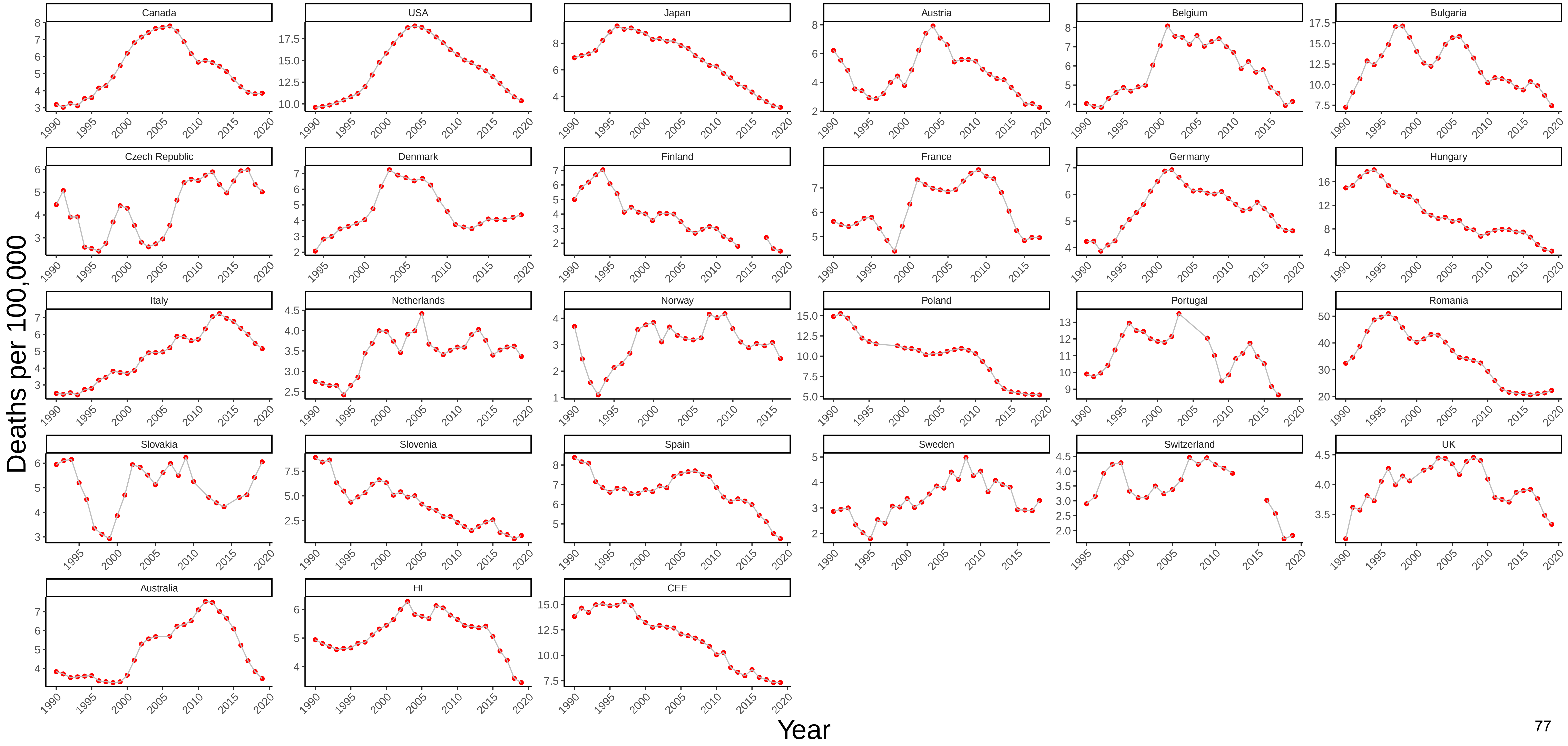

### Figure 30 Three−Year Moving Average of Female Mortality from Infectious and Parasitic Diseases at Ages 45−54

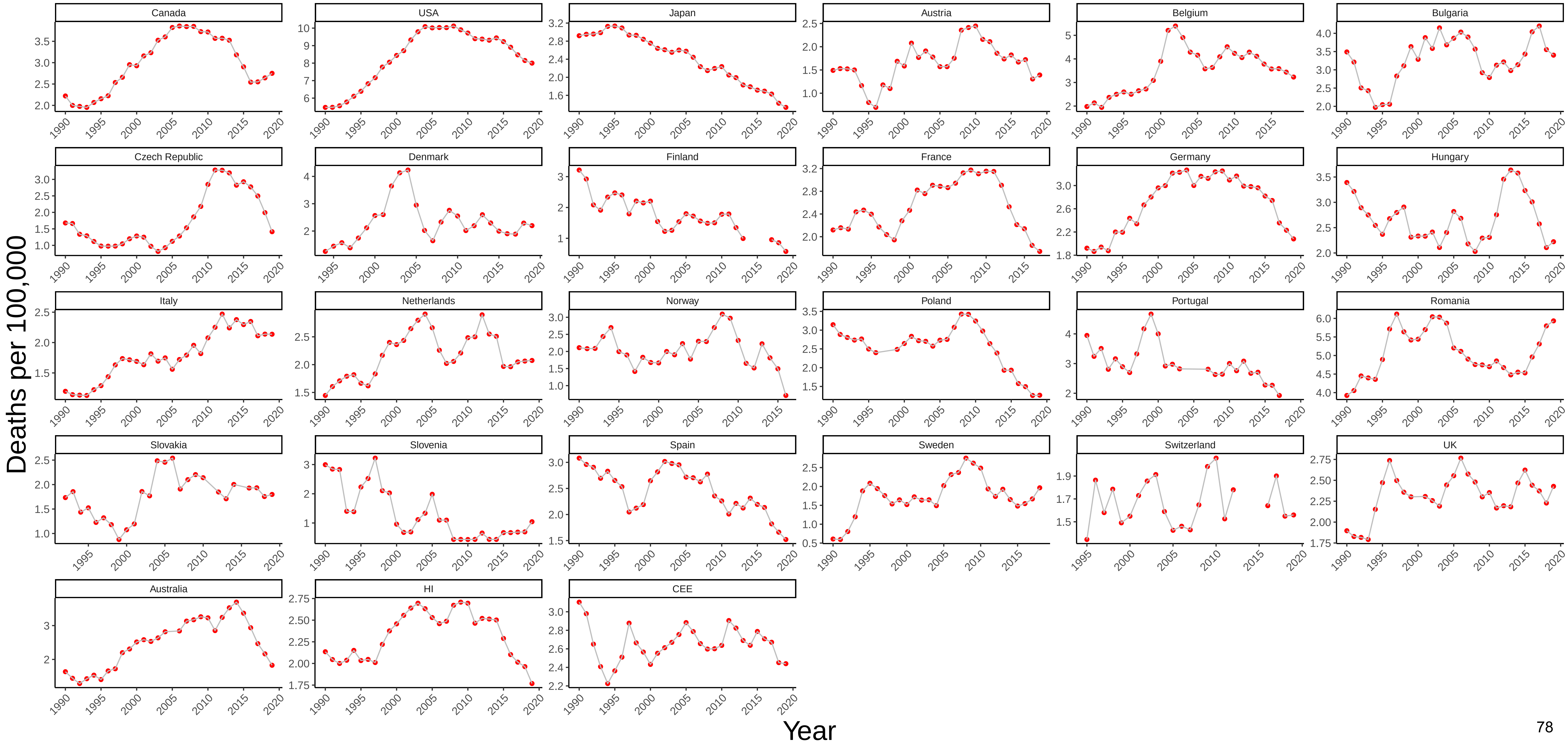

Figure 31 Three−Year Moving Average of Male Mortality from HIV/AIDS at Ages 45−54

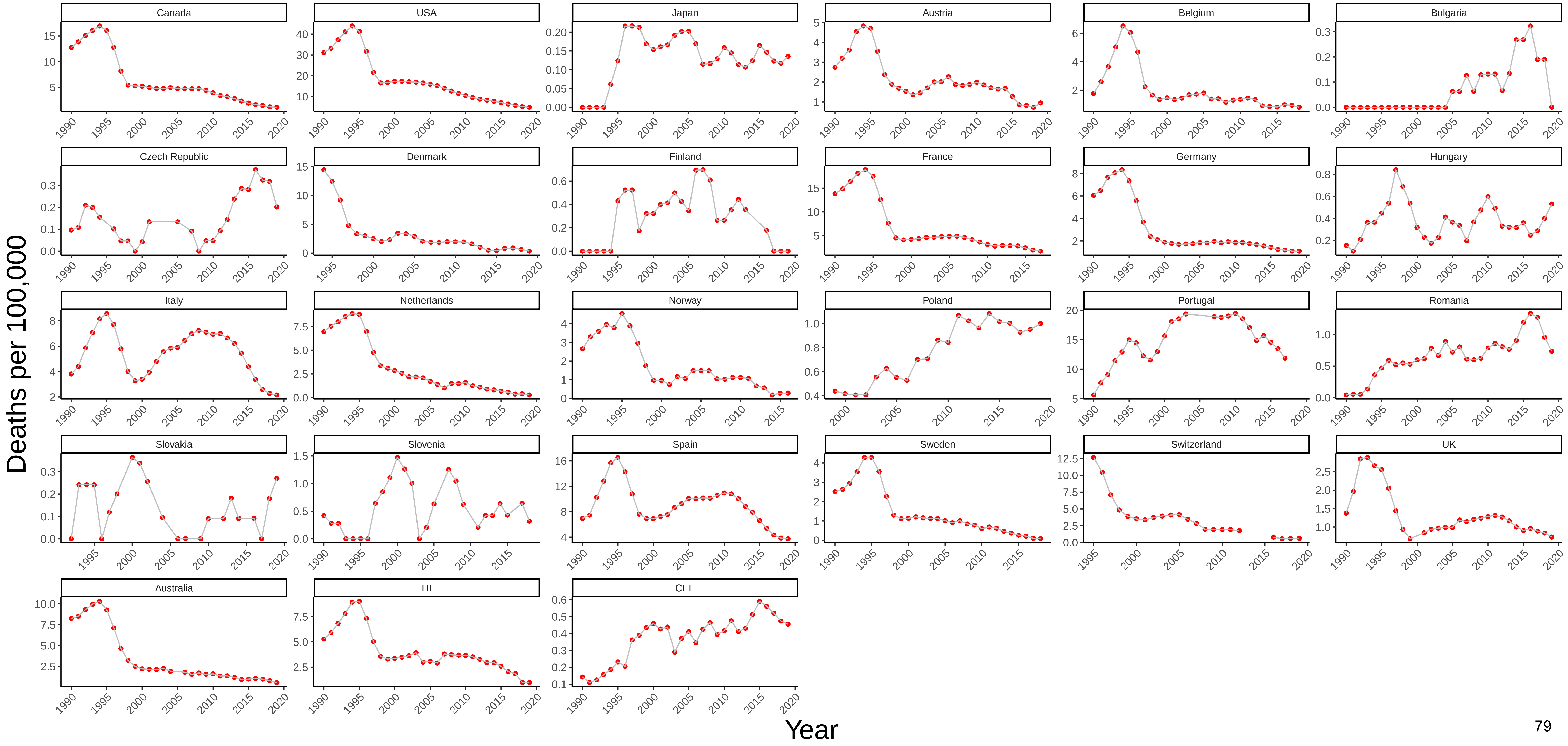

### Figure 32 Three−Year Moving Average of Female Mortality from HIV/AIDS at Ages 45−54

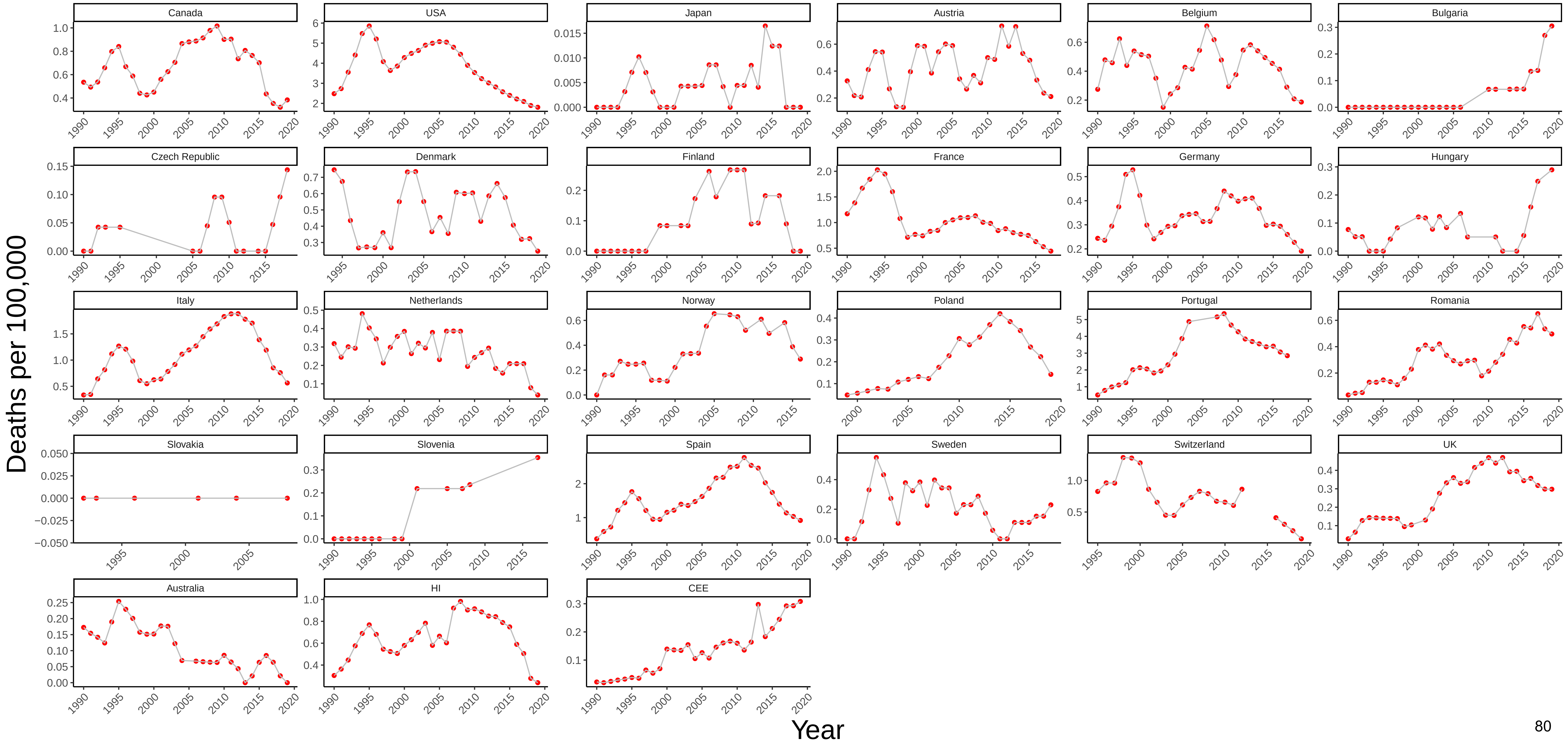

### Figure 33 Three−Year Moving Average of Male Mortality from Respiratory Diseases at Ages 45−54

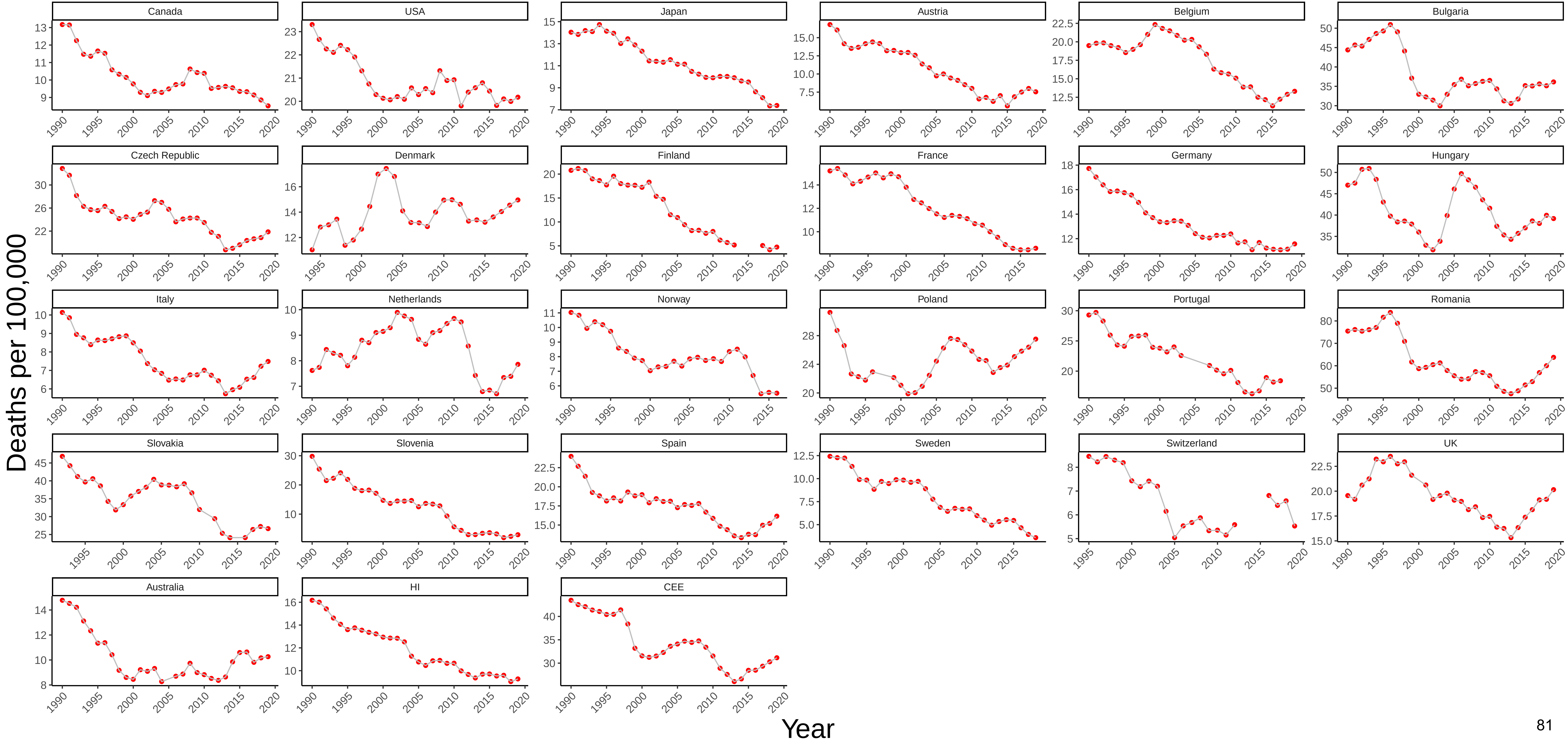

### Figure 34 Three−Year Moving Average of Female Mortality from Respiratory Diseases at Ages 45−54

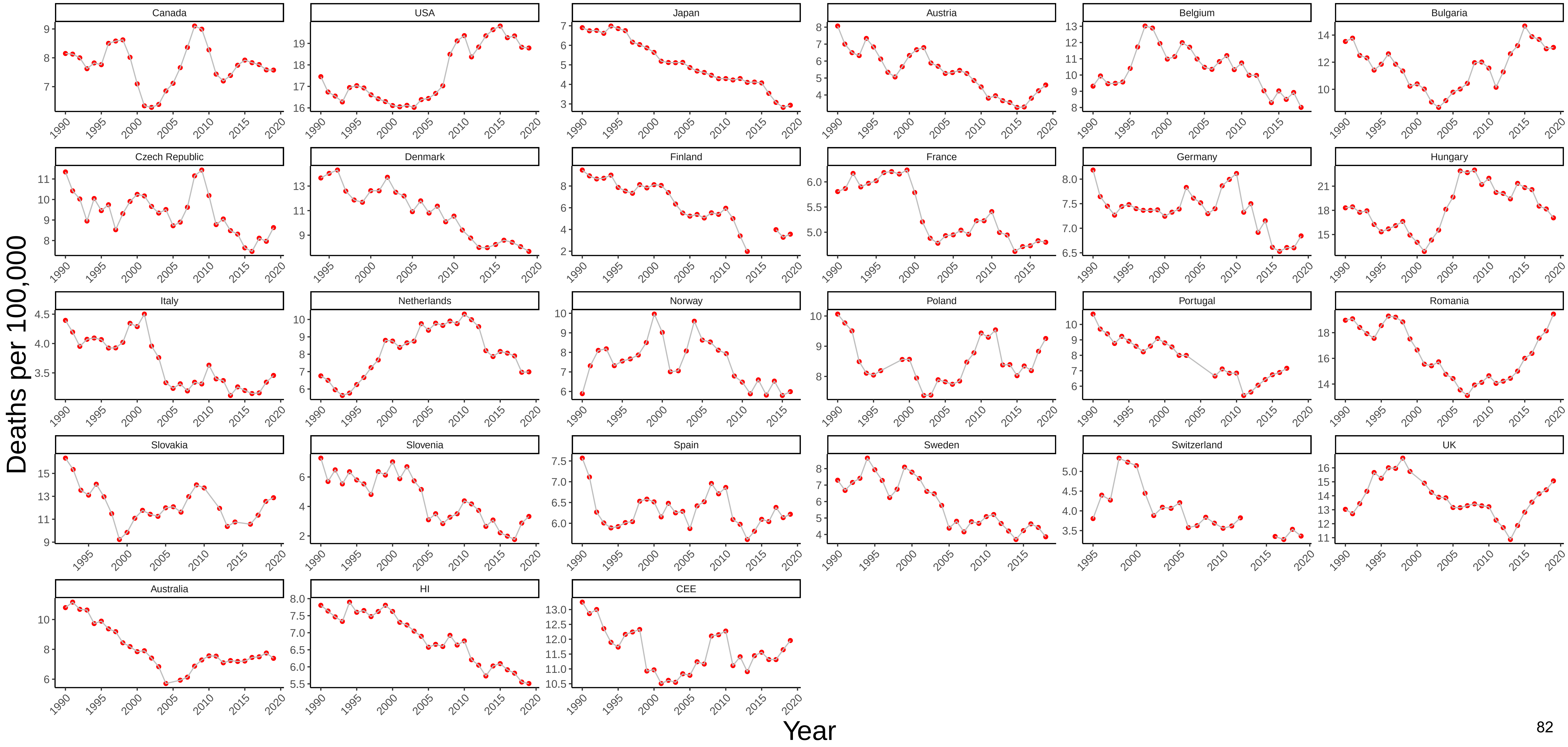

### Figure 35 Three−Year Moving Average of Male Mortality from Trachea/Bronchus, Lung Cancers at Ages 45−54

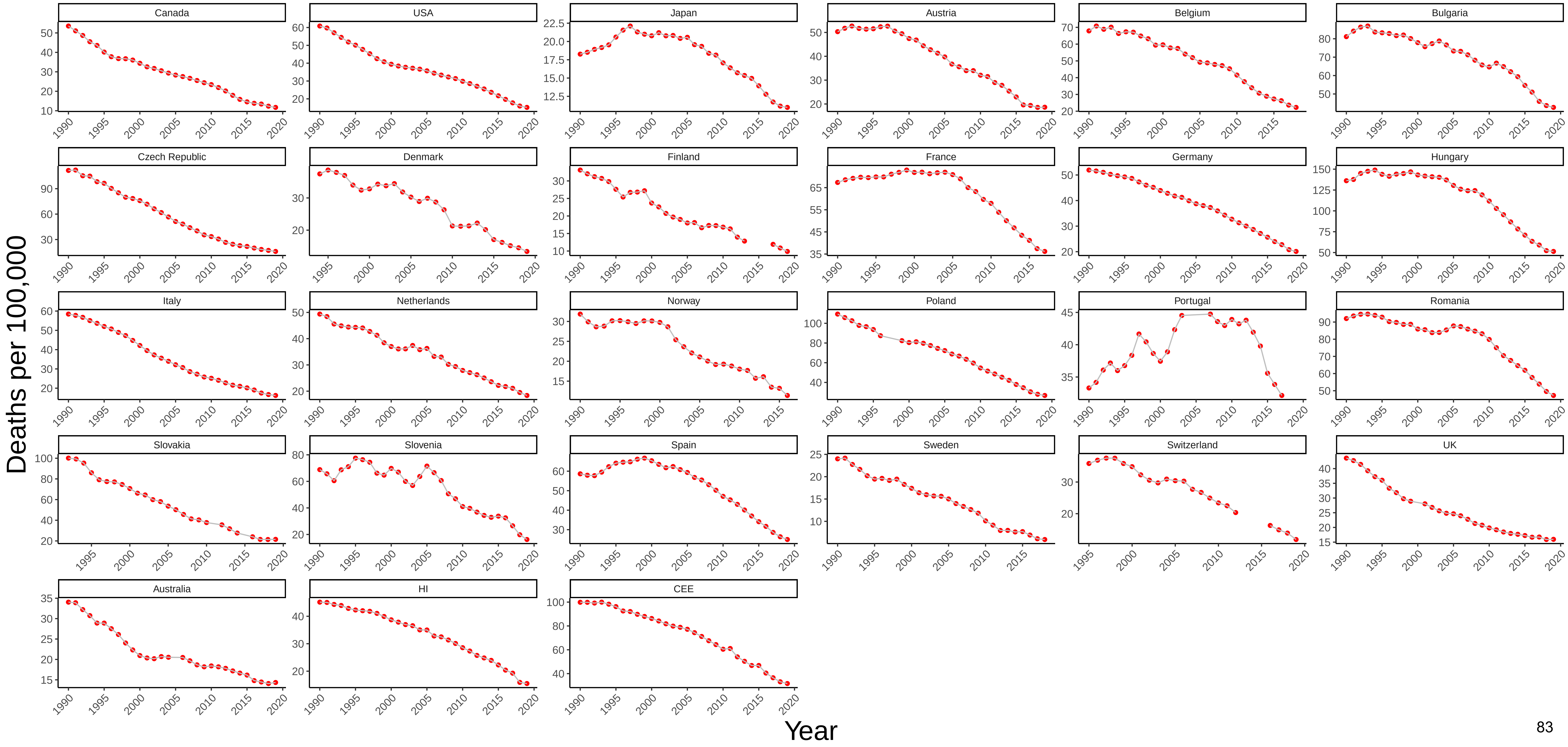

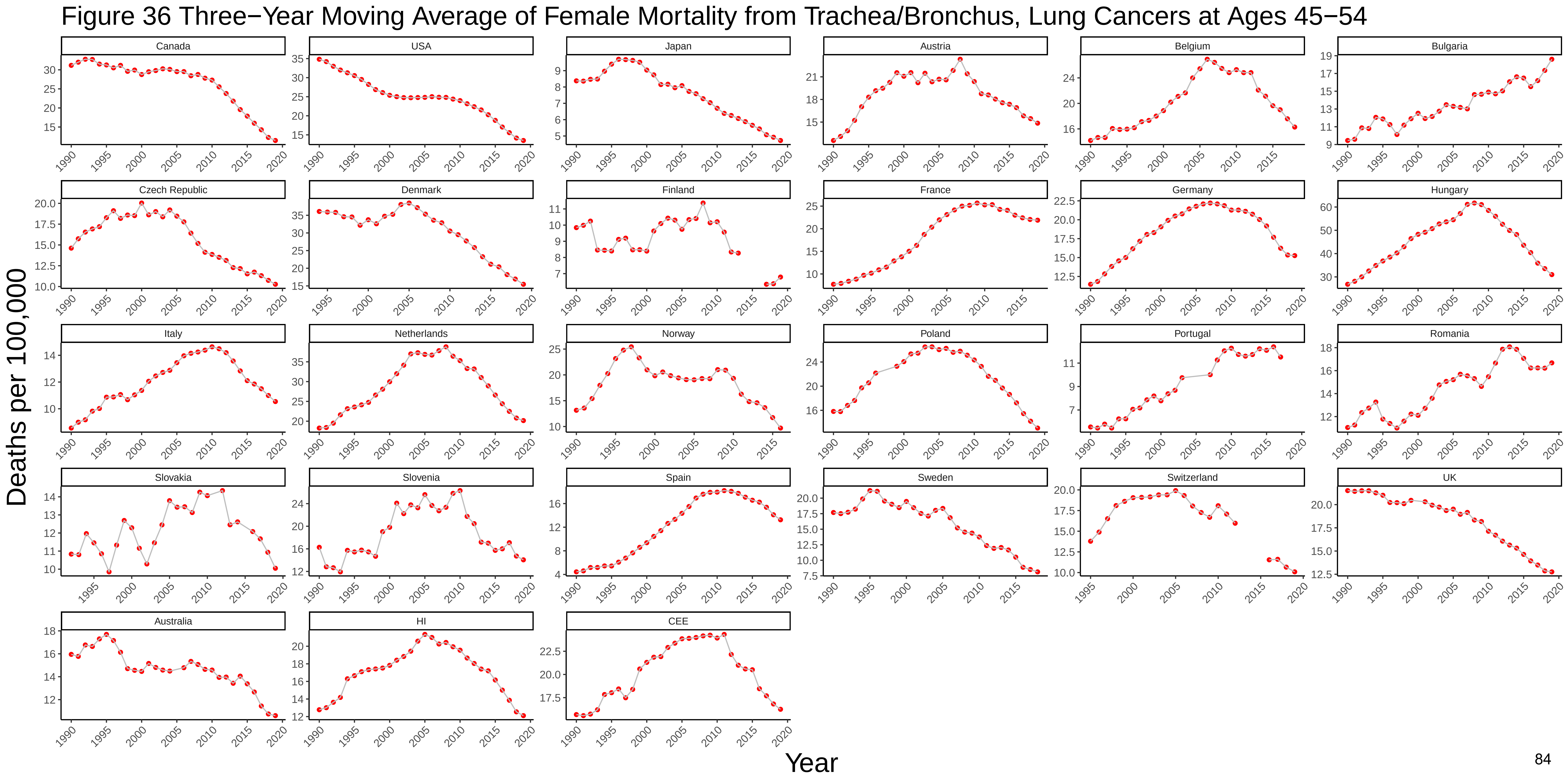

Figure 36 Three−Year Moving Average of Female Mortality from Trachea/Bronchus, Lung Cancers at Ages 45−54
Canada
USA
Japan
Austria
Belgium
Bulgaria
Czech Republic
Denmark
Finland
France
Germany
Hungary
Italy
Netherlands
Norway
Poland
Portugal
Romania
Slovakia
Slovenia
Spain
Sweden
Switzerland
UK
Australia
HI
CEE
Deaths per 100,000
Year

### Figure 37 Three−Year Moving Average of Male Mortality from All Other Cancers at Ages 45−54

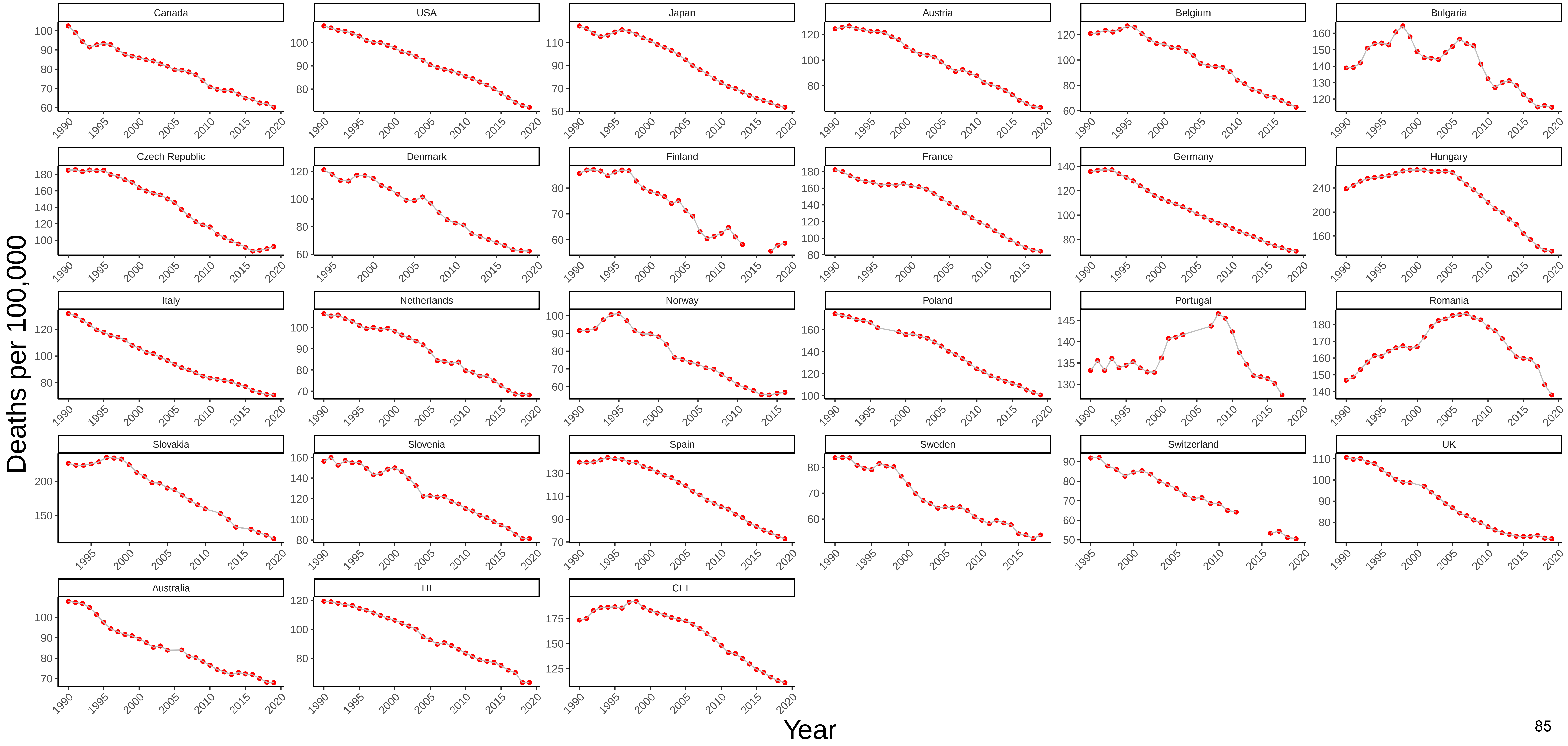

### Figure 38 Three−Year Moving Average of Female Mortality from All Other Cancers at Ages 45−54

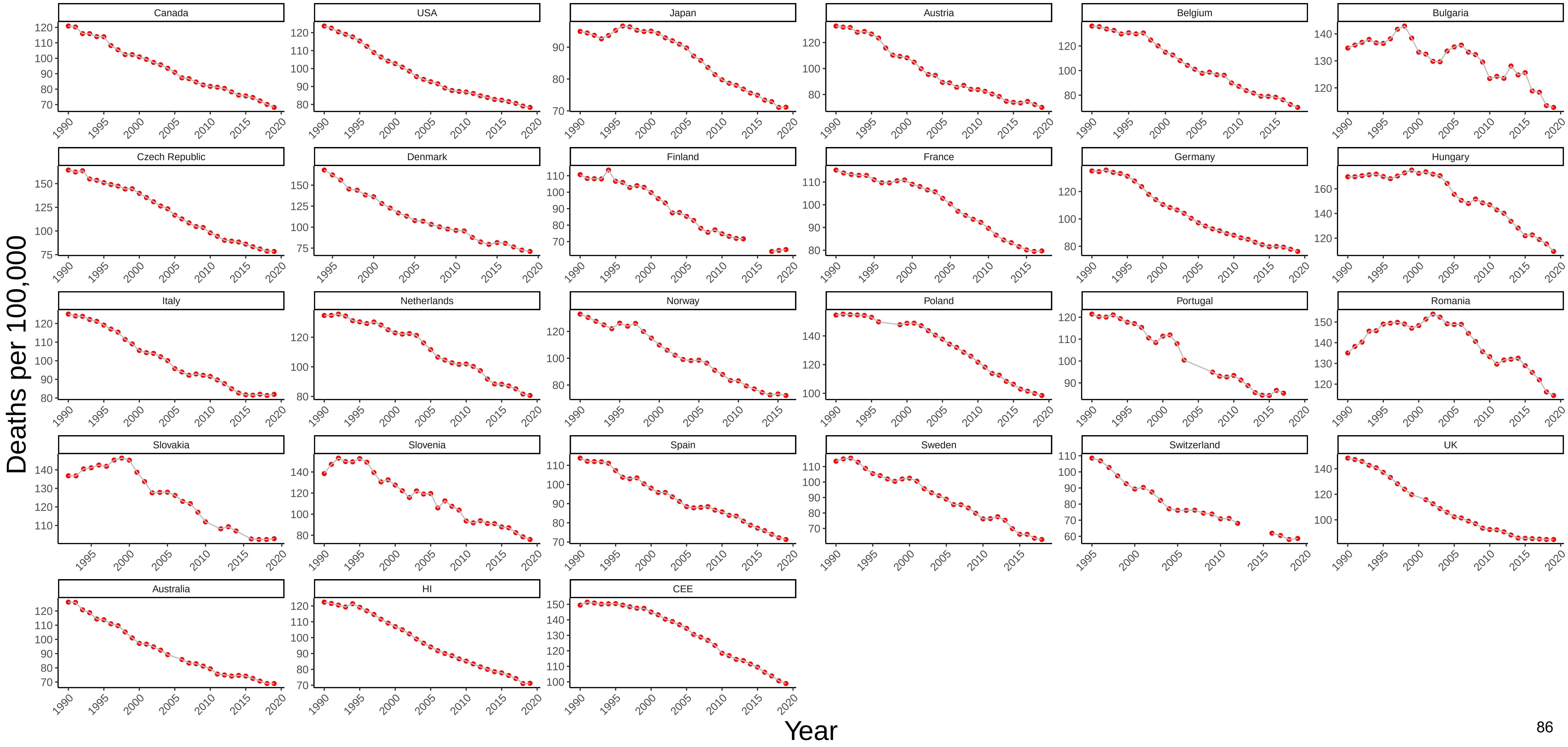

### Figure 39 Three−Year Moving Average of Male Mortality from Nervous System Diseases at Ages 45−54

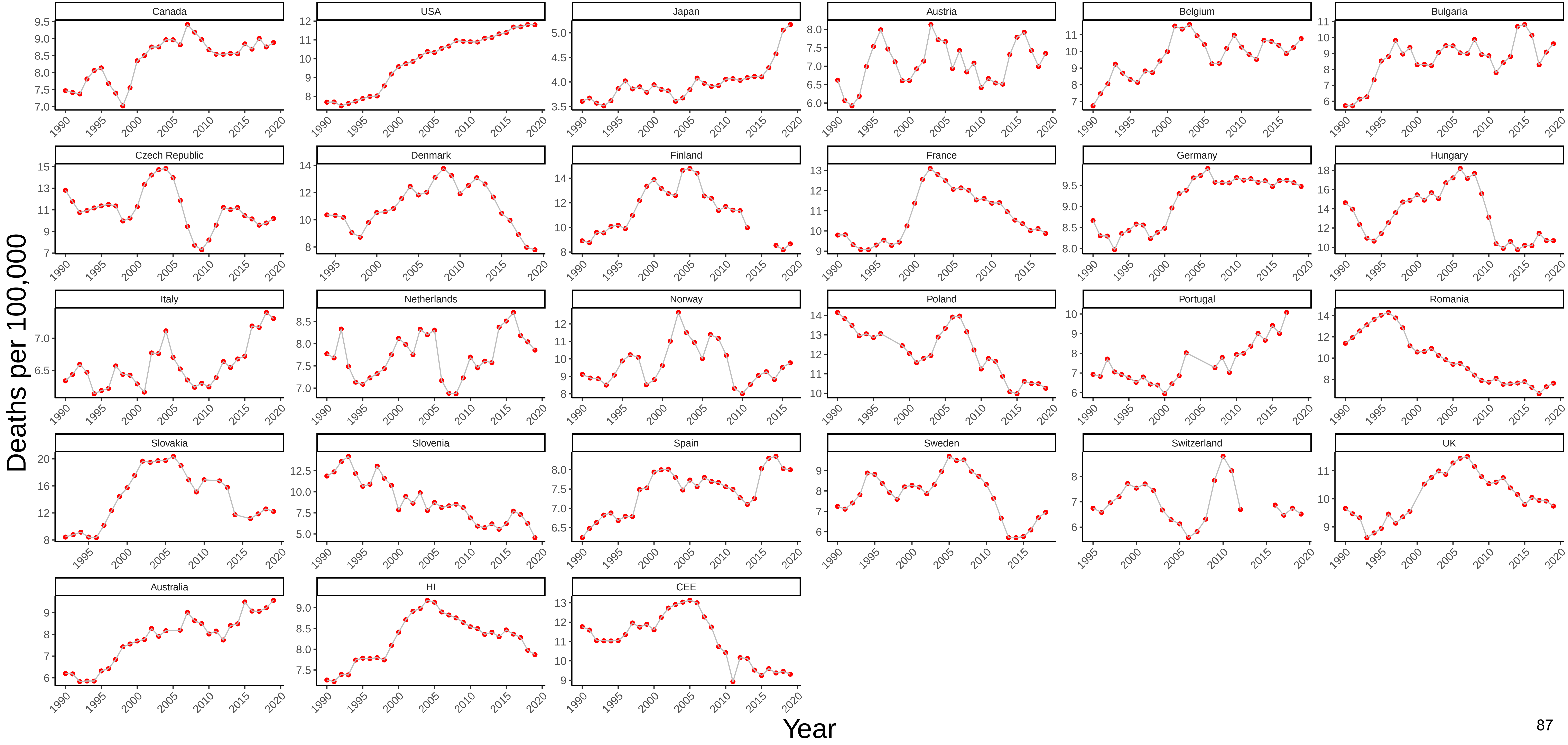

### Figure 40 Three−Year Moving Average of Female Mortality from Nervous System Diseases at Ages 45−54

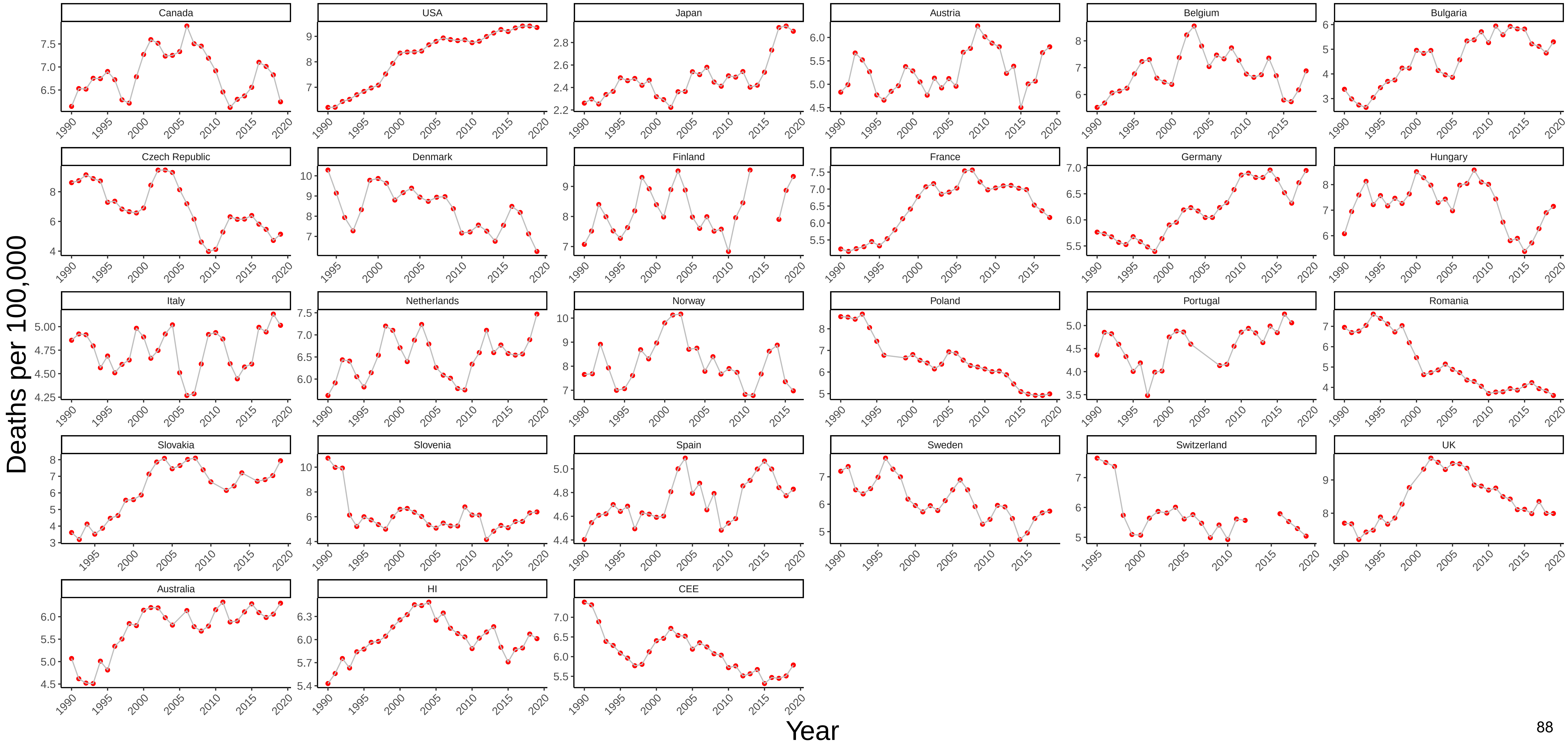

Figure 41 Three−Year Moving Average of Male Mortality from Metabolic Diseases at Ages 45−54

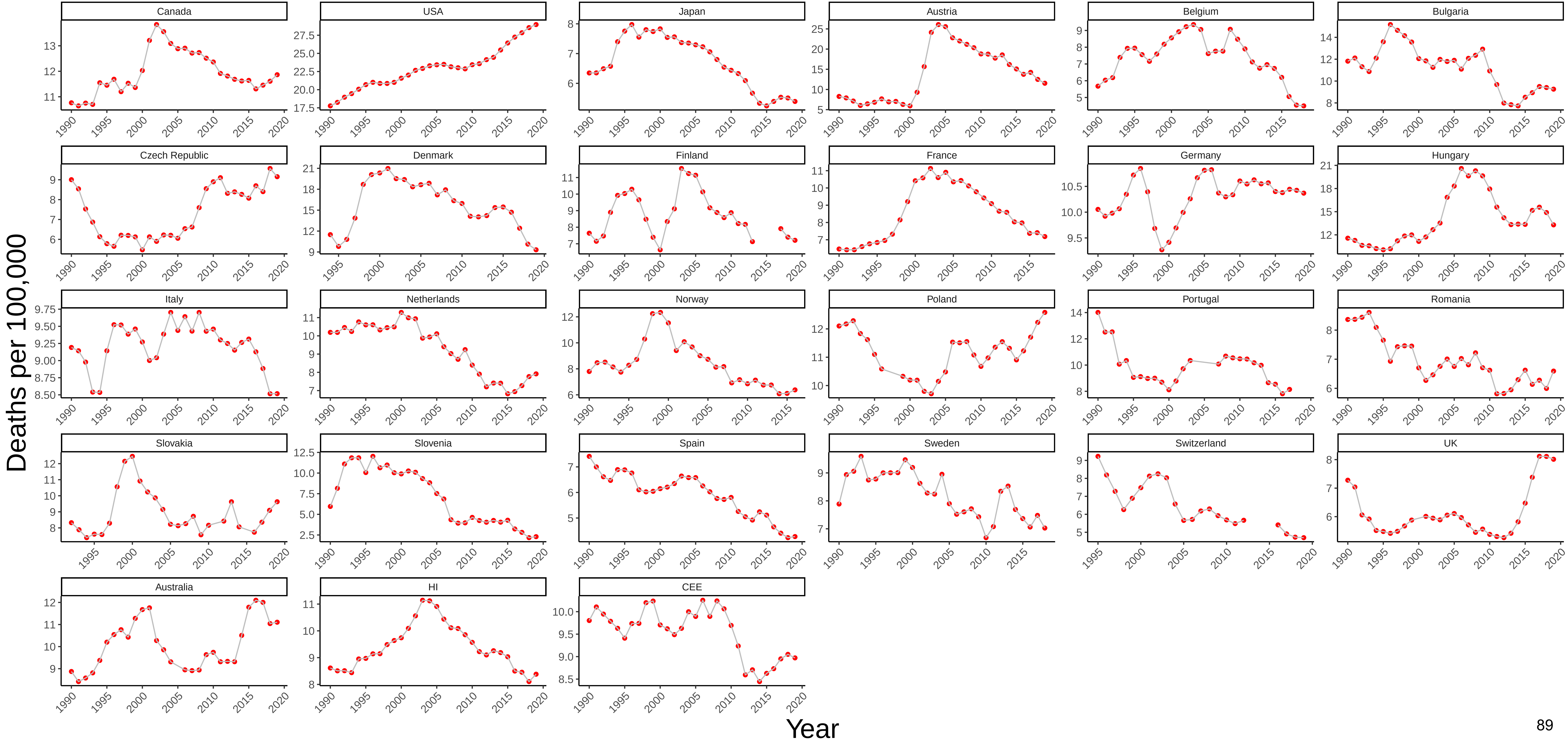

### Figure 42 Three−Year Moving Average of Female Mortality from Metabolic Diseases at Ages 45−54

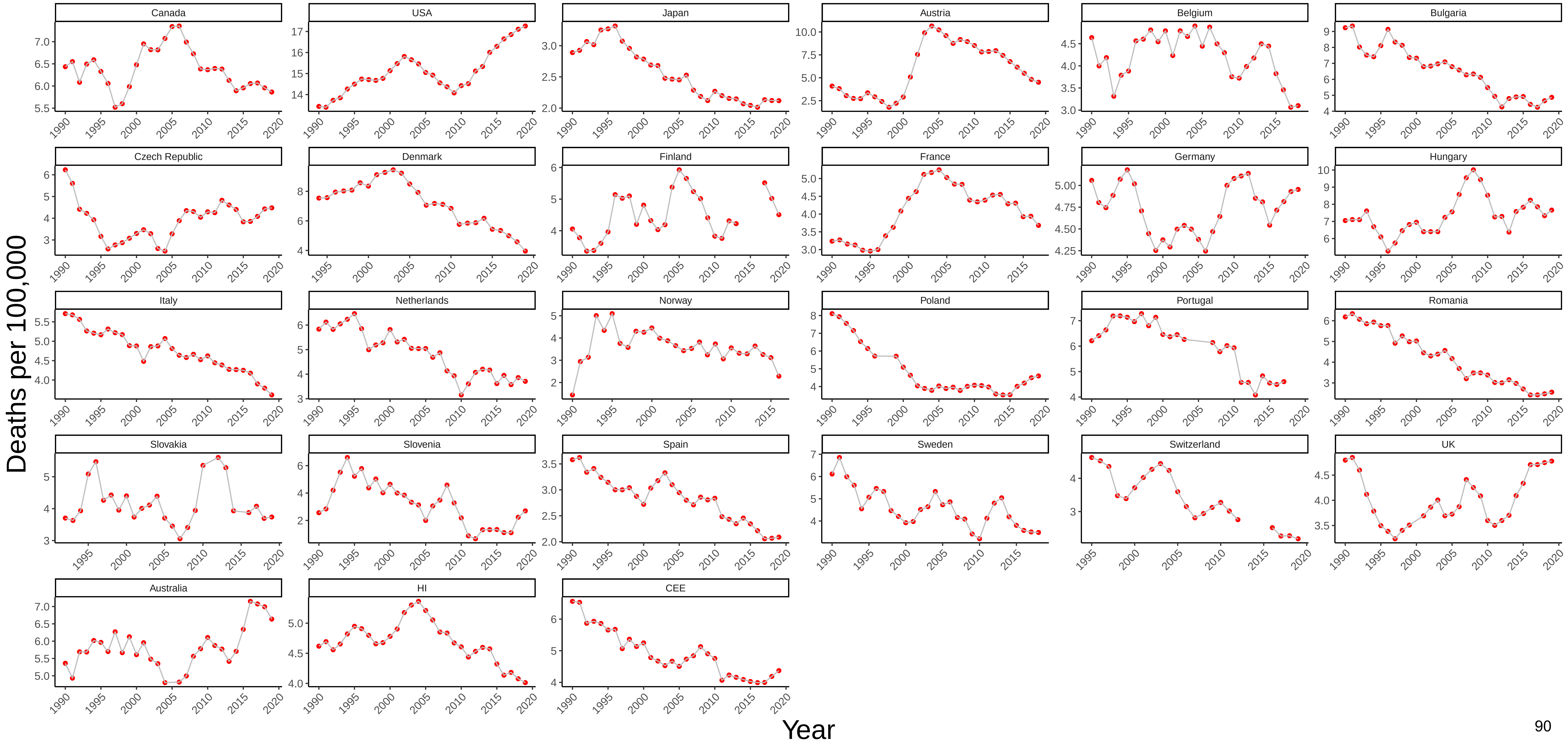

### Figure 43 Three−Year Moving Average of Male Mortality from Cardiovascular Disease at Ages 45−54

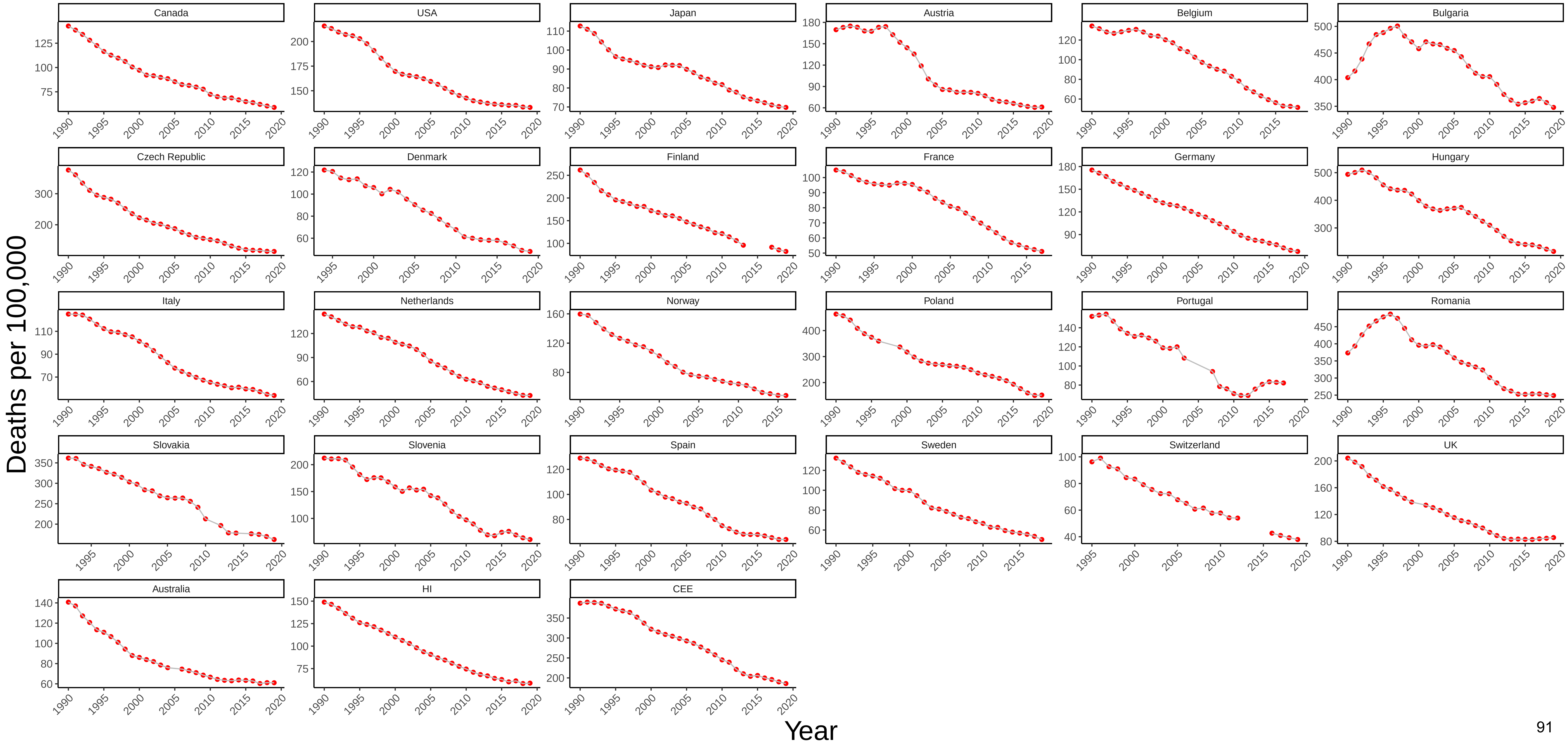

Figure 44 Three−Year Moving Average of Female Mortality from Cardiovascular Disease at Ages 45−54

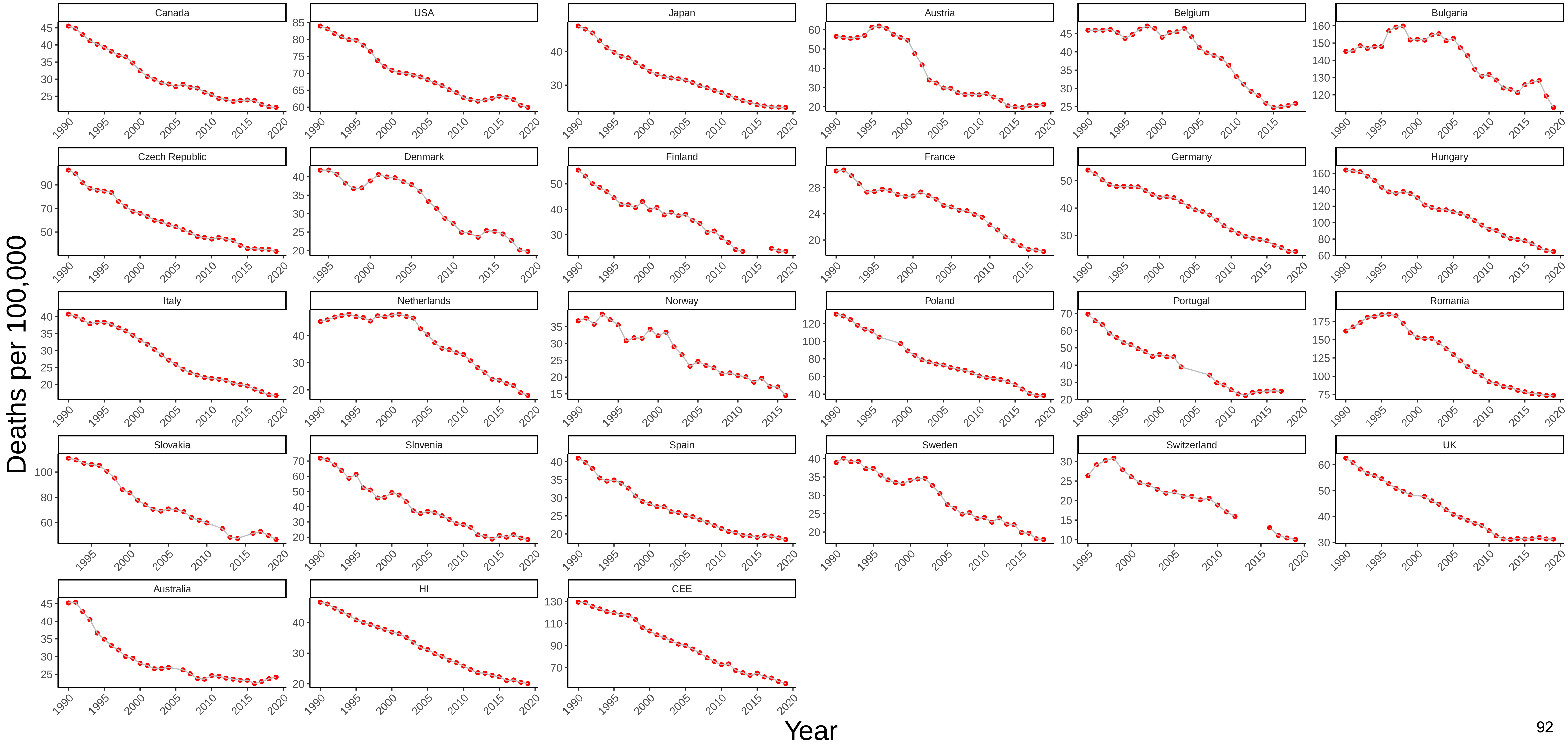

### Figure 45 Three−Year Moving Average of Male Mortality from Suicide at Ages 45−54

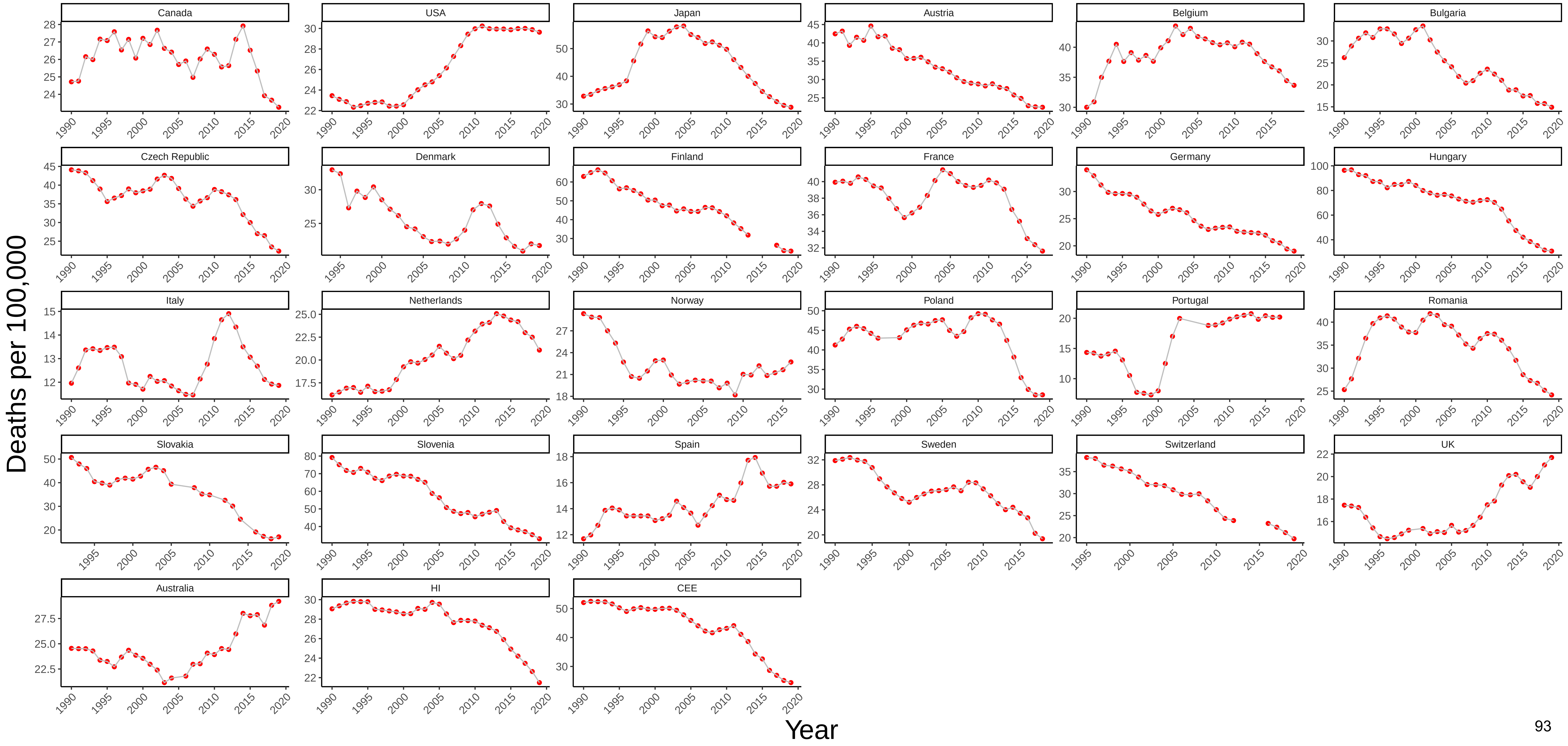

### Figure 46 Three−Year Moving Average of Female Mortality from Suicide at Ages 45−54

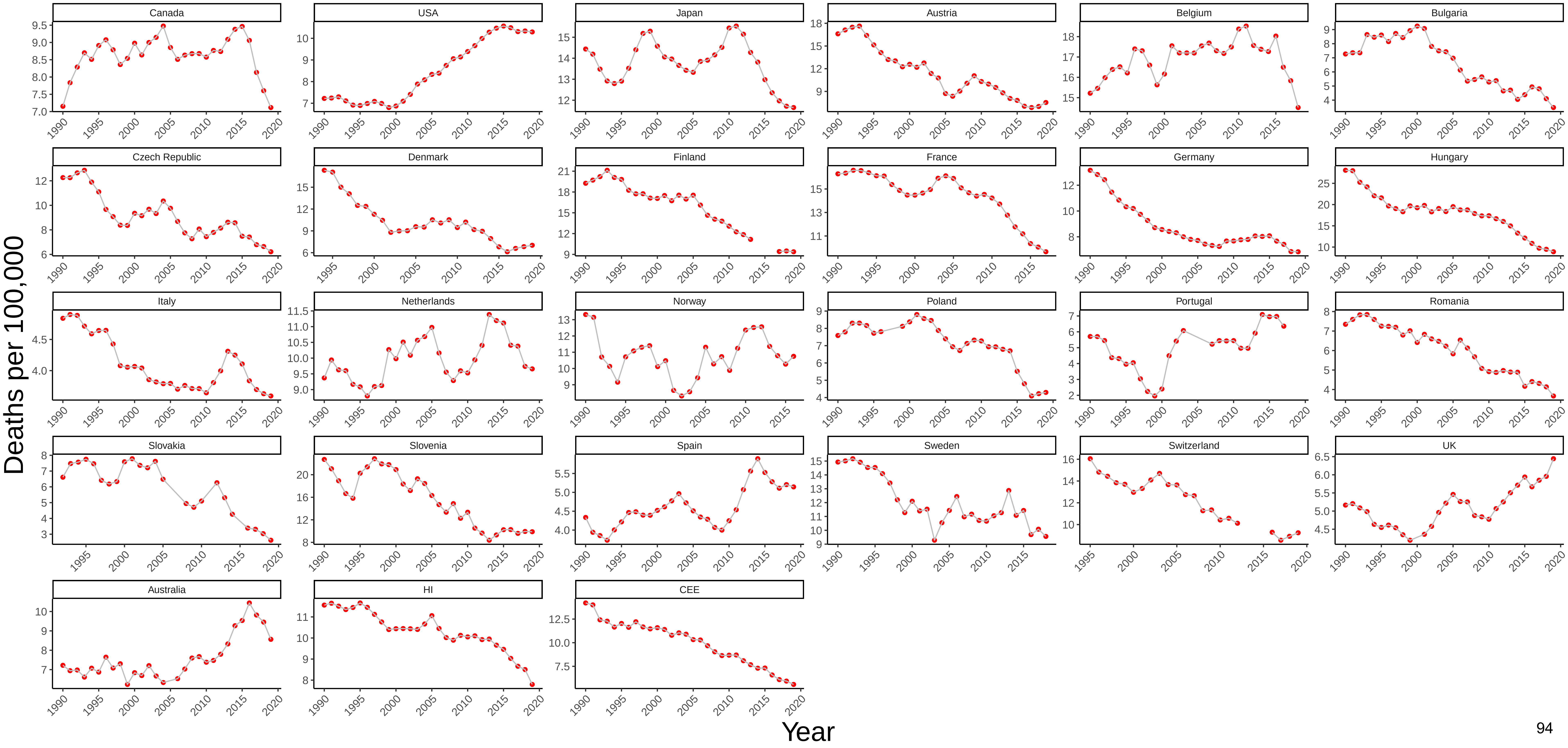

Figure 47 Three−Year Moving Average of Male Mortality from Homicide at Ages 45−54

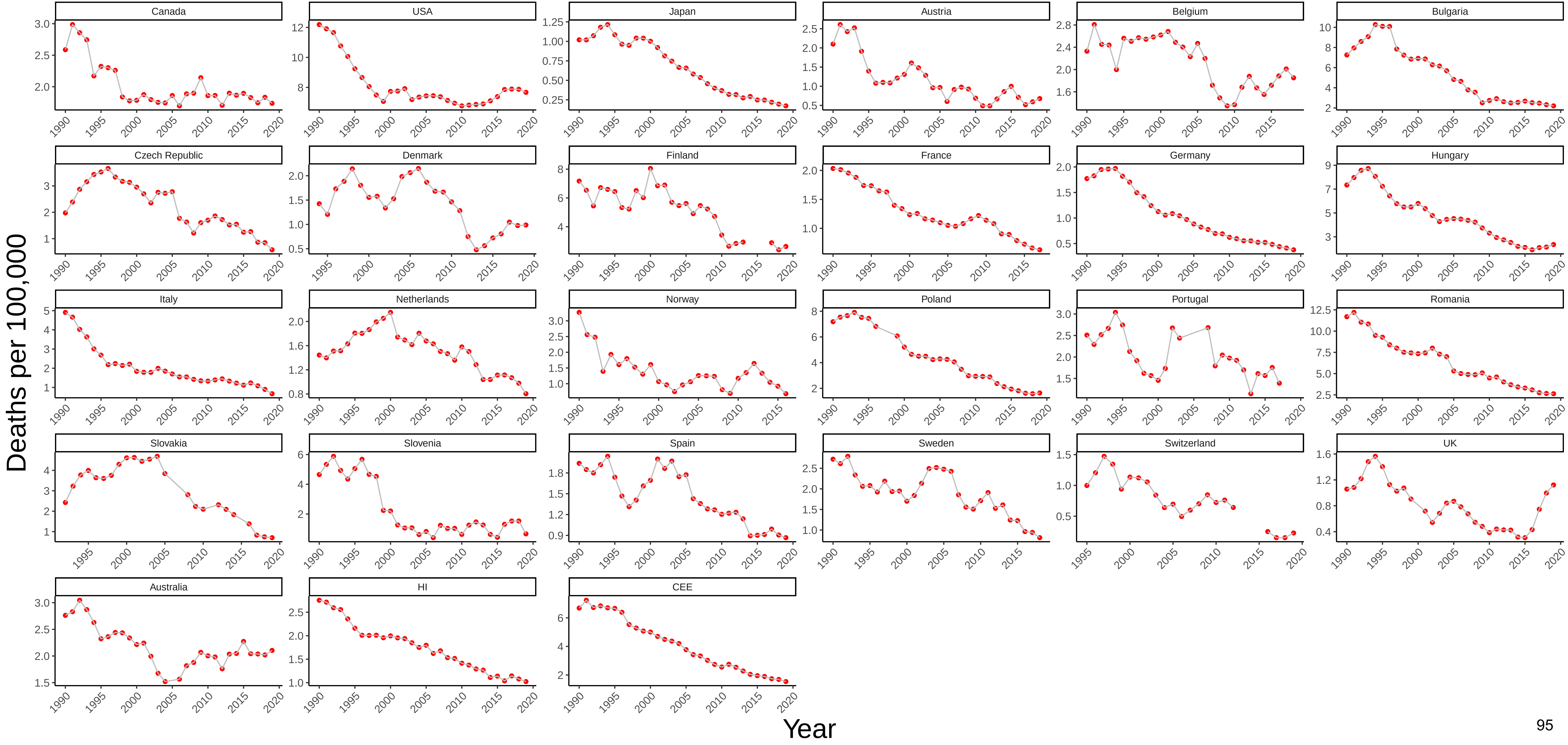

### Figure 48 Three−Year Moving Average of Female Mortality from Homicide at Ages 45−54

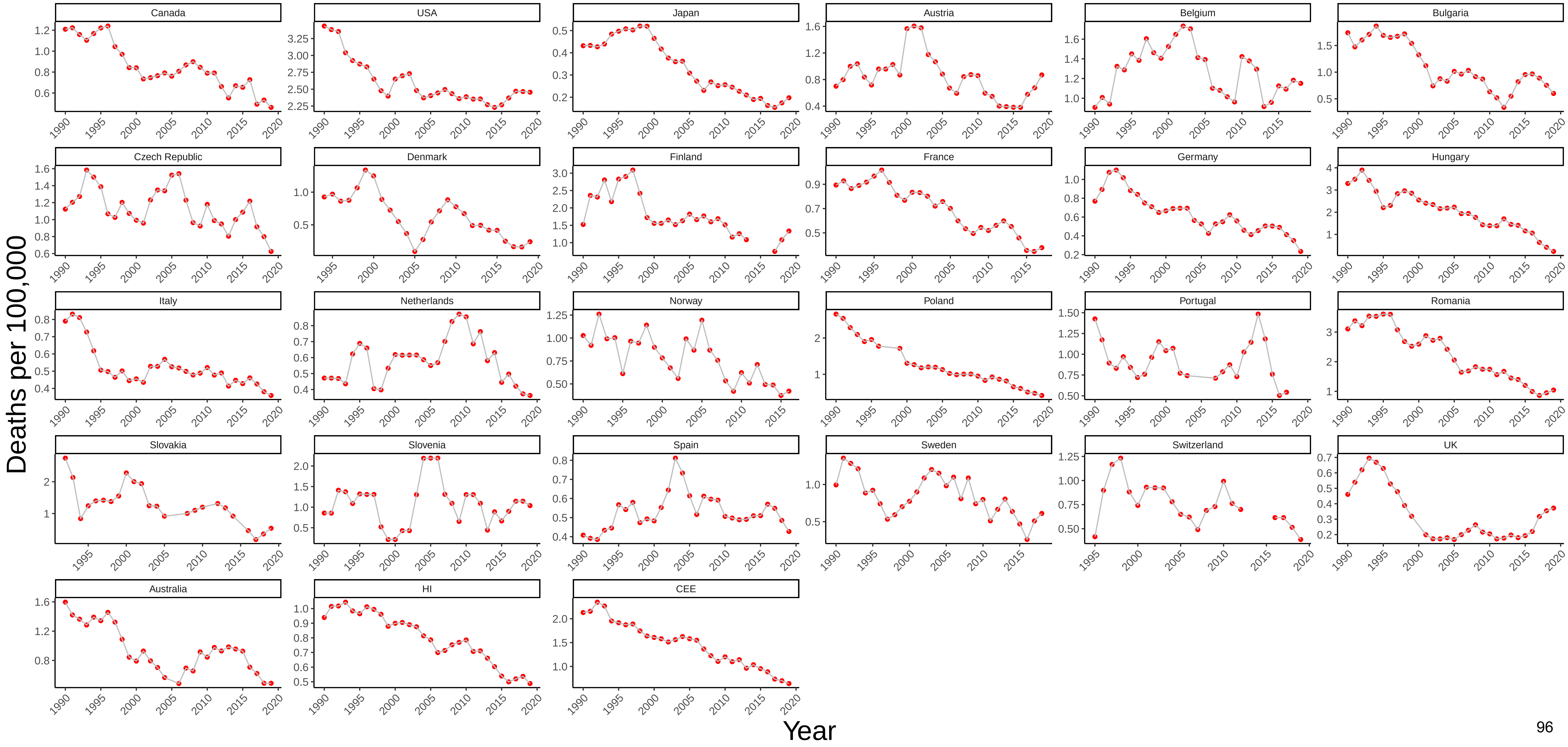

### Figure 49 Three−Year Moving Average of Male Mortality from Transport Accidents at Ages 45−54

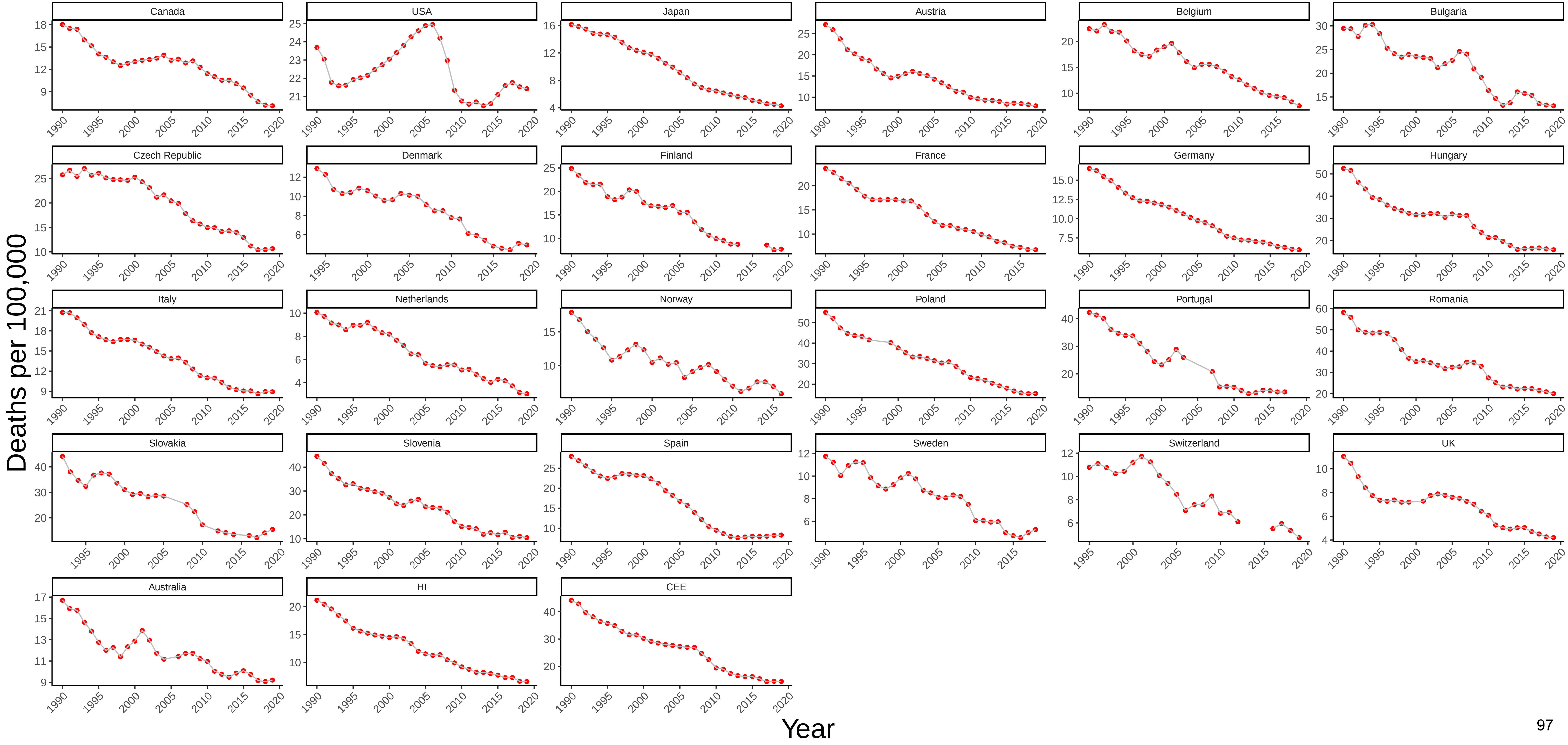

Figure 50 Three−Year Moving Average of Female Mortality from Transport Accidents at Ages 45−54

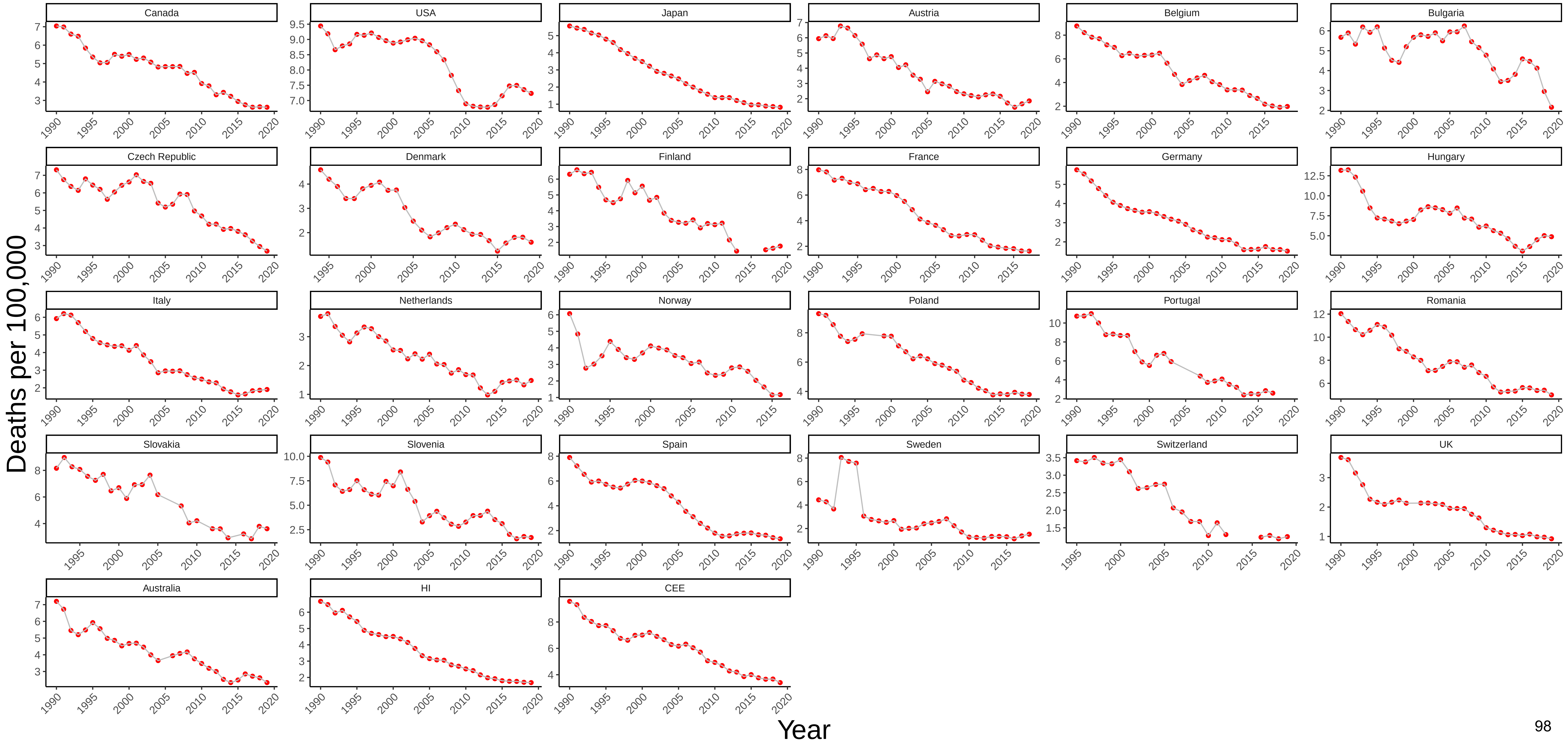

Figure 51 Three−Year Moving Average of Male Mortality from Other External Causes at Ages 45−54

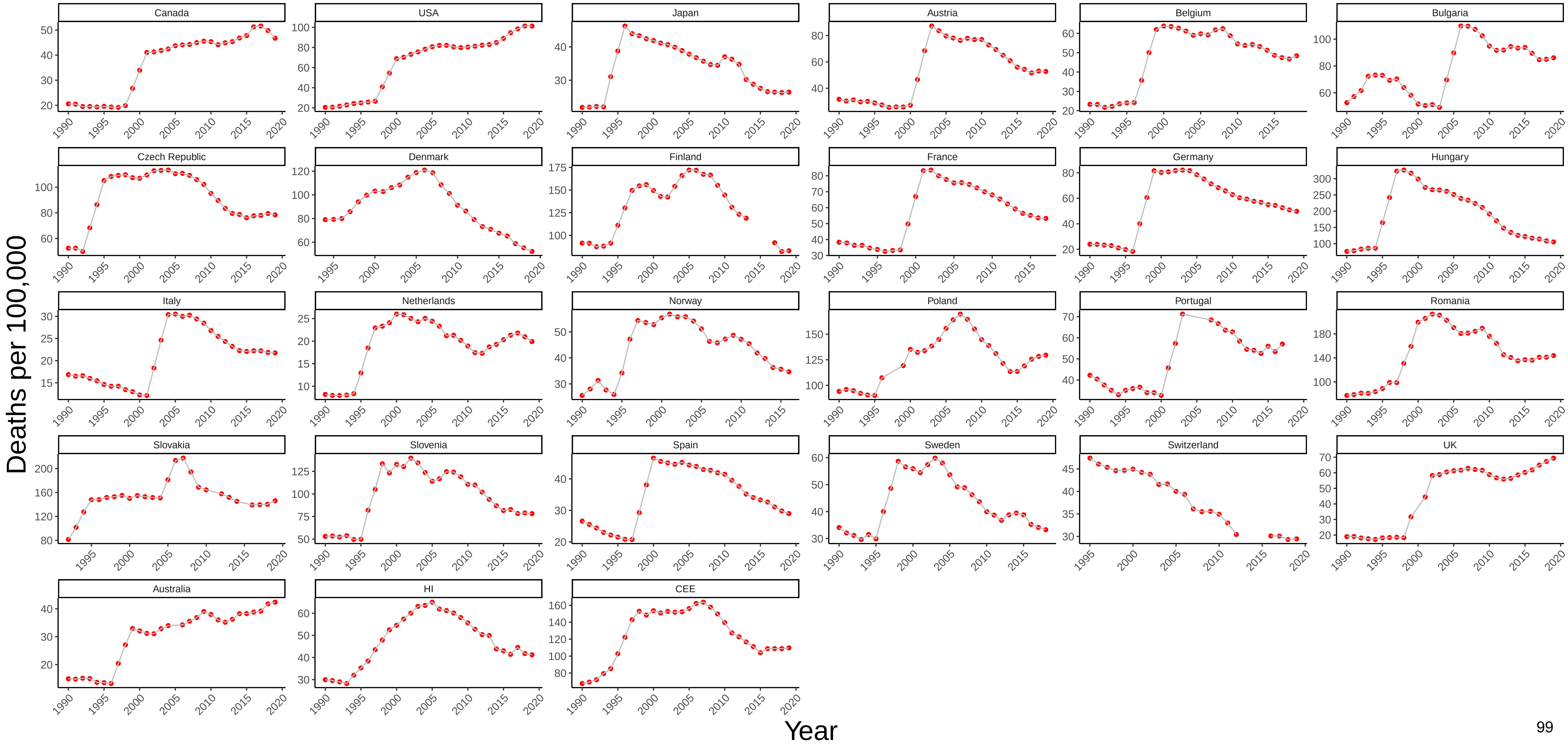

### Figure 52 Three−Year Moving Average of Female Mortality from Other External Causes at Ages 45−54

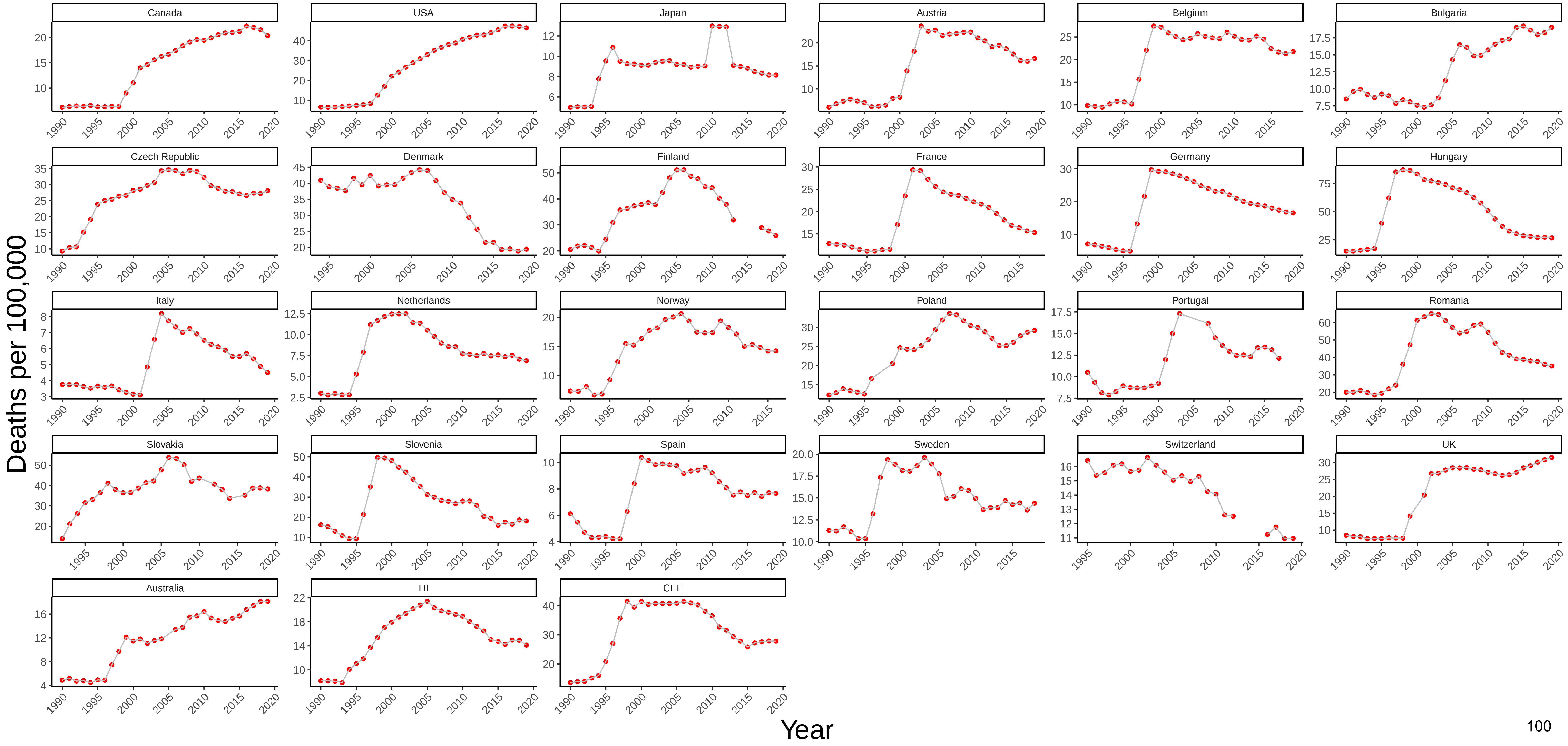

### Figure 53 Three−Year Moving Average of Male Mortality from All Other Causes at Ages 45−54

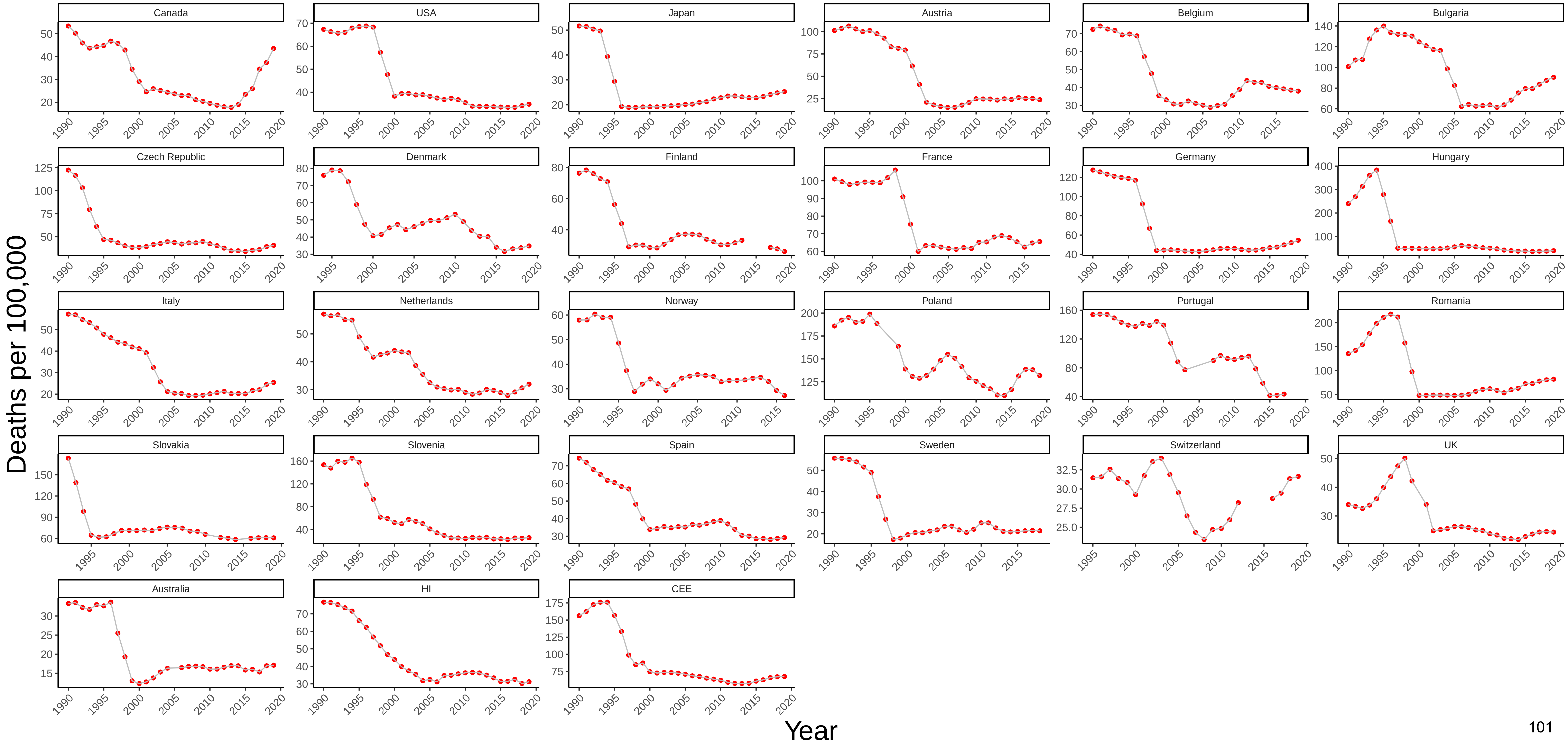

Figure 54 Three−Year Moving Average of Female Mortality from All Other Causes at Ages 45−54

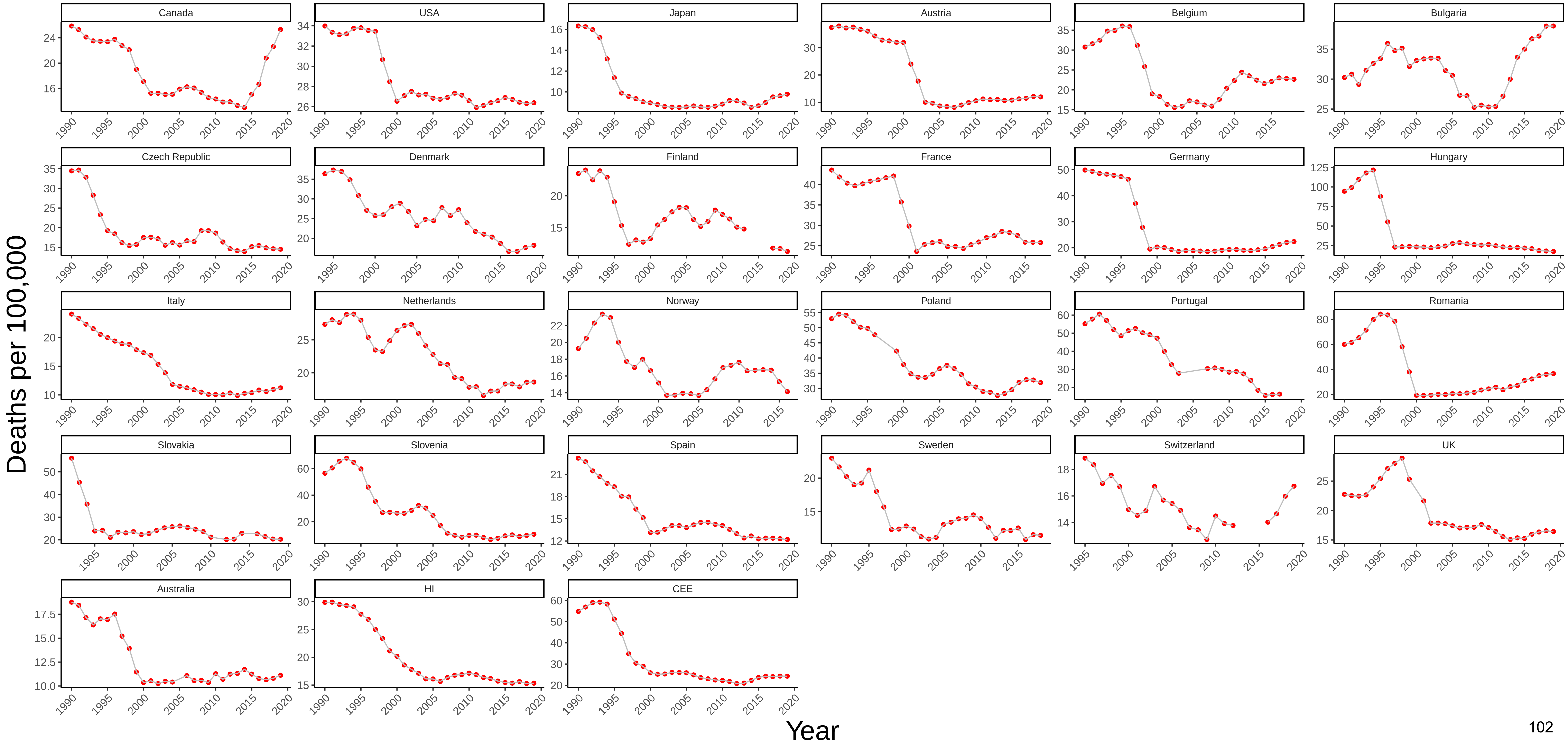

Figure 55 Three−Year Moving Average of Male Mortality from All Causes at Ages 45−54

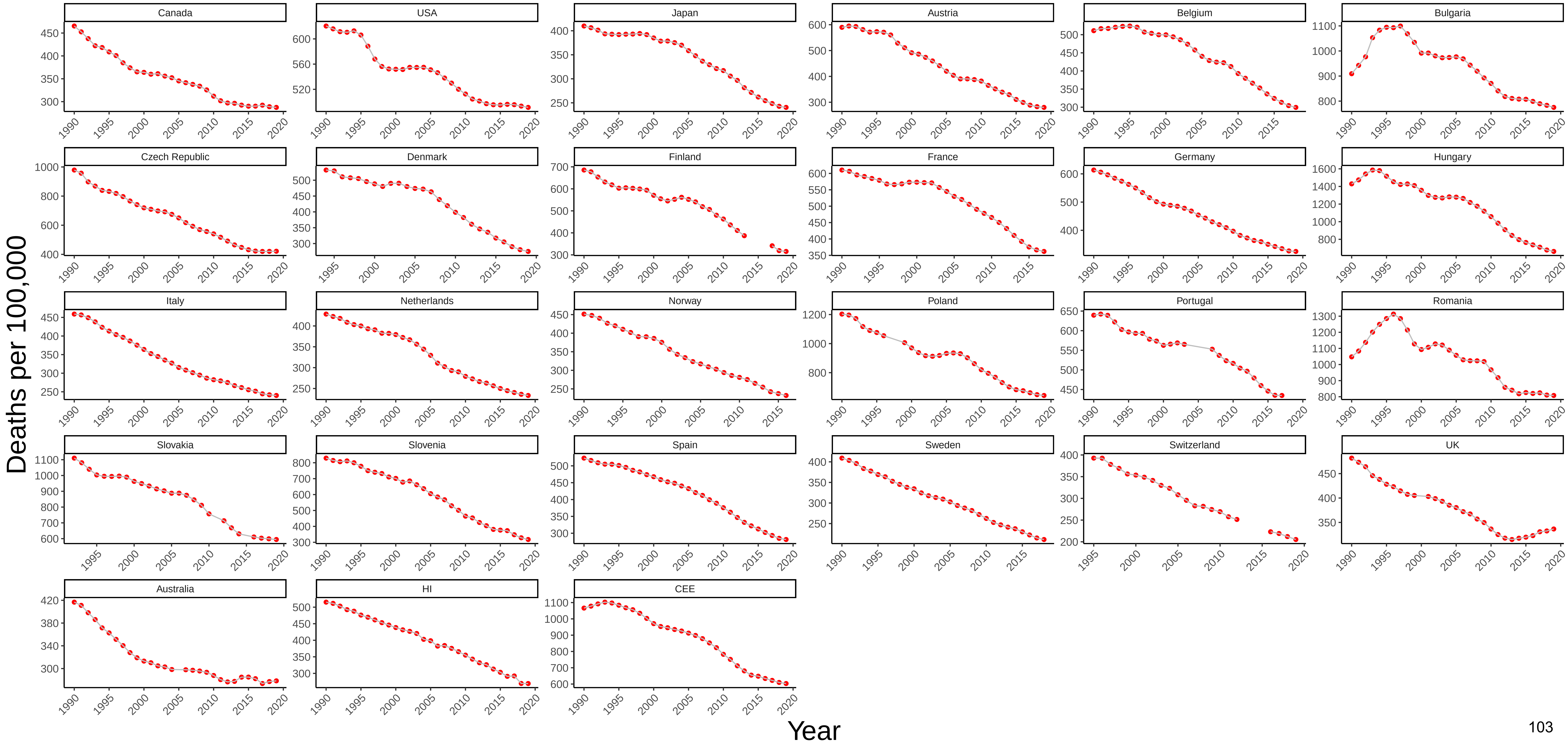

### Figure 56 Three−Year Moving Average of Female Mortality from All Causes at Ages 45−54

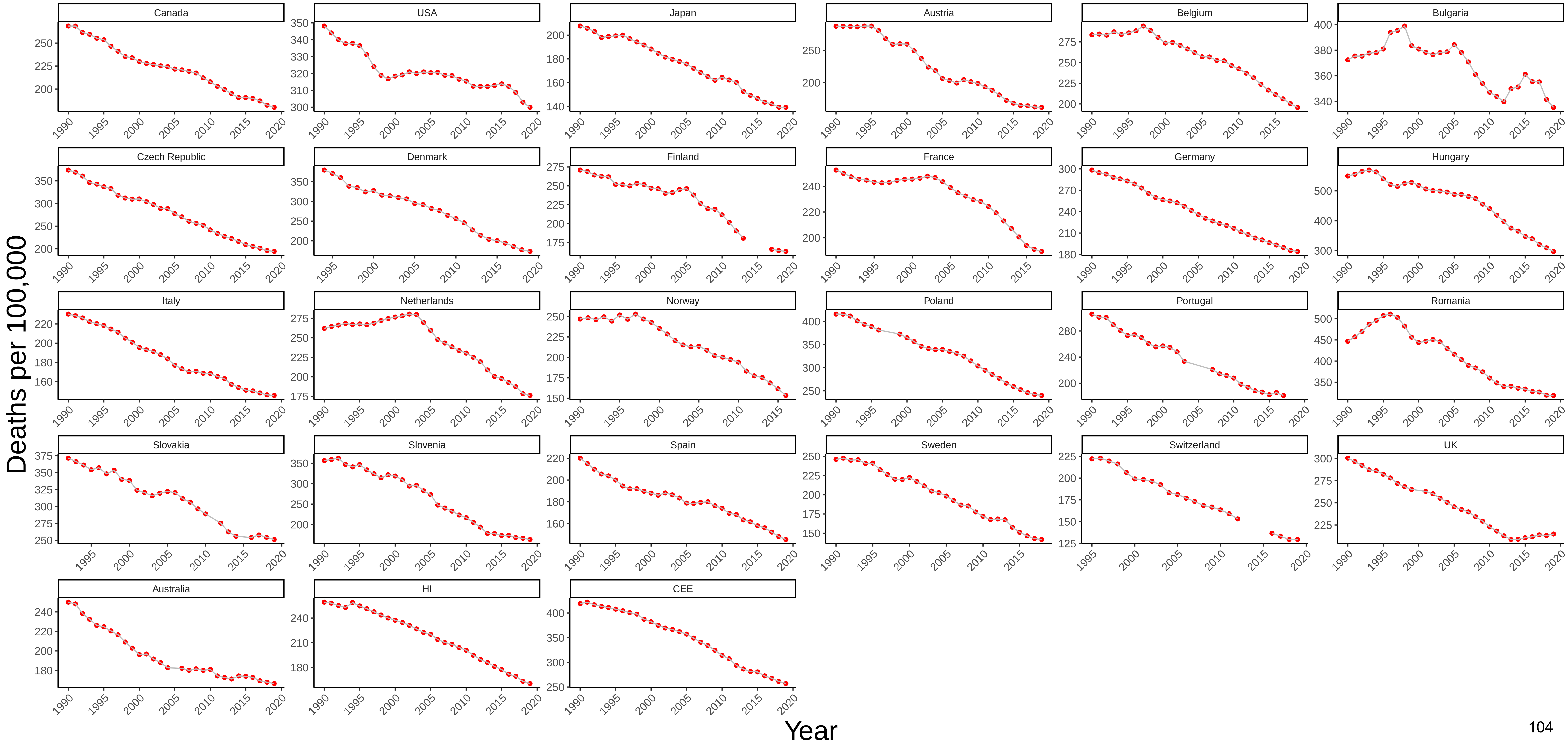

### Figure 57 Three−Year Moving Average of Male Mortality from Infectious and Parasitic Diseases at Ages 55−64

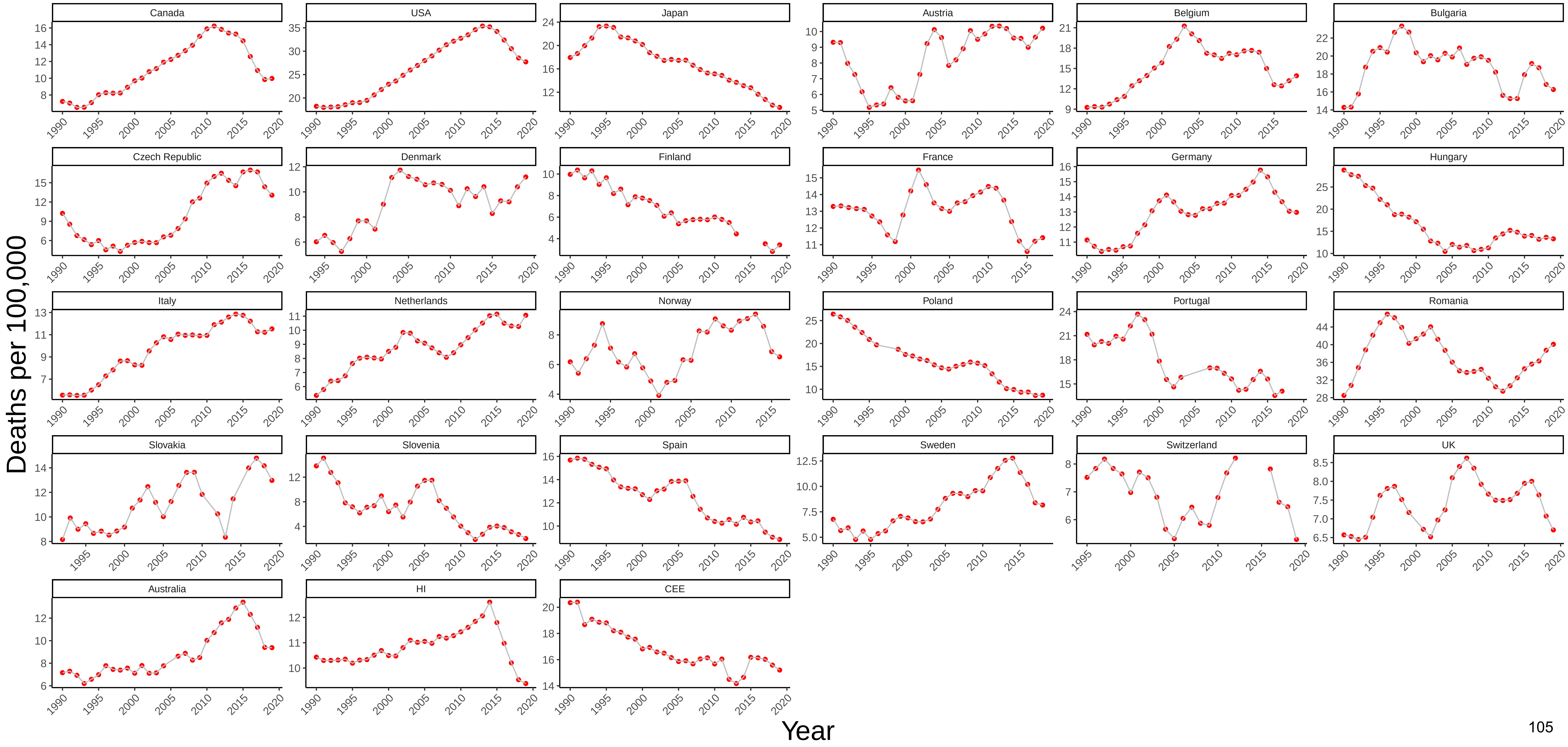

Figure 58 Three−Year Moving Average of Female Mortality from Infectious and Parasitic Diseases at Ages 55−64

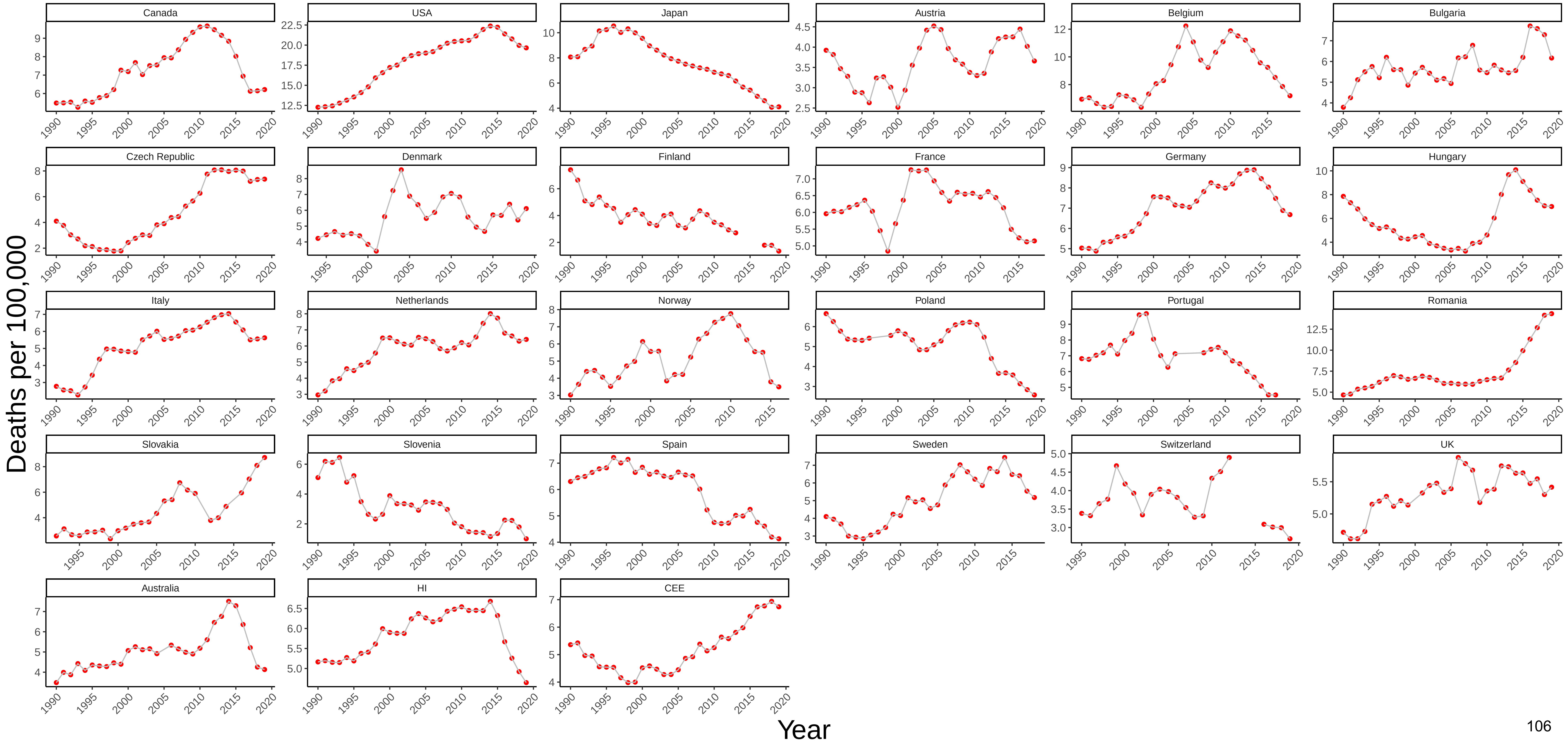

### Figure 59 Three−Year Moving Average of Male Mortality from HIV/AIDS at Ages 55−64

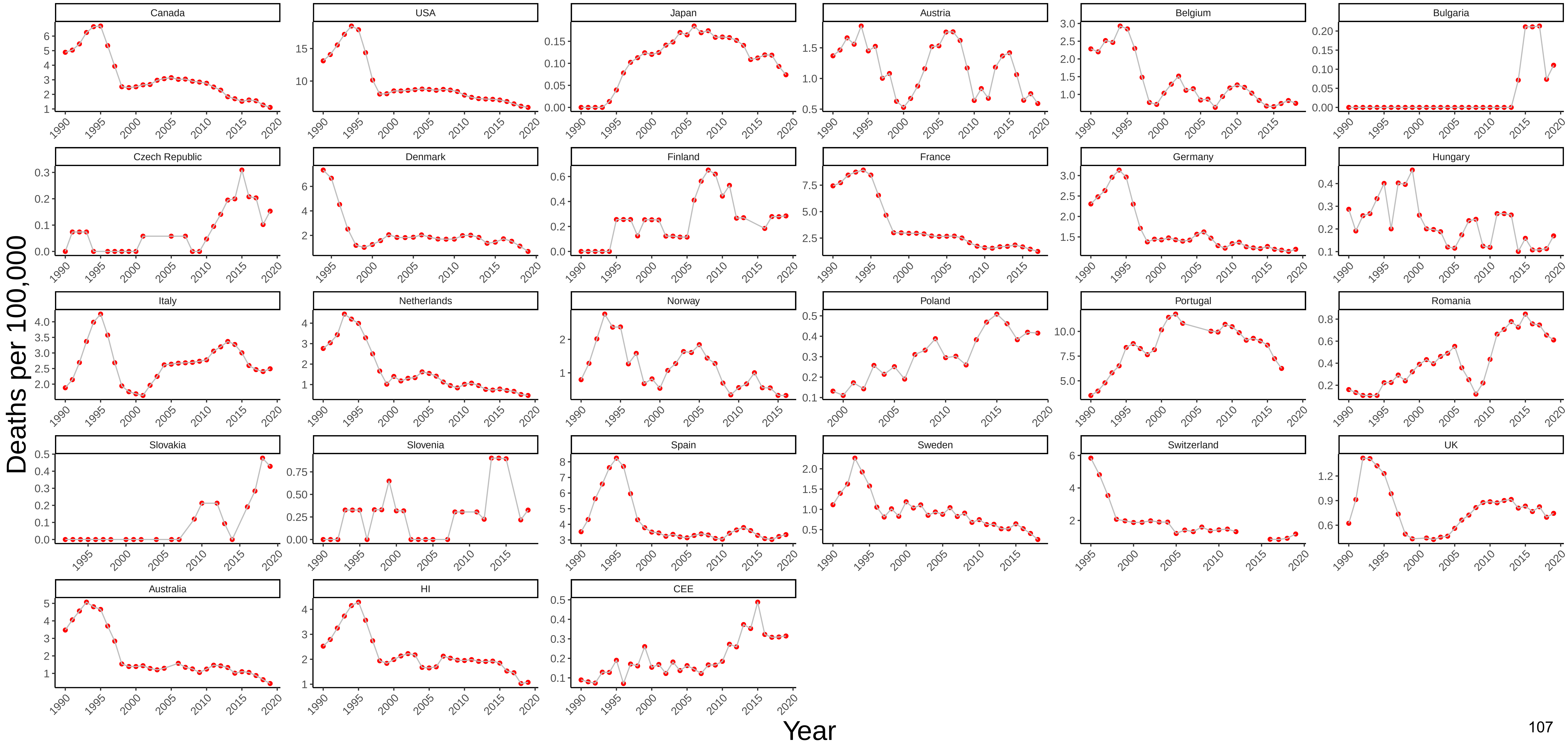

### Figure 60 Three−Year Moving Average of Female Mortality from HIV/AIDS at Ages 55−64

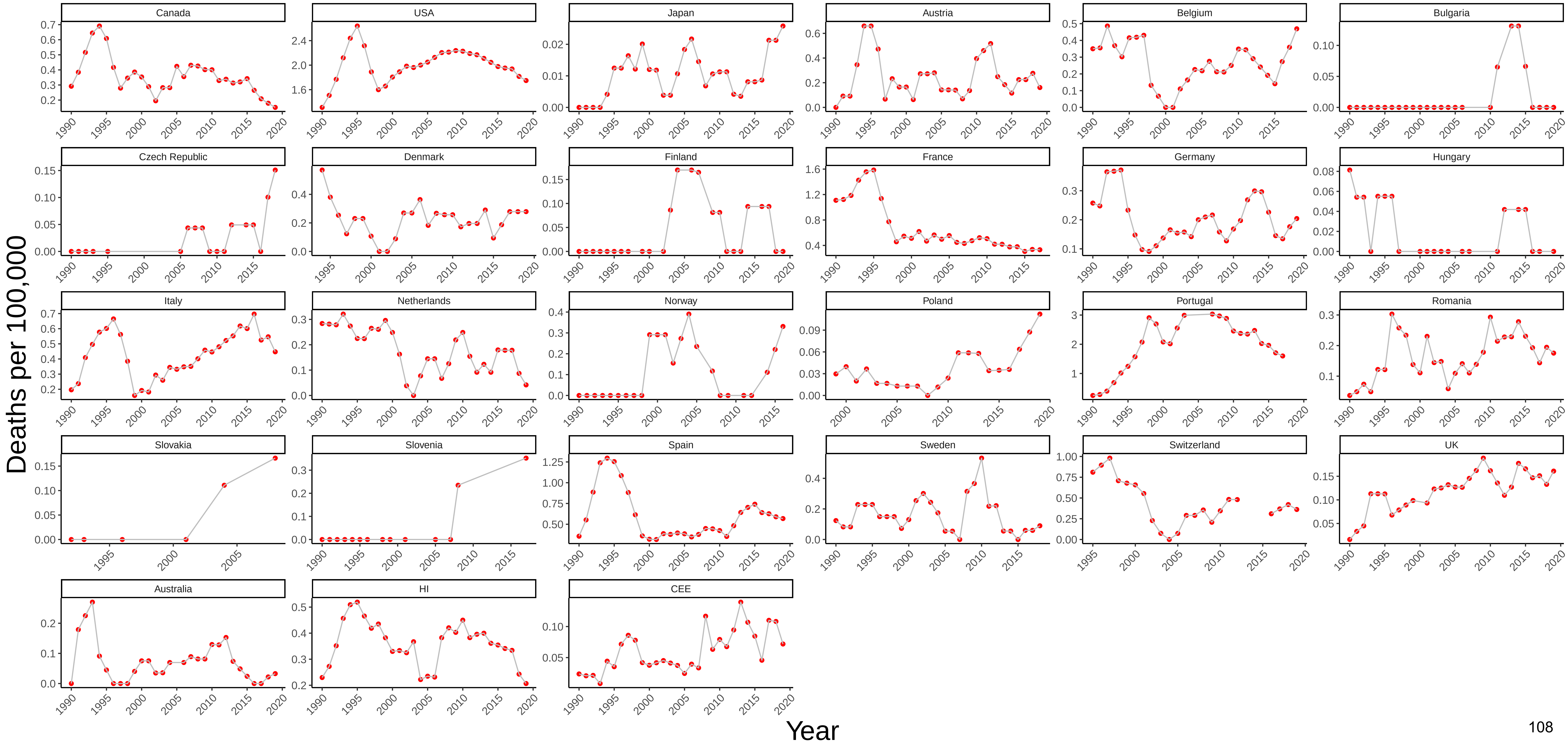

### Figure 61 Three−Year Moving Average of Male Mortality from Respiratory Diseases at Ages 55−64

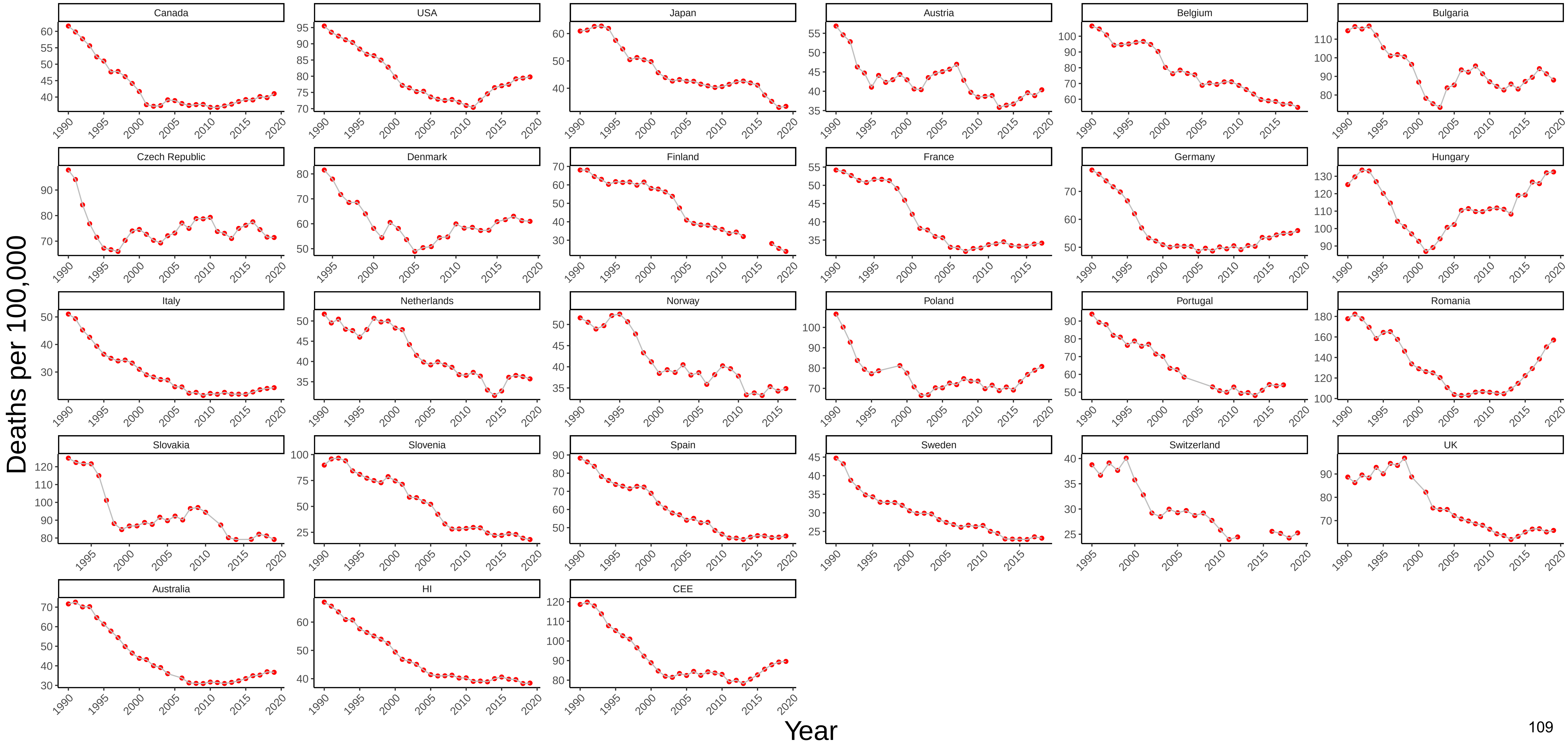

### Figure 62 Three−Year Moving Average of Female Mortality from Respiratory Diseases at Ages 55−64

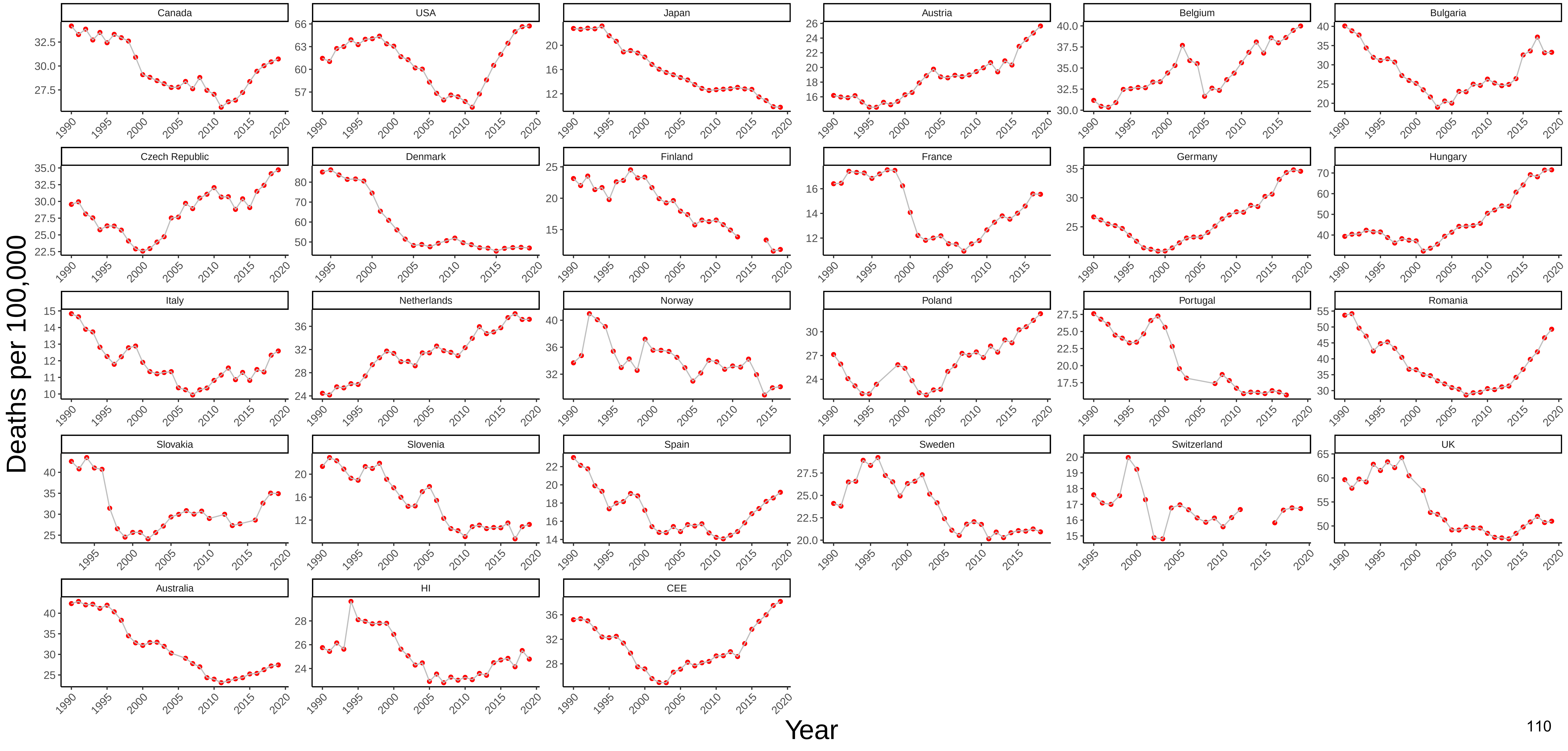

Figure 63 Three−Year Moving Average of Male Mortality from Trachea/Bronchus, Lung Cancers at Ages 55−64

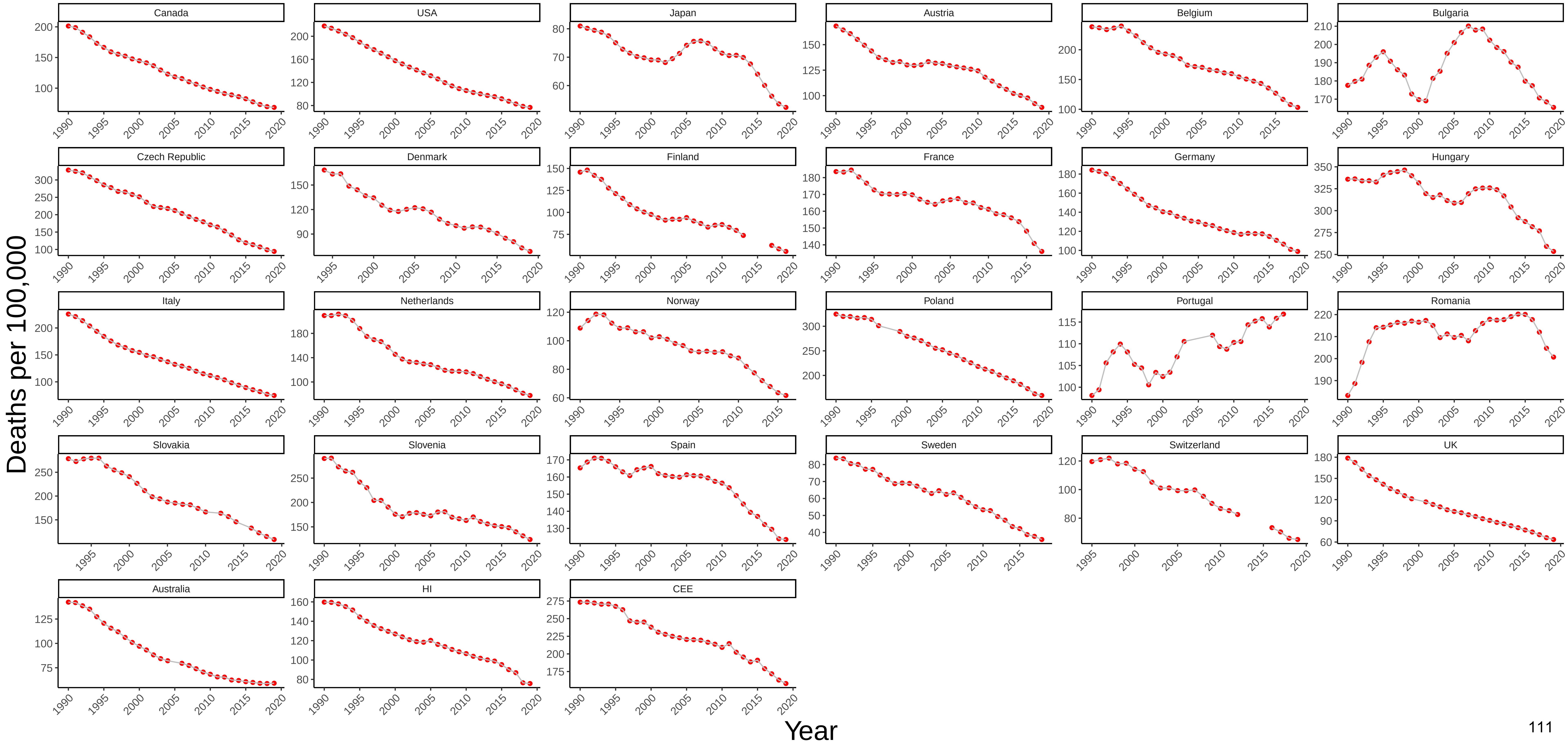

Figure 64 Three−Year Moving Average of Female Mortality from Trachea/Bronchus, Lung Cancers at Ages 55−64

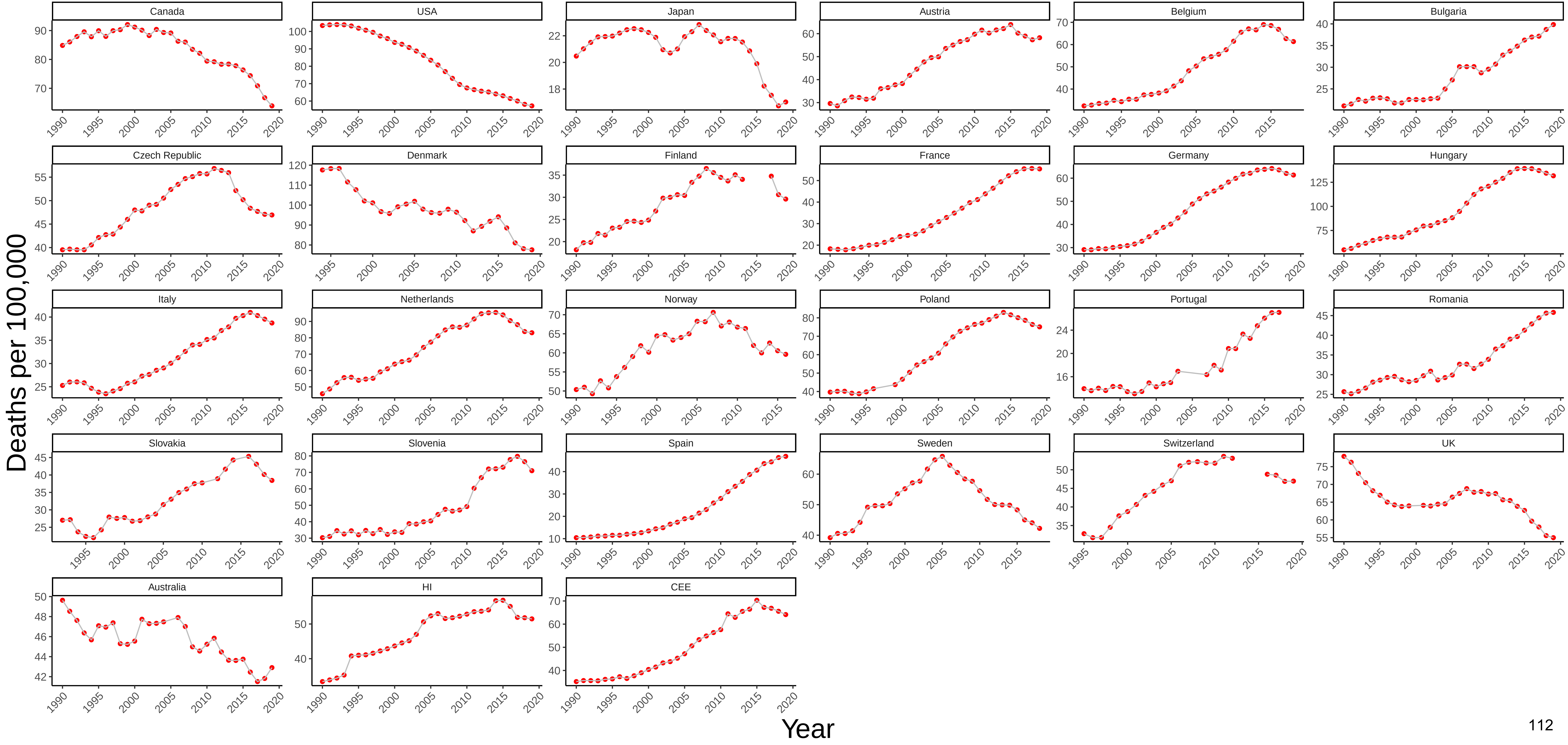

### Figure 65 Three−Year Moving Average of Male Mortality from All Other Cancers at Ages 55−64

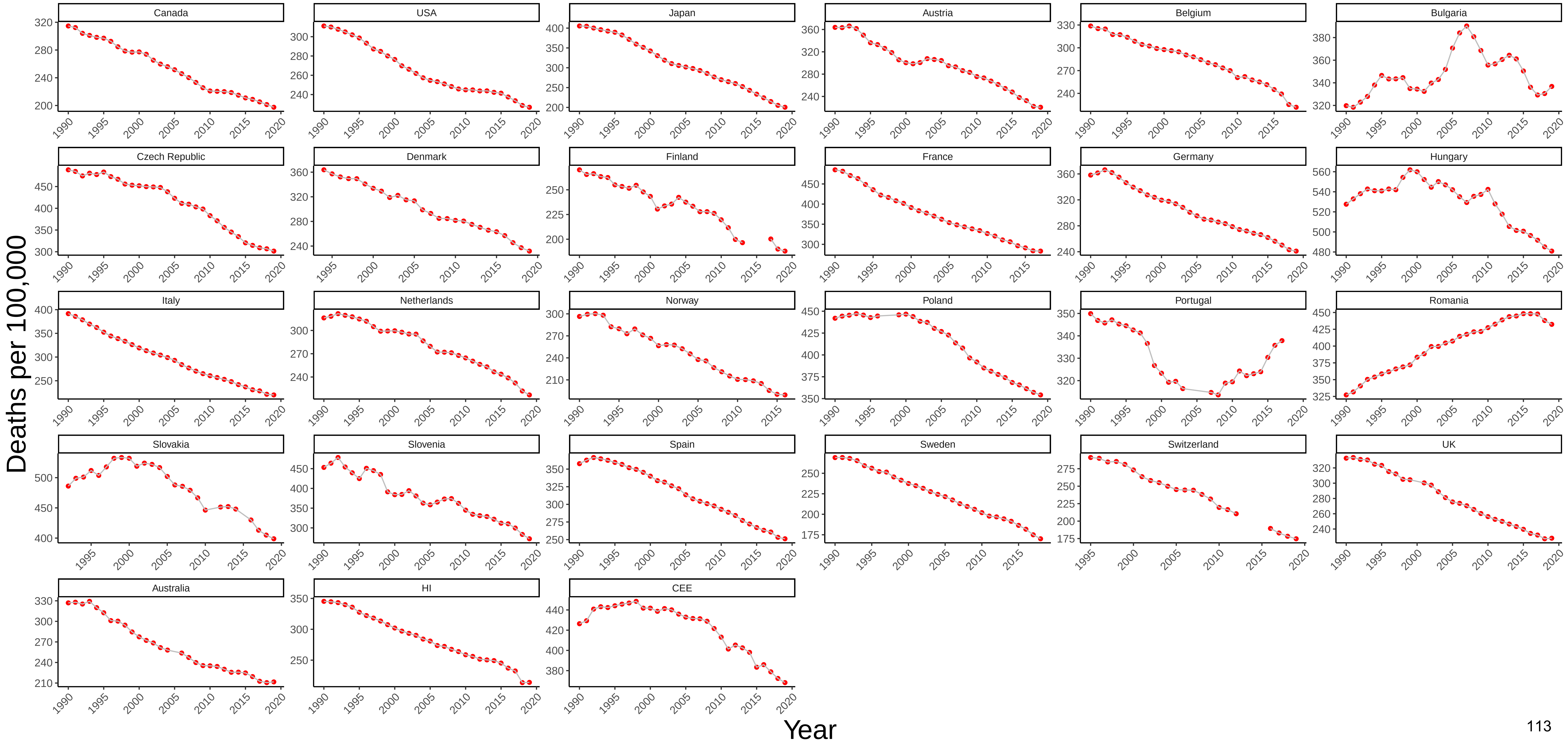

### Figure 66 Three−Year Moving Average of Female Mortality from All Other Cancers at Ages 55−64

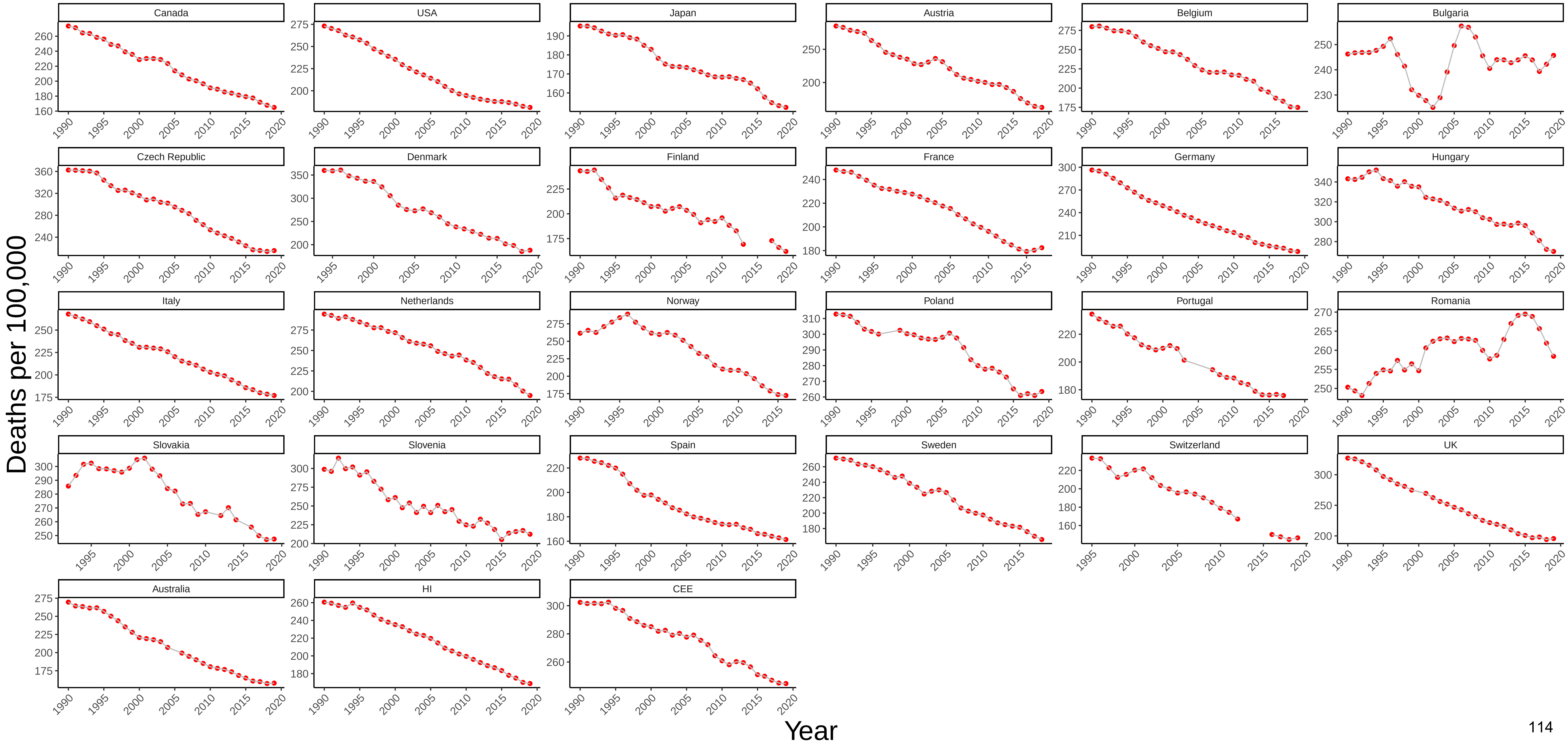

Figure 67 Three−Year Moving Average of Male Mortality from Nervous System Diseases at Ages 55−64

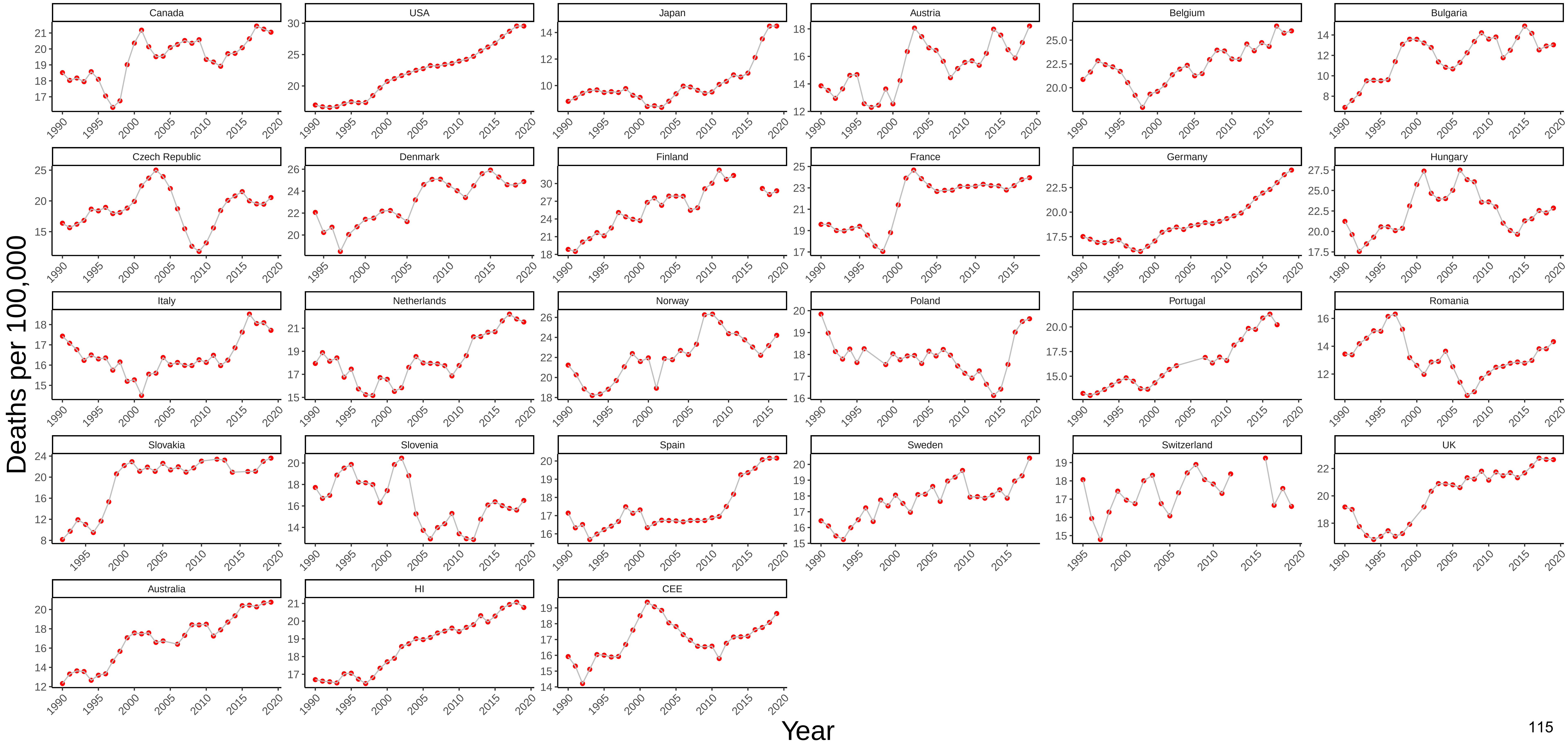

### Figure 68 Three−Year Moving Average of Female Mortality from Nervous System Diseases at Ages 55−64

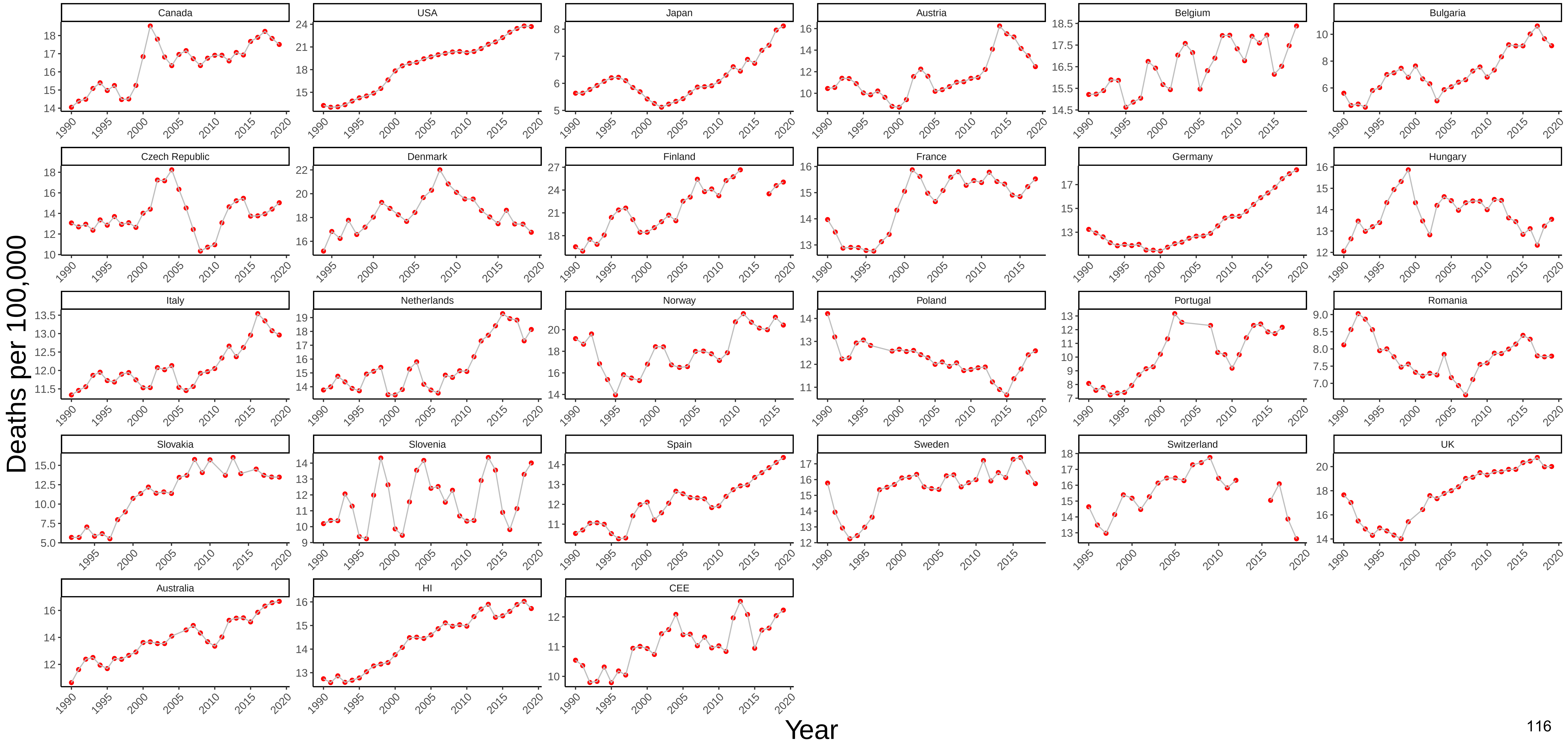

### Figure 69 Three−Year Moving Average of Male Mortality from Metabolic Diseases at Ages 55−64

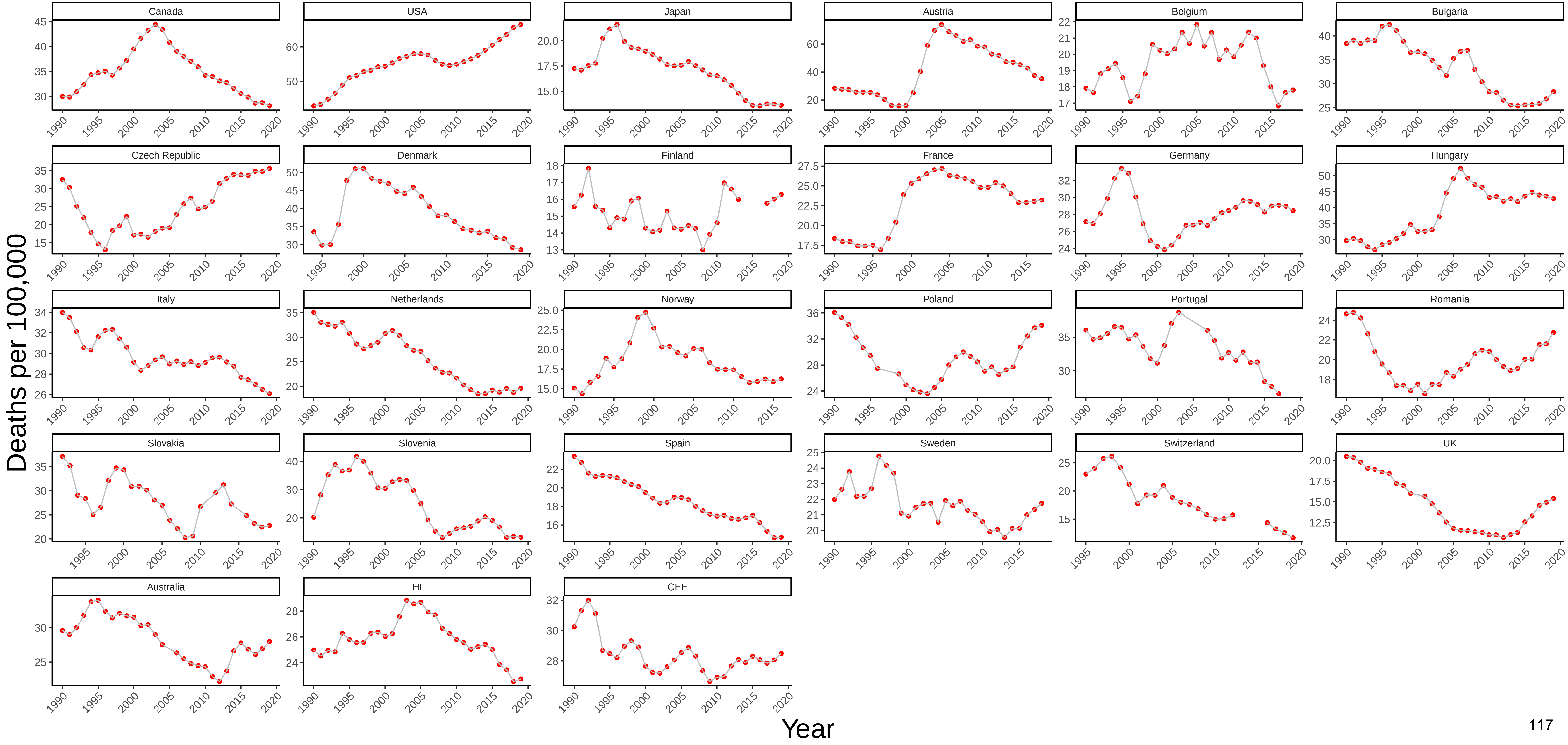

### Figure 70 Three−Year Moving Average of Female Mortality from Metabolic Diseases at Ages 55−64

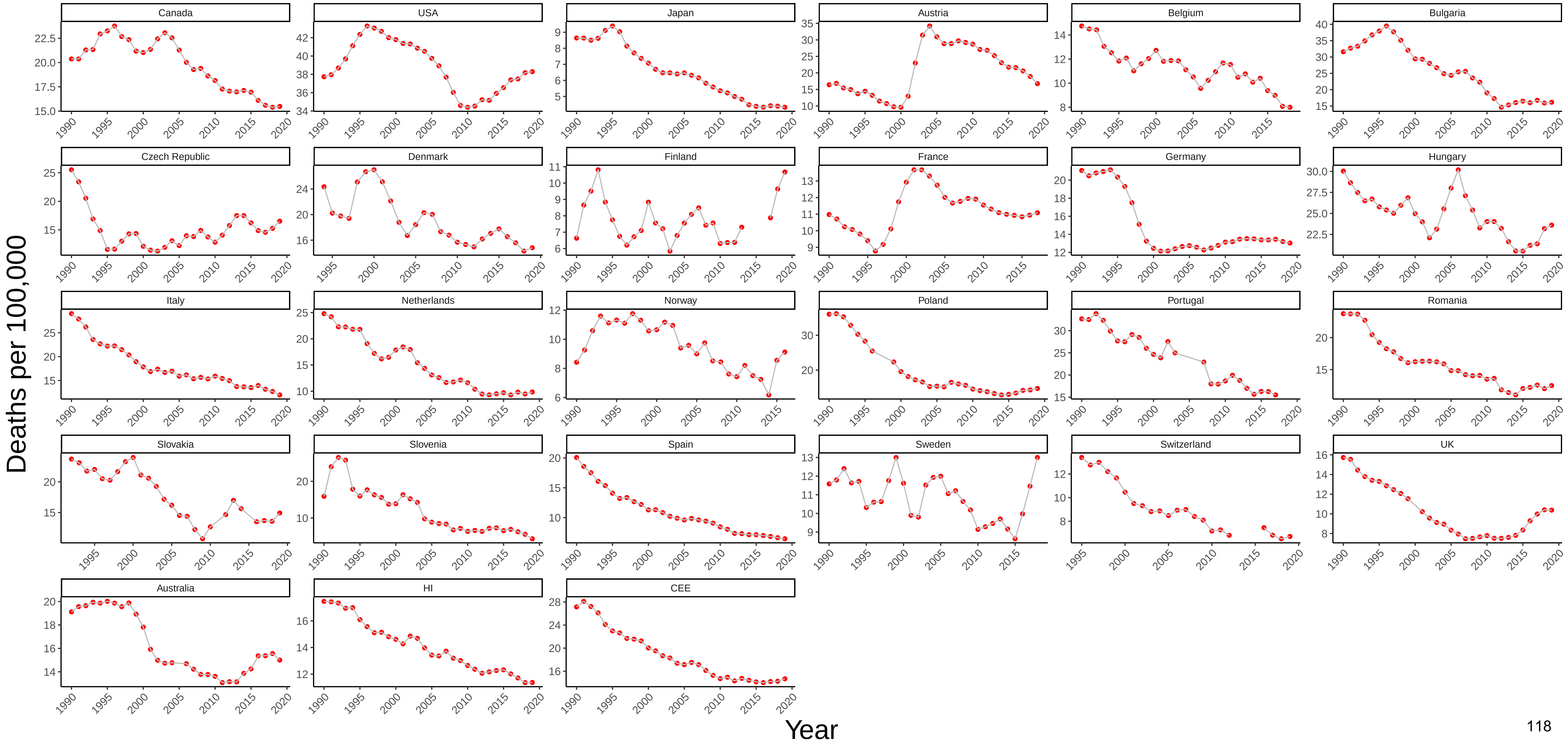

### Figure 71 Three−Year Moving Average of Male Mortality from Cardiovascular Disease at Ages 55−64

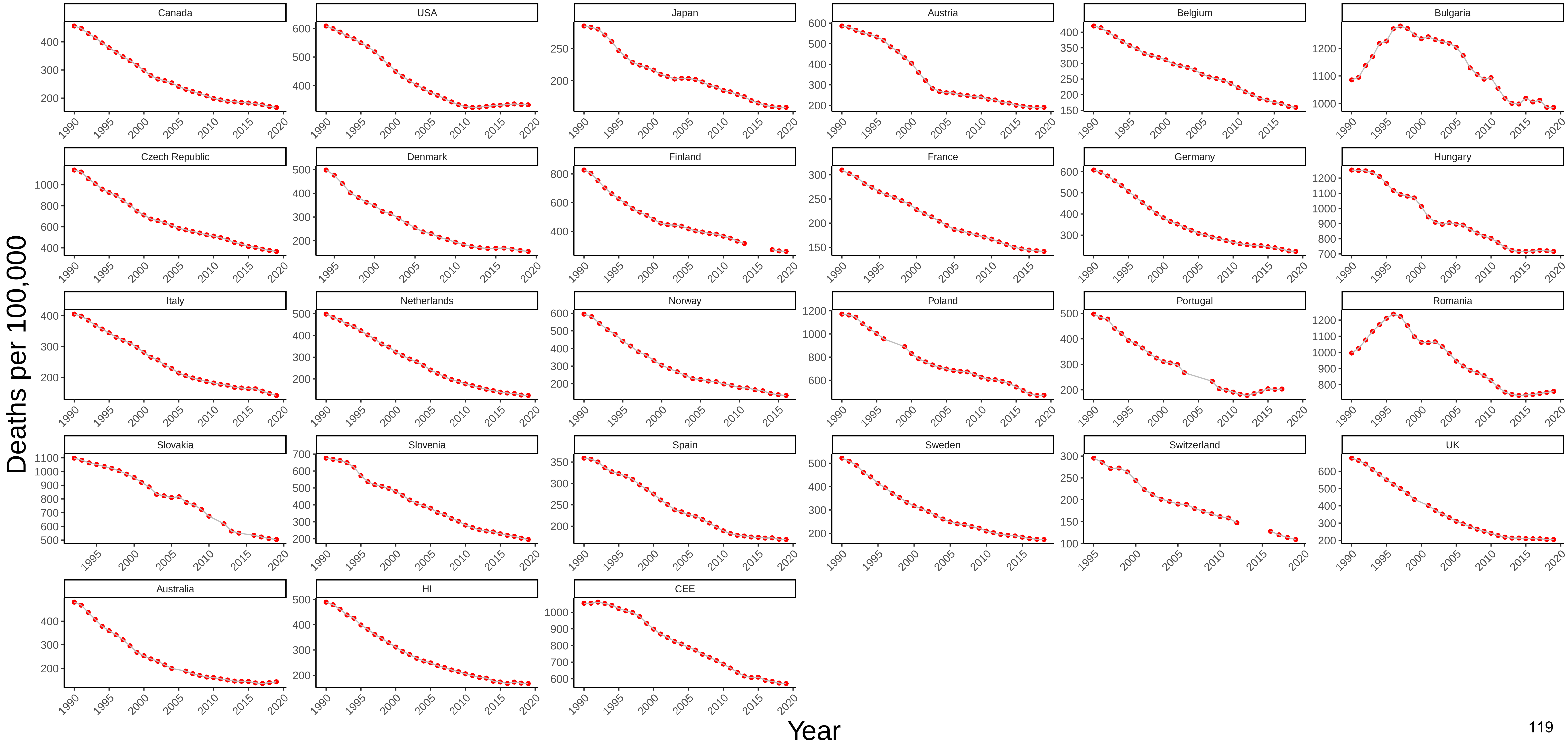

Figure 72 Three−Year Moving Average of Female Mortality from Cardiovascular Disease at Ages 55−64

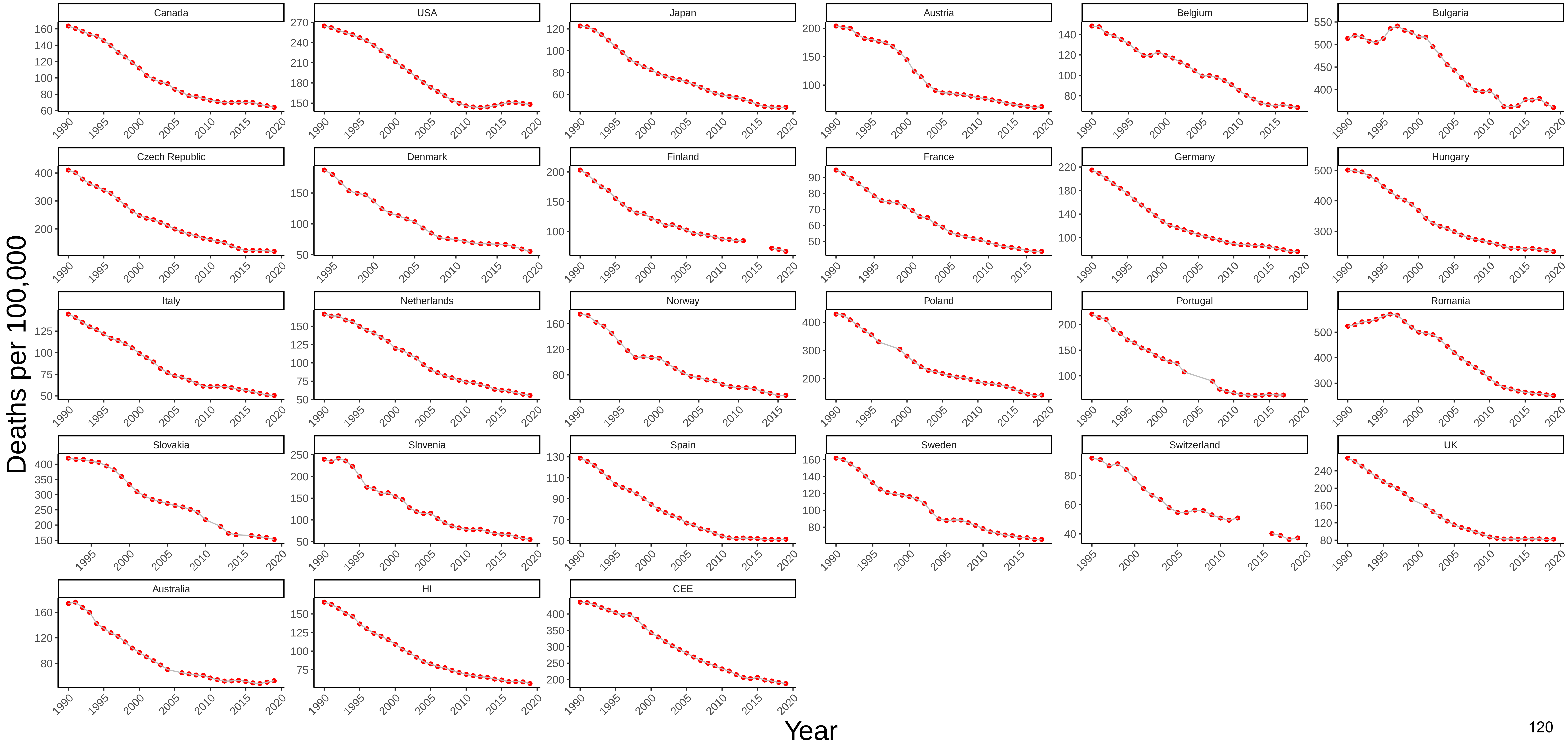

### Figure 73 Three−Year Moving Average of Male Mortality from Suicide at Ages 55−64

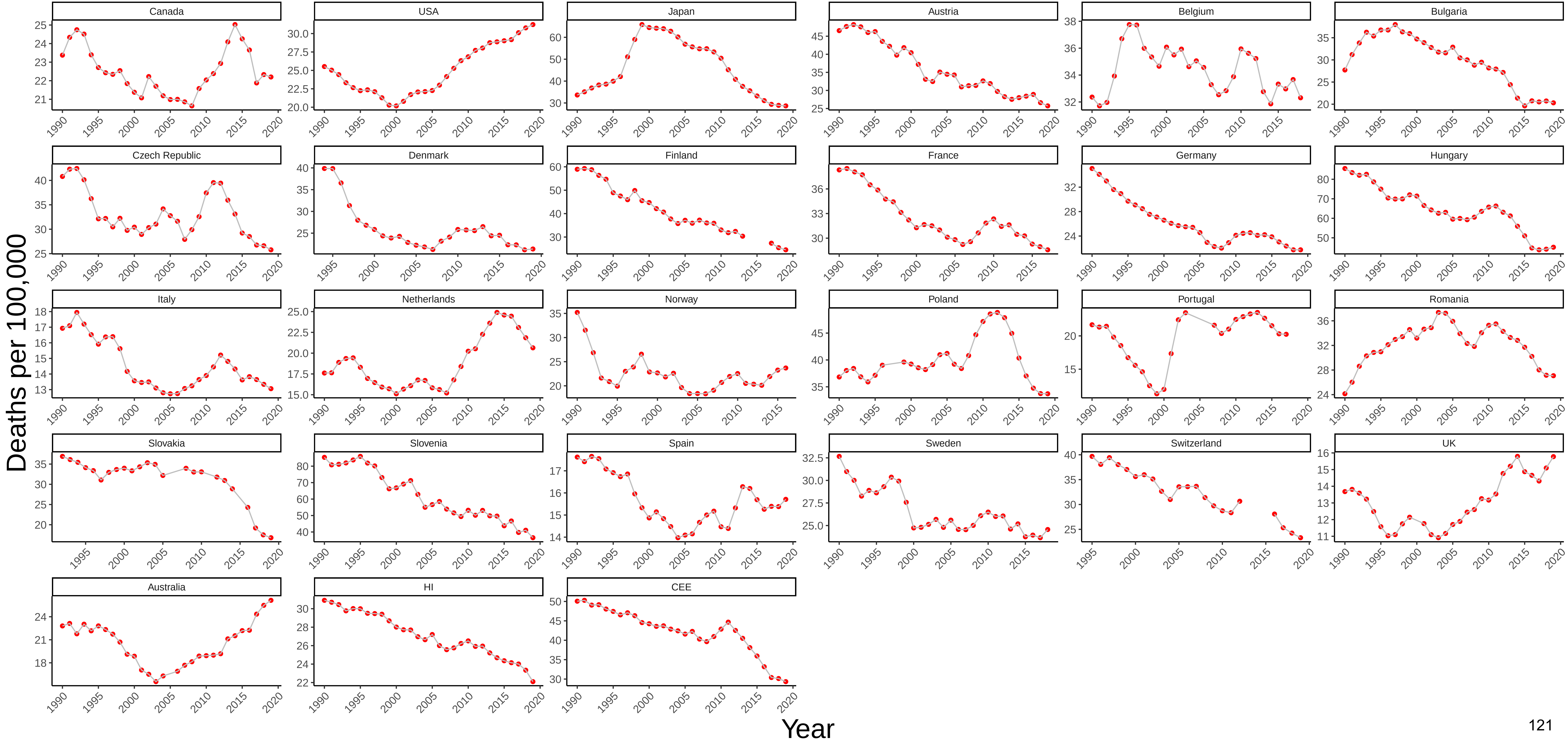

### Figure 74 Three−Year Moving Average of Female Mortality from Suicide at Ages 55−64

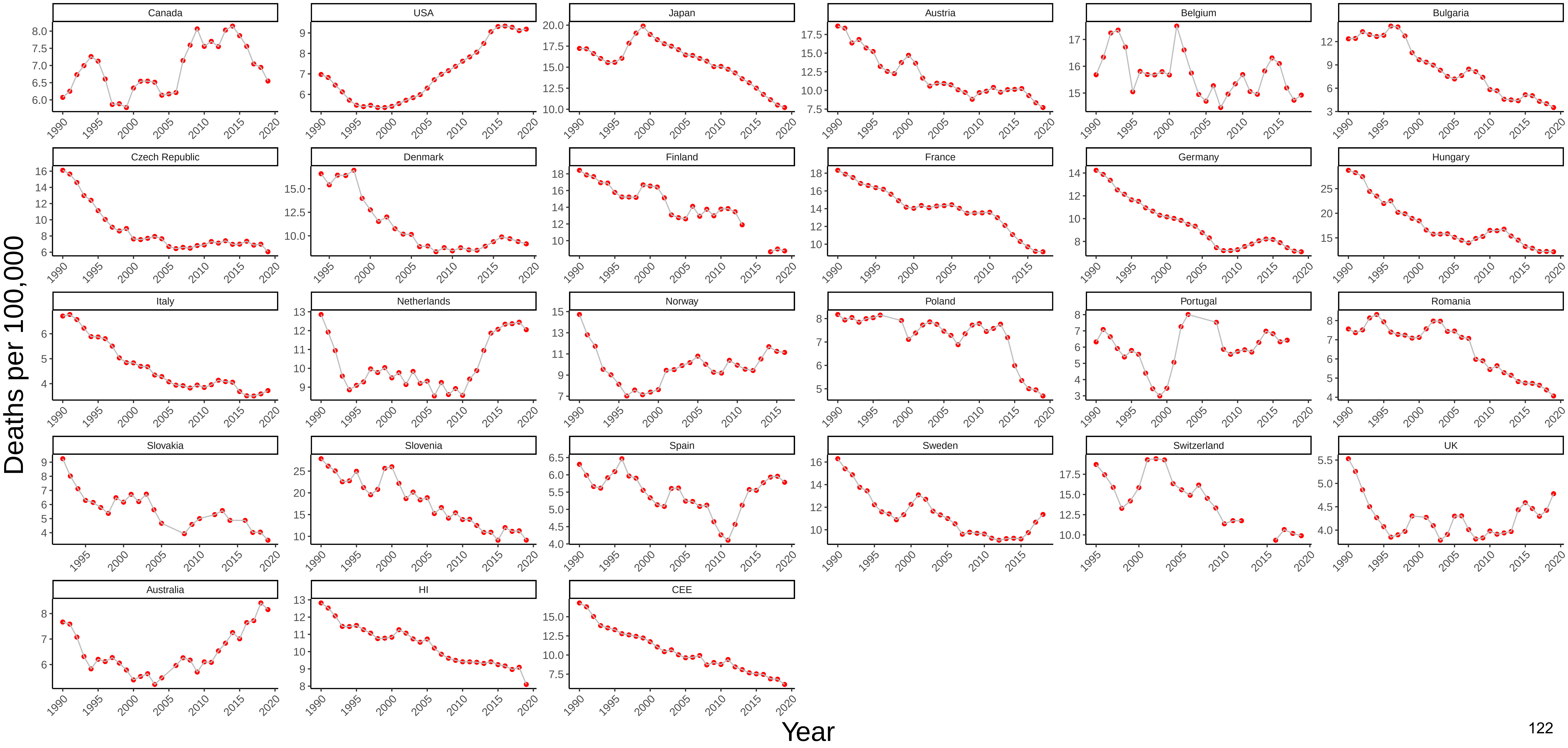

### Figure 75 Three−Year Moving Average of Male Mortality from Homicide at Ages 55−64

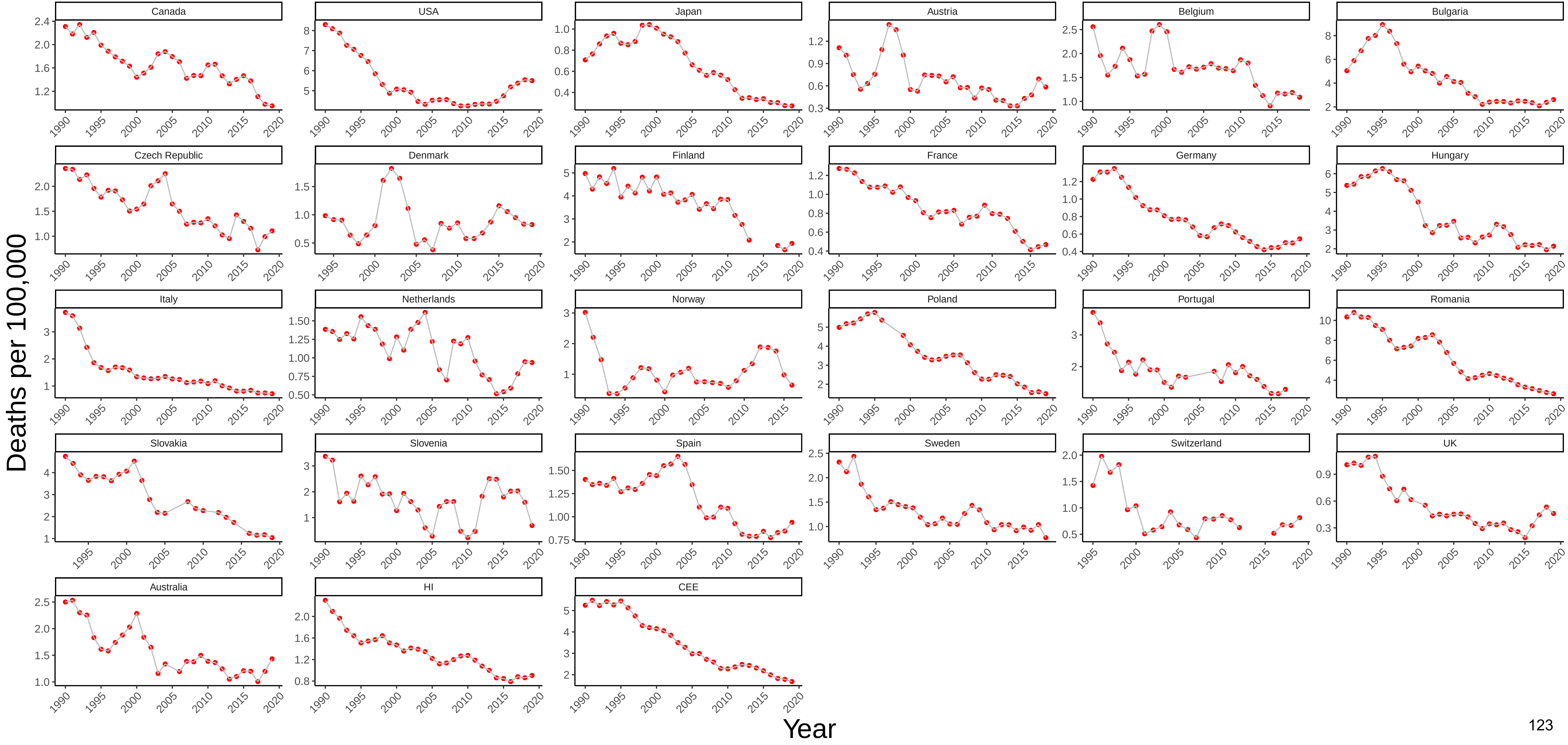

### Figure 76 Three−Year Moving Average of Female Mortality from Homicide at Ages 55−64

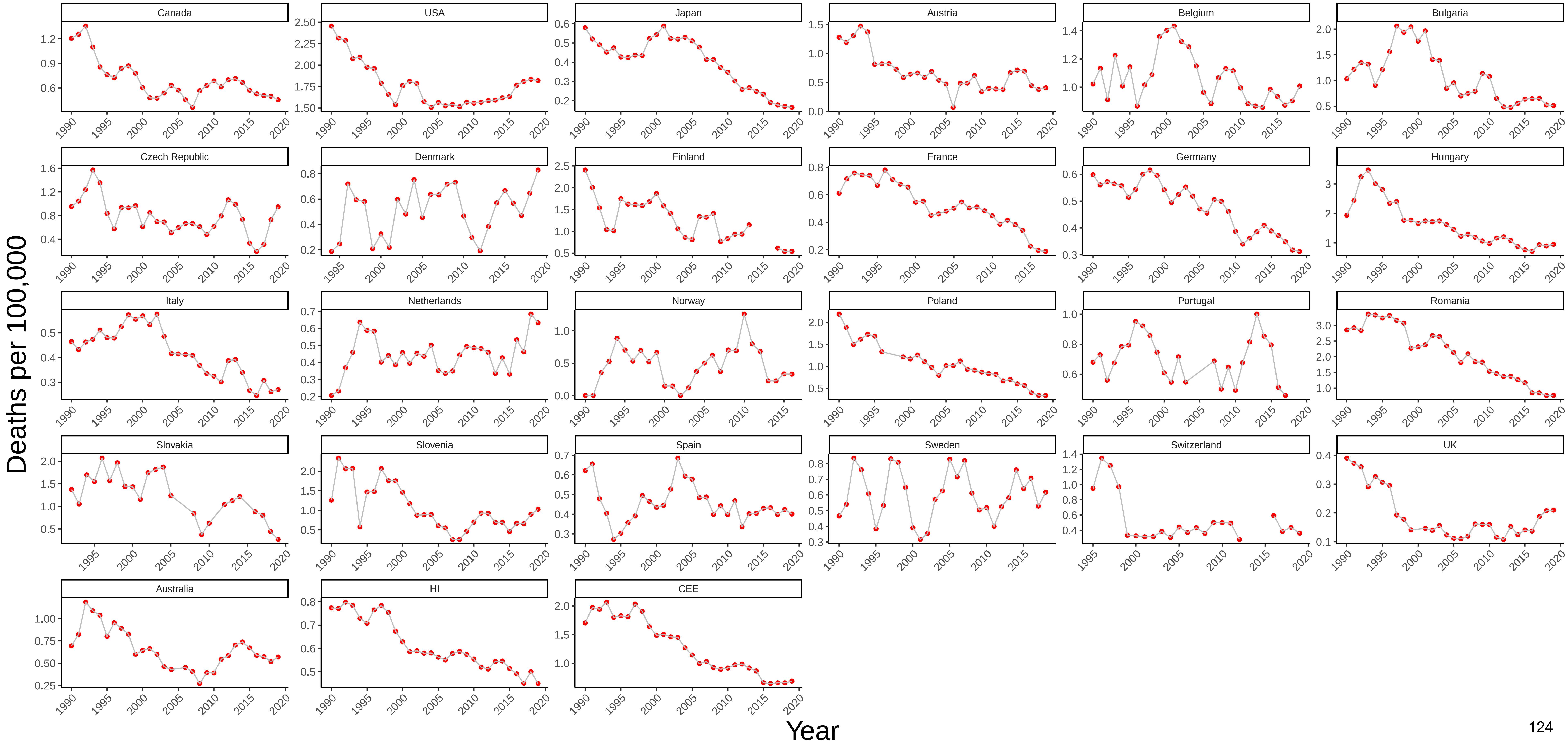

### Figure 77 Three−Year Moving Average of Male Mortality from Transport Accidents at Ages 55−64

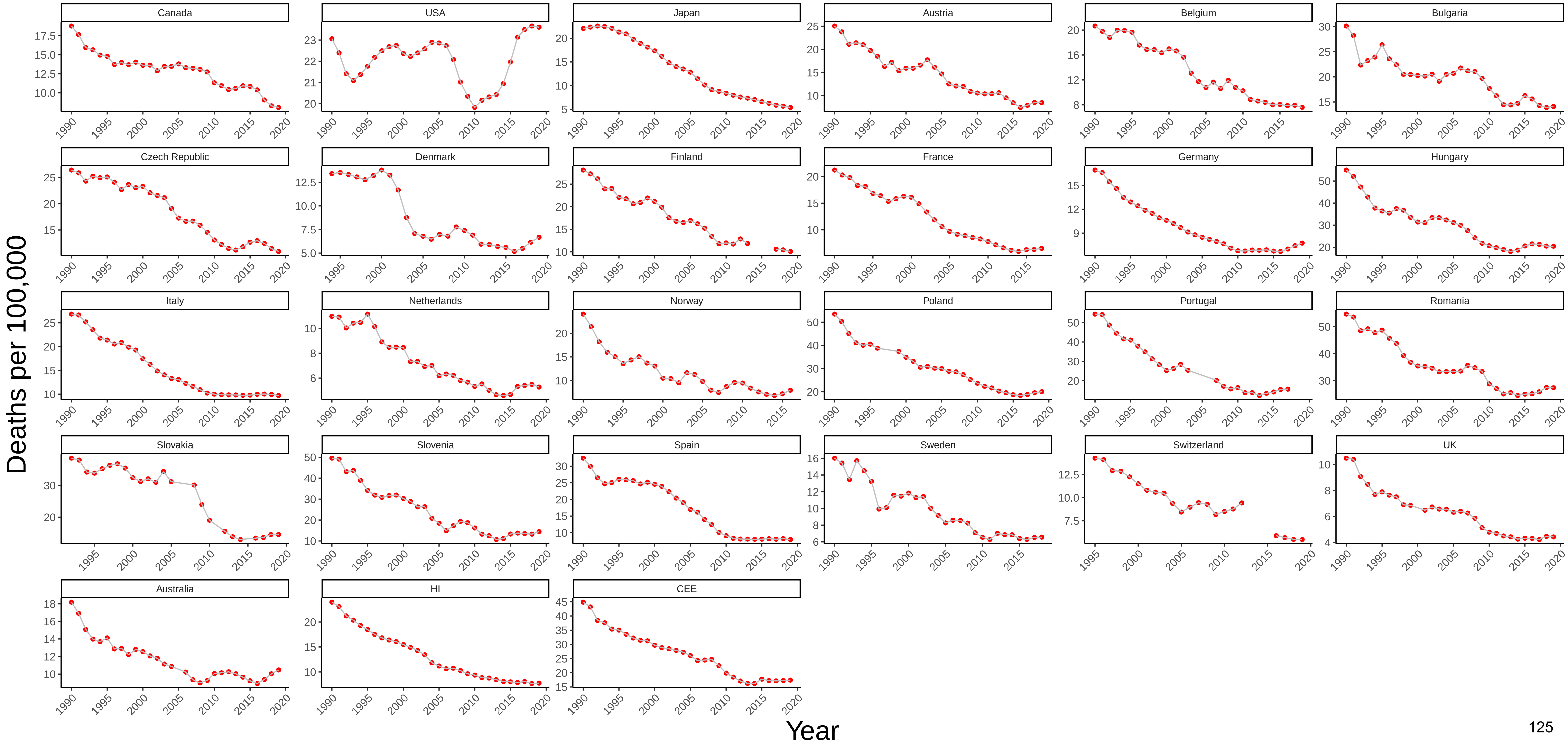

### Figure 78 Three−Year Moving Average of Female Mortality from Transport Accidents at Ages 55−64

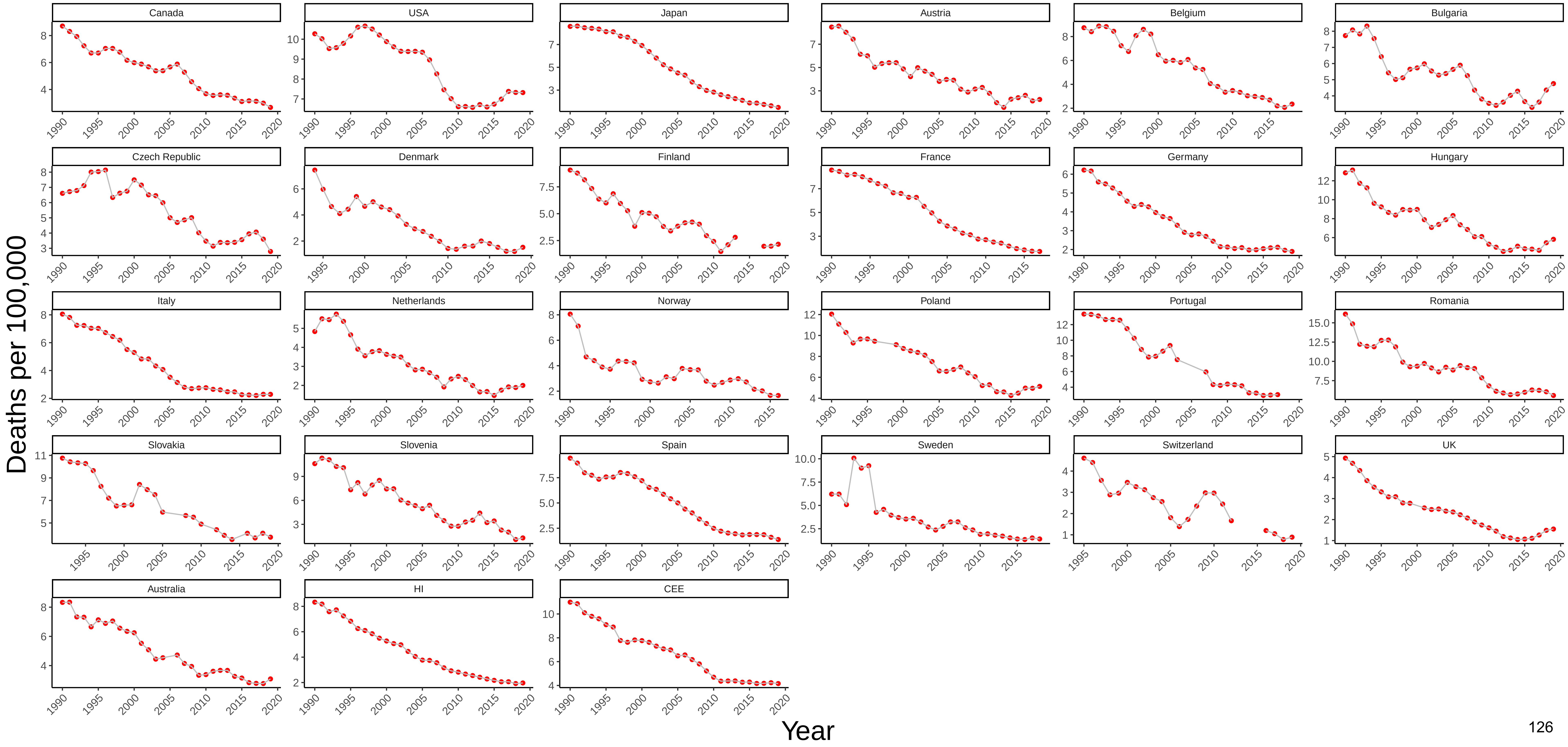

### Figure 79 Three−Year Moving Average of Male Mortality from Other External Causes at Ages 55−64

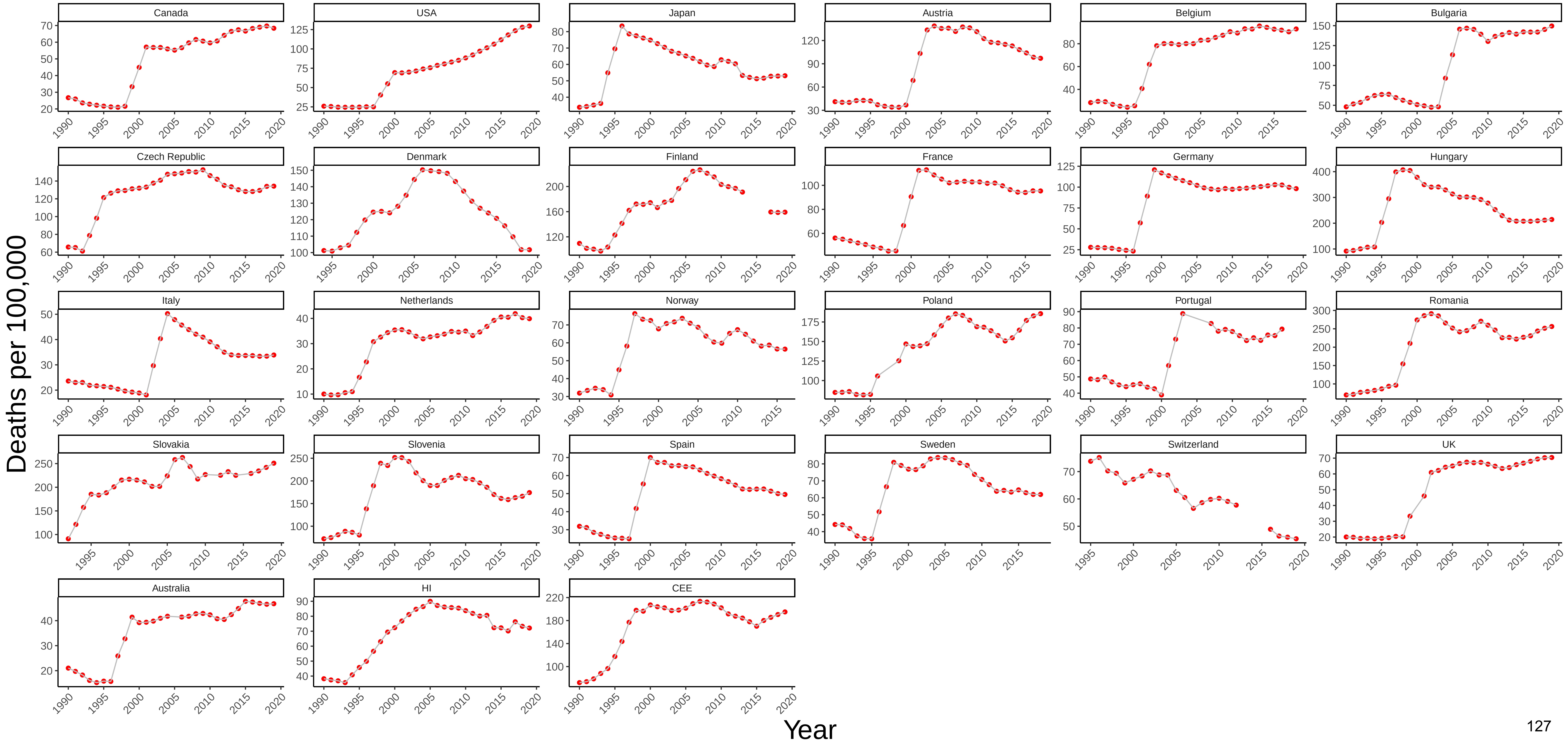

### Figure 80 Three−Year Moving Average of Female Mortality from Other External Causes at Ages 55−64

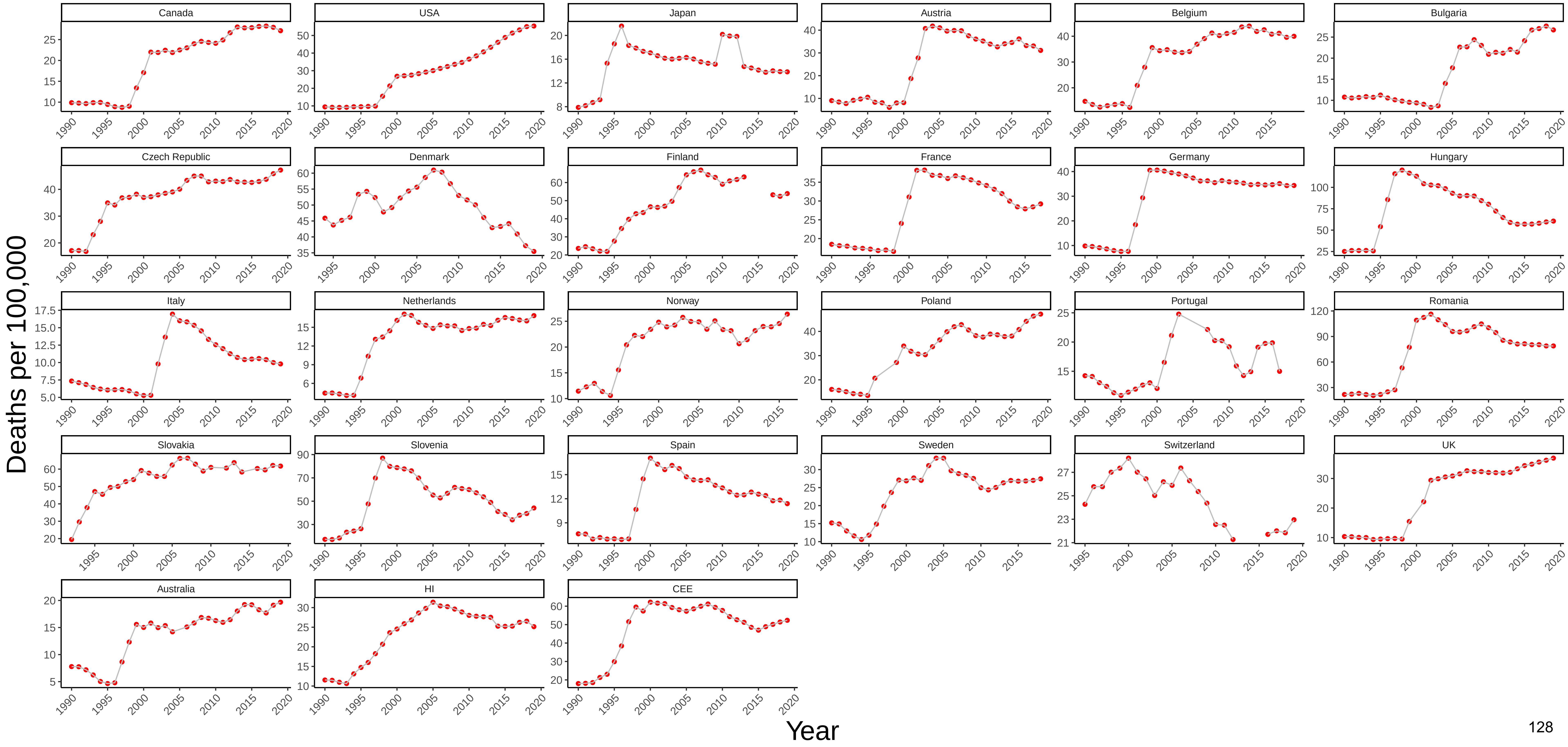

### Figure 81 Three−Year Moving Average of Male Mortality from All Other Causes at Ages 55−64

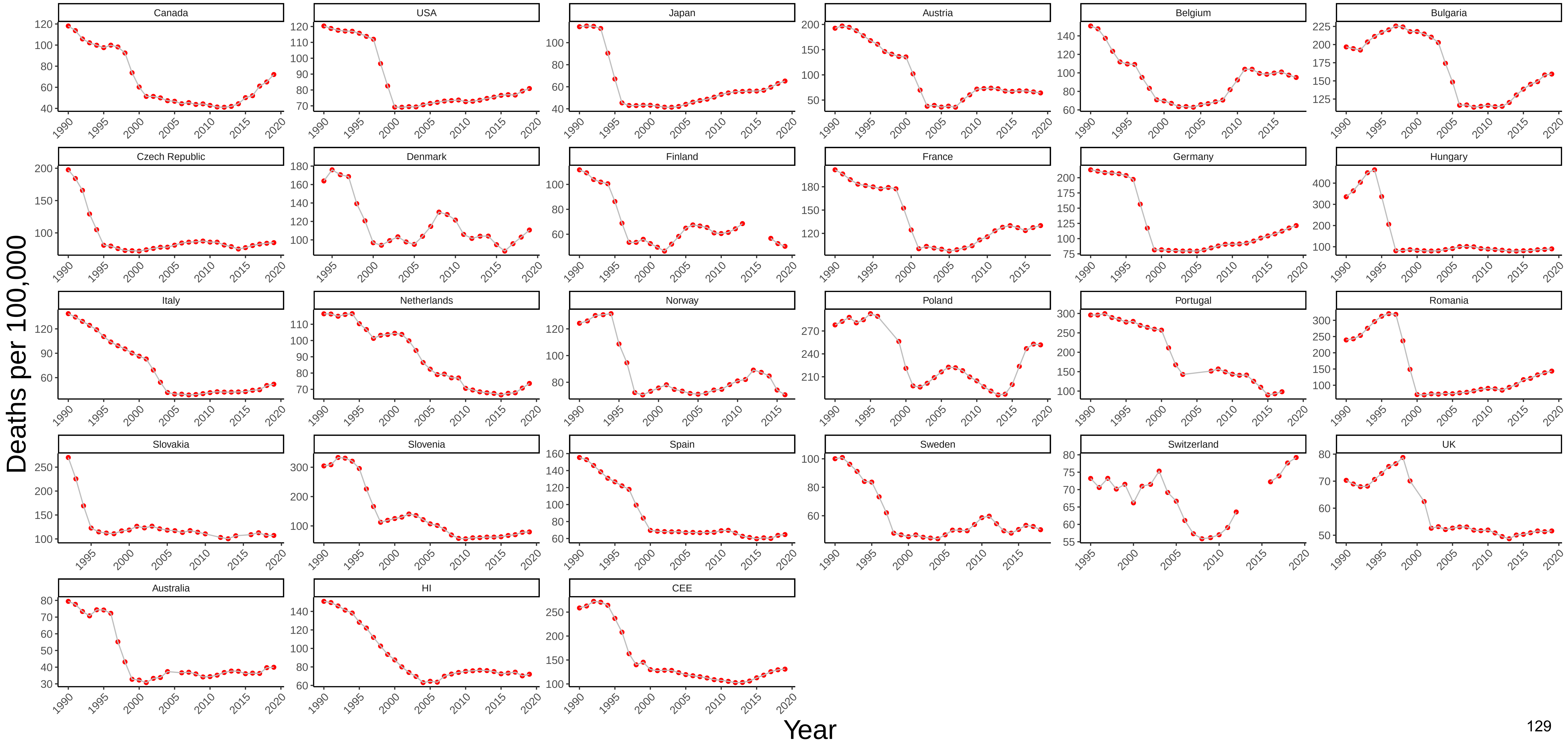

### Figure 82 Three−Year Moving Average of Female Mortality from All Other Causes at Ages 55−64

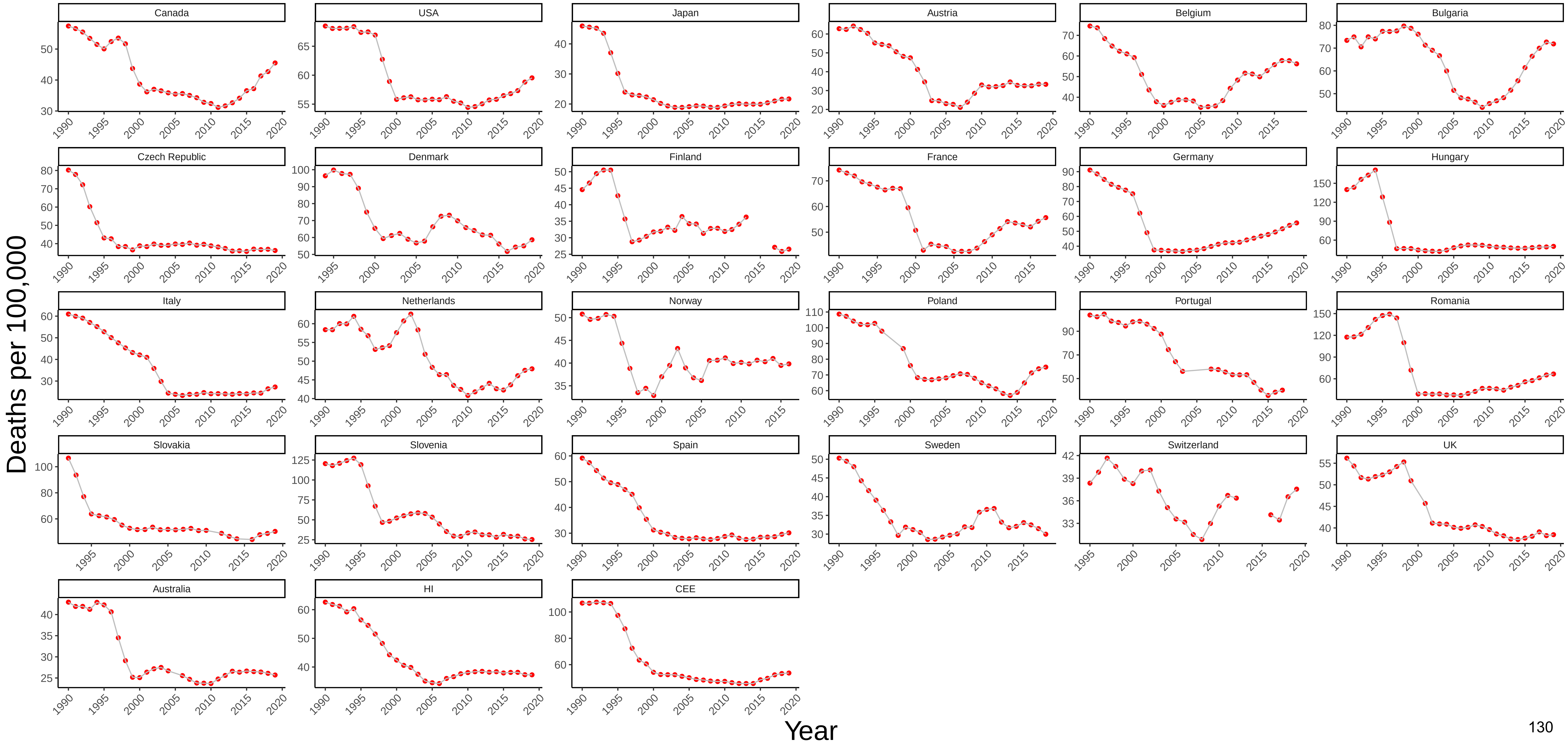

### Figure 83 Three−Year Moving Average of Male Mortality from All Causes at Ages 55−64

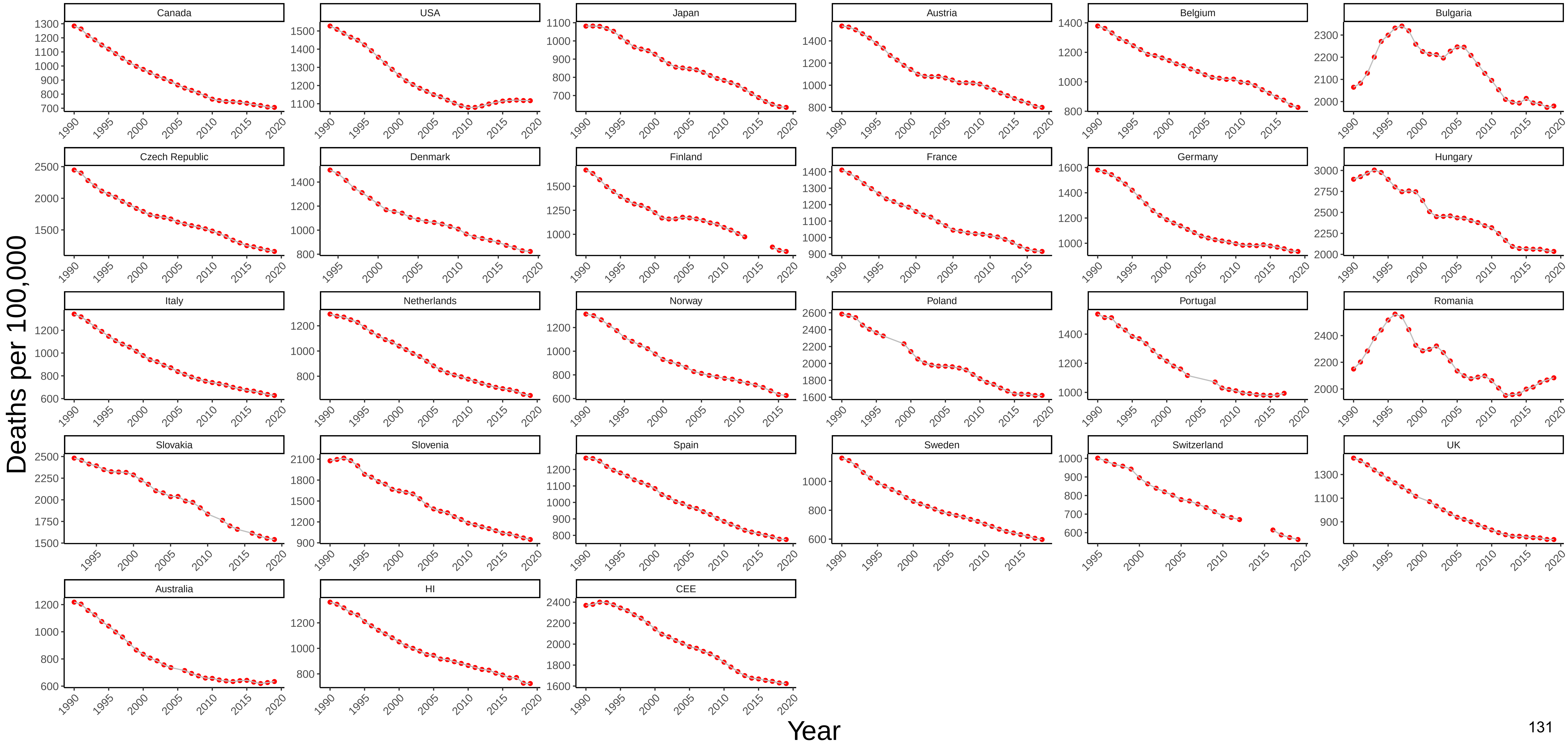

### Figure 84 Three−Year Moving Average of Female Mortality from All Causes at Ages 55−64

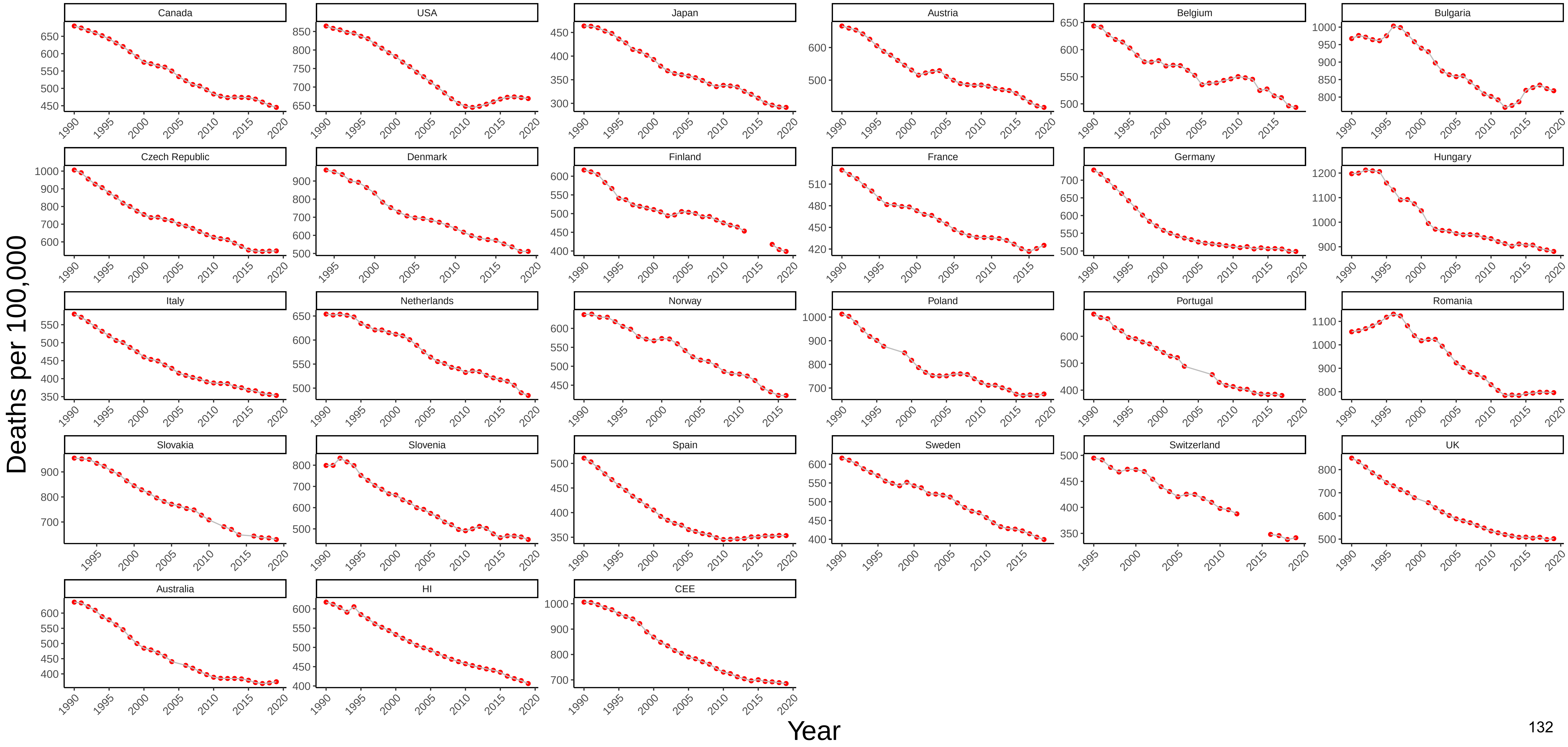

### Figure 85 Three−Year Moving Average of Male Mortality from Drug−Related Causes at Ages 25−44

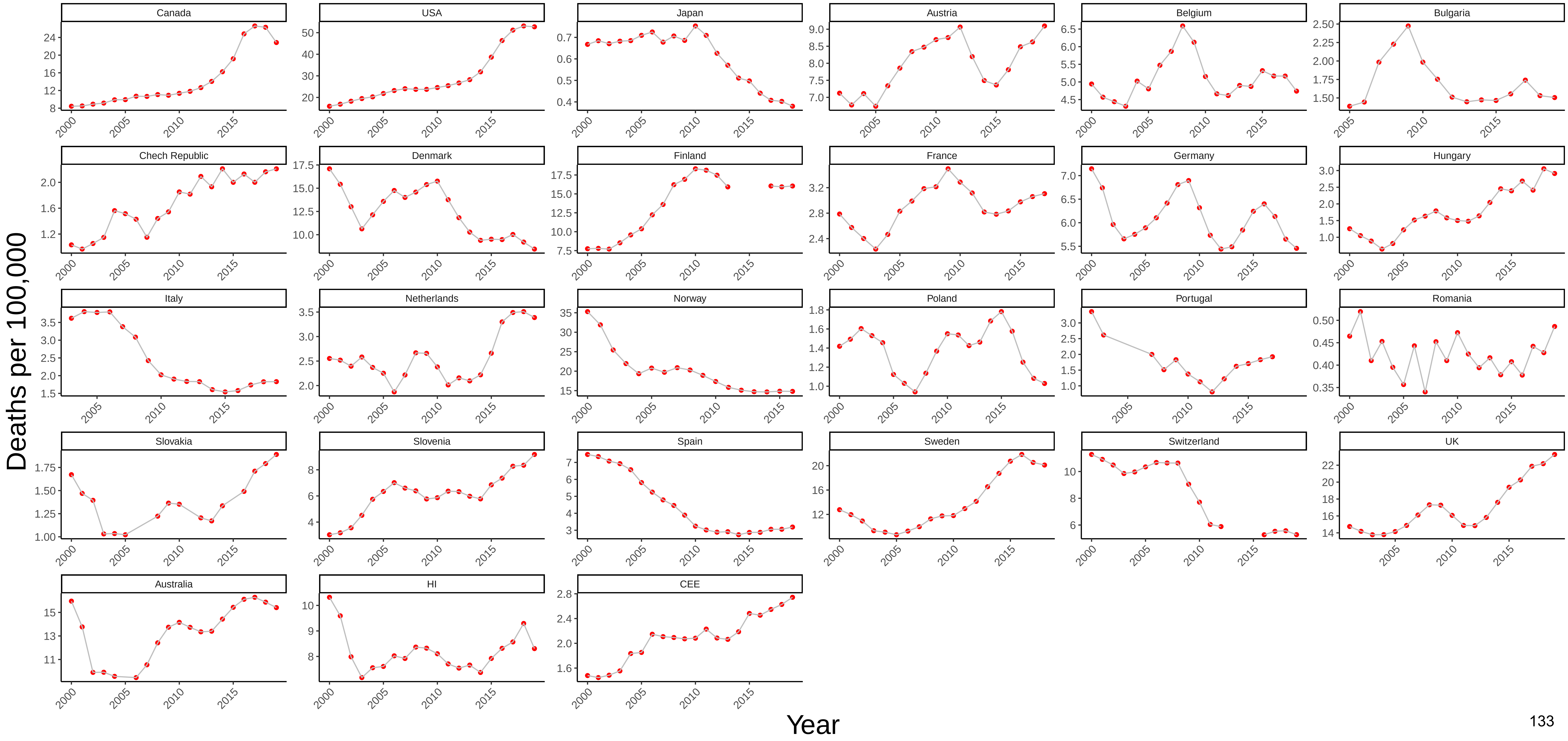

### Figure 86 Three−Year Moving Average of Female Mortality from Drug−Related Causes at Ages 25−44

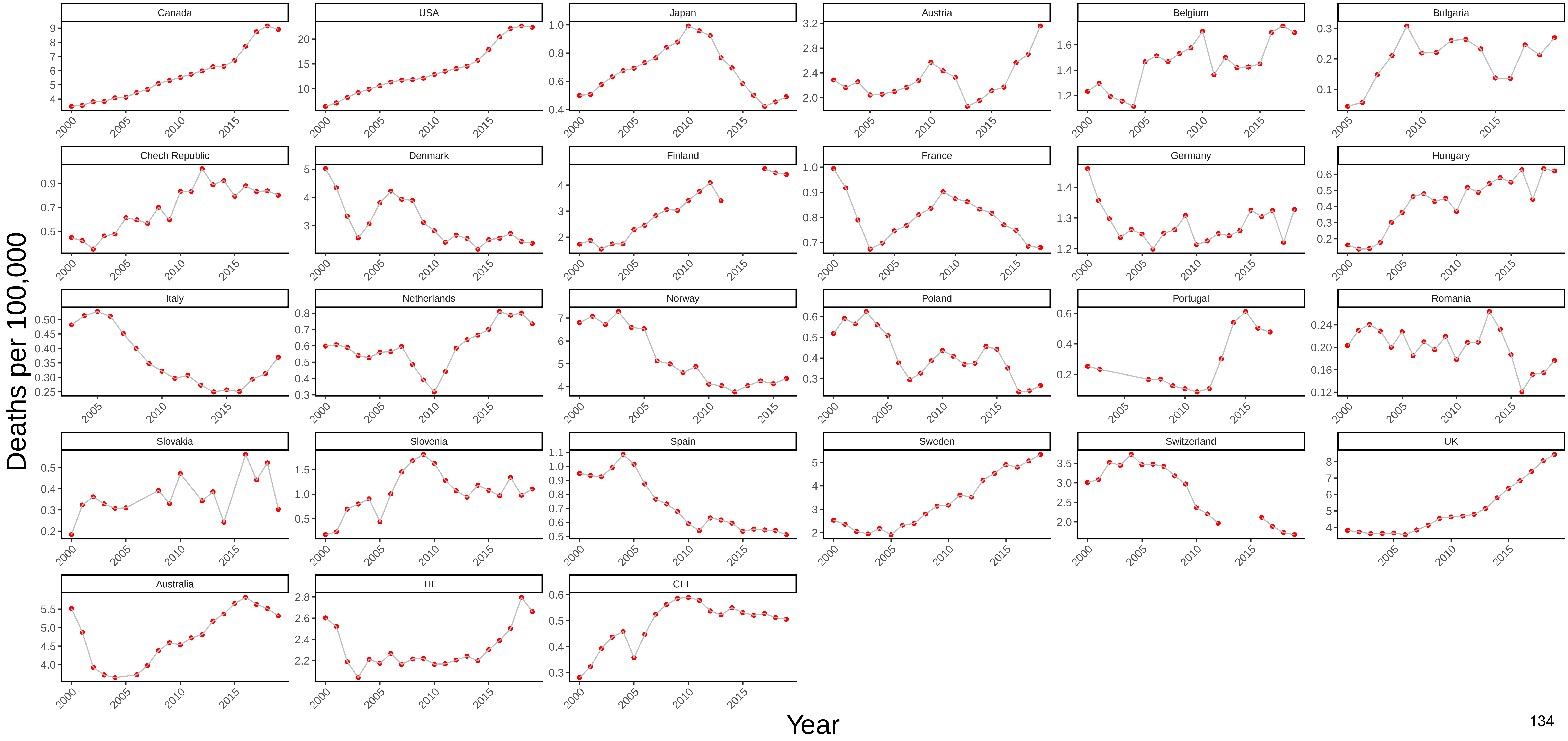

### Figure 87 Three−Year Moving Average of Male Mortality from Alcohol−Related Causes at Ages 25−44

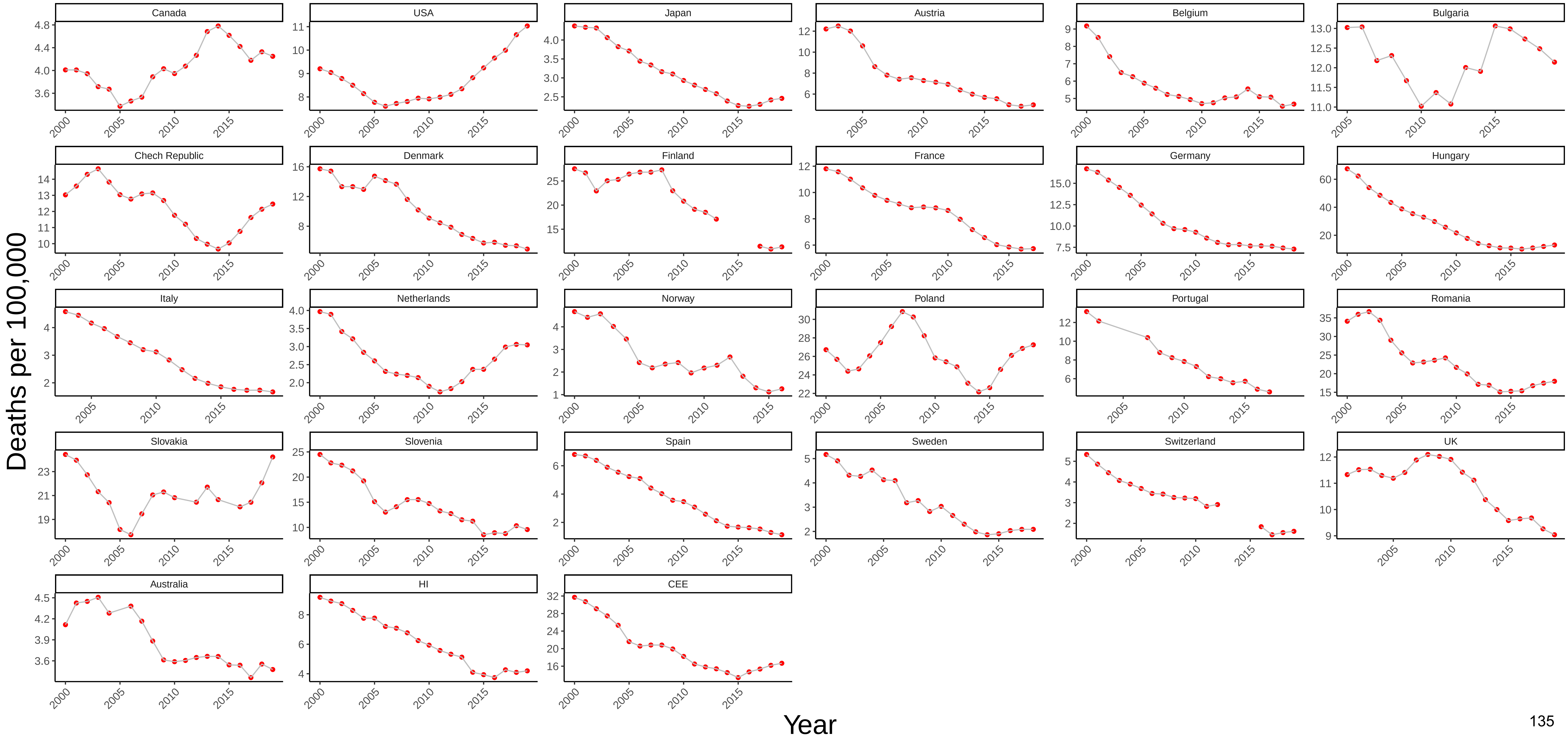

### Figure 88 Three−Year Moving Average of Female Mortality from Alcohol−Related Causes at Ages 25−44

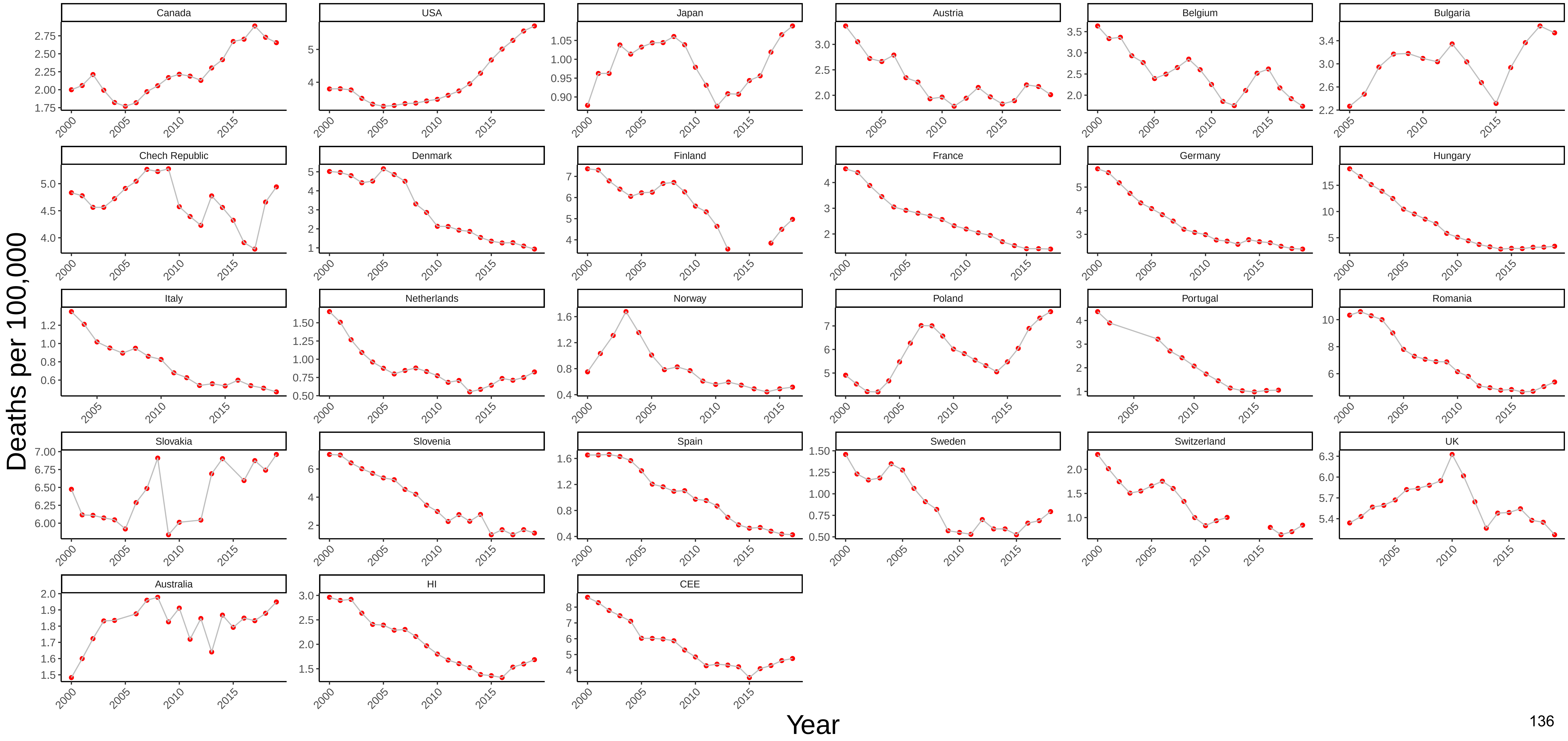

### Figure 89 Three−Year Moving Average of Male Mortality from Drug−Related Causes at Ages 45−54

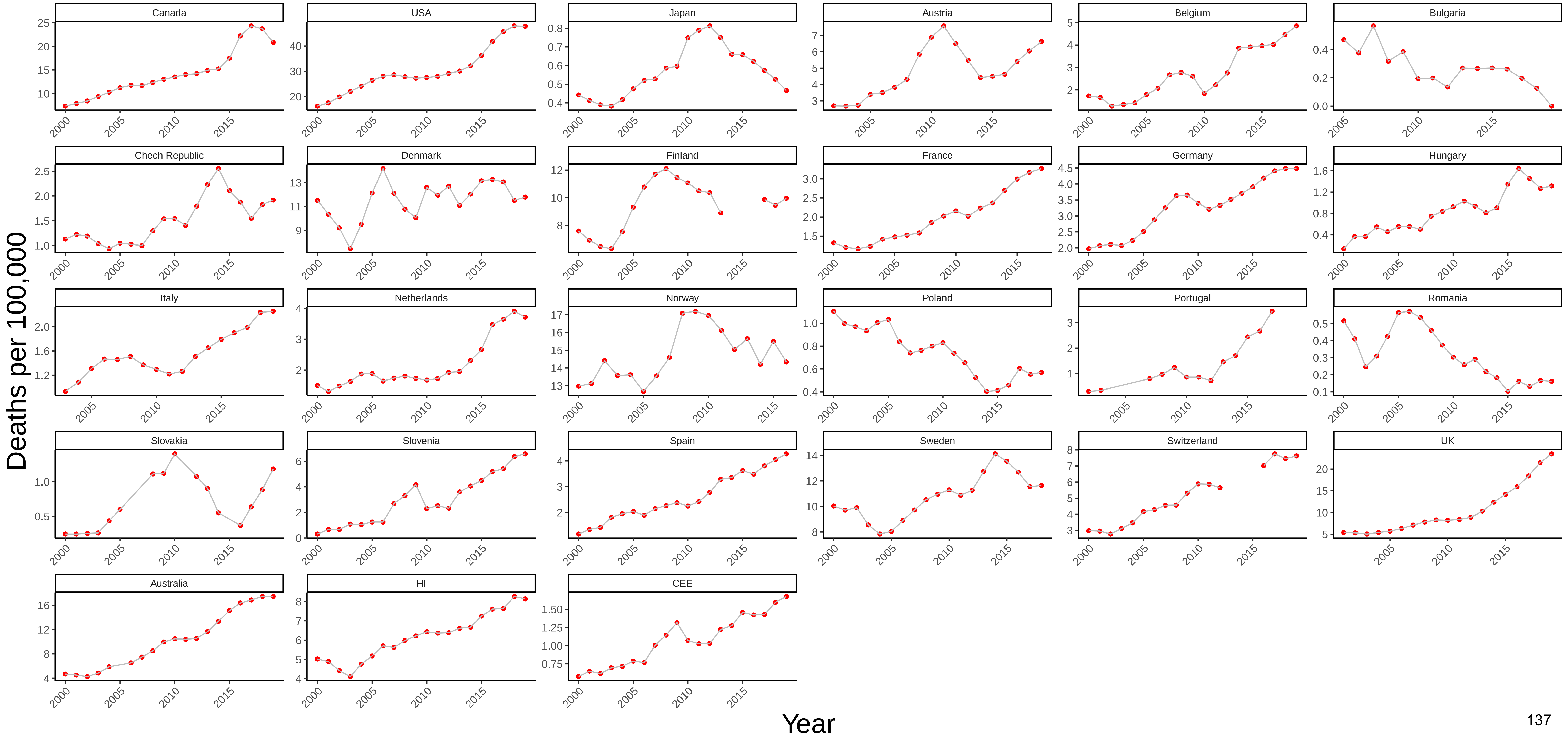

### Figure 90 Three−Year Moving Average of Female Mortality from Drug−Related Causes at Ages 45−54

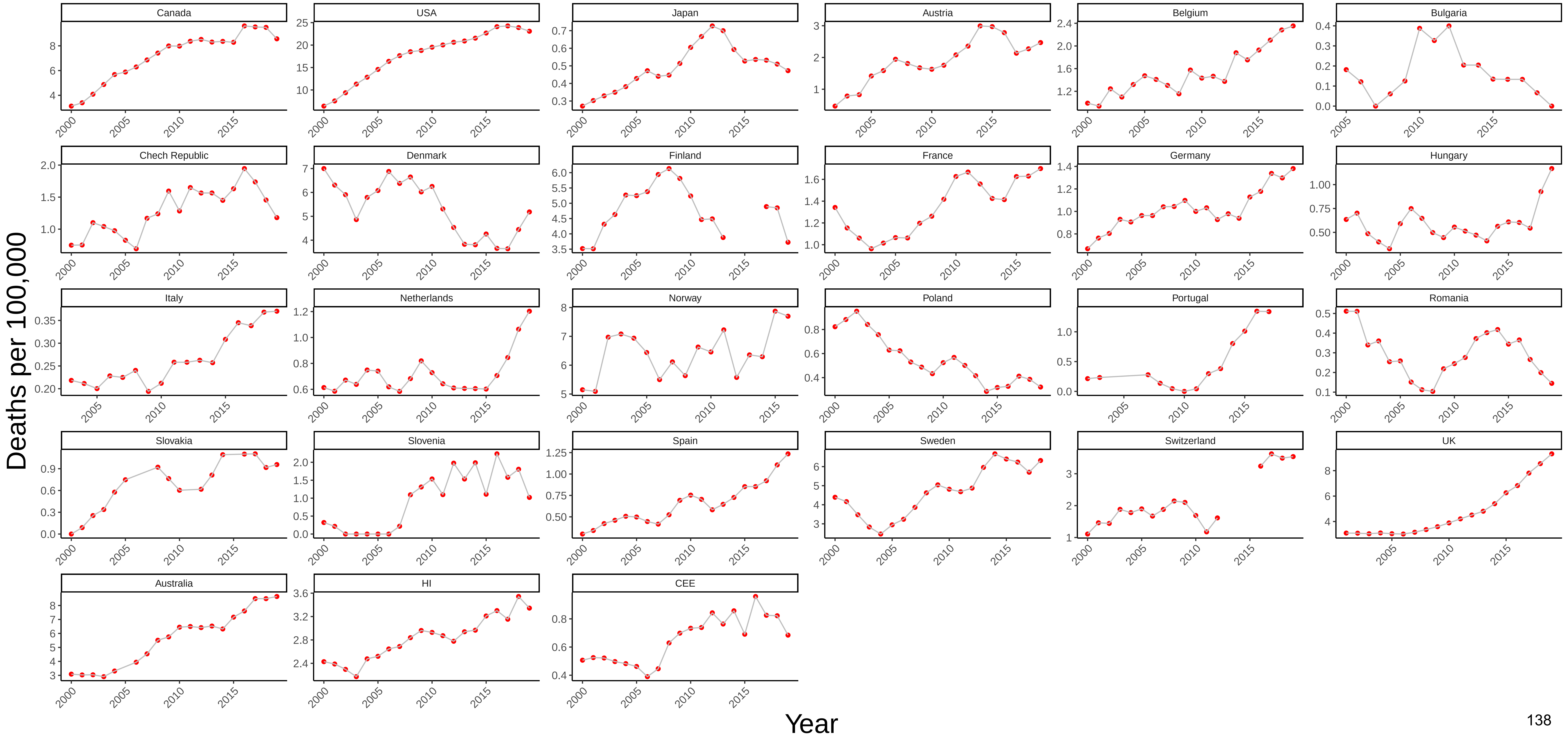

### Figure 91 Three−Year Moving Average of Male Mortality from Alcohol−Related Causes at Ages 45−54

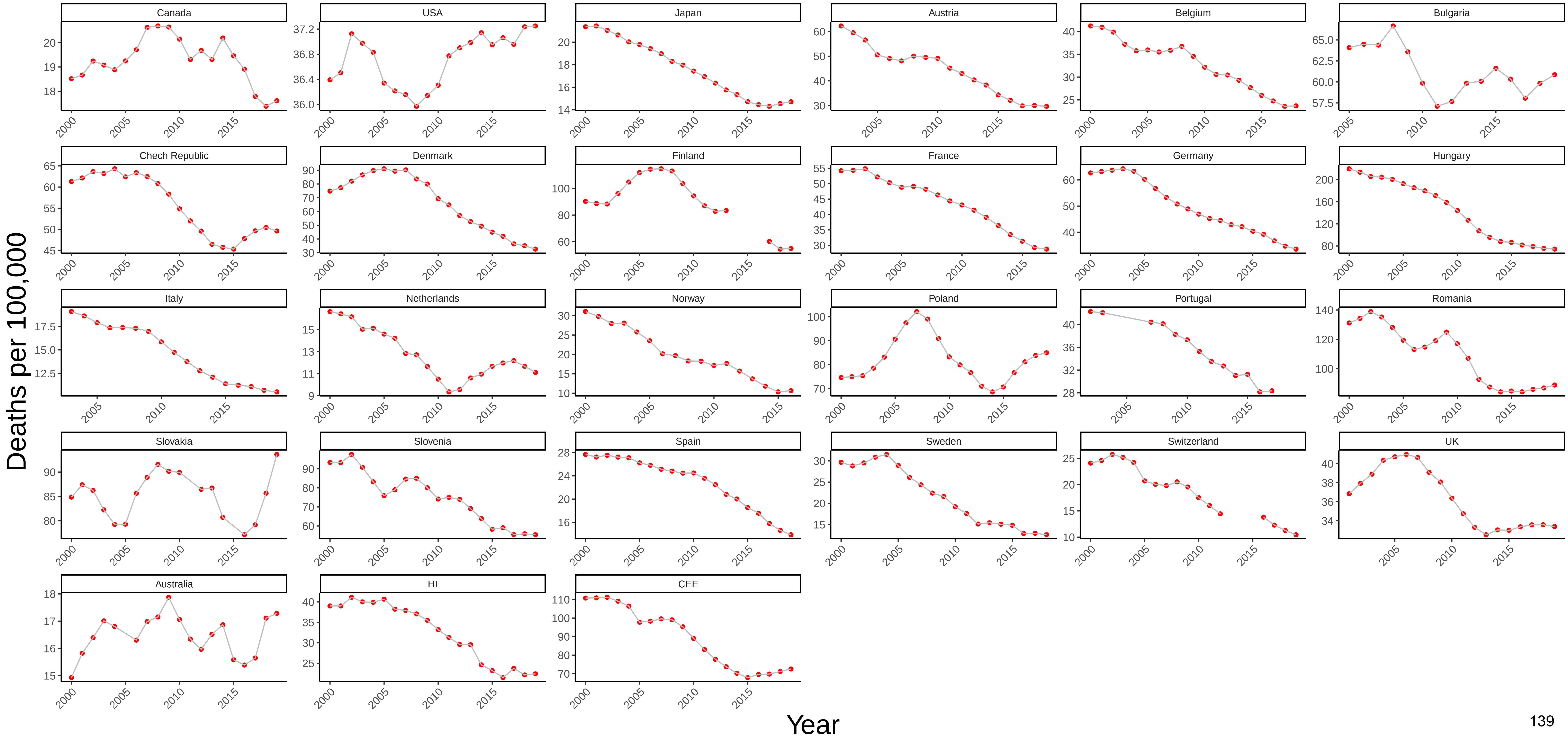

### Figure 92 Three−Year Moving Average of Female Mortality from Alcohol−Related Causes at Ages 45−54

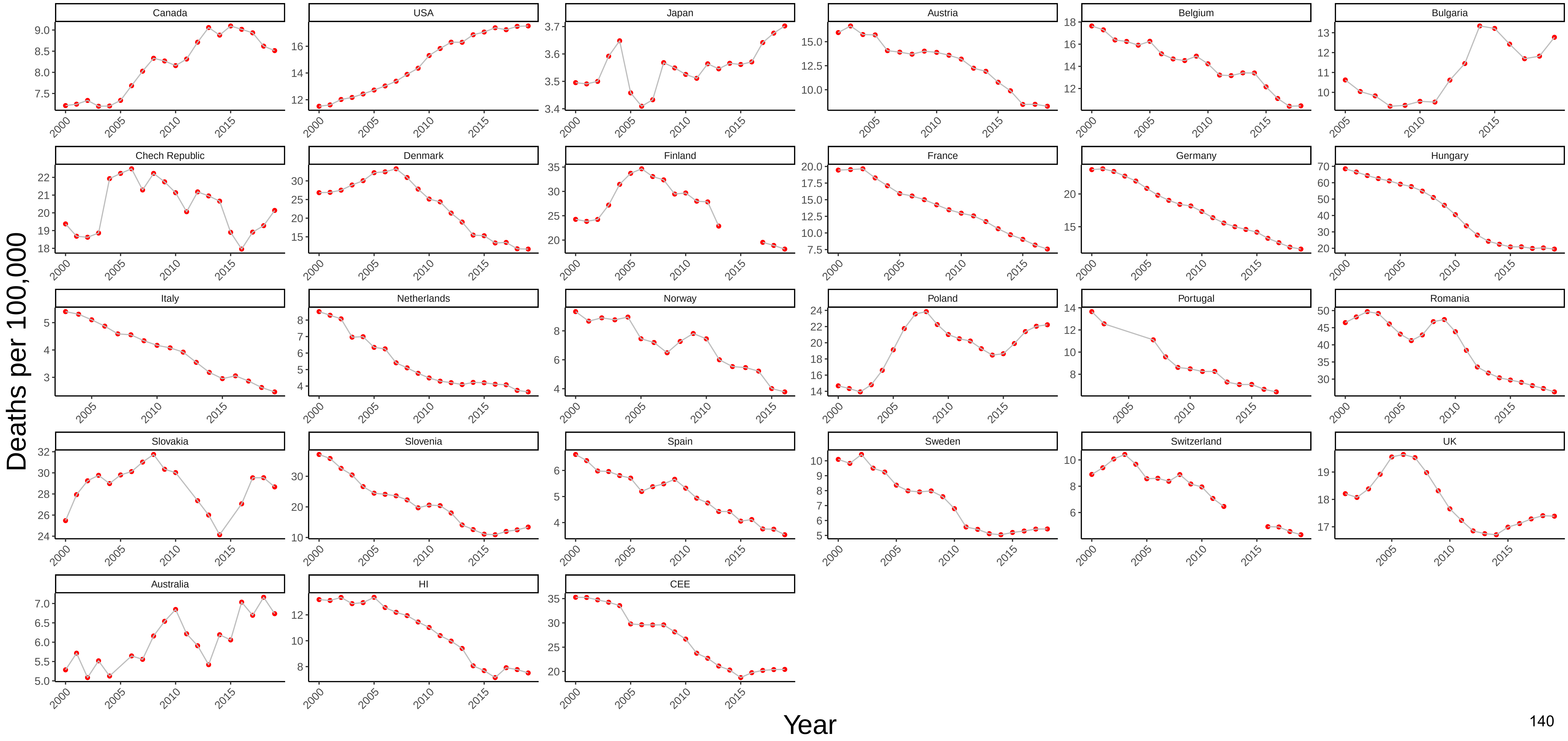

### Figure 93 Three−Year Moving Average of Male Mortality from Drug−Related Causes at Ages 55−64

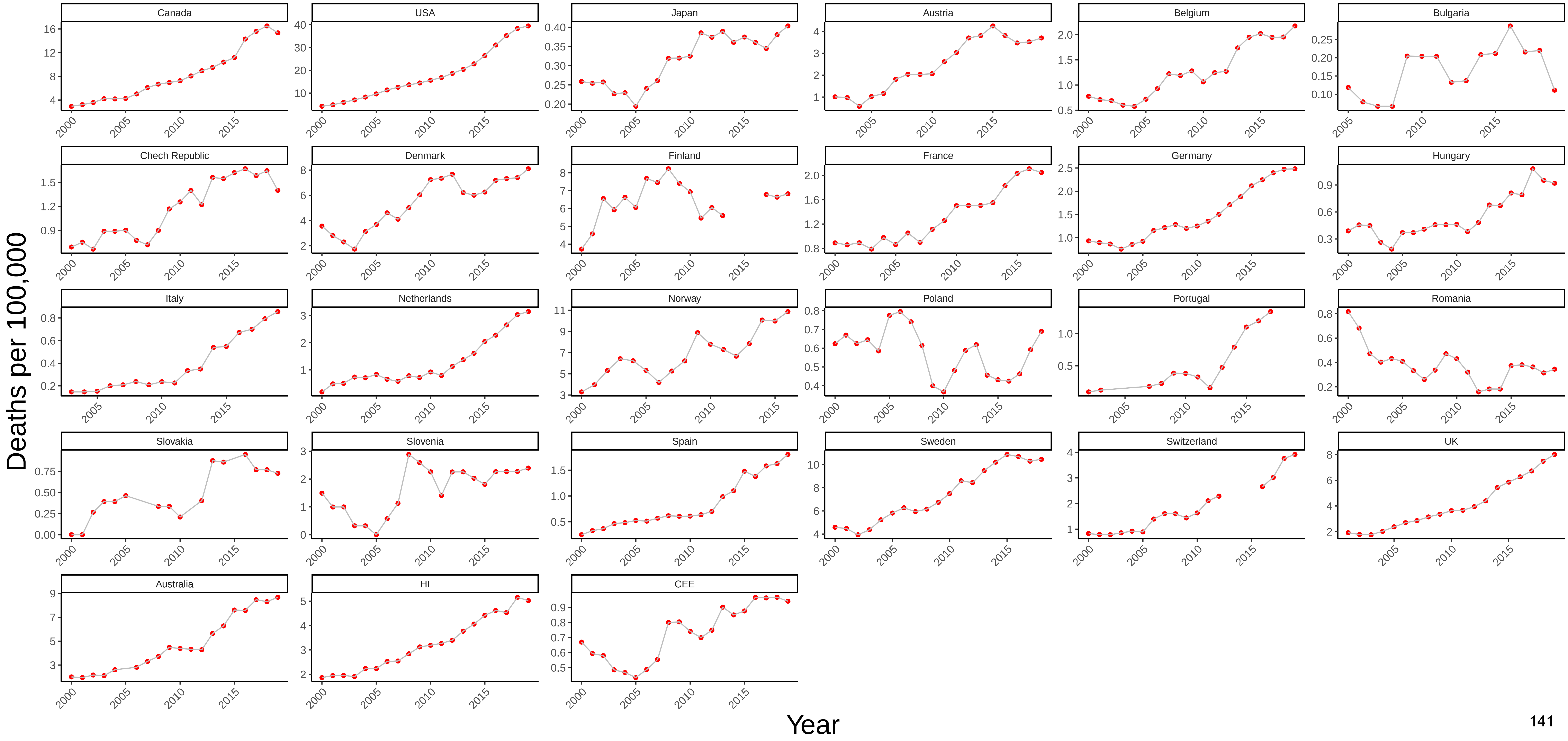

### Figure 94 Three−Year Moving Average of Female Mortality from Drug−Related Causes at Ages 55−64

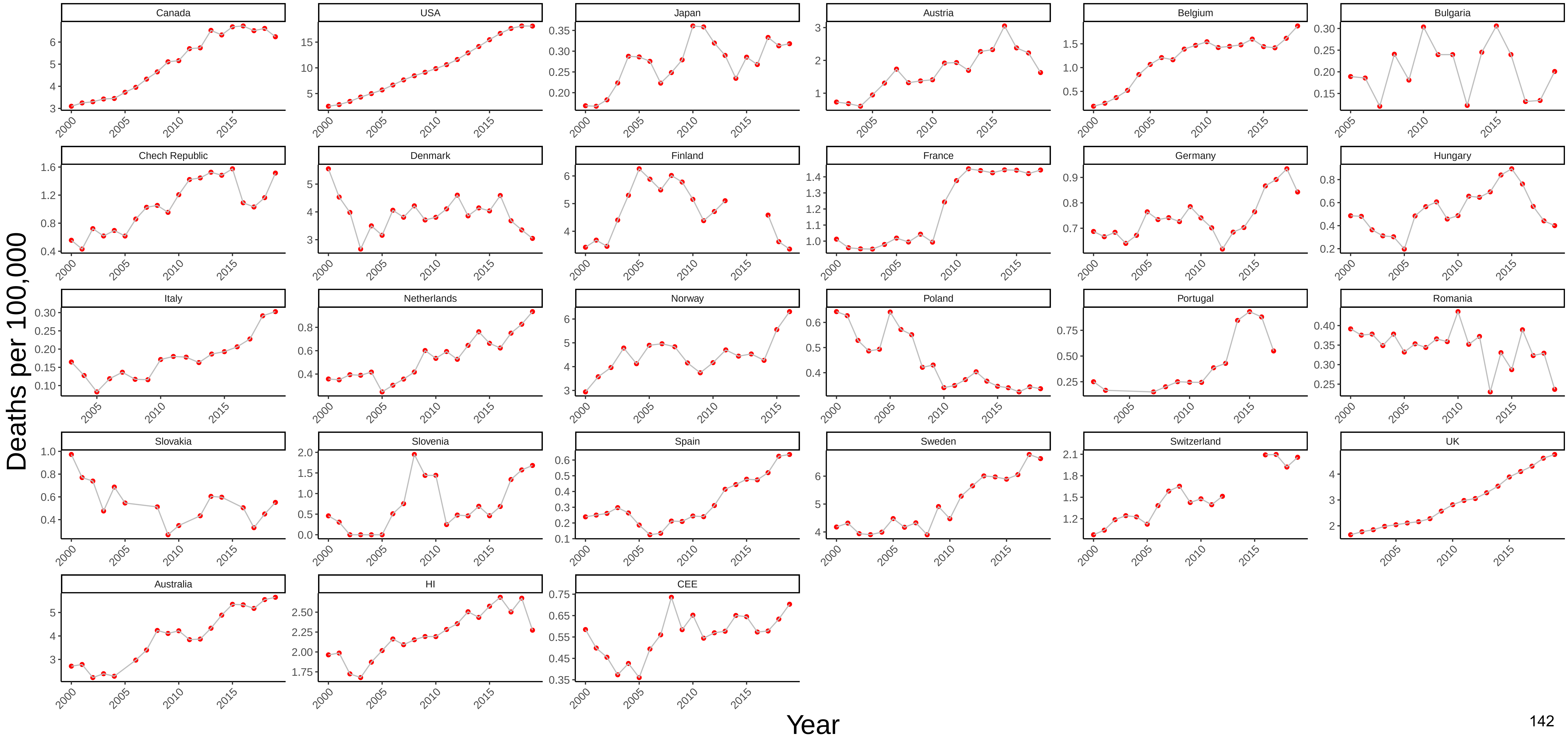

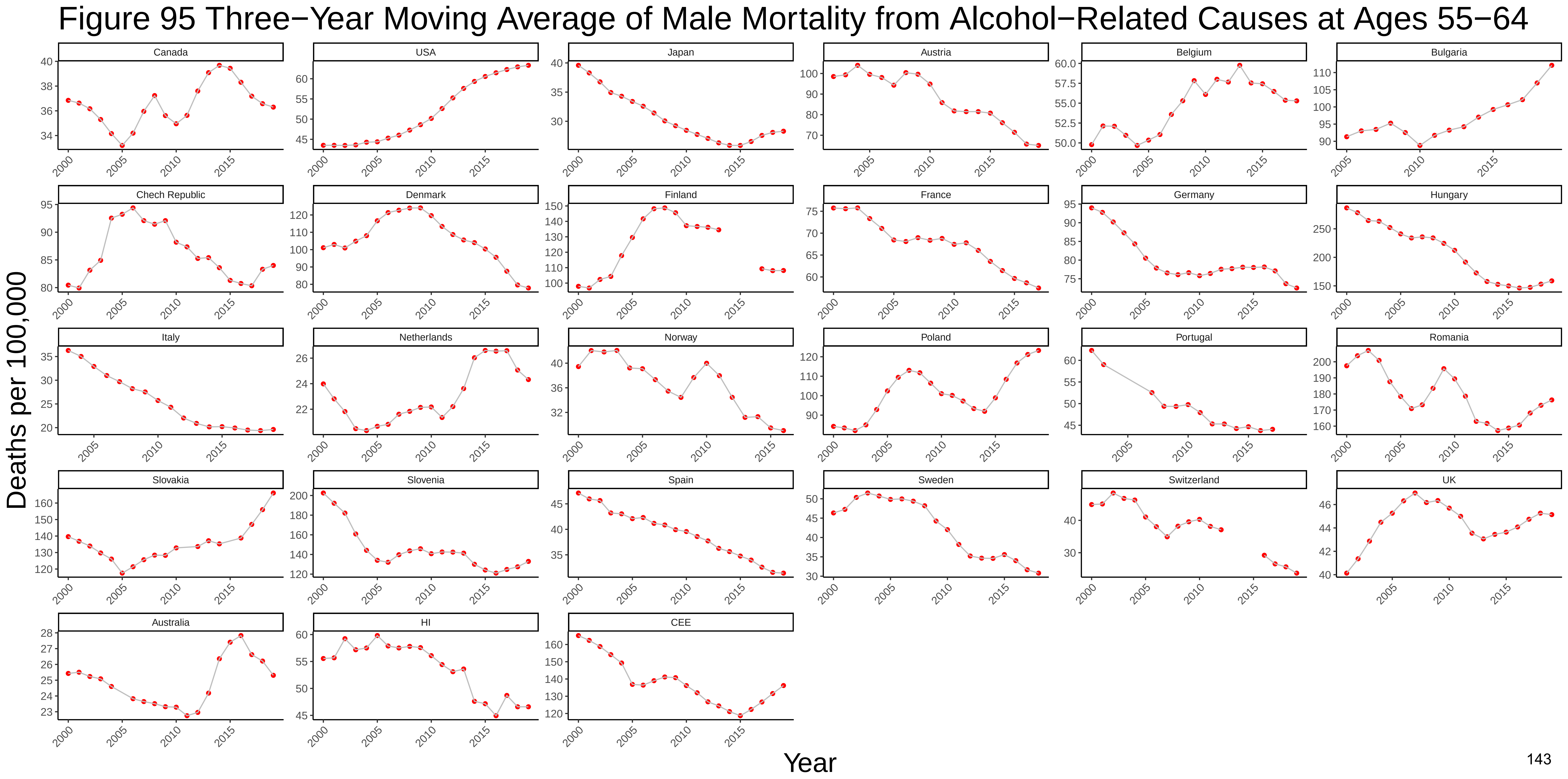

Figure 95 Three−Year Moving Average of Male Mortality from Alcohol−Related Causes at Ages 55−64
Canada
USA
Japan
Austria
Belgium
Bulgaria
Chech Republic
Denmark
Finland
France
Germany
Hungary
Italy
Netherlands
Norway
Poland
Portugal
Romania
Slovakia
Slovenia
Spain
Sweden
Switzerland
UK
Australia
HI
CEE
Deaths per 100,000
Year

### Figure 96 Three−Year Moving Average of Female Mortality from Alcohol−Related Causes at Ages 55−64

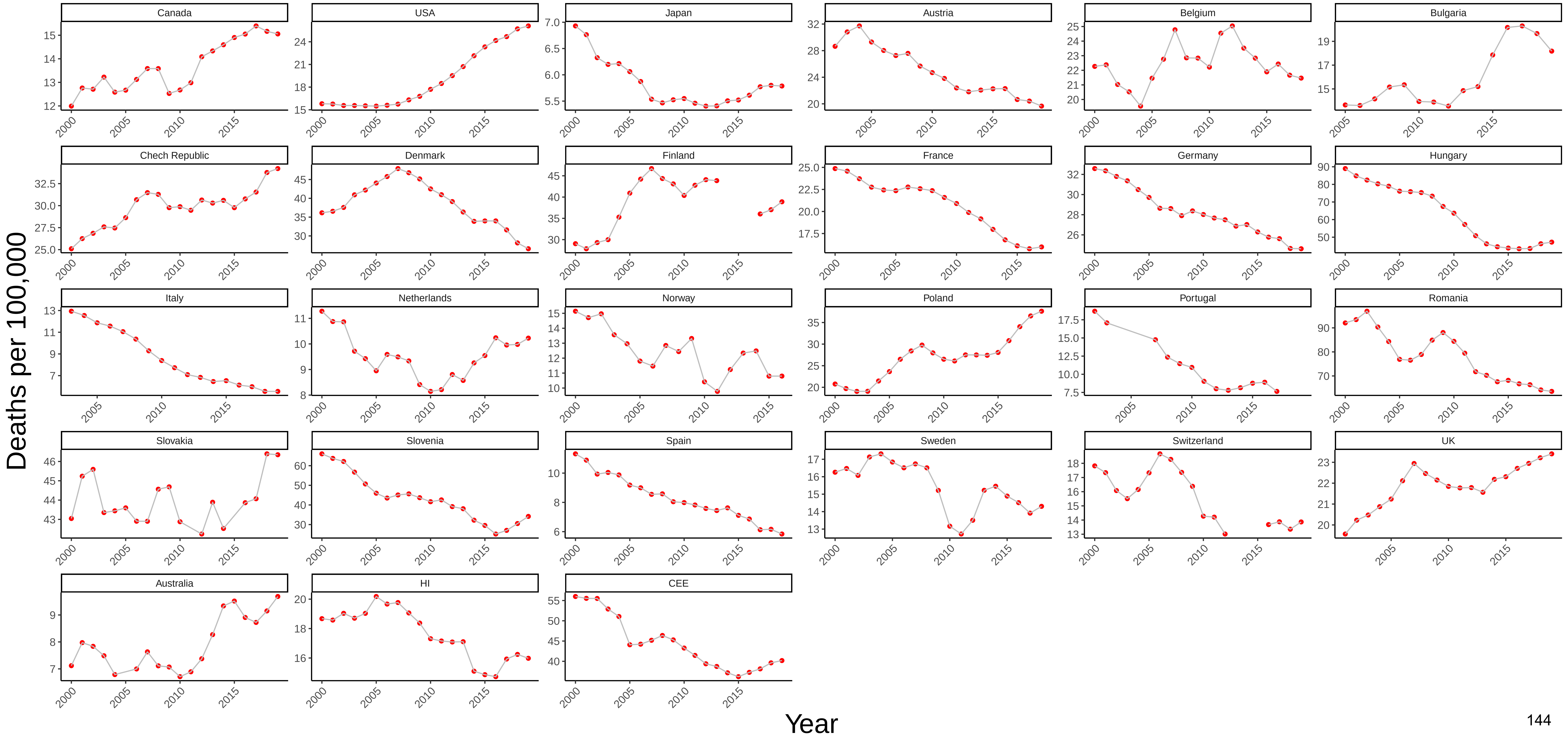
